# Head-to-head comparison of leading blood tests for Alzheimer’s disease pathology

**DOI:** 10.1101/2024.06.12.24308839

**Authors:** Suzanne E. Schindler, Kellen K. Petersen, Benjamin Saef, Duygu Tosun, Leslie M. Shaw, Henrik Zetterberg, Jeffrey L. Dage, Kyle Ferber, Gallen Triana-Baltzer, Lei Du-Cuny, Yan Li, Janaky Coomaraswamy, Michael Baratta, Yulia Mordashova, Ziad S. Saad, David L. Raunig, Nicholas J. Ashton, Emily A. Meyers, Carrie E. Rubel, Erin G. Rosenbaugh, Anthony W. Bannon, William Z. Potter, Alzheimer’s Disease Neuroimaging Initiative (ADNI), Foundation for the National Institutes of Health (FNIH) Biomarkers Consortium Plasma Aβ and Phosphorylated Tau as Predictors of Amyloid and Tau Positivity in Alzheimer’s Disease Project Team

**Affiliations:** Department of Neurology, Washington University in St. Louis, 660 S. Euclid Ave., St. Louis, MO, USA 63110; Department of Radiology and Biomedical Imaging, University of California San Francisco, 505 Parnassus Ave, San Francisco, CA, USA 94143; Department of Pathology and Laboratory Medicine, Perelman School of Medicine, University of Pennsylvania, 3400 Spruce St., Philadelphia, PA, USA 19104; Institute of Neuroscience and Physiology, Department of Psychiatry and Neurochemistry, The Sahlgrenska Academy at University of Gothenburg, Wallinsgatan 6, 431 41, Mölndal, Sweden; Clinical Neurochemistry Laboratory, Sahlgrenska University Hospital, Wallinsgatan 6, 431 41, Mölndal, Sweden; UK Dementia Research Institute Fluid Biomarkers Laboratory, UK DRI at UCL, 6th Floor, Maple House, Tottenham Ct Rd, London W1T 7NF, United Kingdom; Department of Neurodegenerative Disease, UCL Queen Square Institute of Neurology, Queen Square, London WC1N 3BG, United Kingdom; Hong Kong Center for Neurodegenerative Diseases, Clear Water Bay, Units 1501-1502, 1512-1518, 15/F Building 17W, 17 Science Park W Ave, Science Park, Hong Kong, China; Wisconsin Alzheimer’s Disease Research Center, University of Wisconsin School of Medicine and Public Health, University of Wisconsin-Madison, 600 Highland Ave J5/1 Mezzanine, Madison, WI, USA 53792; Department of Neurology, Indiana University School of Medicine, 355 W 16th St Indianapolis, IN, USA 46202; Stark Neurosciences Research Institute, Indiana University School of Medicine, NB Building, 320 W 15th St #414, Indianapolis, IN, USA 46202; Biogen, 225 Binney St., Cambridge, MA, USA 02142; Neuroscience Biomarkers, Johnson and Johnson Innovative Medicine, 3210 Merryfield Row, San Diego, CA, USA 20892; AbbVie, Knollstraße 50, 67061, Ludwigshafen am Rhein, Rheinland-Pfalz, Germany; Takeda Pharmaceutical Company Ltd., 650 E Kendall St., Cambridge, MA, USA 02142; Banner Alzheimer’s Institute, 901 E Willetta St., Phoenix, AZ, USA 85006; Banner Sun Health Research Institute, 10515 W Santa Fe Dr., Sun City, AZ, USA 85351; Alzheimer’s Association, 225 N Michigan Ave., Chicago, IL, USA 60603; The Foundation for the National Institutes of Health, 11400 Rockville Pike, Suite 600, North Bethesda, MD, USA 20852; AbbVie, 1400 Sheridan Rd., North Chicago, IL, USA 60064; Highly qualified expert

**Keywords:** Blood, plasma, biomarkers, amyloid, tau, p-tau, A/T/N, neurofilament light, glial fibrillary acidic protein

## Abstract

**Introduction:** Blood tests have the potential to improve the accuracy of Alzheimer disease (AD) clinical diagnosis, which will enable greater access to AD-specific treatments. This study compared leading commercial blood tests for amyloid pathology and other AD-related outcomes.

**Methods:** Plasma samples from the Alzheimer’s Disease Neuroimaging Initiative were assayed with AD blood tests from C2N Diagnostics, Fujirebio Diagnostics, ALZPath, Janssen, Roche Diagnostics, and Quanterix. Outcomes measures were amyloid positron emission tomography (PET), tau PET, cortical thickness, and dementia severity. Logistic regression models assessed the classification accuracies of individual or combined plasma biomarkers for binarized outcomes, and Spearman correlations evaluated continuous relationships between individual plasma biomarkers and continuous outcomes.

**Results:** Measures of plasma p-tau217, either individually or in combination with other plasma biomarkers, had the strongest relationships with all AD outcomes.

**Discussion:** This study identified the plasma biomarker analytes and assays that most accurately classified amyloid pathology and other AD-related outcomes.

## 1. Background

Blood biomarker (BBM) tests for Alzheimer disease (AD) have rapidly advanced over the past five years and are now being increasingly used in research studies, clinical trials, and clinical practice [1, 2]. The high acceptability of blood collection by patients and the wide availability of blood collection for clinical care are major advantages of BBM tests compared to cerebrospinal fluid (CSF) tests and positron emission tomography (PET) [3]. Moreover, the recent FDA-approval of AD-specific treatments that require biomarker confirmation of amyloid pathology has greatly increased the need for biomarker testing [4, 5], and BBM tests may be the only modality that can enable the scale of AD biomarker testing that is necessary for widespread use of AD-specific treatments [4–6]. Given these recent major changes in the field, the 2018 National Institute on Aging-Alzheimer’s Association (NIA-AA) AT(N) (amyloid, tau, neurodegeneration) research framework for AD is being updated in 2024 and includes guidelines for use of BBM tests [7].

One issue that has become an obstacle for broader use of BBM tests is widely varying levels of accuracy and validation for different BBM tests. Studies have demonstrated that different assays have accuracies that range from equivalent to FDA-approved CSF tests to not much better than chance [8–10]. These differences may be related to a variety of factors including the type of assay (e.g., mass spectrometry-based or immunoassay), the characteristics of the antibodies used, and the precision of the assay. Differences in study cohorts further complicate attempts to validate assays, as performance metrics may vary depending on the composition of the cohort [11]. Relatively few head-to-head studies, in which the exact same samples are analyzed with multiple assays, have been performed because these studies require a large sample volume and are expensive [8–10, 12]. However, head-to-head studies are essential to rigorously comparing performance between assays.

Anticipating the increasing importance of AD BBM tests, the Biomarker Consortium for the Foundation for the National Institutes of Health (FNIH) coordinated a head-to-head comparison of leading plasma assays for Aβ42/Aβ40 in 2020 to determine which assays added value to covariates in classification of amyloid PET status. The Biomarker Consortium included stakeholders from academia, industry, and patient advocacy groups. After evaluating six different plasma Aβ42/Aβ40 assays [8], the FNIH Biomarker Consortium then expanded its focus to additional plasma analytes, including phosphorylated tau species that had been demonstrated to classify amyloid PET status even more accurately than Aβ42/Aβ40 [13].

For the current project, the FNIH Biomarker Consortium sought to compare leading assays for Aβ42/Aβ40 and tau phosphorylated at positions 217 and 181 (p-tau217 and p-tau181). Additionally, glial fibrillary acidic protein (GFAP) and neurofilament light (NfL), which may add value to Aβ42/Aβ40 and/or p-tau species in predicting some AD-related outcomes [14, 15], were included in some assay panels.

Leading commercial assays for each analyte were identified by scientists involved in assay selection for clinical trials: C2N Diagnostics’ PrecivityAD2 %p-tau217, p-tau217, and Aβ42/Aβ40; Fujirebio Diagnostics’ Lumipulse p-tau217 and Aβ42/Aβ40; ALZpath’s Quanterix p-tau217; Janssen’s LucentAD Quanterix p-tau217; Roche Diagnostics’ NeuroToolKit p-tau181, Aβ42/Aβ40, GFAP, and NfL; and Quanterix’s Neurology 4-Plex p-tau181, Aβ42/Aβ40, GFAP, and NfL. Plasma samples were provided by the Alzheimer’s Disease Neuroimaging Initiative (ADNI) biorepository and a prespecified analytic plan was drafted for a head-to-head comparison of these assays with amyloid PET status as the primary outcome. This analytic plan was expanded to include the degree to which the different assays predicted tau PET status, cortical thickness, and cognitive impairment. Data from this study is available to researchers interested in performing further studies to compare the performance of these assays, as well as to evaluate other scientific questions (adni.loni.usc.edu).

## 2. Methods

### 2.1 Participants

The over-arching goal of this FNIH Biomarker Consortium effort is to evaluate the longitudinal trajectories of p-tau217, p-tau181 and Aβ42/Aβ40 in relation to amyloid PET. Therefore, ADNI participants were selected for inclusion who had plasma samples collected within 6 months of an amyloid PET scan for three distinct timepoints. For individuals with more than three time points, plasma samples were selected that represented the earliest, latest, and an intermediate time point. Based on these selection criteria, 393 ADNI participants had at least six plasma aliquots (1,179 samples total) that underwent analysis with the C2N Diagnostics PrecivityAD2 (C2N), Fujirebio Diagnostics Lumipulse (Fujirebio), ALZpath Quanterix (ALZpath), Janssen LucentAD Quanterix (Janssen), and Roche Diagnostics NeuroToolKit (Roche) assays. A seventh aliquot was available for 355 participants (1,063 participant time points, two participants had a seventh plasma aliquot for only two time points), and this aliquot was used for the remaining Quanterix Neurology 4-Plex (Quanterix) assays, which were given a lower priority due to lower accuracy reported by prior studies [9, 10].

For this current head-to-head study, samples were considered for inclusion if 1) no data for the C2N PrecivityAD2, Fujirebio Lumipulse, ALZpath Quanterix, Janssen LucentAD Quanterix, and Roche NeuroToolKit assays were missing (e.g., due to quality control failures or low volume) and if clinical data was available within one year of the sample collection. For samples that met these criteria, the most recently collected sample was chosen from each individual in order to increase the availability of associated tau PET data. One individual was excluded due to missing data from each of the three plasma samples, yielding 392 individuals in this study. Quanterix Neurology 4-Plex data was available for 342 of these individuals.

The current analyses utilized data from the ADNI database (adni.loni.usc.edu), which studies memory and aging in older adults, including both cognitively unimpaired and cognitively impaired individuals. ADNI was launched in 2003 as a public-private partnership led by Principal Investigator Michael W. Weiner, MD. The primary goal of ADNI has been to test whether serial magnetic resonance imaging (MRI), positron emission tomography (PET), other biological markers, and clinical and neuropsychological assessment can be combined to measure the progression of mild cognitive impairment (MCI) and early Alzheimer’s disease (AD). Written informed consent was obtained from each participant or their legally authorized representative. Race and sex were self-identified.

### 2.2 Diagnostic and CDR variables

Participants underwent a clinical assessment within one year of blood collection that included a detailed interview of a collateral source, a neurological examination of the participant, the Clinical Dementia Rating® (CDR®) and the Clinical Dementia Rating Sum of Boxes (CDR-SB) [16]. Individuals with a CDR of 0 were categorized as “cognitively unimpaired.” Individuals with a CDR>0 were categorized as “cognitively impaired;” this group includes individuals with mild cognitive impairment and dementia.

### 2.3 Plasma biomarker assays

The FNIH Biomarker Consortium project team selected assays based on recently published [17–24] as well as currently unpublished data presented by diagnostics companies. This included information on the analytical performance of each assay, such as the measurement range, dilutional linearity, and repeatability. Ultimately, four Aβ42/Aβ40, two p-tau181, four p-tau217, two GFAP, and two NfL assays were selected for this project.

Plasma was collected, processed, and stored according to ADNI protocols. Sample aliquots were pulled from the ADNI biorepository and shipped to four laboratories for analysis on dry ice. Pooled plasma controls with known higher or lower plasma p-tau217 values as measured by the ALZpath Quanterix assay were supplied by the University of Gothenburg to be run with each lot of samples. All laboratories were blinded to information about the participants.

The C2N PrecivityAD2 liquid chromatography tandem mass spectrometry (LC-MS) based assays for Aβ42, Aβ40, p-tau217, and non-phosphorylated tau217 (np-tau217) were run in singlicate at the C2N Diagnostics commercial laboratory in St. Louis, Missouri, USA. The %p-tau217 measure was calculated as the p-tau217 concentration divided by the np-tau217 concentration times 100, which is also described as the percent phosphorylation occupancy [20, 25]. The Fujirebio Lumipulse assays for Aβ42, Aβ40, and p-tau217 were run in singlicate with commercially available kits on a Fujirebio Lumipulse G1200 analyzer at the Indiana University National Centralized Repository for Alzheimer’s Disease and Related Dementias Biomarker Assay Laboratory (NCRAD-BAL). The Roche NeuroToolKit assays for Aβ42, Aβ40, p-tau181, NfL, and GFAP were run in singlicate on a Cobas® e 801 analyzer (Roche Diagnostics International Ltd, Rotkreuz, Switzerland) at the University of Gothenburg. The ALZpath Quanterix and Janssen LucentAD Quanterix assays for p-tau217, as well as the Quanterix Neurology 4-Plex E (N4PE) assays for Aβ42, Aβ40, p-tau181 (v2.1), NfL, and GFAP, were run in duplicate on a Quanterix Simoa-HD-X analyzer at the Quanterix Accelerator Laboratory. Further details are included in the study methodology report, which may be accessed from the ADNI database (adni.loni.usc.edu).

### 2.4 CSF collection and analysis and APOE genotyping

Data on CSF biomarkers and *APOE* genotype were obtained from ADNI. CSF Aβ42, total tau, and p-tau181 were measured using the Elecsys® β-amyloid (1–42) CSF, Elecsys Total-Tau CSF, and Elecsys Phospho-Tau (181P) CSF immunoassays, respectively, on a Cobas e 601 analyzer (Roche Diagnostics International Ltd, Rotkreuz, Switzerland). *APOE* genotyping was performed as part of the ADNI protocol.

### 2.5 Amyloid PET, tau PET and MRI variables

Amyloid PET imaging was conducted at each ADNI site following standardized protocols for florbetapir and florbetaben [26]. A global standardized uptake value ratio (SUVR) was estimated across cortical summary regions, specifically the frontal, cingulate, parietal, and lateral temporal cortices, as defined by FreeSurfer v7.1. This ratio was normalized to the whole cerebellum. To estimate the standardized global cortical Aβ burden in Centiloid units, transformations provided by the ADNI PET Core were applied to florbetaben and florbetapir. The threshold for amyloid PET positivity was set at >20 Centiloids, which corresponds to the recommended thresholds for florbetapir (whole cerebellum-normalized SUVR of 1.11 [27]) and florbetaben (whole cerebellum-normalized SUVR of 1.08 [28]).

Tau PET imaging was conducted at each ADNI site following standardized protocols for flortaucipir. The four 5-minute flortaucipir PET scans were corrected for motion, averaged, smoothed to a uniform resolution of 6 mm, and co-registered to the individual participant’s accompanying T1-weighted MRI scan. Using the CenTauR standardized masks [29], SUVRs were estimated for regions of interest (ROIs) as defined by FreeSurfer v7.1. The mesial temporal meta-ROI included regions with early tau PET signal (entorhinal, parahippocampus and amygdala), and the temporal-parietal meta-ROI included regions with both early and later tau PET signal (the entorhinal, parahippocampus, amygdala, fusiform, inferior and middle temporal gyri). These SUVRs were normalized to the inferior cerebellar gray matter reference region, as defined by the SUIT template [30]. The derivation of cut-offs for early and late tau PET positivity are described in **Appendix A**.

Structural brain MRI data included a 3D MP-RAGE or IR-SPGR T1-weighted 3T MRI with sagittal slices. The full cortex of each subject brain was parcellated into 41 bilateral ROIs using a volumetric Desikan-Killiany-Tourville atlas, using FreeSurfer v5.1 for ADNI-GO/2 data and v6.0 for ADNI-3 data. A composite meta-ROI cortical thickness measurement was calculated that included the entorhinal, fusiform, parahippocampal, mid-temporal, inferior-temporal, and angular gyrus [31]. To address differences in image acquisition protocol and image processing, the meta-ROI cortical thickness measure was harmonized using the ComBat-GAM method [32]. Harmonization parameters specific to each batch were estimated using data from the cognitively unimpaired individuals, while considering normal variance due to age, and sex. The age effect was modeled as a non-linear effect over the study age range. The estimated harmonization parameters were then applied to the full dataset. Finally, normal aging effects on the meta-ROI cortical thickness (harmonized) was estimated within cognitively unimpaired and Aβ-negative individuals and regressed out from the corresponding measures from all individuals (using age and quadratic age as predictors). The steps for ComBAT-GAM harmonization and adjusting for normal effects of age utilized all available ADNI data for unbiased modeling. The derivation of cut-offs for cortical thickness abnormality are described in **Appendix A**.

### 2.6 Statistical methods

The specific ADNI datasets used by this study are listed and the annotated code used for study analyses is provided in full so that other investigators can easily replicate and extended our analyses (see **Appendix B**). For participant characteristics tables, the significance of differences by amyloid PET status were evaluated with Wilcoxon ranked sum tests for continuous variables and Chi-Square or Fisher exact tests for categorical variables. The percent difference by amyloid PET status in the median plasma biomarker level was calculated as the difference between the median of the amyloid PET positive and amyloid PET negative groups divided by the median for the amyloid PET negative group times 100.

Unadjusted logistic regression models for a binary outcome (amyloid PET status, tau PET status, cortical thickness status, or cognitive impairment status [CDR=0 or CDR>0]) were used to assess the classification accuracy of plasma biomarker analytes separately or combined within a company/assay panel. Brier scores were used to quantify the prediction accuracy of the logistic regression models [33]. Logistic regression models adjusted for the covariates of age, sex, and *APOE* genotype were also examined. Differences between receiver operating characteristic areas under the curves (AUCs) were evaluated using DeLong tests [34]. When multiple AUCs were compared, the significance was adjusted using the Benjamini-Hochberg procedure [35]. Cut-off values for separate analytes that yielded the highest combined sensitivity and specificity (Youden Index) for an outcome were found. For each cut-off, the following metrics were calculated: sensitivity, specificity, overall accuracy, positive predictive value, and negative predictive value.

Spearman correlations were used to evaluate the continuous relationships of plasma biomarkers with amyloid PET Centiloid, tau PET measures, cortical thickness measure, and dementia severity as measured by the CDR-SB. For partial Spearman correlations, analyses included covariates of age, sex and *APOE* genotype. Comparisons between Spearman correlations were performed by bootstrapping. When multiple measures were compared, the significance was adjusted using the Benjamini-Hochberg procedure [35].

## 3. Results

### Participants

The full cohort included data from 392 individuals with a median age of 78.1 years (interquartile range 72.9-83.3 years); 193 (49.2%) were female; 132 (33.7%) carried an *APOE* ε4 allele; 366 (93.4%) self-identified as their race as white; 191 (48.7%) were amyloid PET positive; and 192 (49.0%) were cognitively impaired (CDR>0), (**Table 1**). Of the participants with cognitive impairment, 148 had mild cognitive impairment or very mild dementia (CDR 0.5) and 44 had dementia (CDR 1 or higher), (see **Tables S1-S2** for characteristics of the cognitively impaired and unimpaired sub-cohorts). Within one year of plasma collection, all individuals in the full cohort had amyloid PET, 188 had tau PET, 266 had structural brain MRI, and 122 had CSF collection with Elecsys data for CSF p-tau181 and Aβ42 (see **Tables S3-5** for characteristics of the tau PET, brain MRI, and CSF sub-cohorts).

**Table 1.** Cohort characteristics. Continuous values are presented as the median with the interquartile range. The significance of differences by amyloid PET status were evaluated with Wilcoxon’s rank-sum tests for continuous variables and Chi square or Fisher’s exact tests for categorical variables. All tests were two sided and not adjusted for covariates or multiple comparisons. Please see the methods and **Appendix A** for definitions of early tau PET, late tau PET, and cortical thickness.

| Characteristic | Full cohort |  | Amyloid PET negative |  | Amyloid PET positive |  |  |
| --- | --- | --- | --- | --- | --- | --- | --- |
|  | n= | Values | n= | Values | n= | Values | p= |
| **Demographics** |  |  |  |  |  |  |  |
| Age (years) | 392 | 78.1 (72.9-83.3) | 201 | 76.1 (71.8-81.9) | 191 | 79.0 (74.5-84.2) | 0.0026 |
| Sex (n, % female) | 392 | 193 (49.2%) | 201 | 98 (48.8%) | 191 | 95 (49.7%) | 0.80 |
| APOE genotype (22/23/24/33/34/44) | 392 | 1/35/6/224/108/18 | 201 | 1/29/1/133/31/6 | 191 | 0/6/5/91/77/12 | <0.0001 |
| APOE ε4 carrier status (n, % carrier) | 392 | 132 (33.7%) | 201 | 38 (18.9%) | 191 | 94 (49.2%) | <0.0001 |
| Years of education | 392 | 16 (14-18) | 201 | 16 (14-19) | 191 | 16 (14-18) | 0.10 |
| Race (Black/White/Other) | 392 | 14/366/12 | 201 | 8/184/9 | 191 | 6/182/3 | 0.22 |
| CDR (0/0.5/1+) | 392 | 200/148/44 | 201 | 124/47/3 | 191 | 76/74/41 | 0.025 |
| CDR Sum of Boxes | 392 | 0 (0-1.5) | 201 | 0 (0-1) | 191 | 1 (0-3) | <0.0001 |
| Plasma collection to CDR (years) | 392 | 0 (0-0.02) | 201 | 0 (0-0.02) | 191 | 0 (0-0.02) | 0.36 |
| **Amyloid PET** |  |  |  |  |  |  |  |
| Amyloid PET Centiloid | 392 | 19 (-2-68) | 201 | -2 (-9-6) | 191 | 69 (47-99) | <0.0001 |
| Plasma collection to amyloid PET (years) | 392 | 0.02 (0-0.04) | 201 | 0.02 (0-0.04) | 191 | 0.02 (0.003-0.04) | 0.88 |
| **Tau PET** |  |  |  |  |  |  |  |
| Early Tau PET | 188 | 1.18 (1.1-1.3) | 97 | 1.14 (1.06-1.23) | 91 | 1.28 (1.15-1.44) | <0.0001 |
| Early Tau PET positivity | 188 | 40, 21.3% | 97 | 4, 4.1% | 91 | 36, 39.6% | <0.0001 |
| Late Tau PET | 188 | 1.13 (1.07-1.2) | 97 | 1.11 (1.05-1.15) | 91 | 1.17 (1.09-1.32) | <0.0001 |
| Late Tau PET positivity | 188 | 29, 15.4% | 97 | 2, 2.1% | 91 | 27, 29.7% | <0.0001 |
| Plasma collection to tau PET (years) | 188 | 0.02 (0.003-0.09) | 97 | 0.02 (0.003-0.08) | 91 | 0.02 (0.003-0.11) | 0.54 |
| **Cortical thickness** |  |  |  |  |  |  |  |
| Cortical thickness | 266 | 2.8 (2.69-2.89) | 144 | 2.82 (2.75-2.89) | 122 | 2.76 (2.62-2.88) | 0.00284 |
| Cortical thickness positivity | 266 | 27, 10.2% | 144 | 5, 3.5% | 122 | 22, 18.0% | <0.0001 |
| Plasma collection to MRI (years) | 266 | 0.01 (0-0.04) | 144 | 0.008 (0-0.04) | 122 | 0.01 (0-0.05) | 0.053 |
| **CSF Elecsys** |  |  |  |  |  |  |  |
| CSF p-tau181/Aβ42 | 122 | 0.0267 (0.0127-0.0506) | 52 | 0.0127 (0.00963-0.0162) | 70 | 0.0482 (0.0304-0.063) | <0.0001 |
| CSF p-tau181 (pg/ml) | 123 | 23.3 (17.7-30.9) | 52 | 19.2 (15.1-24.5) | 71 | 27.1 (22.2-36.6) | <0.0001 |
| CSF Aβ42 (pg/ml) | 125 | 860 (647-1360) | 55 | 1360 (1030-1830) | 70 | 691 (528-837) | <0.0001 |
| Plasma collection to CSF (years) | 122 | 0 (0-0.005) | 52 | 0 (0-0.005) | 70 | 0 (0-0) | 0.49 |

### Sensitivity and variance of assays

Of the 392 samples included, 110 samples had p-tau217 values that were below the level of detection for the C2N PrecivityAD2 assay and were assigned values of 0.65 pg/ml (half the value of the limit of detection). There were 12 samples with p-tau217 values that were below the level of detection for the Fujirebio Lumipulse assay and were assigned values of 0.030 pg/ml. Analyte values were within range for the other assays.

The variance in measurements was assessed using aliquots of two pooled control plasma samples with known higher or lower p-tau217 values that were run with each batch of samples. For most assays, the coefficient of variance (CV) ranged from 3-10% (see **Appendix C**). The exception was for Fujirebio Lumipulse p-tau217, which had CVs of 11% and 34% for the higher and lower controls, respectively. The low control was near the lower range of the Fujirebio Lumipulse p-tau217 assay, which is an explanation for the higher imprecision.

### Correlations between plasma biomarkers

Correlations between plasma biomarker levels across platforms were evaluated with samples from the 392 individuals (**Figure 1**). C2N PrecivityAD2 %p-tau217 and C2N PrecivityAD2 p-tau217 values were highly correlated (unadjusted Spearman ρ=0.94 [0.93-0.95]); notably, the numerator in %p-tau217 is p-tau217, so a high correlation was expected. ALZpath Quanterix and Janssen LucentAD Quanterix p-tau217 concentrations were also highly correlated (ρ=0.94 [0.92-0.95]). Compared to C2N PrecivityAD2 p-tau217, C2N PrecivityAD2 %p-tau217 had lower correlations with p-tau217 concentrations as measured by the three immunoassays: ALZpath Quanterix, ρ=0.89 versus 0.81, p=0.0002; Fujirebio Lumipulse, ρ=0.88 versus 0.81, p=0.001; Janssen LucentAD Quanterix ρ=0.88 versus 0.80, p=0.0002. C2N PrecivityAD2 %p-tau217 had much lower correlations with Roche NeuroToolKit p-tau181 (ρ=0.69 [0.63-0.74]) and Quanterix Neurology 4-Plex p-tau181 (ρ=0.54 [0.45-0.62]). The four Aβ42/Aβ40 measures had pairwise correlations that ranged from 0.63 to 0.43; the Fujirebio Lumipulse and Quanterix Neurology 4-Plex had the highest correlation (ρ=0.63 [0.55-0.70]). Roche NeuroToolKit and Quanterix Neurology 4-Plex concentrations of GFAP and NfL were highly correlated (ρ=0.94 [0.91-0.97] and ρ=0.96 [0.94-0.97], respectively).

**Figure 1.**
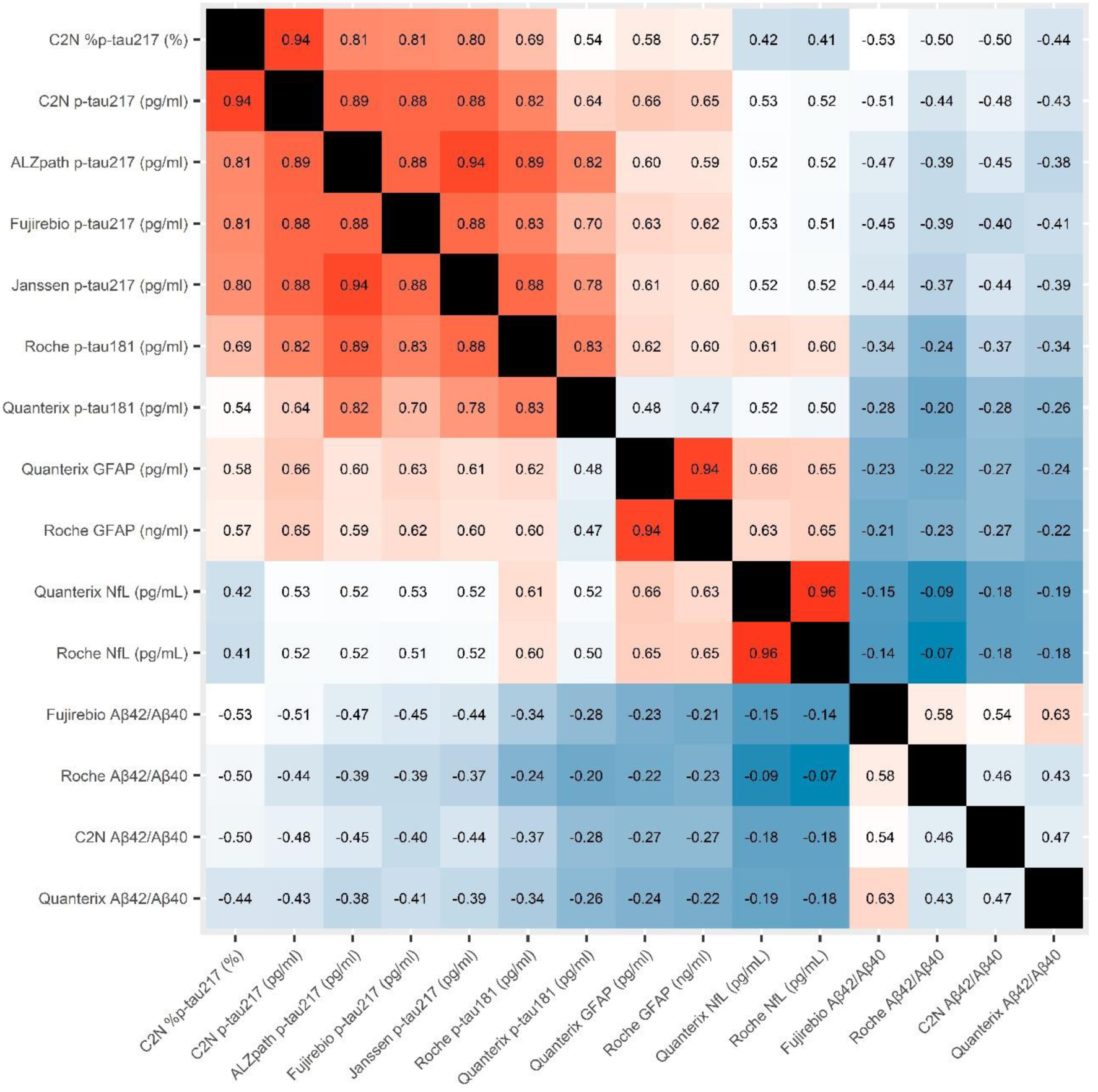
Correlation matrix of plasma biomarker measures in the full cohort. All individuals (n=392) had plasma biomarker measures for the C2N PrecivityAD2, Fujirebio Lumipulse, ALZpath Quanterix, Janssen LucentAD Quanterix, and Roche NeuroToolKit assays. A sub-cohort (n=342) additionally had plasma biomarker measures for the Quanterix Neurology 4-Plex assays. The unadjusted Spearman correlation of plasma biomarker measures with one another is shown. Redder colors are used for higher absolute correlations and bluer colors are used for lower absolute correlations. Black boxes represent identity.

### Differences in plasma biomarkers by amyloid PET status

Differences in plasma biomarker levels associated with positive amyloid PET status were examined. Because plasma biomarker values tend to be highly skewed and therefore mean levels may overestimate the degree of abnormality for the typical amyloid PET positive individual, percent differences were calculated for the median plasma biomarker levels (**Table 2**). Compared to the amyloid PET negative group, median plasma p-tau217 was higher in the amyloid PET positive group by 255% (Fujirebio Lumipulse), 245% (C2N PrecivityAD2 %p-tau217), 228% (C2N PrecivityAD2 p-tau217), 170% (ALZpath Quanterix) and 139% (Janssen LucentAD Quanterix). The percent difference in median p-tau181 was 66% (Roche NeuroToolKit) and 55% (Quanterix Neurology 4-Plex). Differences in median plasma Aβ42/Aβ40 levels were much smaller in magnitude, ranging from 13% to 10% lower in the amyloid PET positive group, but were still highly significant due to low variance in values. Median plasma GFAP and NfL were higher in the amyloid PET positive group (64 to 61% for GFAP, 28% to 27% for NfL).

**Table 2.** Plasma biomarker measures stratified by amyloid PET status. All individuals (n=392) had plasma biomarker measures for the C2N PrecivityAD2, Fujirebio Lumipulse, ALZpath Quanterix, Janssen LucentAD Quanterix, and Roche NeuroToolKit assays. A sub-cohort (n=342) additionally had plasma biomarker measures for the Quanterix Neurology 4-Plex assays. Continuous values are presented as the median with the interquartile range. The significance of unadjusted differences by amyloid PET status were evaluated with Wilcoxon’s rank-sum tests. All tests were two sided. The percent difference by amyloid PET status in the median plasma biomarker level is shown (difference in the medians for amyloid PET positive and negative groups divided by the median for the amyloid PET negative group times 100).

| Characteristic | Full cohort |  | Amyloid PET negative |  | Amyloid PET positive |  | Unadjusted p= | Percent difference |
| --- | --- | --- | --- | --- | --- | --- | --- | --- |
|  | n= | Values | n= | Values | n= | Values |  |  |
| C2N PrecivityAD2 |  |  |  |  |  |  |  |  |
| Aβ42 (pg/ml) | 392 | 40.1 (35.5-46.3) | 201 | 42.0 (37.2-47.5) | 191 | 39.2 (34.2-43.8) | 0.0001 | -6.7% |
| Aβ40 (pg/ml) | 392 | 441 (390-488) | 201 | 435 (385-487) | 191 | 447 (399-491) | 0.23 | 2.8% |
| Aβ42/Aβ40 | 392 | 0.0915 (0.0852-0.0997) | 201 | 0.0974 (0.0894-0.104) | 191 | 0.0871 (0.0821-0.0936) | <0.0001 | -10.6% |
| p-tau217 (pg/ml) | 392 | 2.27 (0.65-4.29) | 201 | 1.31 (0.65-1.86) | 191 | 4.3 (2.95-6.5) | <0.0001 | 228% |
| %p-tau217 (%) | 392 | 3.98 (1.86-7.54) | 201 | 2.20 (1.45-3.27) | 191 | 7.58 (5.24-10.5) | <0.0001 | 245% |
| np-tau217 (pg/ml) | 392 | 54.6 (44.3-66) | 201 | 51.8 (41.4-62.7) | 191 | 58.1 (48.7-71.7) | <0.0001 | 12.2% |
| Fujirebio Lumipulse |  |  |  |  |  |  |  |  |
| Aβ42 (pg/ml) | 392 | 26.9 (23.4-31) | 201 | 28.1 (24.2-32.6) | 191 | 25.5 (22.2-28.8) | <0.0001 | -9.3% |
| Aβ40 (pg/ml) | 392 | 301 (267-342) | 201 | 300 (267-338) | 191 | 307 (267-345) | 0.43 | 2.3% |
| Aβ42/Aβ40 | 392 | 0.0883 (0.0804-0.0965) | 201 | 0.0944 (0.0865-0.102) | 191 | 0.082 (0.0781-0.0887) | <0.0001 | -13.1% |
| p-tau217 (pg/ml) | 392 | 0.158 (0.089-0.326) | 201 | 0.094 (0.065-0.143) | 191 | 0.334 (0.196-0.52) | <0.0001 | 255% |
| ALZpath Quanterix |  |  |  |  |  |  |  |  |
| p-tau217 (pg/ml) | 392 | 0.432 (0.251-0.746) | 201 | 0.275 (0.202-0.395) | 191 | 0.741 (0.53-1.09) | <0.0001 | 170% |
| Janssen LucentAD Quanterix |  |  |  |  |  |  |  |  |
| p-tau217 (pg/ml) | 392 | 0.059 (0.038-0.1) | 201 | 0.041 (0.029-0.055) | 191 | 0.098 (0.069-0.143) | <0.0001 | 139% |
| Roche NeuroToolKit |  |  |  |  |  |  |  |  |
| Aβ42 (pg/ml) | 392 | 37.7 (32.4-42.7) | 201 | 39.3 (34.2-44.9) | 191 | 36.1 (30.7-40.9) | <0.0001 | -8.1% |
| Aβ40 (ng/ml) | 392 | 0.308 (0.276-0.342) | 201 | 0.305 (0.276-0.339) | 191 | 0.310 (0.278-0.343) | 0.54 | 1.6% |
| Aβ42/Aβ40 | 392 | 0.122 (0.111-0.133) | 201 | 0.131 (0.12-0.141) | 191 | 0.115 (0.108-0.122) | <0.0001 | -12.2% |
| p-tau181 (pg/ml) | 392 | 1.15 (0.855-1.58) | 201 | 0.922 (0.739-1.15) | 191 | 1.53 (1.15-1.95) | <0.0001 | 65.9% |
| GFAP (ng/ml) | 392 | 0.116 (0.0826-0.164) | 201 | 0.0908 (0.0712-0.126) | 191 | 0.149 (0.108-0.192) | <0.0001 | 64.1% |
| NfL (pg/ml) | 392 | 4.02 (3.11-5.39) | 201 | 3.64 (2.77-4.64) | 191 | 4.64 (3.57-6.12) | <0.0001 | 27.5% |
| Quanterix Neurology 4-Plex |  |  |  |  |  |  |  |  |
| Aβ42 (pg/ml) | 342 | 6.32 (5.36-7.43) | 183 | 6.68 (5.67-8.09) | 159 | 6.01 (5.04-6.87) | <0.0001 | -10.0% |
| Aβ40 (pg/ml) | 342 | 108 (91.8-130) | 183 | 106 (90-130) | 159 | 111 (93.3-130) | 0.60 | 4.7% |
| Aβ42/Aβ40 | 342 | 0.0578 (0.0518-0.0651) | 183 | 0.0617 (0.0556-0.0678) | 159 | 0.0545 (0.0483-0.0589) | <0.0001 | -11.7% |
| p-tau181 (pg/ml) | 342 | 20.3 (14.6-30.8) | 183 | 17.1 (12.4-22.5) | 159 | 26.5 (19.2-36.3) | <0.0001 | 55.0% |
| GFAP (pg/ml) | 342 | 168 (114-246) | 183 | 134 (103-184) | 159 | 216 (155-283) | <0.0001 | 61.2% |
| NfL (pg/ml) | 342 | 22.9 (16.6-30.8) | 183 | 20.1 (14.4-26.7) | 159 | 25.6 (19.8-37) | <0.0001 | 27.4% |

### Classification of amyloid PET status by plasma biomarkers

The classification accuracies of individual plasma biomarkers for amyloid PET status as defined by Centiloid >20 or ≤20 were evaluated using logistic regression models in the full cohort (**Figure 2 and Figure 3**). With the expectation that companies may provide a panel of plasma biomarkers, models that combined plasma biomarkers from the same company were also examined. The highest classification accuracies of amyloid PET status were achieved by C2N PrecivityAD2 and Fuijirebio Lumipulse models that included measures of both p-tau217 and Aβ42/Aβ40 (**Table 3**). However, the C2N PrecivityAD2 %p-tau217 and Aβ42/Aβ40 combined model (AUC 0.929 [0.902-0.956]) was not superior to the model with %p-tau217 alone (AUC 0.927 [0.900-0.955]), p=0.54. For C2N PrecivityAD2 %p-tau217, a cut-off of 4.06% yielded a positive percent agreement (PPA) of 88.5%, a negative percent agreement (NPA) of 87.1%, an overall percent accuracy of 87.8%, a positive predictive value (PPV) of 86.7%, and a negative predictive value (NPV) of 88.8% for amyloid PET status (**Table 4**). The Fujirebio Lumipulse p-tau217 and Aβ42/Aβ40 combined model (AUC 0.911 [0.882-0.940]) did not have significantly different performance from the C2N PrecivityAD2 %p-tau217 and Aβ42/Aβ40 combined model (p=0.16), or the model with Fujirebio Lumipulse p-tau217 alone (AUC 0.896 [0.864-0.928], p=0.12), (**Table 3**). For Fujirebio Lumipulse p-tau217, a cut-off of 0.158 yielded a PPA of 84.3%, a NPA of 82.1%, an accuracy of 83.2%, a PPV of 81.7%, and a NPV of 84.6% (**Table 4**).

**Figure 2.**
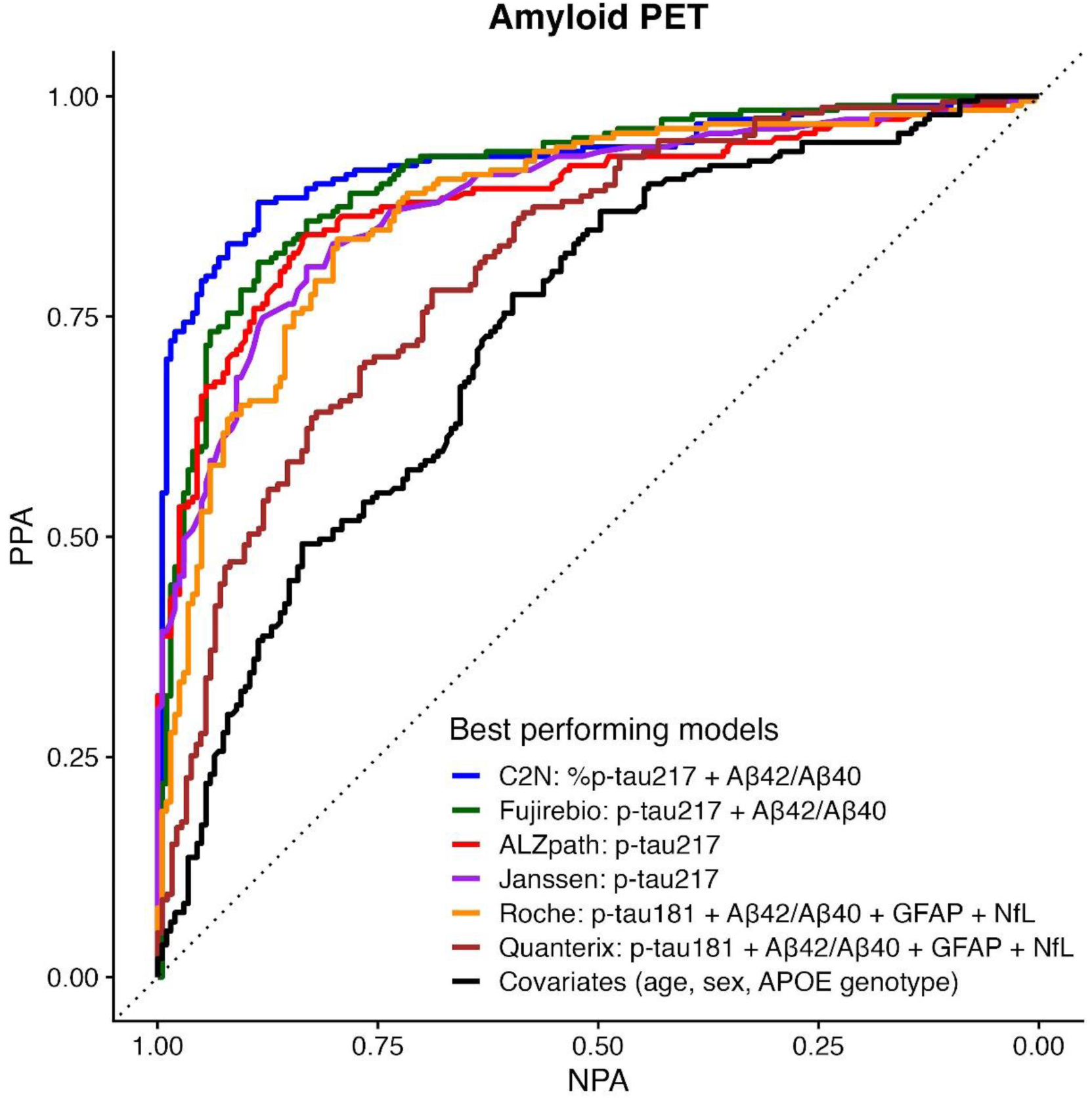
Receiver operating characteristics area under the curve plots for amyloid PET status in the full cohort. All individuals (n=392) had plasma biomarker measures for the C2N PrecivityAD2, Fujirebio Lumipulse, ALZpath Quanterix, Janssen LucentAD Quanterix, and Roche NeuroToolKit assays. A sub-cohort (n=342) additionally had plasma biomarker measures for the Quanterix Neurology 4-Plex assays. The best performing plasma biomarker measure or combination of measures from each company is shown for unadjusted models of amyloid PET > or ≤20 Centiloids. The solid black line represents a model of covariates alone. The dotted black line represents chance classification.

**Figure 3.**
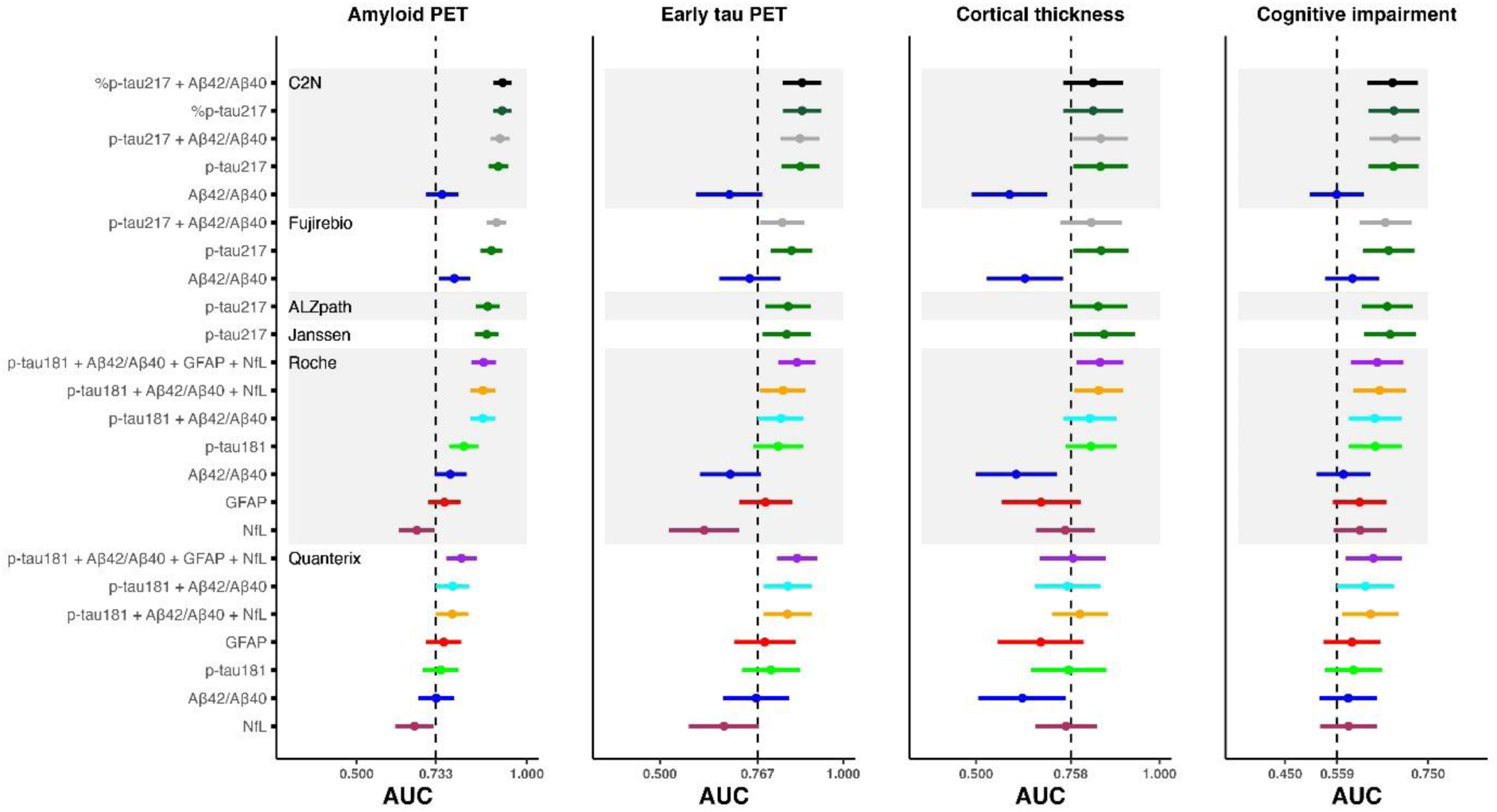
Classification accuracies of individual and combined plasma biomarker measures for key outcomes in the full cohort. All individuals (n=392) had plasma biomarker measures for the C2N PrecivityAD2, Fujirebio Lumipulse, ALZpath Quanterix, Janssen LucentAD Quanterix, and Roche NeuroToolKit assays. A sub-cohort (n=342) additionally had plasma biomarker measures for the Quanterix Neurology 4-Plex assays. The receiver operating characteristics area under the curve (AUC) point estimate (midpoint) and 95% confidence intervals are shown for classification of amyloid PET status (> or ≤20 Centiloids), early tau PET status, cortical thickness status, and cognitive impairment status (CDR>0 or =0) by individual or combined plasma biomarker measures. The dashed vertical reference lines represent the AUCs for models including only covariates (age, sex, *APOE genotype*) as predictors. Please see methods and **Appendix A** for definitions of early tau PET status and cortical thickness status.

**Table 3.** Classification accuracies of individual and combined plasma biomarker measures for amyloid PET status defined by Centiloid>20 or ≤20 in the full cohort. The receiver operating characteristics area under the curve (AUC) point estimate (midpoint) and 95% confidence intervals are shown for classification of amyloid PET status (> or ≤20 Centiloids) by individual or combined plasma biomarker measures. Both the unadjusted AUC and the AUC adjusted for age, sex, and *APOE* genotype are provided. AUCs were compared using DeLong’s test. The Benjamin-Hochberg procedure was used to adjust for multiple comparisons with the reference measure, either within a company or across companies. The AUC for a model with covariates only was 0.733 (0.684-0.782).

| Company | Analytes | Unadjusted for covariates |  |  | Adjusted for covariates |  |  |
| --- | --- | --- | --- | --- | --- | --- | --- |
|  |  | AUC | Within-company comparisons<br>p= | Across-company comparisons<br>p= | AUC | Within-company comparisons<br>p= | Across-company comparisons<br>p= |
| C2N<br>PrecivityAD2 | %p-tau217 + Aβ42/Aβ40 | 0.929 (0.902-0.956) | REFERENCE | REFERENCE | 0.932 (0.905-0.958) | REFERENCE | REFERENCE |
|  | %p-tau217 | 0.927 (0.900-0.955) | 0.54 |  | 0.931 (0.904-0.957) | 0.59 |  |
|  | p-tau217 + Aβ42/Aβ40 | 0.921 (0.893-0.950) | 0.21 |  | 0.926 (0.898-0.953) | 0.38 |  |
|  | p-tau217 | 0.916 (0.887-0.946) | 0.11 |  | 0.923 (0.895-0.951) | 0.35 |  |
|  | Aβ42/Aβ40 | 0.751 (0.703-0.799) | <0.0001 |  | 0.788 (0.743-0.833) | <0.0001 |  |
| Fujirebio<br>Lumipulse | p-tau217 + Aβ42/Aβ40 | 0.911 (0.882-0.940) | REFERENCE | 0.16 | 0.913 (0.886-0.941) | REFERENCE | 0.12 |
|  | p-tau217 | 0.896 (0.864-0.928) | 0.12 |  | 0.903 (0.873-0.932) | 0.17 |  |
|  | Aβ42/Aβ40 | 0.787 (0.741-0.833) | <0.0001 |  | 0.816 (0.774-0.859) | <0.0001 |  |
| ALZpath<br>Quanterix | p-tau217 | 0.885 (0.851-0.920) |  | 0.0006 | 0.903 (0.873-0.934) |  | 0.011 |
| Janssen<br>LucentAD<br>Quanterix | p-tau217 | 0.882 (0.848-0.916) |  | 0.0006 | 0.896 (0.864-0.927) |  | 0.0047 |
| Roche<br>NeuroToolKit | p-tau181 + Aβ42/Aβ40 + GFAP + NfL | 0.873 (0.838-0.909) | REFERENCE | 0.0006 | 0.888 (0.855-0.921) | REFERENCE | 0.0047 |
|  | p-tau181 + Aβ42/Aβ40 + NfL | 0.871 (0.835-0.908) | 0.67 |  | 0.884 (0.850-0.918) | 0.29 |  |
|  | p-tau181 + Aβ42/Aβ40 | 0.871 (0.834-0.907) | 0.67 |  | 0.884 (0.850-0.917) | 0.29 |  |
|  | p-tau181 | 0.815 (0.772-0.858) | 0.0003 |  | 0.851 (0.813-0.888) | 0.0011 |  |
|  | Aβ42/Aβ40 | 0.775 (0.728-0.823) | <0.0001 |  | 0.806 (0.763-0.850) | <0.0001 |  |
|  | GFAP | 0.758 (0.710-0.806) | <0.0001 |  | 0.814 (0.772-0.857) | <0.0001 |  |
|  | NfL | 0.677 (0.624-0.729) | <0.0001 |  | 0.775 (0.730-0.820) | <0.0001 |  |
| Quanterix<br>Neurology<br>4-Plex | p-tau181 + Aβ42/Aβ40 + GFAP + NfL | 0.808 (0.763-0.854) | REFERENCE | <0.0001 | 0.846 (0.805-0.887) | REFERENCE | 0.0032 |
|  | p-tau181 + Aβ42/Aβ40 | 0.782 (0.733-0.830) | 0.0832 |  | 0.820 (0.775-0.864) | 0.036 |  |
|  | p-tau181 + Aβ42/Aβ40 + NfL | 0.781 (0.733-0.829) | 0.0528 |  | 0.823 (0.779-0.867) | 0.036 |  |
|  | GFAP | 0.756 (0.704-0.807) | 0.0112 |  | 0.815 (0.769-0.860) | 0.016 |  |
|  | p-tau181 | 0.747 (0.694-0.799) | 0.0120 |  | 0.796 (0.750-0.843) | 0.0043 |  |
|  | Aβ42/Aβ40 | 0.734 (0.681-0.787) | 0.0046 |  | 0.786 (0.737-0.835) | 0.0022 |  |
|  | NfL | 0.670 (0.613-0.727) | <0.0001 |  | 0.769 (0.719-0.818) | <0.0001 |  |

**Table 4.** Classification accuracies of individual plasma biomarker measures for amyloid PET status defined by Centiloid>20 or ≤20 in the full cohort. The receiver operating characteristics area under the curve (AUC) point estimate (midpoint) and 95% confidence intervals are shown for classification of amyloid PET status (> or ≤20 Centiloids) by plasma biomarker measures. The single cut-off for the plasma biomarker that best distinguishes amyloid PET status based on the Youden index is shown, as well as the positive percent agreement (PPA) and negative percent agreement (NPA), overall accuracy, Brier score (confidence of model prediction), positive predictive value (PPV) and negative predictive value (NPV) of the cut-off for amyloid PET status in the full cohort, which has a 48.7% rate of amyloid PET positivity based on a cut-off of >20 Centiloids.

| Company | Analyte | AUC | Cut-off | Brier Score | PPA | NPA | Accuracy | PPV | NPV |
| --- | --- | --- | --- | --- | --- | --- | --- | --- | --- |
| **C2N**<br>**PrecivityAD2** | %p-tau217 | 0.927 (0.900-0.955) | 4.06 (%) | 0.098 | 0.885 | 0.871 | 0.878 | 0.867 | 0.888 |
|  | p-tau217 | 0.916 (0.887-0.946) | 2.34 (pg/ml) | 0.107 | 0.864 | 0.871 | 0.867 | 0.864 | 0.871 |
|  | Aβ42/Aβ40 | 0.751 (0.703-0.799) | 0.0924 | 0.205 | 0.717 | 0.672 | 0.694 | 0.675 | 0.714 |
| **Fujirebio**<br>**Lumipulse** | p-tau217 | 0.896 (0.864-0.928) | 0.158 (pg/ml) | 0.129 | 0.843 | 0.821 | 0.832 | 0.817 | 0.846 |
|  | Aβ42/Aβ40 | 0.787 (0.741-0.833) | 0.0869 | 0.190 | 0.723 | 0.751 | 0.737 | 0.734 | 0.740 |
| **ALZpath**<br>**Quanterix** | p-tau217 | 0.885 (0.851-0.920) | 0.444 (pg/ml) | 0.129 | 0.843 | 0.831 | 0.837 | 0.826 | 0.848 |
| **Janssen**<br>**LucentAD**<br>**Quanterix** | p-tau217 | 0.882 (0.848-0.916) | 0.0615 (pg/ml) | 0.138 | 0.806 | 0.831 | 0.819 | 0.819 | 0.819 |
| **Roche**<br>**NeuroToolKit** | p-tau181 | 0.815 (0.772-0.858) | 1.14 (pg/ml) | 0.177 | 0.775 | 0.746 | 0.760 | 0.744 | 0.777 |
|  | Aβ42/Aβ40 | 0.775 (0.728-0.823) | 0.126 | 0.197 | 0.864 | 0.627 | 0.742 | 0.688 | 0.829 |
|  | GFAP | 0.758 (0.710-0.806) | 0.113 (ng/ml) | 0.203 | 0.728 | 0.682 | 0.704 | 0.685 | 0.725 |
|  | NfL | 0.677 (0.624-0.729) | 4.29 (pg/ml) | 0.228 | 0.592 | 0.682 | 0.638 | 0.638 | 0.637 |
| **Quanterix**<br>**Neurology**<br>**4-Plex** | GFAP | 0.756 (0.704-0.807) | 205 (pg/ml) | 0.204 | 0.597 | 0.814 | 0.713 | 0.736 | 0.700 |
|  | p-tau181 | 0.747 (0.694-0.799) | 20.5 (pg/ml) | 0.211 | 0.717 | 0.694 | 0.705 | 0.671 | 0.738 |
|  | Aβ42/Aβ40 | 0.734 (0.681-0.787) | 0.0582 | 0.214 | 0.730 | 0.689 | 0.708 | 0.671 | 0.746 |
|  | NfL | 0.670 (0.613-0.727) | 21.7 (pg/ml) | 0.229 | 0.686 | 0.574 | 0.626 | 0.583 | 0.677 |

Compared to the C2N PrecivityAD2 combined model, the ALZpath Quanterix p-tau217 and Janssen LucentAD Quanterix p-tau217 models had significantly lower performance (ALZpath Quanterix AUC 0.885 [0.851-0.920]; Janssen LucentAD Quanterix AUC 0.882 [0.848-0.916]), p=0.0006 for both versus the C2N PrecivityAD2 combined model (**Table 3**). For the Roche NeuroToolKit and Quanterix Neurology 4-Plex assays, models including p-tau181, Aβ42/Aβ40, NfL, and GFAP were most accurate (Roche NeuroToolKit AUC 0.873 [0.838-0.909]; Quanterix Neurology 4-Plex AUC 0.808 [0.763-0.854]), but were not significantly better than models including only p-tau181 and Aβ42/Aβ40 (Roche NeuroToolKit AUC 0.871 [0.834-0.907], p=0.67 for the two compared to the four analyte model; Quanterix Neurology 4-Plex AUC 0.782 [0.733-0.830], p=0.083 for the two compared to the four analyte model), (**Table 3**).

In a sub-cohort of 122 individuals with CSF data available, the classification accuracies of the plasma biomarker models for amyloid PET status were directly compared to CSF Elecsys p-tau181/Aβ42. In this smaller sample, the classification accuracy of the CSF Elecsys p-tau181/Aβ42 model (AUC 0.915 [0.864-0.967]) was not significantly different from the classification accuracies of the top performing plasma models for all six companies: C2N PrecivityAD2 %p-tau217 + Aβ42/Aβ40 (0.907 [0.851-0.963], p=0.67); Fujirebio Lumipulse p-tau217 + Aβ42/Aβ40 (AUC 0.897 [0.840-0.955], p=0.58); ALZpath Quanterix p-tau217 (AUC 0.883 [0.822-0.945], p=0.22); Janssen LucentAD Quanterix p-tau217 (AUC 0.858 [0.792-0.924], p=0.12); Roche NeuroToolKit p-tau181 + Aβ42/Aβ40 + GFAP + NfL (AUC 0.834 [0.760-0.908], p=0.098); Quanterix Neurology 4-Plex p-tau181 + Aβ42/Aβ40 + GFAP + NfL (AUC 0.822 [0.740-0.905], p=0.13), (**Tables S6 and S7**).

The classification accuracies of plasma biomarker models of amyloid PET status as defined by Centiloid >20 were evaluated in the sub-cohort of 192 individuals with cognitive impairment (CDR>0), which is relevant to clinical practice in which biomarker testing is limited to patients with cognitive impairment (**Table S8**). Classification accuracies were slightly higher than in the full cohort: the C2N PrecivityAD2 %p-tau217 and Aβ42/Aβ40 combined model had an AUC of 0.960 [0.934-0.986] and the Fujirebio Lumipulse p-tau217 and Aβ42/Aβ40 combined model had an AUC of 0.952 [0.925-0.980]. The PPA, NPA, and overall percent accuracies for the C2N PrecivityAD2 and Fujirebio Lumipulse p-tau217 measures were approximately 90% (**Table S9**). Conversely, classification accuracies were lower in the sub-cohort of 200 individuals who were cognitively unimpaired (CDR=0), which is relevant to clinical trials of preventative treatments (**Tables S10 and S11**). The C2N PrecivityAD2 combined model had an AUC of 0.890 [0.839-0.942] and was statistically superior to the best performing models from other companies including the Fujirebio Lumipulse combined model, which had an AUC of 0.840 [0.782-0.897], p=0.024. If a more abnormal cut-off value were chosen for amyloid PET positivity (Centiloid >37), the classification accuracies were similar or slightly higher for p-tau217 and p-tau181 models and were similar or slightly lower for Aβ42/Aβ40 models in the full cohort (**Tables S12 and S13**), cognitively impaired sub-cohort (**Tables S14 and S15**), and cognitively unimpaired sub-cohort (**Tables S16 and S17**).

Cut-offs for amyloid status that were previously published or reported by the company were examined in the full cohort (**Table S18**). Notably, these cut-offs were developed in different cohorts and may have been determined using different standards for amyloid positivity than in the current study (amyloid PET Centiloid >20). For C2N PrecivityAD2 %p-tau217, the cut-off of 4.2% for distinguishing amyloid status that was previously reported [20] was similar to the cut-off determined in the current study (4.06%), with similar accuracy for classifying amyloid PET status (86.7% for Meyer cut-off, 87.8% for current study cut-off). For ALZpath Quanterix p-tau217, the cut-off of 0.42 pg/ml that was previously reported [21] was also similar to the cut-off determined in the current study (0.444 pg/ml), with similar accuracy (82.4% for Ashton cut-off, 83.7% for current study cut-off).

### Correlations between amyloid PET Centiloid and plasma biomarkers

Correlations between amyloid PET Centiloid and individual plasma biomarkers were examined with Spearman correlations because of expected non-linearity in these relationships (**Figure 4**). In the full cohort, amyloid PET Centiloid had the strongest correlation with the C2N PrecivityAD2 %p-tau217 (unadjusted Spearman ρ=0.771 [95% confidence intervals 0.735 to 0.804]), which was superior to correlations of analytes from other companies including with Fujirebio Lumipulse p-tau217 (ρ=0.715 [0.663 to 0.760], p=0.011), ALZpath Quanterix p-tau217 (ρ=0.702 [0.649 to 0.748], p=0.0040), and Janssen LucentAD Quanterix p-tau217 (ρ=0.699 [0.647 to 0.748], p=0.0029) (**Figure 5**, **Table 5**). Correlations between amyloid PET Centiloid and p-tau181 were much lower as compared to C2N PrecivityAD2 %p-tau217: Roche NeuroToolKit p-tau181 (ρ=0.547 [0.472 to 0.618], p<0.0001) and Quanterix Neurology 4-Plex p-tau181 (ρ=0.438 (0.351 to 0.512), p<0.0001). Correlations between amyloid PET Centiloid and Aβ42/Aβ40 were also lower, ranging from a correlation of ρ=-0.531 (−0.595 to −0.455) for the Roche NeuroToolKit to ρ=-0.404 (−0.486 to −0.316) for the Quanterix Neurology 4-Plex.

**Figure 4.**
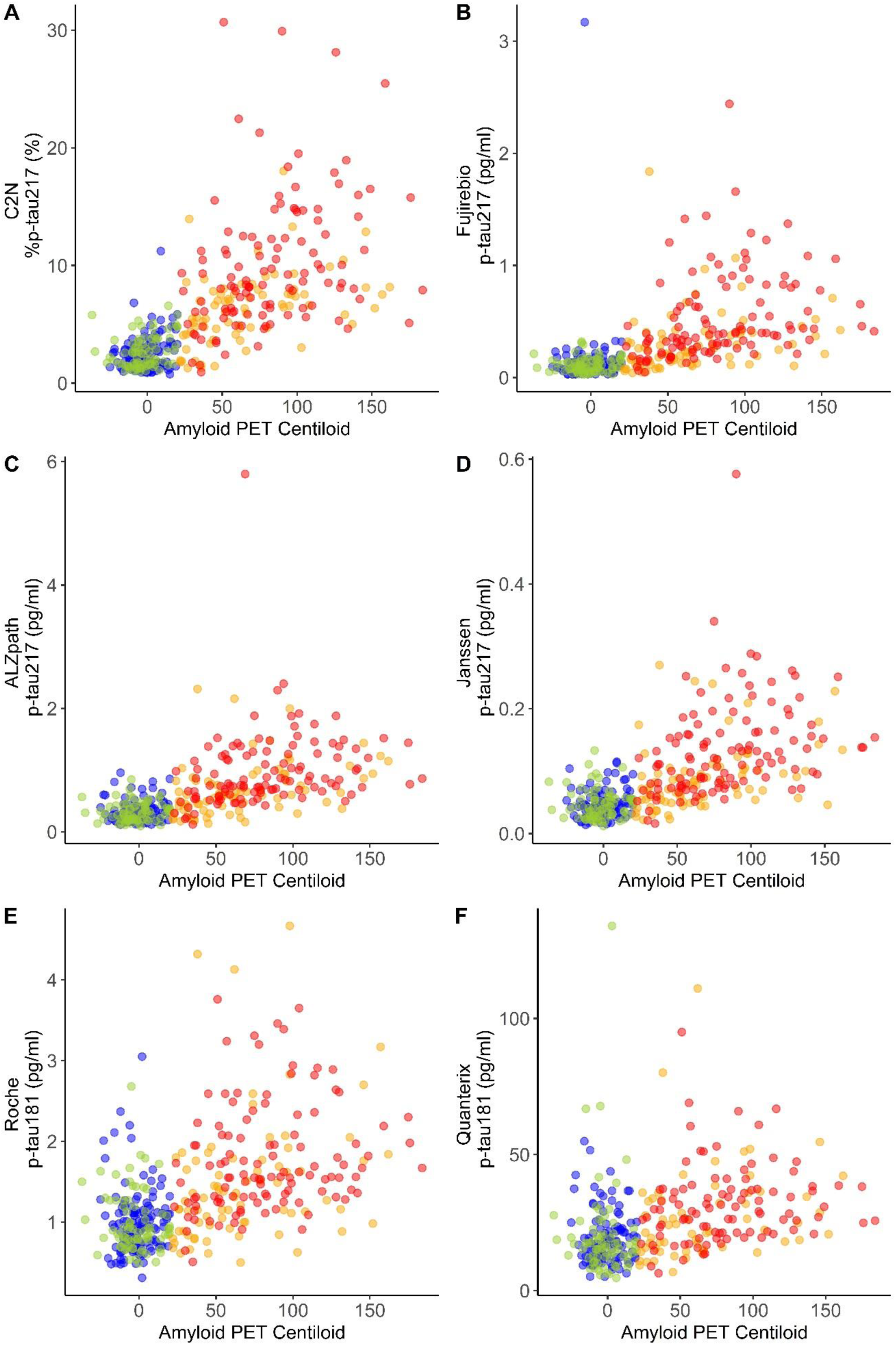
Scatterplots of plasma biomarker measures and amyloid PET Centiloid. The best performing plasma biomarker measure from each company is shown. All individuals (n=392) had plasma biomarker measures for the C2N PrecivityAD2, Fujirebio Lumipulse, ALZpath Quanterix, Janssen LucentAD Quanterix, and Roche NeuroToolKit assays. A sub-cohort (n=342) additionally had plasma biomarker measures for the Quanterix Neurology 4-Plex assays. Point colors represent the following: blue, cognitively unimpaired and amyloid PET negative; green, cognitively impaired and amyloid PET negative; orange, cognitively unimpaired and amyloid PET positive; red, cognitively impaired and amyloid PET positive.

**Figure 5.**
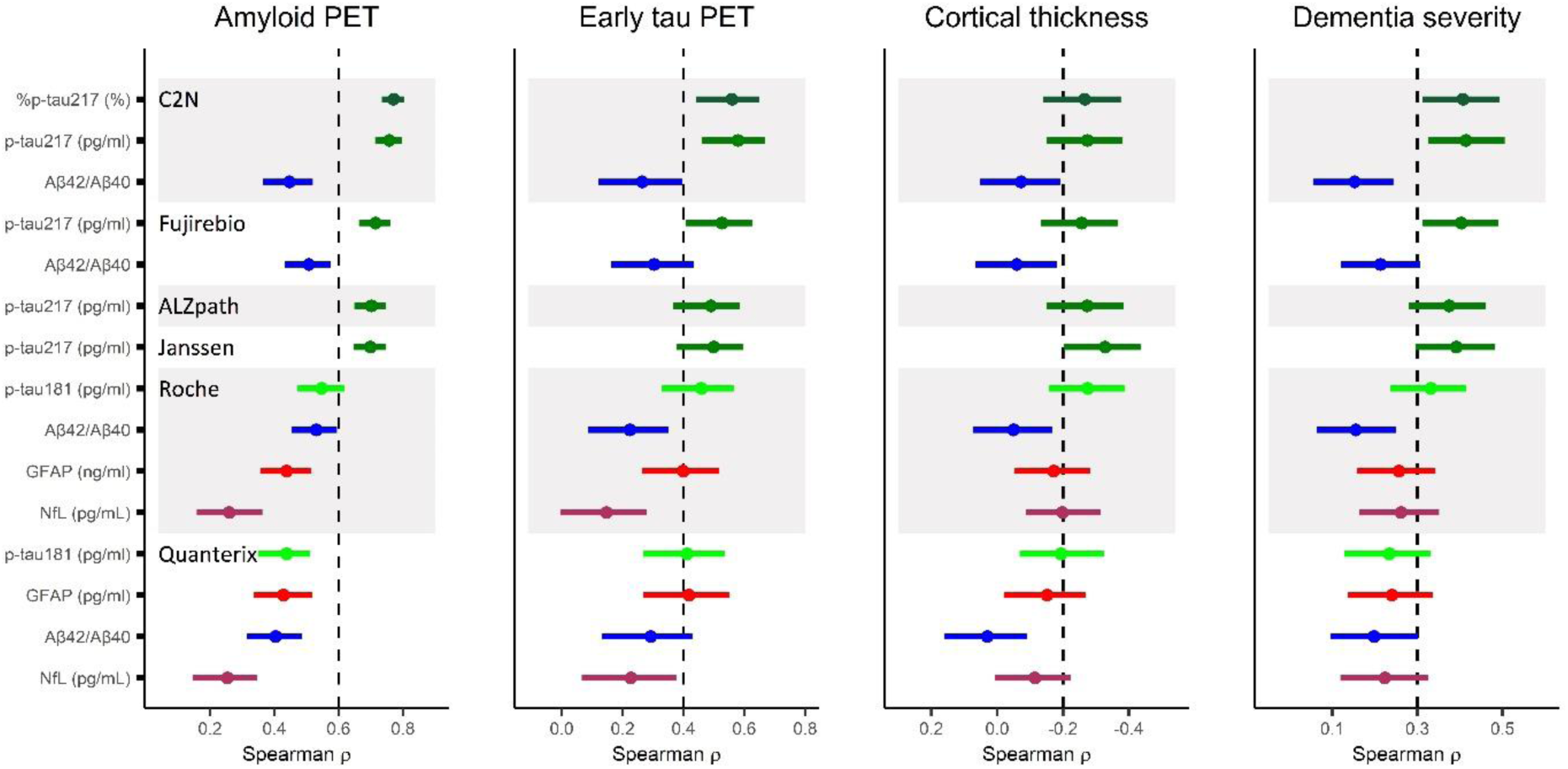
Correlations between individual plasma biomarker measures and key outcomes in the full cohort. All individuals (n=392) had plasma biomarker measures for the C2N PrecivityAD2, Fujirebio Lumipulse, ALZpath Quanterix, Janssen LucentAD Quanterix, and Roche NeuroToolKit assays. A sub-cohort (n=342) additionally had plasma biomarker measures for the Quanterix Neurology 4-Plex assays. The unadjusted Spearman correlation point estimate (midpoint) and 95% confidence intervals with plasma biomarker measures is shown for amyloid PET Centiloid, the early tau PET measure, cortical thickness, and dementia severity by the Clinical Dementia Rating Sum of Boxes. The dashed vertical reference lines are arbitrary and presented to aide in visual interpretation.

**Table 5.** Correlations between individual plasma biomarker measures and amyloid PET Centiloid in the full cohort. The unadjusted Spearman correlation with amyloid PET Centiloid, and the partial Spearman correlation adjusting for age, sex, and *APOE* genotype, are shown with 95% confidence intervals. Correlations between the top performing measure and other measures were compared by bootstrapping.

| Company | Measure | Unadjusted for covariates |  | Adjusted for covariates |  |
| --- | --- | --- | --- | --- | --- |
|  |  | Spearman rho (95% CI) | p= | Partial Spearman rho (95% CI) | p= |
| C2N<br>PrecivityAD2 | %p-tau217 (%) | 0.771 (0.735 to 0.804) | REFERENCE | 0.762 (0.721 to 0.797) | REFERENCE |
|  | p-tau217 (pg/ml) | 0.757 (0.713 to 0.798) | 0.25 | 0.754 (0.707 to 0.793) | 0.50 |
| | Aβ42/Aβ40 | -0.447 (-0.521 to -0.372) | <0.0001 | -0.425 (-0.499 to -0.349) | <0.0001 |
| Fujirebio<br>Lumipulse | p-tau217 (pg/ml) | 0.715 (0.663 to 0.760) | 0.011 | 0.706 (0.652 to 0.755) | 0.017 |
| | Aβ42/Aβ40 | -0.507 (-0.574 to -0.433) | <0.0001 | -0.491 (-0.561 to -0.411) | <0.0001 |
| ALZpath<br>Quanterix | p-tau217 (pg/ml) | 0.702 (0.649 to 0.748) | 0.0040 | 0.704 (0.645 to 0.755) | 0.013 |
| Janssen<br>LucentAD<br>Quanterix | p-tau217 (pg/ml) | 0.699 (0.647 to 0.748) | 0.0029 | 0.693 (0.634 to 0.745) | 0.0038 |
| Roche<br>NeuroToolKit | p-tau181 (pg/ml) | 0.547 (0.472 to 0.618) | <0.0001 | 0.545 (0.463 to 0.621) | <0.0001 |
| | Aβ42/Aβ40 | -0.531 (-0.595 to -0.455) | <0.0001 | -0.513 (-0.590 to -0.434) | <0.0001 |
|  | GFAP (ng/ml) | 0.438 (0.356 to 0.516) | <0.0001 | 0.426 (0.341 to 0.505) | <0.0001 |
|  | NfL (pg/mL) | 0.260 (0.159 to 0.363) | <0.0001 | 0.241 (0.144 to 0.333) | <0.0001 |
| Quanterix<br>Neurology<br>4-Plex | p-tau181 (pg/ml) | 0.438 (0.351 to 0.512) | <0.0001 | 0.427 (0.335 to 0.510) | <0.0001 |
|  | GFAP (pg/ml) | 0.428 (0.335 to 0.516) | <0.0001 | 0.400 (0.305 to 0.486) | <0.0001 |
| | Aβ42/Aβ40 | -0.404 (-0.486 to -0.316) | <0.0001 | -0.363 (-0.454 to -0.266) | <0.0001 |
|  | NfL (pg/mL) | 0.254 (0.146 to 0.351) | <0.0001 | 0.217 (0.112 to 0.328) | <0.0001 |

In the cognitively impaired sub-cohort, the correlation of C2N PrecivityAD2 %p-tau217 with amyloid PET Centiloid was ρ=0.792 [0.744 to 0.832], which was not significantly different from Fujirebio Lumipulse p-tau217 (ρ=0.792 [0.746 to 0.829], p=0.97), ALZpath Quanterix p-tau217 (ρ=0.768 [0.714 to 0.807], p=0.35), or Janssen LucentAD Quanterix p-tau217 (ρ=0.744 [0.680 to 0.792], 0.087), (**Figure 6, Table S19)**. Correlations between amyloid PET Centiloid and p-tau181 remained lower as compared to C2N PrecivityAD2 %p-tau217: Roche NeuroToolKit p-tau181 (ρ=0.613 [0.519 to 0.685], p<0.0001) and Quanterix Neurology 4-Plex p-tau181 (ρ=0.512 [0.401 to 0.606], p<0.0001). Correlations between amyloid PET Centiloid and Aβ42/Aβ40 were also low, ranging from a correlation of ρ=-0.521 (−0.636 to −0.412) for the Roche NeuroToolKit to ρ=-0.384 (−0.506 to −0.248) for the Quanterix Neurology 4-Plex.

**Figure 6.**
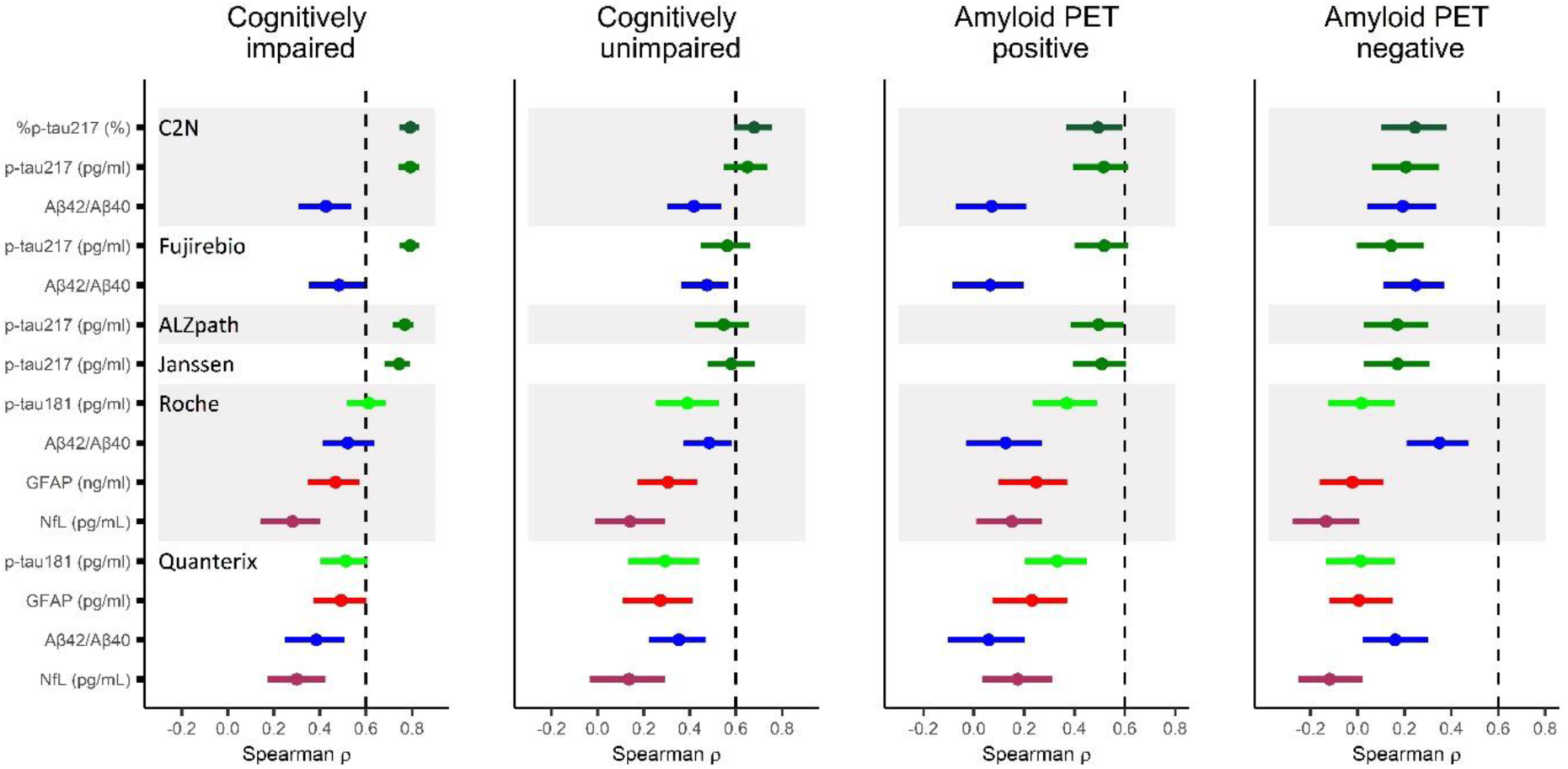
Correlations between individual plasma biomarker measures and amyloid PET Centiloid in sub-cohorts. All individuals (n=392) had plasma biomarker measures for the C2N PrecivityAD2, Fujirebio Lumipulse, ALZpath Quanterix, Janssen LucentAD Quanterix, and Roche NeuroToolKit assays. A sub-cohort (n=342) additionally had plasma biomarker measures for the Quanterix Neurology 4-Plex assays. The unadjusted Spearman correlation point estimate (midpoint) and 95% confidence intervals are shown for amyloid PET Centiloid, the early tau PET measure, cortical thickness, and dementia severity by the Clinical Dementia Rating Sum of Boxes. The dashed vertical reference lines are arbitrary and presented to aide in visual interpretation.

In the cognitively unimpaired sub-cohort, C2N PrecivityAD2 %p-tau217 had a significantly higher correlation with amyloid PET Centiloid (ρ=0.679 [0.592 to 0.756]) compared to the other p-tau217 measures: Janssen LucentAD Quanterix (ρ=0.580 [0.477 to 0.683), p=0.030), Fujirebio Lumipulse (ρ=0.563 [0.447 to 0.661], p=0.019) and ALZpath Quanterix (ρ=0.546 [0.422 to 0.658], p=0.0078), (**Figure 6, Table S20**). Correlations between amyloid PET Centiloid and p-tau181 dropped much lower in this sub-cohort as compared to C2N PrecivityAD2 %p-tau217: Roche NeuroToolKit p-tau181 (ρ=0.389 [0.253 to 0.525], p<0.0001) and Quanterix Neurology 4-Plex p-tau181 (ρ=0.293 [0.132 to 0.440], p<0.0001). In contrast, correlations between amyloid PET Centiloid and Aβ42/Aβ40 remained relatively similar, ranging from a correlation of ρ=-0.484 (−0.584 to −0.371) for the Roche NeuroToolKit to ρ=-0.352 (−0.470 to −0.221) for the Quanterix Neurology 4-Plex.

In a sub-cohort with amyloid PET >20 Centiloids, Fujirebio Lumipulse p-tau217 had a correlation of ρ=0.518 [0.401 to 0.615] with amyloid PET Centiloid, which was not statistically different than correlations for the C2N PrecivityAD2 %p-tau217 (ρ=0.494 [0.366 to 0.593], p=0.70), Janssen LucentAD Quanterix p-tau217 (ρ=0.509 [0.393 to 0.606], p=0.83), or ALZpath Quanterix p-tau217 (ρ=0.496 [0.384 to 0.596], p=0.62) (**Figure 6, Table S21**). In a sub-cohort with amyloid PET ≤20 Centiloids, very low levels of amyloid burden had the highest correlations with Roche NeuroToolKit Aβ42/Aβ40 (ρ=-0.349 [-0.472 to −0.211]), Fujirebio Lumipulse Aβ42/Aβ40 (ρ=-0.248 [-0.372 to −0.109]), and C2N PrecivityAD2 %p-tau217 (ρ=0.245 [0.102 to 0.379]), (**Figure 6, Table S22**).

### Classification of tau PET status by plasma biomarkers

The classification accuracies of individual and combined plasma biomarkers for early tau PET status as defined by flortaucipir signal in the mesial temporal meta-ROI (entorhinal, parahippocampus and amygdala) were evaluated using logistic regression models in the full cohort (**Figure 3, Tables S23 and S24**). The highest performing models from each company included either p-tau217 or p-tau181 and were not significantly different from one another: C2N PrecivityAD2 %p-tau217 (AUC 0.888 [0.836-0.940]), Fujirebio Lumipulse p-tau217 (AUC 0.859 [0.802-0.916]), ALZpath Quanterix p-tau217 (AUC 0.850 [0.788-0.912]), Janssen LucentAD Quanterix p-tau217 (AUC 0.846 [0.780-0.912]), Roche NeuroToolKit p-tau181 + Aβ42/Aβ40 + GFAP + NfL (AUC 0.874 [0.823-0.925]), and Quanterix Neurology 4-Plex p-tau181 + Aβ42/Aβ40 + GFAP + NfL (AUC 0.874 [0.819-0.930]). Notably, the Roche NeuroToolKit p-tau181 (0.823 [0.755-0.891]) and Quanterix Neurology 4-Plex p-tau181 (0.803 [0.724-0.883]) models were not significantly different from the models combining all four analytes (p=0.095 for difference in Roche NeuroToolKit 2 versus 4 analyte models; p=0.10 for difference in Quanterix Neurology 4-Plex 2 versus 4 analyte models).

In the cognitively impaired sub-cohort, classification accuracies for early tau PET status were slightly higher, including for the C2N PrecivityAD2 %p-tau217 (AUC 0.921 [0.857-0.985]) and Janssen LucentAD Quanterix p-tau217 (AUC 0.921 [0.855-0.986]) models, and again there were no significant differences between the top performing models from each company (**Tables S25 and S26**). In the cognitively unimpaired sub-cohort, classification accuracies were lower for the C2N PrecivityAD2 %p-tau217 (AUC 0.805 [0.707-0.903]) and Janssen LucentAD Quanterix p-tau217 (AUC 0.677 [0.522-0.833]) models (**Table S27**). In the sub-cohort with amyloid PET >20 Centiloids, classification accuracies for models of early tau PET status were lower than in the full cohort, including for the C2N PrecivityAD2 %p-tau217 (AUC 0.792 [0.694-0.891]) and Janssen LucentAD Quanterix p-tau217 (AUC 0.744 [0.638-0.850]) models, (**Tables S28 and S29)**.

The classification accuracies of individual and combined plasma biomarkers for late tau PET status as defined by flortaucipir signal in the temporal-parietal meta-ROI (entorhinal, parahippocampus, amygdala, fusiform, inferior and middle temporal gyri) were evaluated using logistic regression models in the full cohort (**Table 30**). The highest performing models were C2N PrecivityAD2 %p-tau217 + Aβ42/Aβ40 (AUC 0.901 [0.823-0.980]), Janssen LucentAD Quanterix p-tau217 (AUC 0.879 [0.796-0.961]), and Fujirebio Lumipulse p-tau217 (AUC 0.878 [0.797-0.958]). As with early tau PET, classification accuracies for late tau PET status were slightly higher in the cognitively impaired sub-cohort (**Table 31**), and lower in the cognitively unimpaired sub-cohort (**Table S32**) and sub-cohort with amyloid PET > 20 Centiloids (**Table 33**).

### Correlations between tau PET and plasma biomarkers

Correlations between the early tau PET measure and individual plasma biomarkers were examined with Spearman correlations because of expected non-linearity in these relationships (**Figure 7**). In the full cohort, the early tau PET measure had the highest correlation with C2N PrecivityAD2 p-tau217 (ρ=0.579 [0.468 to 0.666]), which was not different from correlations with Fujirebio Lumipulse p-tau217 (ρ=0.526 [0.416 to 0.620], p=0.11), but was stronger than correlations with Janssen LucentAD Quanterix p-tau217 (ρ=0.499 [0.384 to 0.594], p=0.022), ALZpath Quanterix p-tau217 (ρ=0.490 [0.371 to 0.586], p=0.011), Roche NeuroToolKit p-tau181 (ρ=0.459 [0.341 to 0.568], p=0.0061) and Quanterix Neurology 4-Plex p-tau181 (0.411 [0.266 to 0.535], p=0.013), (**Figure 5, Table S34**).

**Figure 7.**
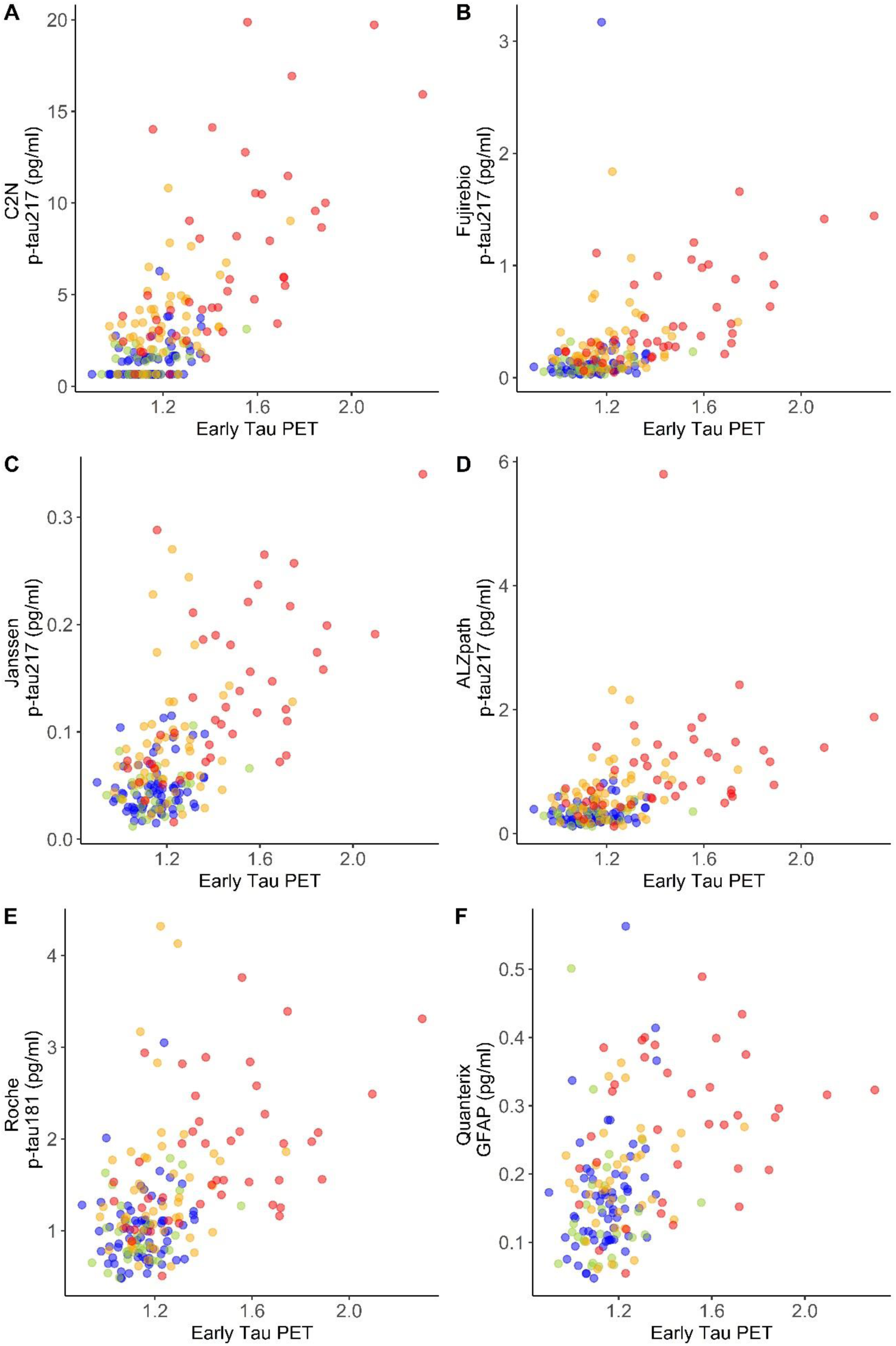
Scatterplots of plasma biomarker measures and the early tau PET measure. The best performing plasma biomarker measure from each company is shown. All individuals (n=392) had plasma biomarker measures for the C2N PrecivityAD2, Fujirebio Lumipulse, ALZpath Quanterix, Janssen LucentAD Quanterix, and Roche NeuroToolKit assays. A sub-cohort (n=342) additionally had plasma biomarker measures for the Quanterix Neurology 4-Plex assays. Point colors represent the following: blue, cognitively unimpaired and amyloid PET negative; green, cognitively impaired and amyloid PET negative; orange, cognitively unimpaired and amyloid PET positive; red, cognitively impaired and amyloid PET positive.

In the cognitively impaired sub-cohort, correlations with the early tau PET measure were stronger, including for C2N PrecivityAD2 p-tau217 (ρ=0.755 [0.626 to 0.833]) and Fujirebio Lumipulse p-tau217 (ρ=0.726 [0.591 to 0.818]), (**Table S35**). In the cognitively unimpaired cohort, correlations with the early tau PET measure were much weaker including for C2N PrecivityAD2 p-tau217 (ρ=0.401 [0.249 to 0.548]) and Fujirebio Lumipulse p-tau217 (ρ=0.324 [0.154 to 0.473]), (**Table S36**). In the sub-cohort with amyloid PET >20 Centiloids, correlations with the early tau PET measure were lower, but still strong for C2N PrecivityAD2 p-tau217 (ρ=0.600 [0.450 to 0.719]), Fujirebio Lumipulse p-tau217 (ρ=0.543 [0.395 to 0.663]), and Janssen LucentAD Quanterix p-tau217 (ρ=0.545 [0.391 to 0.676]), (**Table S37**).

Correlations were also examined for the late tau PET measure. In the full cohort, the late tau PET measure had similar patterns of correlations as the early tau PET measure but weaker correlations: the highest correlations were with C2N PrecivityAD2 p-tau217 (ρ=0.377 [0.243 to 0.507]) and Fujirebio Lumipulse p-tau217 (ρ=0.327 [0.187 to 0.456]), (**Table S38**). Patterns of correlations for the late tau PET measure were also similar but weaker than with the early tau PET measure in the cognitively impaired sub-cohort (**Table S39**) and the sub-cohort with amyloid PET >20 Centiloids (**Table S40**). Correlations between the late tau PET measures and plasma biomarkers in the cognitively unimpaired sub-cohort were not significant.

### Classification of cortical thickness status by plasma biomarkers

Cortical thickness status was evaluated using logistic regression models in the full cohort (**Figure 3, Tables S41 and S42**). As with tau PET, the highest performing models from each company included either p-tau217 or p-tau181 and did not vary significantly from one another in classification accuracy: Janssen LucentAD Quanterix p-tau217 (AUC 0.849 [0.765-0.933]), Fujirebio Lumipulse p-tau217 (AUC 0.841 [0.765-0.916]), C2N PrecivityAD2 p-tau217 + Aβ42/Aβ40 (AUC 0.840 [0.765-0.914]), Roche NeuroToolKit p-tau181 + Aβ42/Aβ40 + GFAP + NfL (AUC 0.838 [0.774-0.902]), ALZpath Quanterix p-tau217 (AUC 0.833 [0.755-0.912]), and Quanterix Neurology 4-Plex p-tau181 + Aβ42/Aβ40 + NfL (AUC 0.783 [0.707-0.860]). In contrast, classification accuracy of cortical thickness status was lower for plasma NfL than covariates alone (AUC 0.758 [0.664-0.852] for covariates; AUC 0.743 [0.663-0.823] for Roche NeuroToolKit NfL; AUC 0.745 [0.662-0.829] for Quanterix Neurology 4-Plex NfL). Classification accuracies were very similar in the cognitively impaired sub-cohort (**Tables S43 and S44**) and similar but slightly lower in the sub-cohort with amyloid PET >20 Centiloids (**Tables S45 and S46**), with no significant differences between the top performing models for each company.

### Correlations between cortical thickness and plasma biomarkers

Correlations between the cortical thickness measure and individual plasma biomarkers were examined with Spearman correlations. In the full cohort, the cortical thickness measure had the highest correlation with Janssen LucentAD Quanterix p-tau217 (ρ=-0.327 [-0.436 to −0.217]), which was not different from correlations with Roche NeuroToolKit p-tau181 (ρ=-0.275 [-0.386 to −0.165], p=0.10) or C2N PrecivityAD2 p-tau217 (ρ=-0.274 [-0.388 to −0.159], p=0.79), but was higher than correlations with ALZpath Quanterix p-tau217 (ρ=-0.273 [-0.383 to −0.166], p=0.016), Fujirebio Lumipulse p-tau217 (ρ=-0.256 [-0.369 to −0.138], p=0.018), or Quanterix Neurology 4-Plex p-tau181 (ρ=-0.194 [-0.317 to −0.066], p=0.010), (**Table S47**). Correlations between the cortical thickness measure and NfL were inferior to Janssen LucentAD Quanterix p-tau217 (ρ=-0.198 [-0.314 to −0.085] for Roche NeuroToolKit NfL, p=0.041; ρ=-0.114 (−0.234 to 0.000) for Quanterix Neurology 4-Plex NfL, p=0.0008).

In the cognitively impaired sub-cohort, correlations with cortical thickness measure were higher, including for Janssen LucentAD Quanterix p-tau217 (ρ=-0.425 [-0.568 to −0.277]), Fujirebio Lumipulse p-tau217 (ρ=-0.384 [-0.520 to −0.222], and C2N PrecivityAD2 p-tau217 (ρ=-0.356 [-0.497 to −0.196]), (**Figure 5, Table S48**). Correlations were even higher in the sub-cohort with amyloid PET >20 Centiloids, with the strongest correlations for Janssen LucentAD Quanterix p-tau217 (ρ=-0.527 [-0.650 to −0.386]), Fujirebio Lumipulse p-tau217 (ρ=-0.477 [-0.608 to −0.334]), and C2N PrecivityAD2 p-tau217 (ρ=-0.458 [-0.585 to −0.317]), (**Table S49**).

### Classification of cognitive status by plasma biomarkers

Cognitive status (CDR>0, cognitively impaired; CDR=0, cognitively normal) was evaluated using logistic regression models in the full cohort (**Figure 3, Tables S50 and S51**). The classification accuracy of all models was relatively low. The highest performing model was C2N PrecivityAD2 p-tau217 + Aβ42/Aβ40 (AUC 0.680 [0.627-0.733]), but it was not significantly different from the top performing models from the other companies: Janssen LucentAD Quanterix p-tau217 (AUC 0.670 [0.615-0.724], p=0.46); Fujirebio Lumipulse p-tau217 (AUC 0.667 [0.613-0.721], p=0.40); ALZpath Quanterix p-tau217 (AUC 0.664 [0.610-0.718], p=0.40); Roche NeuroToolKit p-tau181 + Aβ42/Aβ40 + NfL (AUC 0.648 [0.593-0.703], p=0.40); and Quanterix Neurology 4-Plex p-tau181 + Aβ42/Aβ40 + GFAP + NfL (AUC 0.635 [0.576-0.695], p=0.40). Classification accuracies were slightly higher in the sub-cohort with amyloid PET >20 Centiloids, but there was still no distinction between the top performing models (**Tables S52 and S53**).

### Correlations between dementia symptoms and plasma biomarkers

Correlations between dementia symptoms as measured by the CDR-SB and individual plasma biomarkers were examined with Spearman correlations. In the full cohort, dementia symptoms had the highest correlation with C2N PrecivityAD2 p-tau217 (ρ=0.414 [0.325 to 0.505]), which was not different from correlations with Fujirebio Lumipulse p-tau217 (ρ=0.403 [0.311 to 0.490], p=0.65), Janssen LucentAD Quanterix p-tau217 (ρ=0.391 [0.295 to 0.482], p=0.34), or ALZpath Quanterix p-tau217 (ρ=0.374 [0.280 to 0.461], p=0.074), but was stronger than correlations with Roche NeuroToolKit p-tau181 (ρ=0.331 [0.237 to 0.415], p=0.0038), Roche NeuroToolKit NfL (ρ=0.261 [0.164 to 0.351], p=0.0022), Quanterix Neurology 4-Plex p-tau181 (ρ=0.234 [0.131 to 0.332], p<0.0001), and Quanterix Neurology 4-Plex NfL (ρ=0.224 [0.118 to 0.321], p=0.0006), (**Table S54**). In the sub-cohort with amyloid PET >20 Centiloids, correlations were slightly higher and the findings were similar, with stronger correlations between dementia symptoms and p-tau217 as compared to p-tau181, (**Table S55**).

## 4. Discussion

Head-to-head studies are essential to compare the performance of plasma assays and analytes. For the current study, the FNIH Biomarker Consortium sought to compare several leading commercial assays for plasma Aβ42/Aβ40 and tau phosphorylated at positions 217 and 181 (p-tau217 and p-tau181). Additionally, two assays for glial fibrillary acidic protein (GFAP) and neurofilament light (NfL) were included. Based on CSF biomarker groupings described in the 2018 NIA-AA AT(N) research framework for AD [7], it might have been expected that plasma Aβ42/Aβ40 would best classify amyloid PET, plasma p-tau217 and p-tau181 would be most strongly associated with tau PET, and plasma NfL would have highest correlations with cortical thickness and cognitive impairment. However, the major finding of this study was that regardless of the specific assay, plasma p-tau217 had the highest classification accuracies and correlations with all key outcomes studied—amyloid PET, tau PET, cortical thickness, and cognitive impairment. Thus, it is possible that research studies and clinical trials could rely on a single or limited number of blood measures that are informative regarding multiple aspects of AD.

The models with the highest classification accuracy for amyloid PET status included both a plasma p-tau217 measure (%p-tau217 for PrecivityAD2, p-tau217 for Fujirebio Lumipulse) and Aβ42/Aβ40, although these models were not statistically superior to the p-tau217 measure alone in either the full cohort or the cognitively impaired sub-cohort. Notably, both the C2N PrecivityAD2 and Fujirebio Lumipulse p-tau217 assays had limited sensitivity, resulting in imputed values for samples with low values, but this did not affect the performance of the assays in classification accuracy for amyloid PET status, which depends on distinguishing individuals with higher levels of p-tau217. Additionally, C2N PrecivityAD2 %p-tau217 had the highest continuous correlation with amyloid PET burden, suggesting that very low levels of %p-tau217 correspond to very low levels of amyloid burden. In the cognitively unimpaired cohort, models including %p-tau217 had superior performance in classifying amyloid PET status and %p-tau217 had the highest correlations with amyloid PET burden. Notably, plasma Aβ42/Aβ40 had much lower classification accuracies for amyloid PET status, likely because of the low magnitude of change in Aβ42/Aβ40 levels associated with amyloid pathology (13-10%) that may be related to non-brain derived Aβ in the plasma [36, 37]. However, in a sub-cohort with very low amyloid burden (≤20 Centiloids), amyloid PET had the highest correlations with Aβ42/Aβ40, suggesting that Aβ42/Aβ40 is altered very early in the course of amyloid deposition.

In addition to being superior classifiers of amyloid PET status, plasma p-tau217 assays also had high classification accuracies for early tau PET status, but these accuracies were lower when only amyloid PET positive individuals were considered. Notably, levels of plasma p-tau217 increase starting very early in the AD disease course in response to amyloid pathology at a time when neurofibrillary tangles are not yet present in high amounts [38, 39], indicating that plasma p-tau217 levels do not only reflect neurofibrillary tangles. High plasma p-tau217 levels likely mainly reflect significant amyloid burden, which is often associated with tau pathology [40]. Novel fluid biomarkers of AD, such as MTBR-tau243 [41] and N-terminal containing tau fragments (NTA-tau) [42], may be less associated with amyloid pathology and more reflective of tau pathology.

While plasma NfL is often used to represent neurodegeneration, measures of p-tau217 had stronger associations with cortical thickness and cognitive impairment, although even these associations were relatively weak. Novel fluid biomarkers of AD such as MTBR-tau243 and NTA-tau are reported to be more strongly associated with brain structure and cognition [41, 42]. However, structural brain changes and cognitive impairment, including in patients with AD pathology, may be complicated by the presence of other conditions such as cerebrovascular disease and non-AD neurodegenerative diseases.

Therefore, plasma biomarkers of non-AD causes of cognitive impairment, including those that reflect cerebrovascular disease, TDP-43, and α-synuclein pathology [1], could potentially be very helpful in improving prediction of brain structure and cognition.

As AD clinical trials and treatments continue to evolve, it is likely that different treatments will be targeted to individuals based on disease stage [43]. For example, the TRAILBLAZER-ALZ2 study enrolled participants with amyloid PET Centiloids≥37 and tau PET positivity, and individuals with low/medium tau had a greater slowing of disease progression compared to individuals with high tau [4]. Plasma biomarkers including p-tau217 will be very helpful in identifying cognitively impaired patients who are likely to benefit from specific treatments [40], and in the future may also be helpful in identifying cognitively unimpaired individuals who are likely to progress to dementia and are candidates for preventative treatments [44, 45]. Further, it may be possible to stage individuals with plasma biomarkers to understand their location on a disease progression model [38, 46].

Beyond their use in research and clinical trials, AD blood tests are increasingly being used in clinical practice to assist clinicians in determining whether AD pathology is present and may be causing or contributing to a patient’s cognitive impairment [3, 47, 48]. AD biomarker testing is now of greater importance in clinical practice to determine if patients may be candidates for new FDA-approved AD-specific treatments [49]. The assays examined in this study are either currently available for clinical use (C2N PrecivityAD2, Fujirebio Lumipulse, ALZpath Quanterix, and Janssen LucentAD Quanterix) or are in the process of being developed for clinical use (Roche NeuroToolKit and Quanterix Neurology 4-Plex). In cognitively impaired individuals, all four p-tau217 assays had high classification accuracies (AUC >0.92) for amyloid PET status. The top-performing C2N PrecivityAD2 and Fujirebio Lumipulse measures that included p-tau217 had approximately 90% PPA, NPA, and accuracy for amyloid PET status, which is similar to the performance of FDA-approved CSF tests and is the recommended minimum performance of clinical AD blood tests (Schindler et al., *Nature Rev Neurol in press)*; [50]. It will be important for clinicians to use only accurate and well-validated tests for clinical care, as these test results have major implications for patients and their families. If using these blood tests as part of patient care, clinicians should consider the predictive value of the test after integration with all other clinical findings (Schindler et al., *Nature Rev Neurol in press)*.

Limitations of the current study include a relatively small sample size for some sub-cohorts. In particular, the CSF sub-cohort (n=122) did not provide adequate power to definitively determine whether the blood tests had equivalent performance to CSF tests in classification of amyloid PET status. The combinations of biomarkers tested were pre-specified and limited to a single company, with the expectation that the company would offer their analytes as a panel, and did not evaluate combinations that might be possible across companies such as p-tau217 combined with NfL or GFAP. Additionally, various methods for deriving cut-offs, such as the two cut-off approach, were not examined [1]. Further, although analyses adjusting for covariates (age, sex, *APOE* genotype) were provided for review, the focus of this study was on unadjusted models, and the results of the adjusted models were not examined in this study. Previous studies have reported that some medical conditions have major effects on plasma biomarker concentrations, including p-tau217 and p-tau181 [51, 52], while other biomarker measures such as Aβ42/Aβ40 and the %p-tau217 may be less affected [52, 53]. Additional studies are needed that carefully examine the effects of age, sex, *APOE* genotype, medical conditions, medications, race, and social determinants of health on plasma biomarker levels and test performance. Another limitation of the current study is very low racial diversity; only 14 of 392 participants (4%) self-identified as Black. In recent years the ADNI has made major efforts to increase the diversity of their cohort [54], but because this project only included individuals with longitudinal data, it did not benefit from more recent improvements in cohort diversity.

While this cohort and the FNIH Biomarkers Consortium project was designed to evaluate the longitudinal trajectories of AD blood biomarkers in relation to amyloid PET, the current study was a cross-sectional head-to-head comparison of leading commercially available blood tests. We expect these analyses will be helpful to other investigators and to clinicians as they seek to understand the important but increasingly complex topic of AD blood tests. Given that these plasma biomarker data will be widely available to investigators via ADNI, the current study is intended to provide a starting point for many future studies that will analyze these data in much greater detail. It is our hope that these data will accelerate development of additional diagnostics and treatments for Alzheimer disease.

## Data Availability

Data used in the preparation of this article may be obtained from the Alzheimers Disease Neuroimaging Initiative (ADNI) database (adni.loni.usc.edu)

https://adni.loni.usc.edu/

## Acknowledgements

The results of the study represent results of the Foundation for the National Institutes of Health (FNIH) Biomarkers Consortium “Biomarkers Consortium, Plasma Aβ and Phosphorylated Tau as Predictors of Amyloid and Tau Positivity in Alzheimer’s Disease” project. The study was made possible through the scientific and financial support of industry, academic, patient advocacy, and governmental partners. We are grateful for the contributions of the following project team members: Anthony Bannon (AbbVie), Michael Baratta (Takeda), Janaky Coomaraswamy (Takeda), Jeff Dage (Indiana University), Iwona Dobler (Takeda), Lei Du-Cuny (AbbVie), Kyle Ferber (Biogen), John Hsiao (NIA), Hartmuth Kolb (formerly with Johnson and Johnson Innovative Medicine), Emily Meyers (Alzheimer’s Association), Yulia Mordashova (AbbVie), William Potter, Maria Quinton (AbbVie), Dave Raunig (Takeda), Erin Rosenbaugh (FNIH), Carrie Rubel (Biogen), Ziad Saad (Johnson and Johnson Innovative Medicine), Patricia Saletti (Alzheimer’s Drug Discovery Foundation), Suzanne Schindler (Washington University in St. Louis), Leslie Shaw (University of Pennsylvania), Gallen Triana-Baltzer (Johnson and Johnson Innovative Medicine), Christopher Weber (Alzheimer’s Association), Henrik Zetterberg (University of Gothenburg). Funding partners of the project include AbbVie Inc., Alzheimer’s Association®, Diagnostics Accelerator at the Alzheimer’s Drug Discovery Foundation, Biogen, Janssen Research & Development, LLC, and Takeda Pharmaceutical Company Limited. Private-sector funding for the study was managed by the Foundation for the National Institutes of Health.

We recognize C_2_N Diagnostics, Fujirebio Diagnostics with the Indiana University National Centralized Repository for Alzheimer’s Disease and Related Dementias Biomarker Assay Laboratory (NCRAD-BAL), Quanterix, and Roche Diagnostics with the University of Gothenburg for performing the plasma biomarker analysis in this study. Additionally, we are grateful to Drs. Nicholas Ashton and Henrik Zetterberg and the University of Gothenburg for generating the plasma endogenous QC pools confirmed high or low for pTau217. The NCRAD-BAL is supported by a cooperative agreement grant (U24 AG021886) awarded to NCRAD by the National Institute on Aging. Elecsys β-amyloid (1–42) CSF, Elecsys Phospho-Tau (181P) CSF and Elecsys Total-Tau CSF assays are approved for clinical use. COBAS and ELECSYS are trademarks of Roche. All other product names and trademarks are the property of their respective owners. The NeuroToolKit is a panel of exploratory prototype assays designed to robustly evaluate biomarkers associated with key pathologic events characteristic of AD and other neurological disorders, used for research purposes only and not approved for clinical use (Roche Diagnostics International Ltd, Rotkreuz, Switzerland).

Finally, we would like to acknowledge ADNI for the plasma samples and ADNI participant data analyzed in this study. Data collection and sharing for the Alzheimer’s Disease Neuroimaging Initiative (ADNI) is funded by the National Institute on Aging (National Institutes of Health Grant U19 AG024904). The grantee organization is the Northern California Institute for Research and Education. In the past, ADNI has also received funding from the National Institute of Biomedical Imaging and Bioengineering, the Canadian Institutes of Health Research, and private sector contributions through the Foundation for the National Institutes of Health (FNIH) including generous contributions from the following: AbbVie, Alzheimer’s Association; Alzheimer’s Drug Discovery Foundation; Araclon Biotech; BioClinica, Inc.; Biogen; Bristol-Myers Squibb Company; CereSpir, Inc.; Cogstate; Eisai Inc.; Elan Pharmaceuticals, Inc.; Eli Lilly and Company; EuroImmun; F. Hoffmann-La Roche Ltd and its affiliated company Genentech, Inc.; Fujirebio; GE Healthcare; IXICO Ltd.; Janssen Alzheimer Immunotherapy Research & Development, LLC.; Johnson & Johnson Pharmaceutical Research & Development LLC.; Lumosity; Lundbeck; Merck & Co., Inc.; Meso Scale Diagnostics, LLC.; NeuroRx Research; Neurotrack Technologies; Novartis Pharmaceuticals Corporation; Pfizer Inc.; Piramal Imaging; Servier; Takeda Pharmaceutical Company Limited; and Transition Therapeutics.

## Collaborators

Data used in the preparation of this article were obtained from the ADNI database (adni.loni.usc.edu). As such, the investigators within the ADNI contributed to the design and implementation of ADNI and/or provided data but did not participate in the analysis or writing of this report. A complete listing of ADNI investigators can be found at: http://adni.loni.usc.edu/wp-content/uploads/how_to_apply/ADNI_Acknowledgement_List.pdf

## Consent Statement

Written informed consent was obtained from each ADNI participant or their legally authorized representative.

## References

[1] Hansson O, Blennow K, Zetterberg H, Dage J. Blood biomarkers for Alzheimer’s disease in clinical practice and trials. Nat Aging. 2023;3:506–19.

[2] Hampel H, Hu Y, Cummings J, Mattke S, Iwatsubo T, Nakamura A, et al. Blood-based biomarkers for Alzheimer’s disease: Current state and future use in a transformed global healthcare landscape. Neuron. 2023.

[3] Hansson O, Edelmayer RM, Boxer AL, Carrillo MC, Mielke MM, Rabinovici GD, et al. The Alzheimer’s Association appropriate use recommendations for blood biomarkers in Alzheimer’s disease. Alzheimer’s & dementia : the journal of the Alzheimer’s Association. 2022.

[4] Sims JR, Zimmer JA, Evans CD, Lu M, Ardayfio P, Sparks J, et al. Donanemab in Early Symptomatic Alzheimer Disease: The TRAILBLAZER-ALZ 2 Randomized Clinical Trial. JAMA. 2023.

[5] van Dyck CH, Swanson CJ, Aisen P, Bateman RJ, Chen C, Gee M, et al. Lecanemab in Early Alzheimer’s Disease. N Engl J Med. 2022.

[6] Schindler SE, Atri A. The role of cerebrospinal fluid and other biomarker modalities in the Alzheimer’s disease diagnostic revolution. Nat Aging. 2023;3:460–2.

[7] Jack CR, Jr., Bennett DA, Blennow K, Carrillo MC, Dunn B, Haeberlein SB, et al. NIA-AA Research Framework: Toward a biological definition of Alzheimer’s disease. Alzheimer’s & dementia : the journal of the Alzheimer’s Association. 2018;14:535–62.

[8] Zicha S, Bateman RJ, Shaw LM, Zetterberg H, Bannon AW, Horton WA, et al. Comparative analytical performance of multiple plasma Abeta42 and Abeta40 assays and their ability to predict positron emission tomography amyloid positivity. Alzheimer’s & dementia : the journal of the Alzheimer’s Association. 2022.

[9] Janelidze S, Bali D, Ashton NJ, Barthelemy NR, Vanbrabant J, Stoops E, et al. Head-to-head comparison of 10 plasma phospho-tau assays in prodromal Alzheimer’s disease. Brain. 2022.

[10] Janelidze S, Teunissen CE, Zetterberg H, Allue JA, Sarasa L, Eichenlaub U, et al. Head-to-Head Comparison of 8 Plasma Amyloid-beta 42/40 Assays in Alzheimer Disease. JAMA Neurol. 2021.

[11] Brand AL, Lawler PE, Bollinger JG, Li Y, Schindler SE, Li M, et al. The performance of plasma amyloid beta measurements in identifying amyloid plaques in Alzheimer’s disease: a literature review. Alzheimers Res Ther. 2022;14:195.

[12] Ashton NJ, Puig-Pijoan A, Mila-Aloma M, Fernandez-Lebrero A, Garcia-Escobar G, Gonzalez-Ortiz F, et al. Plasma and CSF biomarkers in a memory clinic: Head-to-head comparison of phosphorylated tau immunoassays. Alzheimer’s & dementia : the journal of the Alzheimer’s Association. 2023;19:1913–24.

[13] Mielke MM, Hagen CE, Xu J, Chai X, Vemuri P, Lowe VJ, et al. Plasma phospho-tau181 increases with Alzheimer’s disease clinical severity and is associated with tau- and amyloid-positron emission tomography. Alzheimer’s & dementia : the journal of the Alzheimer’s Association. 2018;14:989–97.

[14] Pereira JB, Janelidze S, Smith R, Mattsson-Carlgren N, Palmqvist S, Teunissen CE, et al. Plasma GFAP is an early marker of amyloid-beta but not tau pathology in Alzheimer’s disease. Brain. 2021;144:3505–16.

[15] Aschenbrenner AJ, Li Y, Henson RL, Volluz K, Hassenstab J, Verghese P, et al. Comparison of plasma and CSF biomarkers in predicting cognitive decline. Ann Clin Transl Neurol. 2022;9:1739–51.

[16] Morris JC. The Clinical Dementia Rating (CDR): current version and scoring rules. Neurology. 1993;43:2412–4.

[17] Dore V, Doecke JD, Saad ZS, Triana-Baltzer G, Slemmon R, Krishnadas N, et al. Plasma p217+tau versus NAV4694 amyloid and MK6240 tau PET across the Alzheimer’s continuum. Alzheimer’s & dementia. 2022;14:e12307.

[18] Therriault J, Servaes S, Tissot C, Rahmouni N, Ashton NJ, Benedet AL, et al. Equivalence of plasma p-tau217 with cerebrospinal fluid in the diagnosis of Alzheimer’s disease. Alzheimer’s & dementia : the journal of the Alzheimer’s Association. 2023;19:4967–77.

[19] Triana-Baltzer G, Moughadam S, Slemmon R, Van Kolen K, Theunis C, Mercken M, et al. Development and validation of a high-sensitivity assay for measuring p217+tau in plasma. Alzheimer’s & dementia. 2021;13:e12204.

[20] Meyer MR, Kirmess KM, Eastwood S, Wente-Roth TL, Irvin F, Holubasch MS, et al. Clinical validation of the PrecivityAD2 blood test: A mass spectrometry-based test with algorithm combining %p-tau217 and Abeta42/40 ratio to identify presence of brain amyloid. Alzheimer’s & dementia : the journal of the Alzheimer’s Association. 2024.

[21] Ashton NJ, Brum WS, Di Molfetta G, Benedet AL, Arslan B, Jonaitis E, et al. Diagnostic Accuracy of a Plasma Phosphorylated Tau 217 Immunoassay for Alzheimer Disease Pathology. JAMA Neurol. 2024;81:255–63.

[22] Therriault J, Ashton NJ, Pola I, Triana-Baltzer G, Brum WS, Di Molfetta G, et al. Comparison of two plasma p-tau217 assays to detect and monitor Alzheimer’s pathology. EBioMedicine. 2024;102:105046.

[23] Benedet AL, Mila-Aloma M, Vrillon A, Ashton NJ, Pascoal TA, Lussier F, et al. Differences Between Plasma and Cerebrospinal Fluid Glial Fibrillary Acidic Protein Levels Across the Alzheimer Disease Continuum. JAMA Neurol. 2021;78:1471–83.

[24] Mattsson N, Cullen NC, Andreasson U, Zetterberg H, Blennow K. Association Between Longitudinal Plasma Neurofilament Light and Neurodegeneration in Patients With Alzheimer Disease. JAMA Neurol. 2019;76:791–9.

[25] Kirmess KM, Meyer MR, Holubasch MS, Knapik SS, Hu Y, Jackson EN, et al. The PrecivityAD test: Accurate and reliable LC-MS/MS assays for quantifying plasma amyloid beta 40 and 42 and apolipoprotein E proteotype for the assessment of brain amyloidosis. Clinica chimica acta; international journal of clinical chemistry. 2021;519:267–75.

[26] Jagust WJ, Landau SM, Koeppe RA, Reiman EM, Chen K, Mathis CA, et al. The Alzheimer’s Disease Neuroimaging Initiative 2 PET Core: 2015. Alzheimer’s & dementia : the journal of the Alzheimer’s Association. 2015;11:757–71.

[27] Joshi AD, Pontecorvo MJ, Clark CM, Carpenter AP, Jennings DL, Sadowsky CH, et al. Performance characteristics of amyloid PET with florbetapir F 18 in patients with alzheimer’s disease and cognitively normal subjects. Journal of nuclear medicine : official publication, Society of Nuclear Medicine. 2012;53:378–84.

[28] Royse SK, Minhas DS, Lopresti BJ, Murphy A, Ward T, Koeppe RA, et al. Validation of amyloid PET positivity thresholds in centiloids: a multisite PET study approach. Alzheimers Res Ther. 2021;13:99.

[29] Villemagne VL, Leuzy A, Bohorquez SS, Bullich S, Shimada H, Rowe CC, et al. CenTauR: Toward a universal scale and masks for standardizing tau imaging studies. Alzheimer’s & dementia. 2023;15:e12454.

[30] Diedrichsen J. A spatially unbiased atlas template of the human cerebellum. NeuroImage. 2006;33:127–38.

[31] Jack CR, Jr., Wiste HJ, Weigand SD, Therneau TM, Lowe VJ, Knopman DS, et al. Defining imaging biomarker cut points for brain aging and Alzheimer’s disease. Alzheimer’s & dementia : the journal of the Alzheimer’s Association. 2017;13:205–16.

[32] Pomponio R, Erus G, Habes M, Doshi J, Srinivasan D, Mamourian E, et al. Harmonization of large MRI datasets for the analysis of brain imaging patterns throughout the lifespan. NeuroImage. 2020;208:116450.

[33] Redelmeier DA, Bloch DA, Hickam DH. Assessing predictive accuracy: how to compare Brier scores. J Clin Epidemiol. 1991;44:1141–6.

[34] DeLong ER, DeLong DM, Clarke-Pearson DL. Comparing the areas under two or more correlated receiver operating characteristic curves: a nonparametric approach. Biometrics. 1988;44:837–45.

[35] Benjamini Y, Hochberg Y. Controlling the False Discovery Rate - a Practical and Powerful Approach to Multiple Testing. J R Stat Soc B. 1995;57:289–300.

[36] Wisch JK, Gordon BA, Boerwinkle AH, Luckett PH, Bollinger JG, Ovod V, et al. Predicting continuous amyloid PET values with CSF and plasma Abeta42/Abeta40. Alzheimer’s & dementia. 2023;15:e12405.

[37] Schindler SE, Bollinger JG, Ovod V, Mawuenyega KG, Li Y, Gordon BA, et al. High-precision plasma beta-amyloid 42/40 predicts current and future brain amyloidosis. Neurology. 2019;93:e1647–e59.

[38] Li Y, Yen D, Hendrix RD, Gordon BA, Dlamini S, Barthelemy NR, et al. Timing of Biomarker Changes in Sporadic Alzheimer’s Disease in Estimated Years from Symptom Onset. Annals of neurology. 2024.

[39] Palmqvist S, Janelidze S, Quiroz YT, Zetterberg H, Lopera F, Stomrud E, et al. Discriminative Accuracy of Plasma Phospho-tau217 for Alzheimer Disease vs Other Neurodegenerative Disorders. JAMA. 2020;324:772–81.

[40] Mattsson-Carlgren N, Collij LE, Stomrud E, Pichet Binette A, Ossenkoppele R, Smith R, et al. Plasma Biomarker Strategy for Selecting Patients With Alzheimer Disease for Antiamyloid Immunotherapies. JAMA Neurol. 2024;81:69–78.

[41] Horie K, Salvado G, Barthelemy NR, Janelidze S, Li Y, He Y, et al. CSF MTBR-tau243 is a specific biomarker of tau tangle pathology in Alzheimer’s disease. Nat Med. 2023;29:1954–63.

[42] Lantero-Rodriguez J, Salvado G, Snellman A, Montoliu-Gaya L, Brum WS, Benedet AL, et al. Plasma N-terminal containing tau fragments (NTA-tau): a biomarker of tau deposition in Alzheimer’s Disease. Mol Neurodegener. 2024;19:19.

[43] Therriault J, Schindler SE, Salvado G, Pascoal TA, Benedet AL, Ashton NJ, et al. Biomarker-based staging of Alzheimer disease: rationale and clinical applications. Nature reviews Neurology. 2024.

[44] Mattsson-Carlgren N, Salvado G, Ashton NJ, Tideman P, Stomrud E, Zetterberg H, et al. Prediction of Longitudinal Cognitive Decline in Preclinical Alzheimer Disease Using Plasma Biomarkers. JAMA Neurol. 2023;80:360–9.

[45] Rissman RA, Langford O, Raman R, Donohue MC, Abdel-Latif S, Meyer MR, et al. Plasma Abeta42/Abeta40 and phospho-tau217 concentration ratios increase the accuracy of amyloid PET classification in preclinical Alzheimer’s disease. Alzheimer’s & dementia : the journal of the Alzheimer’s Association. 2023.

[46] Salvado G, Horie K, Barthelemy NR, Vogel JW, Pichet Binette A, Chen CD, et al. Disease staging of Alzheimer’s disease using a CSF-based biomarker model. Nat Aging. 2024.

[47] Johnson KA, Minoshima S, Bohnen NI, Donohoe KJ, Foster NL, Herscovitch P, et al. Appropriate use criteria for amyloid PET: a report of the Amyloid Imaging Task Force, the Society of Nuclear Medicine and Molecular Imaging, and the Alzheimer’s Association. Alzheimer’s & dementia : the journal of the Alzheimer’s Association. 2013;9:e-1–16.

[48] Shaw LM, Arias J, Blennow K, Galasko D, Molinuevo JL, Salloway S, et al. Appropriate use criteria for lumbar puncture and cerebrospinal fluid testing in the diagnosis of Alzheimer’s disease. Alzheimer’s & dementia : the journal of the Alzheimer’s Association. 2018;14:1505–21.

[49] Cummings J, Apostolova L, Rabinovici GD, Atri A, Aisen P, Greenberg S, et al. Lecanemab: Appropriate Use Recommendations. J Prev Alzheimers Dis. 2023;10:362–77.

[50] Revised Criteria for Diagnosis and Staging of Alzheimer’s Disease: Alzheimer’s Association Workgroup.

[51] Mielke MM, Dage JL, Frank RD, Algeciras-Schimnich A, Knopman DS, Lowe VJ, et al. Performance of plasma phosphorylated tau 181 and 217 in the community. Nat Med. 2022.

[52] Syrjanen JA, Campbell MR, Algeciras-Schimnich A, Vemuri P, Graff-Radford J, Machulda MM, et al. Associations of amyloid and neurodegeneration plasma biomarkers with comorbidities. Alzheimer’s & dementia : the journal of the Alzheimer’s Association. 2021.

[53] Janelidze S, Barthelemy NR, He Y, Bateman RJ, Hansson O. Mitigating the Associations of Kidney Dysfunction With Blood Biomarkers of Alzheimer Disease by Using Phosphorylated Tau to Total Tau Ratios. JAMA Neurol. 2023;80:516–22.

[54] Weiner MW, Veitch DP, Miller MJ, Aisen PS, Albala B, Beckett LA, et al. Increasing participant diversity in AD research: Plans for digital screening, blood testing, and a community-engaged approach in the Alzheimer’s Disease Neuroimaging Initiative 4. Alzheimer’s & dementia : the journal of the Alzheimer’s Association. 2023;19:307–17.

