## Supplementary Materials for "Head-to-head comparison of leading blood tests for Alzheimer’s disease pathology"

#### Table of Contents

Table S1. Characteristics of cognitively impaired sub-cohort. 4
Table S2. Characteristics of cognitively unimpaired sub-cohort. 5
Table S3. Characteristics of tau PET sub-cohort. 6
Table S4. Characteristics of structural brain MRI sub-cohort. 7
Table S5. Characteristics of CSF sub-cohort 8
Table S6. Classification accuracies of individual and combined plasma biomarker analytes for amyloid PET status defined by Centiloid>20 or ≤20 in the CSF sub-cohort 9
Table S7. Classification accuracies of individual plasma biomarker analytes for amyloid PET status defined by Centiloid>20 or ≤20 in the CSF sub-cohort 10
Table S8. Classification accuracies of individual and combined plasma biomarker analytes for amyloid PET status defined by Centiloid>20 or ≤20 in the cognitively impaired sub-cohort 11
Table S9. Classification accuracies of individual plasma biomarker analytes for amyloid PET status defined by Centiloid>20 or ≤20 in the cognitively impaired sub-cohort 12
Table S10. Classification accuracies of individual and combined plasma biomarker analytes for amyloid PET status defined by Centiloid>20 or ≤20 in the cognitively unimpaired sub-cohort 13
Table S11. Classification accuracies of individual plasma biomarker analytes for amyloid PET status defined by Centiloid>20 or ≤20 in the cognitively unimpaired sub-cohort 14
Table S12. Classification accuracies of individual and combined plasma biomarker analytes for amyloid PET status defined by Centiloid >37 or ≤37 in the full cohort 15
Table S13. Classification accuracies of individual plasma biomarker analytes for amyloid PET status defined by Centiloid>37 or ≤37 in the full cohort 16
Table S14. Classification accuracies of individual and combined plasma biomarker analytes for amyloid PET status defined by Centiloid>37 or ≤37 in the cognitively impaired sub-cohort 17
Table S15. Classification accuracies of individual plasma biomarker analytes for amyloid PET status defined by Centiloid>37 or ≤37 in the cognitively impaired sub-cohort 18
Table S16. Classification accuracies of individual and combined plasma biomarker analytes for amyloid PET status defined by Centiloid>37 or ≤37 in the cognitively unimpaired sub-cohort 19
Table S17. Classification accuracies of individual plasma biomarker analytes for amyloid PET status defined by Centiloid>37 or ≤37 in the cognitively unimpaired sub-cohort 20
Table S18. Accuracy of published and/or recommended cut-offs for amyloid PET status defined by Centiloid>20 or ≤20 in the full cohort 21
Table S19. Correlations between individual plasma biomarker analytes and amyloid PET Centiloid in the cognitively impaired sub-cohort 22
Table S20. Correlations between individual plasma biomarker analytes and amyloid PET Centiloid in the cognitively unimpaired sub-cohort 23
Table S21. Correlations between individual plasma biomarker analytes and amyloid PET Centiloid in the sub-cohort with amyloid PET >20 Centiloids 24
Table S22. Correlations between individual plasma biomarker analytes and amyloid PET Centiloid in the sub-cohort with amyloid PET ≤20 Centiloids 25
Table S23. Classification accuracies of individual and combined plasma biomarker analytes for early tau PET status in the full cohort 26
Table S24. Classification accuracies of individual plasma biomarker analytes for early tau PET status in the full cohort 27

**Table S1. Characteristics of cognitively impaired sub-cohort.** Continuous values are presented as the median with the interquartile range. The significance of differences by amyloid PET status were evaluated with Wilcoxon's rank-sum tests for continuous variables and Chi square or Fisher's exact tests for categorical variables. All tests were two sided and not adjusted for covariates or multiple comparisons. Please see the methods and **Appendix A** for definitions of early tau PET, late tau PET, and cortical thickness.

| Characteristic | Sub-cohort | | Amyloid PET negative | | Amyloid PET positive | | |
|---|---|---|---|---|---|---|---|
| | n= | Values | n= | Values | n= | Values | p= |
| **Demographics** | | | | | | | |
| Age (Years) | 192 | 78 (72.3-83.3) | 77 | 74.7 (70.5-81.5) | 115 | 80.5 (74-83.5) | 0.02 |
| Sex (% Female) | 192 | 90, 46.88% | 77 | 33, 42.86% | 115 | 57, 49.57% | 0.4 |
| *APOE Genotype (22/23/24/33/34/44)* | 192 | 0/13/3/105/59/12 | 77 | 0/10/0/52/12/3 | 115 | 0/3/3/53/47/9 | <0.0001 |
| *APOE ε4 carrier status (n, % carrier)* | 192 | 74, 38.54% | 77 | 15, 19.48% | 115 | 59, 51.3% | <0.0001 |
| Years of Education | 192 | 16 (14-18) | 77 | 16 (14-18) | 115 | 16 (14-18) | 0.32 |
| CDR (0/0.5/1+) | 192 | 4/182/6 | 77 | 2/71/4 | 115 | 2/111/2 | 0.36 |
| CDR Sum of Boxes | 192 | 0/148/44 | 77 | 0/74/3 | 115 | 0/74/41 | <0.0001 |
| Race (Black/White/Other) | 192 | 1.5 (1-3.5) | 77 | 1 (0.5-1.5) | 115 | 2.5 (1-5) | <0.0001 |
| Plasma collection to CDR (years) | 192 | 0 (0-0.02) | 77 | 0 (0-0.02) | 115 | 0 (0-0.02) | 0.71 |
| **Amyloid PET** | | | | | | | |
| Amyloid PET Centiloid | 192 | 46 (1-86) | 77 | -4 (-9-5) | 115 | 78 (56-104) | <0.0001 |
| Plasma collection to amyloid PET (years) | 192 | 0.02 (0-0.04) | 77 | 0.02 (0-0.05) | 115 | 0.02 (0-0.04) | 0.79 |
| **Tau PET** | | | | | | | |
| Early tau PET | 72 | 1.26 (1.12-1.51) | 28 | 1.12 (1.05-1.19) | 44 | 1.41 (1.23-1.65) | <0.0001 |
| Early tau PET Positivity | 72 | 29, 40% | 28 | 1, 3.6% | 44 | 28, 64% | <0.0001 |
| Late tau PET | 72 | 1.17 (1.09-1.34) | 28 | 1.12 (1.04-1.18) | 44 | 1.22 (1.11-1.51) | 0.0003 |
| Late tau PET Positivity | 72 | 23, 32% | 28 | 2, 7.1% | 44 | 21, 48% | 0.0003 |
| Plasma collection to tau PET (years) | 72 | 0.03 (0-0.13) | 28 | 0.04 (0-0.10) | 44 | 0.03 (0-0.16) | 0.73 |
| **Cortical thickness** | | | | | | | |
| Cortical thickness | 129 | 2.75 (2.62-2.84) | 58 | 2.8 (2.71-2.85) | 71 | 2.68 (2.54-2.81) | 0.0002 |
| Cortical thickness positivity | 129 | 23, 17% | 58 | 2, 3.5% | 71 | 21, 30% | 0.0001 |
| Plasma collection to MRI (years) | 129 | 0.01 (0-0.05) | 58 | 0.01 (0-0.05) | 71 | 0.01 (0-0.05) | 0.72 |
| **CSF Elecsys** | | | | | | | |
| CSF p-tau181/Aβ42 | 63 | 0.0432 (0.0181-0.0551) | 22 | 0.0134 (0.0106-0.0215) | 41 | 0.0519 (0.0422-0.0652) | <0.0001 |
| CSF p-tau181 (pg/ml) | 63 | 26.3 (19.9-36.6) | 22 | 19.4 (15.9-24.9) | 41 | 30.3 (22.9-38.7) | 0.0003 |
| CSF Aβ42 (pg/ml) | 64 | 767 (561-1130) | 23 | 1180 (1030-1500) | 41 | 628 (455-803) | <0.0001 |
| Plasma collection to CSF (years) | 63 | 0 (0-0.02) | 22 | 0 (0-0.03) | 41 | 0 (0-0.01) | 0.22 |

**Table S2. Characteristics of cognitively unimpaired sub-cohort.** Continuous values are presented as the median with the interquartile range. The significance of differences by amyloid PET status were evaluated with Wilcoxon's rank-sum tests for continuous variables and Chi square or Fisher's exact tests for categorical variables. All tests were two sided and not adjusted for covariates or multiple comparisons. Please see the methods and **Appendix A** for definitions of early tau PET, late tau PET, and cortical thickness.

| Characteristic | Sub-cohort | | Amyloid PET negative | | Amyloid PET positive | | |
|---|---|---|---|---|---|---|---|
| | n= | Values | n= | Values | n= | Values | p= |
| **Demographics** | | | | | | | |
| Age (Years) | 200 | 78.1 (73.1-83.2) | 124 | 76.6 (72.4-81.9) | 76 | 78.6 (74.8-84.4) | 0.05 |
| Sex (% Female) | 200 | 103, 51.5% | 124 | 65, 52.42% | 76 | 38, 50% | 0.7 |
| *APOE Genotype (22/23/24/33/34/44)* | 200 | 1/22/3/119/49/6 | 124 | 1/19/1/81/19/3 | 76 | 0/3/2/38/30/3 | 0.0009 |
| *APOE ε4 carrier status (n, % carrier)* | 200 | 58, 29% | 124 | 23, 18.55% | 76 | 35, 46.05% | <0.0001 |
| Years of Education | 200 | 17 (15-19) | 124 | 17 (15-19) | 76 | 17 (14-18) | 0.63 |
| CDR (0/0.5/1+) | 200 | 10/184/6 | 124 | 6/113/5 | 76 | 4/71/1 | 0.55 |
| CDR Sum of Boxes | 200 | 0.00274 (0-0.0219) | 124 | 0.00274 (0-0.0246) | 76 | 0.00274 (0-0.0219) | 0.61 |
| Race (Black/White/Other) | 200 | 78.1 (73.1-83.2) | 124 | 76.6 (72.4-81.9) | 76 | 78.6 (74.8-84.4) | 0.05 |
| Plasma collection to CDR (years) | 200 | 103, 51.5% | 124 | 65, 52.42% | 76 | 38, 50% | 0.7 |
| **Amyloid PET** | | | | | | | |
| Amyloid PET Centiloid | 200 | 8 (-4-45) | 124 | -2 (-9-6) | 76 | 59 (37-91) | <0.0001 |
| Plasma collection to amyloid PET (years) | 200 | 0.02 (0-0.04) | 124 | 0.02 (0-0.04) | 76 | 0.01 (0-0.03) | 0.78 |
| **Tau PET** | | | | | | | |
| Early tau PET | 116 | 1.16 (1.09-1.24) | 69 | 1.15 (1.07-1.23) | 47 | 1.19 (1.11-1.29) | 0.02 |
| Early tau PET Positivity | 116 | 11, 9.48% | 69 | 3, 4.35% | 47 | 8, 17.02% | 0.02 |
| Late tau PET | 116 | 1.12 (1.06-1.17) | 69 | 1.11 (1.05-1.15) | 47 | 1.14 (1.07-1.2) | 0.06 |
| Late tau PET Positivity | 116 | 6, 5.17% | 69 | 0, 0% | 47 | 6, 12.77% | 0.002 |
| Plasma collection to tau PET (years) | 116 | 0.02 (0-0.08) | 69 | 0.02 (0-0.07) | 47 | 0.02 (0-0.09) | 0.85 |
| **Cortical thickness** | | | | | | | |
| Cortical thickness | 137 | 2.83 (2.76-2.92) | 86 | 2.84 (2.76-2.91) | 51 | 2.83 (2.76-2.94) | 0.7 |
| Cortical thickness positivity | 137 | 4, 2.92% | 86 | 3, 3.49% | 51 | 1, 1.96% | 0.6 |
| Plasma collection to MRI (years) | 137 | 0.01 (0-0.03) | 86 | 0.01 (0-0.02) | 51 | 0.01 (0-0.04) | 0.05 |
| **CSF Elecsys** | | | | | | | |
| CSF p-tau181/Aβ42 | 59 | 0.0162 (0.0106-0.0318) | 30 | 0.0117 (0.00952-0.0144) | 29 | 0.031 (0.0189-0.0475) | <0.0001 |
| CSF p-tau181 (pg/ml) | 60 | 21.2 (16.6-26) | 30 | 18.6 (15.1-23) | 30 | 24.2 (19.7-29.1) | 0.005 |
| CSF Aβ42 (pg/ml) | 61 | 1040 (713-1690) | 32 | 1560 (1200-1830) | 29 | 734 (622-994) | <0.0001 |
| Plasma collection to CSF (years) | 59 | 0 (0-0) | 30 | 0 (0-0) | 29 | 0 (0-0) | 0.93 |

**Table S3. Characteristics of tau PET sub-cohort.** Continuous values are presented as the median with the interquartile range. The significance of differences by amyloid PET status were evaluated with Wilcoxon's rank-sum tests for continuous variables and Chi square or Fisher's exact tests for categorical variables. All tests were two sided and not adjusted for covariates or multiple comparisons. Please see the methods and **Appendix A** for definitions of early tau PET, late tau PET, and cortical thickness.

| Characteristic | Sub-cohort | | Amyloid PET negative | | Amyloid PET positive | | |
|---|---|---|---|---|---|---|---|
| | n= | Values | n= | Values | n= | Values | p= |
| **Demographics** | | | | | | | |
| Age (Years) | 188 | 78.6 (73.1-83.4) | 97 | 77 (71.3-82.9) | 91 | 79.7 (76-84.4) | 0.038 |
| Sex (% Female) | 188 | 90, 47.9% | 97 | 47, 48.5% | 91 | 43, 47.3% | 0.90 |
| *APOE Genotype (22/23/24/33/34/44)* | 188 | 1/19/1/111/46/10 | 97 | 1/15/0/63/15/3 | 91 | 0/4/1/48/31/7 | 0.0038 |
| *APOE ε4 carrier status (n, % carrier)* | 188 | 57, 30.3% | 97 | 18, 18.6% | 91 | 39, 42.9% | 0.0003 |
| Years of Education | 188 | 16 (14-18) | 97 | 16 (14-19) | 91 | 16 (14-18) | 0.27 |
| CDR (0/0.5/1+) | 188 | 116/55/17 | 97 | 69/28/0 | 91 | 47/27/17 | <0.0001 |
| CDR Sum of Boxes | 188 | 0 (0-1) | 97 | 0 (0-0.5) | 91 | 0 (0-2.5) | 0.0001 |
| Race (Black/White/Other) | 188 | 9/172/7 | 97 | 6/86/5 | 91 | 3/86/2 | 0.35 |
| Plasma collection to CDR (years) | 188 | 0.003 (0.000-0.024) | 97 | 0.006 (0.000-0.027) | 91 | 0.003 (0.000-0.022) | 0.14 |
| **Amyloid PET** | | | | | | | |
| Amyloid PET Centiloid | 188 | 18 (-3-62) | 97 | -3 (-9-6) | 91 | 65 (46-94) | <0.0001 |
| Plasma collection to amyloid PET (years) | 188 | 0.016 (0.003-0.044) | 97 | 0.016 (0.000-0.044) | 91 | 0.016 (0.003-0.041) | 0.85 |
| **Tau PET** | | | | | | | |
| Early tau PET | 188 | 1.18 (1.1-1.3) | 97 | 1.14 (1.06-1.23) | 91 | 1.28 (1.15-1.44) | <0.0001 |
| Early tau PET Positivity | 188 | 40, 21.3% | 97 | 4, 4.1% | 91 | 36, 39.6% | <0.0001 |
| Late tau PET | 188 | 1.13 (1.07-1.2) | 97 | 1.11 (1.05-1.15) | 91 | 1.17 (1.09-1.32) | <0.0001 |
| Late tau PET Positivity | 188 | 29, 15.4% | 97 | 2, 2.1% | 91 | 27, 29.7% | <0.0001 |
| Plasma collection to tau PET (years) | 188 | 0.025 (0.003-0.093) | 97 | 0.025 (0.003-0.079) | 91 | 0.022 (0.003-0.112) | 0.54 |
| **Cortical thickness** | | | | | | | |
| Cortical thickness | 136 | 2.8 (2.69-2.89) | 74 | 2.81 (2.73-2.89) | 62 | 2.79 (2.64-2.88) | 0.20 |
| Cortical thickness positivity | 136 | 13, 9.6% | 74 | 4, 5.4% | 62 | 9, 14.5% | 0.07 |
| Plasma collection to MRI (years) | 136 | 0.014 (0-0.055) | 74 | 0.011 (0-0.038) | 62 | 0.019 (0.005-0.085) | 0.034 |
| **CSF Elecsys** | | | | | | | |
| CSF p-tau181/Aβ42 | 60 | 0.0239 (0.0117-0.0461) | 25 | 0.0117 (0.0094-0.015) | 35 | 0.0366 (0.0242-0.0527) | <0.0001 |
| CSF p-tau181 (pg/ml) | 61 | 23 (16.9-30.1) | 25 | 18 (12.8-24.9) | 36 | 25.1 (19.7-34.9) | 0.0020 |
| CSF Aβ42 (pg/ml) | 62 | 867 (706-1440) | 27 | 1340 (867-1830) | 35 | 744 (647-915) | <0.0001 |
| Plasma collection to CSF (years) | 60 | 0.000 (0.000-0.000) | 25 | 0.000 (0.000-0.000) | 35 | 0.000 (0.000-0.000) | 0.80 |

**Table S4. Characteristics of structural brain MRI sub-cohort.** Continuous values are presented as the median with the interquartile range. The significance of differences by amyloid PET status were evaluated with Wilcoxon's rank-sum tests for continuous variables and Chi square or Fisher's exact tests for categorical variables. All tests were two sided and not adjusted for covariates or multiple comparisons. Please see the methods and **Appendix A** for definitions of early tau PET, late tau PET, and cortical thickness.

| **Characteristic** | **Sub-cohort** | | **Amyloid PET negative** | | **Amyloid PET positive** | | |
|---|---|---|---|---|---|---|---|
| | **n=** | **Values** | **n=** | **Values** | **n=** | **Values** | **p=** |
| **Demographics** | | | | | | | |
| Age (Years) | 266 | 76.6 (72.4-82) | 144 | 75.7 (71.2-81) | 122 | 77.8 (73-83.2) | 0.01 |
| Sex (% Female) | 266 | 124, 46.6% | 144 | 69, 47.9% | 122 | 55, 45.1% | 0.60 |
| *APOE Genotype (22/23/24/33/34/44)* | 266 | 1/23/3/148/77/14 | 144 | 1/20/1/93/25/4 | 122 | 0/3/2/55/52/10 | <0.0001 |
| *APOE ε4 carrier status (n, % carrier)* | 266 | 94, 35.3% | 144 | 30, 20.8% | 122 | 64, 52.5% | <0.0001 |
| Years of Education | 266 | 16 (14-18) | 144 | 16 (15-18) | 122 | 16 (14-18) | 0.45 |
| CDR (0/0.5/1+) | 266 | 137/102/27 | 144 | 86/56/2 | 122 | 51/46/25 | 0.015 |
| CDR Sum of Boxes | 266 | 0 (0-1.5) | 144 | 0 (0-0.5) | 122 | 1 (0-3) | <0.0001 |
| Race (Black/White/Other) | 266 | 10/249/7 | 144 | 7/132/5 | 122 | 3/117/2 | 0.37 |
| Plasma collection to CDR (Years) | 266 | 0.000 (0.000-0.022) | 144 | 0.003 (0.000-0.022) | 122 | 0.000 (0.000-0.019) | 0.096 |
| **Amyloid PET** | | | | | | | |
| Amyloid PET Centiloid | 266 | 16 (-3-67) | 144 | -2 (-9-6) | 122 | 70 (49-99) | <0.0001 |
| Plasma collection to amyloid PET (years) | 266 | 0.016 (0.003-0.041) | 144 | 0.016 (0.003-0.041) | 122 | 0.014 (0.003-0.041) | 0.86 |
| **Tau PET** | | | | | | | |
| Early tau PET | 136 | 1.18 (1.1-1.3) | 74 | 1.16 (1.06-1.23) | 62 | 1.23 (1.16-1.43) | <0.0001 |
| Early tau PET Positivity | 136 | 26, 19.1% | 74 | 3, 4.1% | 62 | 23, 37.1% | <0.0001 |
| Late tau PET | 136 | 1.13 (1.06-1.19) | 74 | 1.12 (1.04-1.16) | 62 | 1.15 (1.08-1.26) | 0.0031 |
| Late tau PET Positivity | 136 | 18, 13.2% | 74 | 2, 2.7% | 62 | 16, 25.8% | <0.0001 |
| Plasma collection to tau PET Interval (years) | 136 | 0.022 (0.003-0.085) | 74 | 0.025 (0.003-0.077) | 62 | 0.022 (0.003-0.110) | 0.51 |
| **Cortical thickness** | | | | | | | |
| Cortical thickness | 266 | 2.8 (2.69-2.89) | 144 | 2.82 (2.75-2.89) | 122 | 2.76 (2.62-2.88) | 0.0028 |
| Cortical thickness positivity | 266 | 27, 10.2% | 144 | 5, 3.5% | 122 | 22, 18.0% | <0.0001 |
| Plasma collection to MRI (years) | 266 | 0.011 (0.000-0.041) | 144 | 0.008 (0.000-0.036) | 122 | 0.014 (0.000-0.047) | 0.053 |
| **CSF Elecsys** | | | | | | | |
| CSF p-tau181/Aβ42 | 91 | 0.0275 (0.0129-0.0491) | 38 | 0.0117 (0.0094-0.0153) | 53 | 0.0436 (0.0284-0.0578) | <0.0001 |
| CSF p-tau181 (pg/ml) | 91 | 24.2 (18.1-30.9) | 38 | 19.2 (15.1-26) | 53 | 26.1 (22-34.9) | 0.00014 |
| CSF Aβ42 (pg/ml) | 94 | 870 (676-1360) | 41 | 1360 (1030-1830) | 53 | 700 (470-915) | <0.0001 |
| Plasma collection to CSF (years) | 91 | 0.000 (0.000-0.003) | 38 | 0.000 (0.000-0.006) | 53 | 0.000 (0.000-0.000) | 0.30 |

**Table S5. Characteristics of CSF sub-cohort.** Continuous values are presented as the median with the interquartile range. The significance of differences by amyloid PET status were evaluated with Wilcoxon's rank-sum tests for continuous variables and Chi square or Fisher's exact tests for categorical variables. All tests were two sided and not adjusted for covariates or multiple comparisons. Please see the methods and **Appendix A** for definitions of early tau PET, late tau PET, and cortical thickness.

| Characteristic | Sub-cohort | | Amyloid PET negative | | Amyloid PET positive | | |
|---|---|---|---|---|---|---|---|
| | n= | Values | n= | Values | n= | Values | p= |
| **Demographics** | | | | | | | |
| Age (Years) | 122 | 76.6 (71.6-82.9) | 52 | 74.2 (70-79.3) | 70 | 78.1 (72.5-84) | 0.0067 |
| Sex (% Female) | 122 | 63, 51.6% | 52 | 28, 53.8% | 70 | 35, 50.0% | 0.70 |
| *APOE Genotype (22/23/24/33/34/44)* | 122 | 0/16/2/63/34/7 | 52 | 0/12/1/32/5/2 | 70 | 0/4/1/31/29/5 | 0.0005 |
| *APOE ε4 carrier status (n, % carrier)* | 122 | 43, 35.2% | 52 | 8, 15.4% | 70 | 35, 50.0% | <0.0001 |
| Years of Education | 122 | 16 (14-18) | 52 | 16 (14-19) | 70 | 16 (14-18) | 0.52 |
| CDR (0/0.5/1+) | 122 | 59/48/15 | 52 | 30/22/0 | 70 | 29/26/15 | 0.0015 |
| CDR Sum of Boxes | 122 | 0.5 (0-1.5) | 52 | 0 (0-0.5) | 70 | 1 (0-3) | 0.0035 |
| Race (Black/White/Other) | 122 | 3/117/2 | 52 | 2/49/1 | 70 | 1/68/1 | 0.68 |
| Plasma collection to CDR (years) | 122 | 0.006 (0.000-0.038) | 52 | 0.003 (0.000-0.030) | 70 | 0.006 (0.000-0.038) | 0.82 |
| **Amyloid PET** | | | | | | | |
| Amyloid PET Centiloid | 122 | 35 (2-82) | 52 | 1 (-6-7) | 70 | 77 (45-100) | <0.0001 |
| Plasma collection to amyloid PET (years) | 122 | 0.016 (0.003-0.038) | 52 | 0.014 (0.000-0.033) | 70 | 0.019 (0.006-0.038) | 0.15 |
| **Tau PET** | | | | | | | |
| Early tau PET | 60 | 1.19 (1.10-1.30) | 25 | 1.09 (1.00-1.22) | 35 | 1.29 (1.16-1.45) | <0.0001 |
| Early tau PET Positivity | 60 | 14, 23.3% | 25 | 0, 0% | 35 | 14, 40.0% | 0.0003 |
| Late tau PET | 60 | 1.13 (1.06-1.2) | 25 | 1.07 (1.01-1.13) | 35 | 1.17 (1.09-1.33) | 0.0002 |
| Late tau PET Positivity | 60 | 13, 21.7% | 25 | 1, 4.0% | 35 | 12, 34.3% | 0.005 |
| Plasma collection to tau PET (years) | 60 | 0.033 (0.006-0.080) | 25 | 0.033 (0.003-0.079) | 35 | 0.027 (0.011-0.131) | 0.98 |
| **Cortical thickness** | | | | | | | |
| Cortical thickness | 91 | 2.83 (2.67-2.93) | 38 | 2.88 (2.79-2.94) | 53 | 2.79 (2.62-2.91) | 0.017 |
| Cortical thickness positivity | 91 | 10, 11.0% | 38 | 1, 2.6% | 53 | 9, 17.0% | 0.03 |
| Plasma collection to MRI (years) | 91 | 0.019 (0.003-0.041) | 38 | 0.011 (0.000-0.033) | 53 | 0.019 (0.003-0.047) | 0.096 |
| **CSF Elecsys** | | | | | | | |
| CSF p-tau181/Aβ42 | 122 | 0.0267 (0.0127-0.0506) | 52 | 0.0127 (0.00963-0.0162) | 70 | 0.0482 (0.0304-0.063) | <0.0001 |
| CSF p-tau181 (pg/ml) | 122 | 23.2 (17.7-31.2) | 52 | 19.2 (15.1-24.5) | 70 | 27.1 (22.2-36.6) | <0.0001 |
| CSF Aβ42 (pg/ml) | 122 | 870 (651-1440) | 52 | 1360 (1120-1830) | 70 | 691 (528-837) | <0.0001 |
| Plasma collection to CSF (years) | 122 | 0.000 (0.000-0.005) | 52 | 0.000 (0.000-0.005) | 70 | 0.000 (0.000-0.000) | 0.49 |

**Table S6. Classification accuracies of individual and combined plasma biomarker analytes for amyloid PET status defined by Centiloid>20 or ≤20 in the CSF sub-cohort.** The receiver operating characteristics area under the curve (AUC) point estimate (midpoint) and 95% confidence intervals are shown for classification of amyloid PET status (> or ≤20 Centiloids) by individual or combined plasma biomarker analytes. Both the unadjusted AUC and the AUC adjusted for age, sex, and *APOE* genotype are provided. AUCs were compared using DeLong's test. The Benjamin-Hochberg procedure was used to adjust for multiple comparisons with the reference analyte, either within a company or across companies. The AUC for a model with covariates only was 0.798 (0.716-0.879).

| Company | Analytes | Unadjusted for covariates | | | Adjusted for covariates | | |
|---|---|---|---|---|---|---|---|
| | | AUC | Within-platform comparisons p= | Across-platform comparisons p= | AUC | Within-platform comparisons p= | Across-platform comparisons p= |
| | CSF p-tau181/Aβ42 | 0.915 (0.864-0.967) | REFERENCE | REFERENCE | 0.936 (0.894-0.978) | REFERENCE | REFERENCE |
| C2N PrecivityAD2 | %p-tau217+ Aβ42/Aβ40 | 0.907 (0.851-0.963) | REFERENCE | 0.67 | 0.932 (0.887-0.977) | REFERENCE | 0.81 |
| | %p-tau217 | 0.907 (0.852-0.963) | 0.81 | | 0.928 (0.881-0.975) | 0.50 | |
| | p-tau217 | 0.903 (0.847-0.959) | 0.81 | | 0.920 (0.872-0.968) | 0.50 | |
| | p-tau217 + Aβ42/Aβ40 | 0.902 (0.845-0.959) | 0.81 | | 0.922 (0.877-0.968) | 0.50 | |
| | Aβ42/Aβ40 | 0.686 (0.591-0.781) | <0.0001 | | 0.798 (0.717-0.880) | 0.0022 | |
| Fujirebio Lumipulse | p-tau217 + Aβ42/Aβ40 | 0.897 (0.840-0.955) | REFERENCE | 0.58 | 0.925 (0.882-0.968) | REFERENCE | 0.69 |
| | p-tau217 | 0.895 (0.838-0.951) | 0.75 | | 0.921 (0.876-0.966) | 0.30 | |
| | Aβ42/Aβ40 | 0.688 (0.593-0.784) | <0.0001 | | 0.818 (0.739-0.896) | 0.0025 | |
| ALZpath Quanterix | p-tau217 | 0.883 (0.822-0.945) | | 0.22 | 0.911 (0.860-0.963) | | 0.25 |
| Janssen LucentAD Quanterix | p-tau217 | 0.858 (0.792-0.924) | | 0.12 | 0.894 (0.840-0.948) | | 0.12 |
| Roche NeuroToolKit | p-tau181 + Aβ42/Aβ40 + GFAP + NfL | 0.834 (0.760-0.908) | REFERENCE | 0.098 | 0.877 (0.816-0.938) | REFERENCE | 0.074 |
| | p-tau181 + Aβ42/Aβ40 + NfL | 0.830 (0.755-0.905) | 0.79 | | 0.872 (0.808-0.935) | 0.56 | |
| | p-tau181 + Aβ42/Aβ40 | 0.821 (0.743-0.898) | 0.54 | | 0.862 (0.795-0.928) | 0.38 | |
| | p-tau181 | 0.809 (0.731-0.887) | 0.5 | | 0.860 (0.793-0.927) | 0.38 | |
| | GFAP | 0.788 (0.707-0.869) | 0.33 | | 0.857 (0.791-0.922) | 0.38 | |
| | NfL | 0.711 (0.620-0.802) | 0.015 | | 0.842 (0.771-0.913) | 0.25 | |
| | Aβ42/Aβ40 | 0.677 (0.576-0.778) | 0.015 | | 0.802 (0.721-0.883) | 0.12 | |
| Quanterix Neurology 4-Plex | p-tau181 + Aβ42/Aβ40 + GFAP + NfL | 0.822 (0.740-0.905) | REFERENCE | 0.13 | 0.879 (0.811-0.947) | REFERENCE | 0.25 |
| | GFAP | 0.804 (0.715-0.893) | 0.54 | | 0.870 (0.799-0.941) | 0.54 | |
| | p-tau181 + Aβ42/Aβ40 + NfL | 0.788 (0.699-0.877) | 0.21 | | 0.844 (0.765-0.923) | 0.11 | |
| | p-tau181 + Aβ42/Aβ40 | 0.777 (0.687-0.868) | 0.19 | | 0.844 (0.764-0.924) | 0.11 | |
| | p-tau181 | 0.776 (0.684-0.868) | 0.19 | | 0.846 (0.767-0.925) | 0.11 | |
| | NfL | 0.709 (0.608-0.811) | 0.032 | | 0.836 (0.756-0.916) | 0.11 | |
| | Aβ42/Aβ40 | 0.615 (0.500-0.731) | 0.010 | | 0.812 (0.726-0.898) | 0.11 | |

**Table S7. Classification accuracies of individual plasma biomarker analytes for amyloid PET status defined by Centiloid>20 or ≤20 in the CSF sub-cohort.** The receiver operating characteristics area under the curve (AUC) point estimate (midpoint) and 95% confidence intervals are shown for classification of amyloid PET status (> or ≤20 Centiloids) by plasma biomarker analytes. The single cut-off for the plasma biomarker that best distinguished amyloid PET status based on the Youden index is shown, as well as the positive percent agreement (PPA), negative percent agreement (NPA), overall accuracy, positive predictive value (PPV), and negative predictive value (NPV) of the cut-off for amyloid PET status in the CSF sub-cohort, which had a 57.4% rate of amyloid PET positivity based on a cut-off of >20 Centiloids.

| Company | Analyte | AUC | Cut-off | Brier Score | PPA | NPA | Accuracy | PPV | NPV |
|---|---|---|---|---|---|---|---|---|---|
| **CSF** | CSF p-tau181/Aβ42 | 0.915 (0.864-0.967) | 0.0279 | 0.111 | 0.786 | 0.923 | 0.844 | 0.932 | 0.762 |
| **C2N PrecivityAD2** | %p-tau217 | 0.907 (0.852-0.963) | 3.99 (%) | 0.108 | 0.871 | 0.904 | 0.885 | 0.924 | 0.839 |
| | p-tau217 | 0.903 (0.847-0.959) | 2.34 (pg/ml) | 0.117 | 0.843 | 0.904 | 0.869 | 0.922 | 0.810 |
| | Aβ42/Aβ40 | 0.686 (0.591-0.781) | 0.0932 | 0.227 | 0.729 | 0.577 | 0.664 | 0.699 | 0.612 |
| **Fujirebio Lumipulse** | p-tau217 | 0.895 (0.838-0.951) | 0.201 (pg/ml) | 0.125 | 0.800 | 0.904 | 0.844 | 0.918 | 0.770 |
| | Aβ42/Aβ40 | 0.688 (0.593-0.784) | 0.0847 | 0.219 | 0.571 | 0.788 | 0.664 | 0.784 | 0.577 |
| **ALZpath Quanterix** | p-tau217 | 0.883 (0.822-0.945) | 0.414 (pg/ml) | 0.128 | 0.886 | 0.827 | 0.861 | 0.873 | 0.843 |
| **Janssen LucentAD Quanterix** | p-tau217 | 0.858 (0.792-0.924) | 0.0655 (pg/ml) | 0.153 | 0.771 | 0.846 | 0.803 | 0.871 | 0.733 |
| **Roche NeuroToolKit** | p-tau181 | 0.809 (0.731-0.887) | 1.13 (pg/ml) | 0.179 | 0.743 | 0.788 | 0.762 | 0.825 | 0.695 |
| | GFAP | 0.788 (0.707-0.869) | 0.105 (ng/ml) | 0.185 | 0.729 | 0.750 | 0.738 | 0.797 | 0.672 |
| | NfL | 0.711 (0.620-0.802) | 4.83 (pg/ml) | 0.211 | 0.386 | 0.923 | 0.615 | 0.871 | 0.527 |
| | Aβ42/Aβ40 | 0.677 (0.576-0.778) | 0.122 | 0.227 | 0.743 | 0.635 | 0.697 | 0.732 | 0.647 |
| **Quanterix Neurology 4-Plex** | GFAP | 0.804 (0.715-0.893) | 136 (pg/ml) | 0.183 | 0.818 | 0.667 | 0.750 | 0.750 | 0.750 |
| | p-tau181 | 0.776 (0.684-0.868) | 23.3 (pg/ml) | 0.188 | 0.618 | 0.889 | 0.740 | 0.872 | 0.656 |
| | NfL | 0.709 (0.608-0.811) | 20.0 (pg/ml) | 0.216 | 0.764 | 0.578 | 0.680 | 0.689 | 0.667 |
| | Aβ42/Aβ40 | 0.615 ( 0.500-0.731) | 0.0604 | 0.240 | 0.727 | 0.511 | 0.630 | 0.645 | 0.605 |

**Table S8. Classification accuracies of individual and combined plasma biomarker analytes for amyloid PET status defined by Centiloid>20 or ≤20 in the cognitively impaired sub-cohort.** The receiver operating characteristics area under the curve (AUC) point estimate (midpoint) and 95% confidence intervals are shown for classification of amyloid PET status (> or ≤20 Centiloids) by individual or combined plasma biomarker analytes. Both the unadjusted AUC and the AUC adjusted for age, sex, and *APOE* genotype are provided. AUCs were compared using DeLong's test. The Benjamin-Hochberg procedure was used to adjust for multiple comparisons with the reference analyte, either within a company or across companies. The AUC for a model with covariates only was 0.740 (0.669-0.811).

| Company | Analytes | Unadjusted for covariates | | | Adjusted for covariates | | |
|---|---|---|---|---|---|---|---|
| | | AUC | Within-company comparisons p= | Across-company comparisons p= | AUC | Within-company comparisons p= | Across-company comparisons p= |
| C2N PrecivityAD2 | p-tau217 + Aβ42/Aβ40 | 0.960 (0.934-0.986) | REFERENCE | REFERENCE | 0.962 (0.936-0.987) | REFERENCE | REFERENCE |
| | %p-tau217+ Aβ42/Aβ40 | 0.958 (0.932-0.985) | 0.80 | | 0.961 (0.935-0.986) | 0.87 | |
| | p-tau217 | 0.955 (0.927-0.984) | 0.22 | | 0.958 (0.930-0.986) | 0.41 | |
| | %p-tau217 | 0.952 (0.922-0.981) | 0.45 | | 0.954 (0.926-0.983) | 0.43 | |
| | Aβ42/Aβ40 | 0.759 (0.688-0.829) | <0.0001 | | 0.800 (0.735-0.866) | <0.0001 | |
| Fujirebio Lumipulse | p-tau217 + Aβ42/Aβ40 | 0.952 (0.925-0.980) | REFERENCE | 0.48 | 0.952 (0.924-0.980) | REFERENCE | 0.39 |
| | p-tau217 | 0.951 (0.922-0.979) | 0.46 | | 0.950 (0.922-0.979) | 0.44 | |
| | Aβ42/Aβ40 | 0.803 (0.733-0.872) | <0.0001 | | 0.820 (0.757-0.884) | <0.0001 | |
| ALZpath Quanterix | p-tau217 | 0.942 (0.910-0.974) | | 0.19 | 0.951 (0.922-0.980) | | 0.39 |
| Janssen LucentAD Quanterix | p-tau217 | 0.924 (0.886-0.962) | | 0.032 | 0.932 (0.897-0.966) | | 0.056 |
| Roche NeuroToolKit | p-tau181 + Aβ42/Aβ40 + NfL | 0.921 (0.880-0.962) | REFERENCE | 0.077 | 0.928 (0.889-0.966) | 0.98 | |
| | p-tau181 + Aβ42/Aβ40 + GFAP + NfL | 0.921 (0.880-0.961) | 0.98 | | 0.928 (0.890-0.966) | REFERENCE | 0.091 |
| | p-tau181 + Aβ42/Aβ40 | 0.920 (0.879-0.961) | 0.89 | | 0.926 (0.887-0.965) | 0.92 | |
| | p-tau181 | 0.874 (0.822-0.926) | 0.012 | | 0.905 (0.862-0.947) | 0.084 | |
| | GFAP | 0.805 (0.740-0.870) | 0.0013 | | 0.838 (0.782-0.895) | 0.0015 | |
| | Aβ42/Aβ40 | 0.790 (0.718-0.862) | 0.0004 | | 0.817 (0.754-0.880) | 0.0001 | |
| | NfL | 0.728 (0.654-0.802) | <0.0001 | | 0.808 (0.745-0.871) | 0.0001 | |
| Quanterix Neurology 4-Plex | p-tau181 + Aβ42/Aβ40 + GFAP + NfL | 0.833 (0.770-0.895) | REFERENCE | 0.0014 | 0.871 (0.819-0.923) | REFERENCE | 0.012 |
| | p-tau181 | 0.804 (0.733-0.874) | 0.39 | | 0.832 (0.769-0.894) | 0.13 | |
| | p-tau181 + Aβ42/Aβ40 | 0.801 (0.727-0.874) | 0.37 | | 0.838 (0.775-0.900) | 0.13 | |
| | GFAP | 0.800 (0.731-0.869) | 0.22 | | 0.846 (0.788-0.904) | 0.15 | |
| | p-tau181 + Aβ42/Aβ40 + NfL | 0.794 (0.723-0.865) | 0.22 | | 0.853 (0.794-0.911) | 0.23 | |
| | Aβ42/Aβ40 | 0.750 (0.671-0.828) | 0.14 | | 0.793 (0.723-0.862) | 0.017 | |
| | NfL | 0.719 (0.640-0.798) | 0.0054 | | 0.807 (0.742-0.872) | 0.017 | |

**Table S9. Classification accuracies of individual plasma biomarker analytes for amyloid PET status defined by Centiloid>20 or ≤20 in the cognitively impaired sub-cohort.** The receiver operating characteristics area under the curve (AUC) point estimate (midpoint) and 95% confidence intervals are shown for classification of amyloid PET status (> or ≤20 Centiloids) by plasma biomarker analytes. The single cut-off for the plasma biomarker that best distinguished amyloid PET status based on the Youden index is shown, as well as the positive percent agreement (PPA), negative percent agreement (NPA), overall accuracy, positive predictive value (PPV), and negative predictive value (NPV) of the cut-off for amyloid PET status in the cognitively impaired sub-cohort, which had a 59.9% rate of amyloid PET positivity based on a cut-off of >20 Centiloids.

| Company | Analyte | AUC | Cut-off | Brier Score | PPA | NPA | Accuracy | PPV | NPV |
|---|---|---|---|---|---|---|---|---|---|
| C2N PrecivityAD2 | p-tau217 | 0.955 (0.927-0.984) | 2.41 (pg/ml) | 0.076 | 0.939 | 0.896 | 0.922 | 0.931 | 0.908 |
| | %p-tau217 | 0.952 (0.922-0.981) | 4.85 (%) | 0.080 | 0.878 | 0.922 | 0.896 | 0.944 | 0.835 |
| | Aβ42/Aβ40 | 0.759 (0.688-0.829) | 0.0974 | 0.195 | 0.878 | 0.545 | 0.745 | 0.743 | 0.750 |
| Fujirebio Lumipulse | p-tau217 | 0.951 (0.922-0.979) | 0.207 (pg/ml) | 0.085 | 0.852 | 0.922 | 0.880 | 0.942 | 0.807 |
| | Aβ42/Aβ40 | 0.803 (0.733-0.872) | 0.0862 | 0.179 | 0.730 | 0.792 | 0.755 | 0.840 | 0.663 |
| ALZpath Quanterix | p-tau217 | 0.942 (0.910-0.974) | 0.462 (pg/ml) | 0.090 | 0.922 | 0.844 | 0.891 | 0.898 | 0.878 |
| Janssen LucentAD Quanterix | p-tau217 | 0.924 (0.886-0.962) | 0.0695 (pg/ml) | 0.106 | 0.870 | 0.870 | 0.870 | 0.909 | 0.817 |
| Roche NeuroToolKit | p-tau181 | 0.874 (0.822-0.926) | 1.14 (pg/ml) | 0.139 | 0.870 | 0.740 | 0.818 | 0.833 | 0.792 |
| | GFAP | 0.805 (0.740-0.870) | 0.116 (ng/ml) | 0.177 | 0.783 | 0.753 | 0.771 | 0.826 | 0.699 |
| | Aβ42/Aβ40 | 0.790 (0.718-0.862) | 0.123 | 0.178 | 0.800 | 0.727 | 0.771 | 0.814 | 0.709 |
| | NfL | 0.728 (0.654-0.802) | 4.38 (pg/ml) | 0.210 | 0.661 | 0.727 | 0.688 | 0.784 | 0.589 |
| Quanterix Neurology 4-Plex | p-tau181 | 0.804 (0.733-0.874) | 21.0 (pg/ml) | 0.192 | 0.776 | 0.767 | 0.772 | 0.817 | 0.718 |
| | GFAP | 0.800 (0.731-0.869) | 194 (pg/ml) | 0.181 | 0.714 | 0.808 | 0.754 | 0.833 | 0.678 |
| | Aβ42/Aβ40 | 0.750 (0.671-0.828) | 0.0582 | 0.210 | 0.776 | 0.712 | 0.749 | 0.784 | 0.703 |
| | NfL | 0.719 (0.640-0.798) | 21.7 (pg/ml) | 0.215 | 0.755 | 0.616 | 0.696 | 0.725 | 0.652 |

**Table S10. Classification accuracies of individual and combined plasma biomarker analytes for amyloid PET status defined by Centiloid>20 or ≤20 in the cognitively unimpaired sub-cohort.** The receiver operating characteristics area under the curve (AUC) point estimate (midpoint) and 95% confidence intervals are shown for classification of amyloid PET status (> or ≤20 Centiloids) by individual or combined plasma biomarker analytes. Both the unadjusted AUC and the AUC adjusted for age, sex, and *APOE* genotype are provided. AUCs were compared using DeLong's test. The Benjamin-Hochberg procedure was used to adjust for multiple comparisons with the reference analyte, either within a company or across companies. The AUC for a model with covariates only was 0.727 (0.657-0.797).

| Company | Analytes | Unadjusted for covariates | | | Adjusted for covariates | | |
|---|---|---|---|---|---|---|---|
| | | AUC | Within-company comparisons p= | Across-company comparisons p= | AUC | Within-company comparisons p= | Across-company comparisons p= |
| C2N PrecivityAD2 | %p-tau217 | 0.892 (0.842-0.942) | REFERENCE | REFERENCE | 0.897 (0.848-0.947) | 0.82 | |
| | %p-tau217+ Aβ42/Aβ40 | 0.890 (0.839-0.942) | 0.68 | | 0.898 (0.849-0.947) | REFERENCE | REFERENCE |
| | p-tau217 + Aβ42/Aβ40 | 0.866 (0.808-0.923) | 0.053 | | 0.882 (0.831-0.934) | 0.17 | |
| | p-tau217 | 0.863 (0.807-0.919) | 0.0312 | | 0.877 (0.824-0.930) | 0.13 | |
| | Aβ42/Aβ40 | 0.740 (0.666-0.813) | <0.0001 | | 0.775 (0.708-0.842) | 0.0003 | |
| Fujirebio Lumipulse | p-tau217 + Aβ42/Aβ40 | 0.840 (0.782-0.897) | REFERENCE | 0.024 | 0.853 (0.799-0.908) | REFERENCE | 0.040 |
| | p-tau217 | 0.818 (0.756-0.881) | 0.46 | | 0.828 (0.771-0.884) | 0.19 | |
| | Aβ42/Aβ40 | 0.762 (0.694-0.829) | 0.0001 | | 0.805 (0.744-0.866) | 0.0026 | |
| ALZpath Quanterix | p-tau217 | 0.805 (0.738-0.873) | | 0.0011 | 0.843 (0.786-0.900) | | 0.0089 |
| Janssen LucentAD Quanterix | p-tau217 | 0.812 (0.749-0.875) | | 0.0011 | 0.843 (0.787-0.900) | | 0.0089 |
| Roche NeuroToolKit | p-tau181 + Aβ42/Aβ40 + GFAP + NfL | 0.816 (0.753-0.879) | REFERENCE | 0.0043 | 0.846 (0.790-0.902) | REFERENCE | 0.024 |
| | p-tau181 + Aβ42/Aβ40 + NfL | 0.811 (0.748-0.875) | 0.55 | | 0.842 (0.786-0.898) | 0.48 | |
| | p-tau181 + Aβ42/Aβ40 | 0.811 (0.748-0.875) | 0.55 | | 0.841 (0.785-0.898) | 0.48 | |
| | Aβ42/Aβ40 | 0.752 (0.684-0.821) | 0.018 | | 0.795 (0.733-0.857) | 0.017 | |
| | p-tau181 | 0.721 (0.647-0.796) | 0.0043 | | 0.790 (0.727-0.854) | 0.014 | |
| | GFAP | 0.687 (0.610-0.765) | 0.0024 | | 0.773 (0.707-0.839) | 0.014 | |
| | NfL | 0.593 (0.512-0.675) | <0.0001 | | 0.736 (0.666-0.805) | 0.0019 | |
| Quanterix Neurology 4-Plex | p-tau181 + Aβ42/Aβ40 + GFAP + NfL | 0.753 (0.678-0.828) | REFERENCE | 0.0043 | 0.813 (0.747-0.880) | REFERENCE | 0.046 |
| | p-tau181 + Aβ42/Aβ40 | 0.733 (0.657-0.810) | 0.23 | | 0.798 (0.730-0.867) | 0.25 | |
| | p-tau181 + Aβ42/Aβ40 + NfL | 0.732 (0.656-0.808) | 0.23 | | 0.799 (0.730-0.867) | 0.25 | |
| | Aβ42/Aβ40 | 0.702 (0.623-0.781) | 0.11 | | 0.778 (0.708-0.849) | 0.14 | |
| | GFAP | 0.685 (0.602-0.769) | 0.11 | | 0.771 (0.697-0.845) | 0.14 | |
| | p-tau181 | 0.661 (0.576-0.747) | 0.058 | | 0.758 (0.683-0.833) | 0.12 | |
| | NfL | 0.593 (0.504-0.683) | 0.0098 | | 0.733 (0.657-0.809) | 0.084 | |

**Table S11. Classification accuracies of individual plasma biomarker analytes for amyloid PET status defined by Centiloid>20 or ≤20 in the cognitively unimpaired sub-cohort.** The receiver operating characteristics area under the curve (AUC) point estimate (midpoint) and 95% confidence intervals are shown for classification of amyloid PET status (> or ≤20 Centiloids) by plasma biomarker analytes. The single cut-off for the plasma biomarker that best distinguished amyloid PET status based on the Youden index is shown, as well as the positive percent agreement (PPA), negative percent agreement (NPA), overall accuracy, positive predictive value (PPV), and negative predictive value (NPV) of the cut-off for amyloid PET status in the cognitively unimpaired sub-cohort, which had a 38.0% rate of amyloid PET positivity based on a cut-off of >20 Centiloids.

| Company | Analyte | AUC | Cut-off | Brier Score | PPA | NPA | Accuracy | PPV | NPV |
|---|---|---|---|---|---|---|---|---|---|
| **C2N PrecivityAD2** | %p-tau217 | 0.892 (0.842-0.942) | 4.06 (%) | 0.114 | 0.816 | 0.887 | 0.860 | 0.816 | 0.887 |
| | p-tau217 | 0.863 (0.807-0.919) | 2.18 (pg/ml) | 0.135 | 0.776 | 0.847 | 0.820 | 0.756 | 0.861 |
| | Aβ42/Aβ40 | 0.740 (0.666-0.813) | 0.0892 | 0.198 | 0.605 | 0.782 | 0.715 | 0.630 | 0.764 |
| **Fujirebio Lumipulse** | p-tau217 | 0.818 (0.756-0.881) | 0.158 (pg/ml) | 0.173 | 0.684 | 0.831 | 0.775 | 0.712 | 0.811 |
| | Aβ42/Aβ40 | 0.762 (0.694-0.829) | 0.0887 | 0.190 | 0.711 | 0.718 | 0.715 | 0.607 | 0.802 |
| **Janssen LucentAD Quanterix** | p-tau217 | 0.812 (0.749-0.875) | 0.0535 (pg/ml) | 0.166 | 0.763 | 0.774 | 0.770 | 0.674 | 0.842 |
| **ALZpath Quanterix** | p-tau217 | 0.805 (0.738-0.873) | 0.449 (pg/ml) | 0.162 | 0.697 | 0.847 | 0.790 | 0.736 | 0.820 |
| **Roche NeuroToolKit** | Aβ42/Aβ40 | 0.752 (0.684-0.821) | 0.126 | 0.199 | 0.816 | 0.637 | 0.705 | 0.579 | 0.849 |
| | p-tau181 | 0.721 (0.647-0.796) | 1.13 (pg/ml) | 0.201 | 0.645 | 0.750 | 0.710 | 0.613 | 0.775 |
| | GFAP | 0.687 (0.610-0.765) | 0.146 (ng/ml) | 0.215 | 0.408 | 0.863 | 0.690 | 0.646 | 0.704 |
| | NfL | 0.593 (0.512-0.675) | 3.05 (pg/ml) | 0.227 | 0.803 | 0.339 | 0.515 | 0.427 | 0.737 |
| **Quanterix Neurology 4-Plex** | Aβ42/Aβ40 | 0.702 (0.623-0.781) | 0.0582 | 0.205 | 0.656 | 0.673 | 0.667 | 0.526 | 0.779 |
| | GFAP | 0.685 (0.602-0.769) | 212 (pg/ml) | 0.210 | 0.459 | 0.845 | 0.708 | 0.622 | 0.738 |
| | p-tau181 | 0.661 (0.576-0.747) | 26.1 (pg/ml) | 0.211 | 0.410 | 0.827 | 0.678 | 0.568 | 0.717 |
| | NfL | 0.593 (0.504-0.683) | 15.9 (pg/ml) | 0.222 | 0.820 | 0.336 | 0.509 | 0.407 | 0.771 |

**Table S12. Classification accuracies of individual and combined plasma biomarker analytes for amyloid PET status defined by Centiloid >37 or ≤37 in the full cohort.** The receiver operating characteristics area under the curve (AUC) point estimate (midpoint) and 95% confidence intervals are shown for classification of amyloid PET status (> or ≤37 Centiloids) by individual or combined plasma biomarker analytes. Both the unadjusted AUC and the AUC adjusted for age, sex, and *APOE* genotype are provided. AUCs were compared using DeLong's test. The Benjamin-Hochberg procedure was used to adjust for multiple comparisons with the reference analyte, either within a company or across companies. The AUC for a model with covariates only was 0.706 (0.655-0.758).

| Company | Analytes | Unadjusted for covariates | | | Adjusted for covariates | | |
|---|---|---|---|---|---|---|---|
| | | AUC | Within-company comparisons p= | Across-company comparisons p= | AUC | Within-company comparisons p= | Across-company comparisons p= |
| C2N PrecivityAD2 | %p-tau217+ Aβ42/Aβ40 | 0.941 (0.919-0.964) | REFERENCE | REFERENCE | 0.941 (0.919-0.964) | 0.89 | |
| | %p-tau217 | 0.941 (0.918-0.964) | 0.73 | | 0.942 (0.919-0.964) | REFERENCE | REFERENCE |
| | p-tau217 + Aβ42/Aβ40 | 0.938 (0.915-0.961) | 0.73 | | 0.939 (0.916-0.962) | 0.89 | |
| | p-tau217 | 0.936 (0.912-0.960) | 0.73 | | 0.938 (0.915-0.962) | 0.89 | |
| | Aβ42/Aβ40 | 0.728 (0.677-0.779) | <0.0001 | | 0.761 (0.713-0.808) | <0.0001 | |
| Fujirebio Lumipulse | p-tau217 + Aβ42/Aβ40 | 0.925 (0.898-0.951) | REFERENCE | 0.12 | 0.923 (0.897-0.949) | REFERENCE | 0.083 |
| | p-tau217 | 0.919 (0.892-0.946) | 0.50 | | 0.915 (0.886-0.943) | 0.21 | |
| | Aβ42/Aβ40 | 0.773 (0.726-0.820) | <0.0001 | | 0.797 (0.753-0.841) | <0.0001 | |
| ALZpath Quanterix | p-tau217 | 0.910 (0.880-0.940) | | 0.013 | 0.918 (0.890-0.946) | | 0.035 |
| Janssen LucentAD Quanterix | p-tau217 | 0.904 (0.874-0.933) | | 0.0047 | 0.909 (0.881-0.937) | | 0.010 |
| Roche NeuroToolKit | p-tau181 + Aβ42/Aβ40 + GFAP + NfL | 0.884 (0.852-0.917) | REFERENCE | 0.0004 | 0.896 (0.865-0.928) | REFERENCE | 0.0039 |
| | p-tau181 + Aβ42/Aβ40 | 0.881 (0.847-0.914) | 0.42 | | 0.89 (0.858-0.922) | 0.21 | |
| | p-tau181 + Aβ42/Aβ40 + NfL | 0.880 (0.847-0.914) | 0.42 | | 0.89 (0.858-0.922) | 0.18 | |
| | p-tau181 | 0.843 (0.804-0.882) | 0.0034 | | 0.866 (0.83-0.903) | 0.0051 | |
| | GFAP | 0.781 (0.736-0.827) | <0.0001 | | 0.82 (0.778-0.862) | <0.0001 | |
| | Aβ42/Aβ40 | 0.754 (0.706-0.801) | <0.0001 | | 0.781 (0.735-0.826) | <0.0001 | |
| | NfL | 0.693 (0.639-0.746) | <0.0001 | | 0.773 (0.727-0.819) | <0.0001 | |
| Quanterix Neurology 4-Plex | p-tau181 + Aβ42/Aβ40+ GFAP + NfL | 0.828 (0.786-0.871) | REFERENCE | <0.0001 | 0.854 (0.814-0.894) | REFERENCE | 0.0011 |
| | p-tau181 + Aβ42/Aβ40 + NfL | 0.800 (0.754-0.847) | 0.042 | | 0.829 (0.785-0.872) | 0.034 | |
| | p-tau181 + Aβ42/Aβ40 | 0.799 (0.752-0.846) | 0.048 | | 0.825 (0.782-0.869) | 0.034 | |
| | GFAP | 0.778 (0.728-0.828) | 0.0098 | | 0.818 (0.772-0.865) | 0.0082 | |
| | p-tau181 | 0.774 (0.724-0.824) | 0.014 | | 0.804 (0.757-0.852) | 0.0054 | |
| | Aβ42/Aβ40 | 0.737 (0.684-0.790) | 0.0006 | | 0.774 (0.724-0.824) | 0.0007 | |
| | NfL | 0.690 (0.632-0.747) | <0.0001 | | 0.762 (0.71-0.815) | <0.0001 | |

**Table S13. Classification accuracies of individual plasma biomarker analytes for amyloid PET status defined by Centiloid>37 or ≤37 in the full cohort.** The receiver operating characteristics area under the curve (AUC) point estimate (midpoint) and 95% confidence intervals are shown for classification of amyloid PET status (> or ≤37 Centiloids) by plasma biomarker analytes. The single cut-off for the plasma biomarker that best distinguished amyloid PET status based on the Youden index is shown, as well as the positive percent agreement (PPA), negative percent agreement (NPA), overall accuracy, positive predictive value (PPV), and negative predictive value (NPV) of the cut-off for amyloid PET status in the full cohort, which had a 40.1% rate of amyloid PET positivity based on a cut-off of >37 Centiloids.

| Company | Analyte | AUC | Cut-off | Brier Score | PPA | NPA | Accuracy | PPV | NPV |
|---|---|---|---|---|---|---|---|---|---|
| **C2N PrecivityAD2** | %p-tau217 | 0.941 (0.918-0.964) | 4.60 (%) | 0.093 | 0.930 | 0.838 | 0.875 | 0.793 | 0.947 |
| | p-tau217 | 0.936 (0.912-0.960) | 2.45 (pg/ml) | 0.099 | 0.924 | 0.838 | 0.872 | 0.792 | 0.943 |
| | Aβ42/Aβ40 | 0.728 (0.677-0.779) | 0.0953 | 0.206 | 0.834 | 0.540 | 0.658 | 0.548 | 0.830 |
| **Fujirebio Lumipulse** | p-tau217 | 0.919 (0.892-0.946) | 0.177 (pg/ml) | 0.116 | 0.879 | 0.821 | 0.844 | 0.767 | 0.910 |
| | Aβ42/Aβ40 | 0.773 (0.726-0.820) | 0.0852 | 0.192 | 0.707 | 0.749 | 0.732 | 0.653 | 0.793 |
| **ALZpath Quanterix** | p-tau217 | 0.910 (0.880-0.940) | 0.444 (pg/ml) | 0.117 | 0.917 | 0.783 | 0.837 | 0.738 | 0.934 |
| **Janssen LucentAD Quanterix** | p-tau217 | 0.904 (0.874-0.933) | 0.0695 (pg/ml) | 0.126 | 0.822 | 0.838 | 0.832 | 0.772 | 0.876 |
| **Roche NeuroToolKit** | p-tau181 | 0.843 (0.804-0.882) | 1.23 (pg/ml) | 0.160 | 0.783 | 0.774 | 0.778 | 0.699 | 0.843 |
| | GFAP | 0.781 (0.736-0.827) | 0.113 (ng/ml) | 0.190 | 0.777 | 0.655 | 0.704 | 0.601 | 0.815 |
| | Aβ42/Aβ40 | 0.754 (0.706-0.801) | 0.125 | 0.202 | 0.860 | 0.574 | 0.689 | 0.574 | 0.860 |
| | NfL | 0.693 (0.639-0.746) | 4.43 (pg/ml) | 0.214 | 0.580 | 0.715 | 0.661 | 0.576 | 0.718 |
| **Quanterix Neurology 4-Plex** | GFAP | 0.778 (0.728-0.828) | 205 (pg/ml) | 0.187 | 0.661 | 0.791 | 0.743 | 0.651 | 0.798 |
| | p-tau181 | 0.774 (0.724-0.824) | 20.5 (pg/ml) | 0.191 | 0.780 | 0.670 | 0.711 | 0.582 | 0.837 |
| | Aβ42/Aβ40 | 0.737 (0.684-0.790) | 0.0582 | 0.205 | 0.764 | 0.647 | 0.690 | 0.561 | 0.822 |
| | NfL | 0.690 (0.632-0.747) | 22.8 (pg/ml) | 0.210 | 0.685 | 0.600 | 0.632 | 0.503 | 0.763 |

**Table S14. Classification accuracies of individual and combined plasma biomarker analytes for amyloid PET status defined by Centiloid>37 or ≤37 in the cognitively impaired sub-cohort.** The receiver operating characteristics area under the curve (AUC) point estimate (midpoint) and 95% confidence intervals are shown for classification of amyloid PET status (> or ≤37 Centiloids) by individual or combined plasma biomarker analytes. Both the unadjusted AUC and the AUC adjusted for age, sex, and *APOE* genotype are provided. AUCs were compared using DeLong's test. The Benjamin-Hochberg procedure was used to adjust for multiple comparisons with the reference analyte, either within a company or across companies. The AUC for a model with covariates only was 0.714 (0.641-0.786).

| Company | Analytes | Unadjusted for covariates | | | Adjusted for covariates | | |
|---|---|---|---|---|---|---|---|
| | | AUC | Within-company comparisons p= | Across-company comparisons p= | AUC | Within-company comparisons p= | Across-company comparisons p= |
| C2N PrecivityAD2 | p-tau217 + Aβ42/Aβ40 | 0.956 (0.930-0.982) | REFERENCE | REFERENCE | 0.960 (0.936-0.984) | REFERENCE | REFERENCE |
| | p-tau217 | 0.954 (0.927-0.981) | 0.39 | | 0.959 (0.934-0.983) | 0.60 | |
| | %p-tau217+ Aβ42/Aβ40 | 0.947 (0.917-0.977) | 0.30 | | 0.952 (0.923-0.980) | 0.34 | |
| | %p-tau217 | 0.944 (0.913-0.975) | 0.30 | | 0.946 (0.915-0.977) | 0.26 | |
| | Aβ42/Aβ40 | 0.744 (0.673-0.815) | <0.0001 | | 0.789 (0.723-0.854) | <0.0001 | |
| Fujirebio Lumipulse | p-tau217 + Aβ42/Aβ40 | 0.960 (0.936-0.984) | REFERENCE | 0.69 | 0.961 (0.937-0.985) | REFERENCE | 0.93 |
| | p-tau217 | 0.959 (0.935-0.983) | 0.70 | | 0.961 (0.937-0.984) | 0.92 | |
| | Aβ42/Aβ40 | 0.796 (0.729-0.862) | <0.0001 | | 0.809 (0.746-0.871) | <0.0001 | |
| ALZpath Quanterix | p-tau217 | 0.951 (0.922-0.979) | | 0.69 | 0.961 (0.936-0.986) | | 0.93 |
| Janssen LucentAD Quanterix | p-tau217 | 0.936 (0.904-0.967) | | 0.20 | 0.943 (0.914-0.973) | | 0.28 |
| Roche NeuroToolKit | p-tau181 + Aβ42/Aβ40 + GFAP + NfL | 0.918 (0.877-0.960) | REFERENCE | 0.085 | 0.932 (0.896-0.967) | 0.75 | |
| | p-tau181 + Aβ42/Aβ40 + NfL | 0.916 (0.874-0.958) | 0.64 | | 0.933 (0.897-0.969) | REFERENCE | 0.28 |
| | p-tau181 + Aβ42/Aβ40 | 0.915 (0.873-0.957) | 0.64 | | 0.929 (0.892-0.967) | 0.54 | |
| | p-tau181 | 0.882 (0.835-0.929) | 0.037 | | 0.916 (0.876-0.955) | 0.16 | |
| | GFAP | 0.804 (0.741-0.867) | 0.0002 | | 0.838 (0.782-0.893) | 0.0012 | |
| | Aβ42/Aβ40 | 0.757 (0.686-0.827) | <0.0001 | | 0.782 (0.717-0.847) | <0.0001 | |
| | NfL | 0.726 (0.654-0.797) | <0.0001 | | 0.807 (0.745-0.869) | <0.0001 | |
| Quanterix Neurology 4-Plex | p-tau181 + Aβ42/Aβ40+ GFAP + NfL | 0.839 (0.780-0.898) | REFERENCE | 0.0022 | 0.887 (0.839-0.935) | REFERENCE | 0.043 |
| | GFAP | 0.811 (0.747-0.876) | 0.25 | | 0.854 (0.799-0.909) | 0.076 | |
| | p-tau181 | 0.811 (0.746-0.877) | 0.37 | | 0.846 (0.788-0.905) | 0.076 | |
| | p-tau181 + Aβ42/Aβ40 | 0.804 (0.738-0.871) | 0.25 | | 0.849 (0.792-0.906) | 0.076 | |
| | p-tau181 + Aβ42/Aβ40 + NfL | 0.802 (0.737-0.868) | 0.23 | | 0.862 (0.807-0.918) | 0.13 | |
| | Aβ42/Aβ40 | 0.743 (0.667-0.820) | 0.060 | | 0.778 (0.708-0.848) | 0.0039 | |
| | NfL | 0.716 (0.640-0.793) | 0.0015 | | 0.807 (0.741-0.873) | 0.0039 | |

**Table S15. Classification accuracies of individual plasma biomarker analytes for amyloid PET status defined by Centiloid>37 or ≤37 in the cognitively impaired sub-cohort.** The receiver operating characteristics area under the curve (AUC) point estimate (midpoint) and 95% confidence intervals are shown for classification of amyloid PET status (> or ≤37 Centiloids) by plasma biomarker analytes. The single cut-off for the plasma biomarker that best distinguished amyloid PET status based on the Youden index is shown, as well as the positive percent agreement (PPA), negative percent agreement (NPA), overall accuracy, positive predictive value (PPV), and negative predictive value (NPV) of the cut-off for amyloid PET status in the cognitively impaired sub-cohort, which had a 52.1% rate of amyloid PET positivity based on a cut-off of >37 Centiloids.

| Company | Analyte | AUC | Cut-off | Brier Score | PPA | NPA | Accuracy | PPV | NPV |
|---|---|---|---|---|---|---|---|---|---|
| Fujirebio Lumipulse | p-tau217 | 0.959 (0.935-0.983) | 0.208 (pg/ml) | 0.084 | 0.920 | 0.870 | 0.896 | 0.885 | 0.909 |
| | Aβ42/Aβ40 | 0.796 (0.729-0.862) | 0.0847 | 0.191 | 0.730 | 0.804 | 0.766 | 0.802 | 0.733 |
| C2N PrecivityAD2 | p-tau217 | 0.954 (0.927-0.981) | 2.85 (pg/ml) | 0.087 | 0.920 | 0.880 | 0.901 | 0.893 | 0.910 |
| | %p-tau217 | 0.944 (0.913-0.975) | 4.86 (%) | 0.093 | 0.930 | 0.848 | 0.891 | 0.869 | 0.918 |
| | Aβ42/Aβ40 | 0.744 (0.673-0.815) | 0.0879 | 0.207 | 0.580 | 0.826 | 0.698 | 0.784 | 0.644 |
| ALZpath Quanterix | p-tau217 | 0.951 (0.922-0.979) | 0.596 (pg/ml) | 0.089 | 0.850 | 0.924 | 0.885 | 0.924 | 0.850 |
| Janssen LucentAD Quanterix | p-tau217 | 0.936 (0.904-0.967) | 0.0695 (pg/ml) | 0.107 | 0.930 | 0.815 | 0.875 | 0.845 | 0.915 |
| Roche NeuroToolKit | p-tau181 | 0.882 (0.835-0.929) | 1.31 (pg/ml) | 0.142 | 0.800 | 0.815 | 0.807 | 0.825 | 0.789 |
| | GFAP | 0.804 (0.741-0.867) | 0.117 (ng/ml) | 0.186 | 0.790 | 0.707 | 0.750 | 0.745 | 0.756 |
| | Aβ42/Aβ40 | 0.757 (0.686-0.827) | 0.123 | 0.202 | 0.810 | 0.652 | 0.734 | 0.717 | 0.759 |
| | NfL | 0.726 (0.654-0.797) | 3.73 (pg/ml) | 0.218 | 0.820 | 0.543 | 0.688 | 0.661 | 0.735 |
| Quanterix Neurology 4-Plex | p-tau181 | 0.811 (0.746-0.877) | 18.9 (pg/ml) | 0.192 | 0.893 | 0.632 | 0.760 | 0.701 | 0.859 |
| | GFAP | 0.811 (0.747-0.876) | 150 (pg/ml) | 0.184 | 0.905 | 0.621 | 0.760 | 0.697 | 0.871 |
| | Aβ42/Aβ40 | 0.743 (0.667-0.820) | 0.0582 | 0.222 | 0.810 | 0.667 | 0.737 | 0.701 | 0.784 |
| | NfL | 0.716 (0.640-0.793) | 19.8 (pg/ml) | 0.221 | 0.857 | 0.494 | 0.673 | 0.621 | 0.782 |

**Table S16. Classification accuracies of individual and combined plasma biomarker analytes for amyloid PET status defined by Centiloid>37 or ≤37 in the cognitively unimpaired sub-cohort.** The receiver operating characteristics area under the curve (AUC) point estimate (midpoint) and 95% confidence intervals are shown for classification of amyloid PET status (> or ≤37 Centiloids) by individual or combined plasma biomarker analytes. Both the unadjusted AUC and the AUC adjusted for age, sex, and *APOE* genotype are provided. AUCs were compared using DeLong's test. The Benjamin-Hochberg procedure was used to adjust for multiple comparisons with the reference analyte, either within a company or across companies. The AUC for a model with covariates only was 0.713 (0.639-0.788).

| Company | Analytes | Unadjusted for covariates | | | Adjusted for covariates | | |
|---|---|---|---|---|---|---|---|
| | | AUC | Within-company comparisons p= | Across-company comparisons p= | AUC | Within-company comparisons p= | Across-company comparisons p= |
| C2N PrecivityAD2 | %p-tau217+ Aβ42/Aβ40 | 0.929 (0.889-0.970) | REFERENCE | REFERENCE | 0.932 (0.896-0.969) | REFERENCE | REFERENCE |
| | %p-tau217 | 0.928 (0.889-0.968) | 0.68 | | 0.932 (0.896-0.968) | 0.84 | |
| | p-tau217 + Aβ42/Aβ40 | 0.907 (0.864-0.951) | 0.074 | | 0.914 (0.874-0.954) | 0.091 | |
| | p-tau217 | 0.903 (0.856-0.951) | 0.051 | | 0.914 (0.874-0.954) | 0.091 | |
| | Aβ42/Aβ40 | 0.706 (0.626-0.787) | <0.0001 | | 0.751 (0.679-0.823) | <0.0001 | |
| Fujirebio Lumipulse | p-tau217 | 0.854 (0.799-0.910) | REFERENCE | 0.0011 | 0.843 (0.787-0.900) | 0.60 | |
| | p-tau217 + Aβ42/Aβ40 | 0.848 (0.792-0.905) | 0.82 | | 0.854 (0.801-0.907) | REFERENCE | 0.0004 |
| | Aβ42/Aβ40 | 0.738 (0.665-0.810) | 0.016 | | 0.784 (0.719-0.848) | 0.0003 | |
| ALZpath Quanterix | p-tau217 | 0.841 (0.777-0.906) | | 0.0004 | 0.861 (0.806-0.916) | | 0.0004 |
| Janssen LucentAD Quanterix | p-tau217 | 0.844 (0.785-0.903) | | 0.0004 | 0.860 (0.808-0.912) | | 0.0004 |
| Roche NeuroToolKit | p-tau181 + Aβ42/Aβ40 + GFAP + NfL | 0.832 (0.775-0.888) | REFERENCE | 0.0004 | 0.855 (0.802-0.907) | REFERENCE | 0.0007 |
| | p-tau181 + Aβ42/Aβ40 + NfL | 0.829 (0.773-0.885) | 0.77 | | 0.848 (0.795-0.900) | 0.38 | |
| | p-tau181 + Aβ42/Aβ40 | 0.829 (0.773-0.885) | 0.77 | | 0.847 (0.794-0.899) | 0.38 | |
| | p-tau181 | 0.765 (0.690-0.840) | 0.028 | | 0.807 (0.742-0.871) | 0.038 | |
| | Aβ42/Aβ40 | 0.746 (0.678-0.814) | 0.015 | | 0.788 (0.725-0.851) | 0.015 | |
| | GFAP | 0.731 (0.653-0.809) | 0.0047 | | 0.781 (0.713-0.848) | 0.0064 | |
| | NfL | 0.611 (0.520-0.703) | <0.0001 | | 0.725 (0.650-0.800) | 0.0004 | |
| Quanterix Neurology 4-Plex | p-tau181 + Aβ42/Aβ40 + GFAP + NfL | 0.777 (0.701-0.852) | REFERENCE | 0.0007 | 0.820 (0.753-0.886) | REFERENCE | 0.0040 |
| | p-tau181 + Aβ42/Aβ40 | 0.761 (0.680-0.842) | 0.30 | | 0.806 (0.734-0.878) | 0.32 | |
| | p-tau181 + Aβ42/Aβ40 + NfL | 0.759 (0.677-0.840) | 0.30 | | 0.803 (0.731-0.876) | 0.28 | |
| | Aβ42/Aβ40 | 0.711 (0.625-0.796) | 0.078 | | 0.764 (0.686-0.841) | 0.081 | |
| | GFAP | 0.708 (0.617-0.799) | 0.13 | | 0.763 (0.683-0.842) | 0.11 | |
| | p-tau181 | 0.703 (0.613-0.792) | 0.083 | | 0.758 (0.675-0.842) | 0.10 | |
| | NfL | 0.624 (0.518-0.730) | 0.030 | | 0.711 (0.624-0.799) | 0.041 | |

**Table S17. Classification accuracies of individual plasma biomarker analytes for amyloid PET status defined by Centiloid>37 or ≤37 in the cognitively unimpaired sub-cohort.** The receiver operating characteristics area under the curve (AUC) point estimate (midpoint) and 95% confidence intervals are shown for classification of amyloid PET status (> or ≤37 Centiloids) by plasma biomarker analytes. The single cut-off for the plasma biomarker that best distinguished amyloid PET status based on the Youden index is shown, as well as the positive percent agreement (PPA), negative percent agreement (NPA), overall accuracy, positive predictive value (PPV), and negative predictive value (NPV) of the cut-off for amyloid PET status in the cognitively unimpaired sub-cohort, which had a 28.5% rate of amyloid PET positivity based on a cut-off of >37 Centiloids.

| Company | Analyte | AUC | Cut-off | Brier Score | PPA | NPA | Accuracy | PPV | NPV |
|---|---|---|---|---|---|---|---|---|---|
| **C2N PrecivityAD2** | %p-tau217 | 0.928 (0.889-0.968) | 4.60 (%) | 0.092 | 0.877 | 0.881 | 0.880 | 0.746 | 0.947 |
| | p-tau217 | 0.903 (0.856-0.951) | 2.18 (pg/ml) | 0.111 | 0.895 | 0.811 | 0.835 | 0.654 | 0.951 |
| | Aβ42/Aβ40 | 0.706 (0.626-0.787) | 0.0924 | 0.183 | 0.702 | 0.629 | 0.650 | 0.430 | 0.841 |
| **Fujirebio Lumipulse** | p-tau217 | 0.854 (0.799-0.910) | 0.172 (pg/ml) | 0.147 | 0.737 | 0.818 | 0.795 | 0.618 | 0.886 |
| | Aβ42/Aβ40 | 0.738 (0.665-0.810) | 0.0886 | 0.176 | 0.719 | 0.664 | 0.680 | 0.461 | 0.856 |
| **Janssen LucentAD Quanterix** | p-tau217 | 0.844 (0.785-0.903) | 0.0535 (pg/ml) | 0.142 | 0.825 | 0.727 | 0.755 | 0.547 | 0.912 |
| **ALZpath Quanterix** | p-tau217 | 0.841 (0.777-0.906) | 0.449 (pg/ml) | 0.138 | 0.789 | 0.811 | 0.805 | 0.625 | 0.906 |
| **Roche NeuroToolKit** | p-tau181 | 0.765 (0.690-0.840) | 1.13 (pg/ml) | 0.166 | 0.702 | 0.720 | 0.715 | 0.500 | 0.858 |
| | Aβ42/Aβ40 | 0.746 (0.678-0.814) | 0.131 | 0.183 | 0.947 | 0.469 | 0.605 | 0.415 | 0.957 |
| | GFAP | 0.731 (0.653-0.809) | 0.146 (ng/ml) | 0.181 | 0.474 | 0.853 | 0.745 | 0.563 | 0.803 |
| | NfL | 0.611 (0.520-0.703) | 4.53 (pg/ml) | 0.190 | 0.439 | 0.755 | 0.665 | 0.417 | 0.771 |
| **Quanterix Neurology 4-Plex** | Aβ42/Aβ40 | 0.711 (0.625-0.796) | 0.0562 | 0.168 | 0.605 | 0.727 | 0.696 | 0.426 | 0.845 |
| | GFAP | 0.708 (0.617-0.799) | 212 (pg/ml) | 0.171 | 0.558 | 0.836 | 0.766 | 0.533 | 0.849 |
| | p-tau181 | 0.703 (0.613-0.792) | 20.5 (pg/ml) | 0.167 | 0.674 | 0.656 | 0.661 | 0.397 | 0.857 |
| | NfL | 0.624 (0.518-0.730) | 25.1 (pg/ml) | 0.175 | 0.558 | 0.711 | 0.673 | 0.393 | 0.827 |

**Table S18. Accuracy of published and/or recommended cut-offs for amyloid PET status defined by Centiloid>20 or ≤20 in the full cohort.** The single cut-off (**A**) for the plasma biomarker that best distinguished amyloid PET status based on the Youden index in the current study is shown, as well as the published and/or company-recommended single cut-off value. Two cut-offs (**B**) that correspond to 90% sensitivity or 90% specificity in the current study are shown, as well as the published and/or recommended two cut-off values. For each cut-off, values are shown for the positive percent agreement (PPA), negative percent agreement (NPA), overall accuracy, positive predictive value (PPV), and negative predictive value (NPV) of the cut-off for amyloid PET status in the full cohort, which had a 48.7% rate of amyloid PET positivity based on a cut-off of >20 Centiloids.

**A. Single cut-off**

| Company | Analyte | Source | Cutoff | PPA | NPA | Accuracy | PPV | NPV |
|---|---|---|---|---|---|---|---|---|
| **C2N PrecivityAD2** | %p-tau217 | Current study | 4.06 (%) | 0.885 | 0.871 | 0.878 | 0.867 | 0.888 |
| | | Meyer et al. 2024 | 4.2 (%) | 0.859 | 0.876 | 0.867 | 0.868 | 0.867 |
| | Aβ42/Aβ40 | Current study | 0.0924 | 0.717 | 0.672 | 0.694 | 0.675 | 0.714 |
| | | Meyer et al. 2024 | 0.089 | 0.581 | 0.771 | 0.679 | 0.707 | 0.660 |
| **ALZpath Quanterix** | p-tau217 | Current study | 0.444 (pg/ml) | 0.843 | 0.831 | 0.837 | 0.826 | 0.848 |
| | | Ashton et al. 2024 | 0.42 (pg/ml) | 0.848 | 0.801 | 0.824 | 0.802 | 0.847 |
| **Quanterix Neurology 4-Plex** | p-tau181 | Current study | 20.5 (pg/ml) | 0.717 | 0.694 | 0.705 | 0.671 | 0.738 |
| | | Recommended | 14.2 (pg/ml) | 0.912 | 0.355 | 0.614 | 0.551 | 0.823 |

**B. Two cut-offs**

| Company | Analyte | Source | High sensitivity cut-off | PPA | NPA | High specificity cut-off | PPA | NPA | Accuracy | PPV | NPV |
|---|---|---|---|---|---|---|---|---|---|---|---|
| **ALZpath Quanterix** | p-tau217 | Current study | 0.300 (pg/ml) | 0.901 | 0.552 | 0.540 (pg/ml) | 0.733 | 0.900 | 0.866 | 0.875 | 0.854 |
| | | Ashton et al. 2024 | 0.4 (pg/ml) | 0.864 | 0.756 | 0.63 (pg/ml) | 0.634 | 0.955 | 0.886 | 0.931 | 0.854 |
| **Janssen LucentAD Quanterix** | p-tau217 | Current study | 0.0495 (pg/ml) | 0.901 | 0.647 | 0.0735 (pg/ml) | 0.681 | 0.905 | 0.872 | 0.872 | 0.872 |
| | | Recommended | 0.04 (pg/ml) | 0.942 | 0.463 | 0.09 (pg/ml) | 0.550 | 0.945 | 0.901 | 0.907 | 0.894 |

**Table S19. Correlations between individual plasma biomarker analytes and amyloid PET Centiloid in the cognitively impaired sub-cohort.** The unadjusted Spearman correlation with amyloid PET Centiloid, and the partial Spearman correlation adjusting for age, sex, and *APOE* genotype, are shown with 95% confidence intervals. Correlations between the top performing analyte and other analytes were compared by bootstrapping.

| | | Unadjusted for covariates | | Adjusted for covariates | |
|---|---|---|---|---|---|
| Company | Analyte | Spearman rho | p= | Partial Spearman rho | p= |
| C2N PrecivityAD2 | %p-tau217 (%) | 0.792 (0.744 to 0.832) | REFERENCE | 0.780 (0.722 to 0.827) | 0.73 |
| | p-tau217 (pg/ml) | 0.792 (0.741 to 0.831) | 0.97 | 0.786 (0.735 to 0.825) | REFERENCE |
| | Aβ42/Aβ40 | -0.426 (-0.537 to -0.309) | <0.0001 | -0.398 (-0.518 to -0.262) | <0.0001 |
| Fujirebio Lumipulse | p-tau217 (pg/ml) | 0.792 (0.746 to 0.829) | 0.97 | 0.782 (0.729 to 0.821) | 0.82 |
| | Aβ42/Aβ40 | -0.482 (-0.594 to -0.351) | <0.0001 | -0.460 (-0.598 to -0.326) | <0.0001 |
| ALZpath Quanterix | p-tau217 (pg/ml) | 0.768 (0.714 to 0.807) | 0.35 | 0.771 (0.716 to 0.809) | 0.49 |
| Janssen LucentAD Quanterix | p-tau217 (pg/ml) | 0.744 (0.680 to 0.792) | 0.087 | 0.735 (0.675 to 0.784) | 0.033 |
| Roche NeuroToolKit | p-tau181 (pg/ml) | 0.613 (0.519 to 0.685) | <0.0001 | 0.618 (0.532 to 0.682) | <0.0001 |
| | Aβ42/Aβ40 | -0.521 (-0.636 to -0.412) | <0.0001 | -0.498 (-0.610 to -0.376) | <0.0001 |
| | GFAP (ng/ml) | 0.468 (0.349 to 0.570) | <0.0001 | 0.451 (0.329 to 0.554) | <0.0001 |
| | NfL (pg/mL) | 0.282 (0.145 to 0.403) | <0.0001 | 0.275 (0.138 to 0.403) | <0.0001 |
| Quanterix Neurology 4-Plex | p-tau181 (pg/ml) | 0.512 (0.401 to 0.606) | <0.0001 | 0.512 (0.400 to 0.602) | <0.0001 |
| | GFAP (pg/ml) | 0.492 (0.374 to 0.597) | <0.0001 | 0.462 (0.328 to 0.582) | <0.0001 |
| | Aβ42/Aβ40 | -0.384 (-0.506 to -0.248) | <0.0001 | -0.333 (-0.470 to -0.172) | <0.0001 |
| | NfL (pg/mL) | 0.299 (0.173 to 0.425) | <0.0001 | 0.266 (0.136 to 0.395) | <0.0001 |

**Table S20. Correlations between individual plasma biomarker analytes and amyloid PET Centiloid in the cognitively unimpaired sub-cohort.** The unadjusted Spearman correlation with amyloid PET Centiloid, and the partial Spearman correlation adjusting for age, sex, and *APOE* genotype, are shown with 95% confidence intervals. Correlations between the top performing analyte and other analytes were compared by bootstrapping.

| | | Unadjusted for covariates | | Adjusted for covariates | |
|---|---|---|---|---|---|
| **Company** | **Analyte** | **Spearman rho** | **p=** | **Partial Spearman rho** | **p=** |
| **C2N PrecivityAD2** | %p-tau217 (%) | 0.679 (0.592 to 0.756) | REFERENCE | 0.673 (0.579 to 0.740) | REFERENCE |
| | p-tau217 (pg/ml) | 0.650 (0.549 to 0.737) | 0.26 | 0.647 (0.539 to 0.720) | 0.31 |
| | Aβ42/Aβ40 | -0.418 (-0.538 to -0.300) | <0.0001 | -0.407 (-0.522 to -0.278) | <0.0001 |
| **Janssen LucentAD Quanterix** | p-tau217 (pg/ml) | 0.580 (0.477 to 0.683) | 0.030 | 0.576 (0.461 to 0.664) | 0.038 |
| **Fujirebio Lumipulse** | p-tau217 (pg/ml) | 0.563 (0.447 to 0.661) | 0.019 | 0.553 (0.443 to 0.646) | 0.020 |
| | Aβ42/Aβ40 | -0.474 (-0.569 to -0.362) | 0.0002 | -0.462 (-0.557 to -0.356) | 0.0005 |
| **ALZpath** | p-tau217 (pg/ml) | 0.546 (0.422 to 0.658) | 0.0078 | 0.544 (0.420 to 0.639) | 0.011 |
| **Roche NeuroToolKit** | Aβ42/Aβ40 | -0.484 (-0.584 to -0.371) | 0.0022 | -0.472 (-0.575 to -0.358) | 0.0032 |
| | p-tau181 (pg/ml) | 0.389 (0.253 to 0.525) | <0.0001 | 0.378 (0.242 to 0.506) | <0.0001 |
| | GFAP (ng/ml) | 0.305 (0.171 to 0.433) | <0.0001 | 0.288 (0.149 to 0.413) | <0.0001 |
| | NfL (pg/mL) | 0.141 (-0.012 to 0.291) | <0.0001 | 0.104 (-0.031 to 0.241) | <0.0001 |
| **Quanterix Neurology 4-Plex** | Aβ42/Aβ40 | -0.352 (-0.470 to -0.221) | <0.0001 | -0.320 (-0.446 to -0.193) | 0.0004 |
| | p-tau181 (pg/ml) | 0.293 (0.132 to 0.440) | <0.0001 | 0.278 (0.131 to 0.416) | <0.0001 |
| | GFAP (pg/ml) | 0.272 (0.107 to 0.417) | <0.0001 | 0.238 (0.087 to 0.389) | <0.0001 |
| | NfL (pg/mL) | 0.136 (-0.032 to 0.294) | <0.0001 | 0.091 (-0.073 to 0.245) | <0.0001 |

**Table S21. Correlations between individual plasma biomarker analytes and amyloid PET Centiloid in the sub-cohort with amyloid PET >20 Centiloids.** The unadjusted Spearman correlation with amyloid PET Centiloid, and the partial Spearman correlation adjusting for age, sex, and *APOE* genotype, are shown with 95% confidence intervals. Correlations between the top performing analyte and other analytes were compared by bootstrapping.

| | | Unadjusted for covariates | | Adjusted for covariates | |
|---|---|---|---|---|---|
| **Company** | **Analyte** | **Spearman rho** | **p=** | **Partial Spearman rho** | **p=** |
| **Fujirebio Lumipulse** | p-tau217 (pg/ml) | 0.518 (0.401 to 0.615) | REFERENCE | 0.514 (0.398 to 0.614) | 0.90 |
| | Aβ42/Aβ40 | -0.065 (-0.200 to 0.084) | <0.0001 | -0.069 (-0.212 to 0.072) | <0.0001 |
| **C2N PrecivityAD2** | p-tau217 (pg/ml) | 0.516 (0.395 to 0.612) | 0.95 | 0.518 (0.395 to 0.623) | REFERENCE |
| | %p-tau217 (%) | 0.494 (0.366 to 0.593) | 0.70 | 0.488 (0.358 to 0.601) | 0.48 |
| | Aβ42/Aβ40 | -0.071 (-0.212 to 0.071) | <0.0001 | -0.065 (-0.216 to 0.082) | <0.0001 |
| **Janssen LucentAD Quanterix** | p-tau217 (pg/ml) | 0.509 (0.393 to 0.606) | 0.83 | 0.505 (0.376 to 0.611) | 0.78 |
| **ALZpath Quanterix** | p-tau217 (pg/ml) | 0.496 (0.384 to 0.596) | 0.62 | 0.504 (0.391 to 0.607) | 0.78 |
| **Roche NeuroToolKit** | p-tau181 (pg/ml) | 0.369 (0.235 to 0.490) | 0.0007 | 0.386 (0.262 to 0.510) | 0.0025 |
| | GFAP (ng/ml) | 0.248 (0.098 to 0.371) | <0.0001 | 0.250 (0.106 to 0.395) | 0.0004 |
| | NfL (pg/mL) | 0.151 (0.009 to 0.269) | <0.0001 | 0.164 (0.033 to 0.300) | <0.0001 |
| | Aβ42/Aβ40 | -0.127 (-0.271 to 0.032) | <0.0001 | -0.103 (-0.248 to 0.037) | <0.0001 |
| **Quanterix Neurology 4-Plex** | p-tau181 (pg/ml) | 0.332 (0.202 to 0.448) | 0.0012 | 0.321 (0.175 to 0.449) | 0.0027 |
| | GFAP (pg/ml) | 0.231 (0.074 to 0.375) | <0.0001 | 0.219 (0.041 to 0.360) | 0.0003 |
| | NfL (pg/mL) | 0.174 (0.034 to 0.313) | <0.0001 | 0.162 (-0.002 to 0.304) | <0.0001 |
| | Aβ42/Aβ40 | -0.059 (-0.204 to 0.105) | <0.0001 | -0.025 (-0.193 to 0.118) | <0.0001 |

**Table S22. Correlations between individual plasma biomarker analytes and amyloid PET Centiloid in the sub-cohort with amyloid PET ≤20 Centiloids.** The unadjusted Spearman correlation with amyloid PET Centiloid, and the partial Spearman correlation adjusting for age, sex, and *APOE* genotype, are shown with 95% confidence intervals. Correlations between the top performing analyte and other analytes were compared by bootstrapping.

| | | Unadjusted for covariates | | Adjusted for covariates | |
|---|---|---|---|---|---|
| **Company** | **Analyte** | **Spearman rho** | **p=** | **Partial Spearman rho** | **p=** |
| **Roche NeuroToolKit** | Aβ42/Aβ40 | -0.349 (-0.472 to -0.211) | REFERENCE | -0.357 (-0.476 to -0.225) | REFERENCE |
| | NfL (pg/mL) | -0.134 (-0.276 to 0.008) | 0.030 | -0.103 (-0.234 to 0.037) | 0.010 |
| | GFAP (ng/ml) | -0.022 (-0.162 to 0.111) | 0.0006 | -0.001 (-0.133 to 0.131) | <0.0001 |
| | p-tau181 (pg/ml) | 0.016 (-0.124 to 0.159) | <0.0001 | 0.061 (-0.089 to 0.197) | 0.0045 |
| **Fujirebio Lumipulse bio** | Aβ42/Aβ40 | -0.248 (-0.372 to -0.109) | 0.072 | -0.258 (-0.386 to -0.127) | 0.12 |
| | p-tau217 (pg/ml) | 0.143 (-0.004 to 0.284) | 0.026 | 0.183 (0.053 to 0.314) | 0.076 |
| **C2N PrecivityAD2** | %p-tau217 (%) | 0.245 (0.102 to 0.379) | 0.17 | 0.281 (0.142 to 0.405) | 0.32 |
| | p-tau217 (pg/ml) | 0.207 (0.061 to 0.348) | 0.10 | 0.247 (0.102 to 0.373) | 0.22 |
| | Aβ42/Aβ40 | -0.193 (-0.334 to -0.042) | 0.054 | -0.215 (-0.361 to -0.066) | 0.089 |
| **Janssen LucentAD Quanterix** | p-tau217 (pg/ml) | 0.171 (0.029 to 0.309) | 0.068 | 0.228 (0.098 to 0.347) | 0.20 |
| **ALZpath Quanterix** | p-tau217 (pg/ml) | 0.169 (0.028 to 0.303) | 0.059 | 0.234 (0.093 to 0.352) | 0.20 |
| **Quanterix Neurology 4-Plex** | Aβ42/Aβ40 | -0.161 (-0.304 to -0.024) | 0.023 | -0.170 (-0.299 to -0.020) | 0.081 |
| | NfL (pg/mL) | -0.119 (-0.254 to 0.020) | 0.023 | -0.083 (-0.226 to 0.062) | 0.013 |
| | p-tau181 (pg/ml) | 0.013 (-0.134 to 0.161) | <0.0001 | 0.089 (-0.052 to 0.231) | 0.033 |
| | GFAP (pg/ml) | 0.006 (-0.124 to 0.150) | <0.0001 | 0.027 (-0.110 to 0.160) | 0.0051 |

**Table S23. Classification accuracies of individual and combined plasma biomarker analytes for early tau PET status in the full cohort.** The receiver operating characteristics area under the curve (AUC) point estimate (midpoint) and 95% confidence intervals are shown for classification of early tau PET status by individual or combined plasma biomarker analytes. Both the unadjusted AUC and the AUC adjusted for age, sex, and *APOE* genotype are provided. AUCs were compared using DeLong's test. The Benjamin-Hochberg procedure was used to adjust for multiple comparisons with the reference analyte, either within a company or across companies. The AUC for a model with covariates only was 0.767 (0.680-0.855).

| Company | Analytes | Unadjusted for covariates | | | Adjusted for covariates | | |
|---|---|---|---|---|---|---|---|
| | | AUC | Within-company comparisons p= | Across-company comparisons p= | AUC | Within-company comparisons p= | Across-company comparisons p= |
| C2N PrecivityAD2 | %p-tau217 | 0.888 (0.836-0.940) | REFERENCE | REFERENCE | 0.931 (0.889-0.974) | REFERENCE | REFERENCE |
| | %p-tau217+ Aβ42/Aβ40 | 0.888 (0.835-0.940) | 0.95 | | 0.930 (0.886-0.975) | 0.79 | |
| | p-tau217 | 0.884 (0.831-0.936) | 0.86 | | 0.926 (0.883-0.969) | 0.63 | |
| | p-tau217 + Aβ42/Aβ40 | 0.882 (0.828-0.936) | 0.86 | | 0.926 (0.883-0.969) | 0.63 | |
| | Aβ42/Aβ40 | 0.690 (0.599-0.780) | <0.0001 | | 0.805 (0.729-0.880) | 0.0011 | |
| Fujirebio Lumipulse | p-tau217 | 0.859 (0.802-0.916) | REFERENCE | 0.21 | 0.882 (0.822-0.942) | 0.70 | |
| | p-tau217 + Aβ42/Aβ40 | 0.834 (0.774-0.895) | 0.38 | | 0.887 (0.834-0.941) | REFERENCE | 0.12 |
| | Aβ42/Aβ40 | 0.745 (0.662-0.829) | 0.049 | | 0.830 (0.761-0.899) | 0.026 | |
| ALZpath Quanterix | p-tau217 | 0.850 (0.788-0.912) | | 0.21 | 0.905 (0.858-0.952) | | 0.21 |
| Janssen LucentAD Quanterix | p-tau217 | 0.846 (0.780-0.912) | | 0.21 | 0.895 (0.847-0.943) | | 0.12 |
| Roche NeuroToolKit | p-tau181 + Aβ42/Aβ40 + GFAP + NfL | 0.874 (0.823-0.925) | REFERENCE | 0.73 | 0.917 (0.873-0.961) | REFERENCE | 0.53 |
| | p-tau181 + Aβ42/Aβ40 + NfL | 0.836 (0.773-0.898) | 0.080 | | 0.898 (0.850-0.946) | 0.10 | |
| | p-tau181 + Aβ42/Aβ40 | 0.830 (0.768-0.892) | 0.069 | | 0.891 (0.840-0.941) | 0.050 | |
| | p-tau181 | 0.823 (0.755-0.891) | 0.095 | | 0.885 (0.832-0.939) | 0.034 | |
| | GFAP | 0.788 (0.716-0.861) | 0.011 | | 0.873 (0.812-0.935) | 0.024 | |
| | Aβ42/Aβ40 | 0.692 (0.609-0.775) | <0.0001 | | 0.795 (0.715-0.875) | 0.0012 | |
| | NfL | 0.621 (0.525-0.717) | <0.0001 | | 0.802 (0.725-0.879) | 0.0012 | |
| Quanterix Neurology 4-Plex | p-tau181 + Aβ42/Aβ40 + GFAP + NfL | 0.874 (0.819-0.930) | REFERENCE | 0.73 | 0.903 (0.849-0.957) | REFERENCE | 0.52 |
| | p-tau181 + Aβ42/Aβ40 | 0.849 (0.783-0.914) | 0.27 | | 0.89 (0.833-0.946) | 0.34 | |
| | p-tau181 + Aβ42/Aβ40 + NfL | 0.848 (0.783-0.914) | 0.27 | | 0.89 (0.832-0.947) | 0.34 | |
| | p-tau181 | 0.803 (0.724-0.883) | 0.10 | | 0.864 (0.796-0.932) | 0.078 | |
| | GFAP | 0.786 (0.703-0.870) | 0.036 | | 0.864 (0.791-0.938) | 0.078 | |
| | Aβ42/Aβ40 | 0.763 (0.673-0.852) | 0.013 | | 0.820 (0.741-0.900) | 0.078 | |
| | NfL | 0.675 (0.579-0.771) | 0.0004 | | 0.806 (0.721-0.890) | 0.018 | |

**Table S24. Classification accuracies of individual plasma biomarker analytes for early tau PET status in the full cohort.** The receiver operating characteristics area under the curve (AUC) point estimate (midpoint) and 95% confidence intervals are shown for classification of early tau PET status by plasma biomarker analytes. The single cut-off for the plasma biomarker that best distinguished early tau PET status based on the Youden index is shown, as well as the positive percent agreement (PPA), negative percent agreement (NPA), overall accuracy, positive predictive value (PPV), and negative predictive value (NPV) of the cut-off for early PET status in the full cohort, which had a 21.3% rate of early tau PET positivity.

| Company | Analyte | AUC | Cut-off | Brier Score | PPA | NPA | Accuracy | PPV | NPV |
|---|---|---|---|---|---|---|---|---|---|
| C2N PrecivityAD2 | %p-tau217 | 0.888 (0.836-0.940) | 5.60 (%) | 0.102 | 0.850 | 0.770 | 0.787 | 0.500 | 0.950 |
| | p-tau217 | 0.884 (0.831-0.936) | 2.94 (pg/ml) | 0.110 | 0.900 | 0.770 | 0.798 | 0.514 | 0.966 |
| | Aβ42/Aβ40 | 0.690 (0.599-0.780) | 0.0951 | 0.154 | 0.825 | 0.500 | 0.569 | 0.308 | 0.914 |
| Fujirebio Lumipulse | p-tau217 | 0.859 (0.802-0.916) | 0.177 (pg/ml) | 0.138 | 0.925 | 0.682 | 0.734 | 0.440 | 0.971 |
| | Aβ42/Aβ40 | 0.745 (0.662-0.829) | 0.0869 | 0.148 | 0.800 | 0.649 | 0.681 | 0.381 | 0.923 |
| ALZpath Quanterix | p-tau217 | 0.850 (0.788-0.912) | 0.559 (pg/ml) | 0.128 | 0.825 | 0.770 | 0.782 | 0.493 | 0.942 |
| Janssen LucentAD Quanterix | p-tau217 | 0.846 (0.780-0.912) | 0.0715 (pg/ml) | 0.128 | 0.825 | 0.743 | 0.761 | 0.465 | 0.940 |
| Roche NeuroToolKit | p-tau181 | 0.823 (0.755-0.891) | 1.16 (pg/ml) | 0.143 | 0.875 | 0.669 | 0.713 | 0.417 | 0.952 |
| | GFAP | 0.788 (0.716-0.861) | 0.115 (ng/ml) | 0.146 | 0.900 | 0.574 | 0.644 | 0.364 | 0.955 |
| | Aβ42/Aβ40 | 0.692 (0.609-0.775) | 0.130 | 0.157 | 0.950 | 0.378 | 0.500 | 0.292 | 0.966 |
| | NfL | 0.621 (0.525-0.717) | 4.43 (pg/ml) | 0.167 | 0.625 | 0.649 | 0.644 | 0.325 | 0.865 |
| Quanterix Neurology 4-Plex | p-tau181 | 0.803 (0.724-0.883) | 24.9 (pg/ml) | 0.141 | 0.750 | 0.752 | 0.752 | 0.429 | 0.924 |
| | GFAP | 0.786 (0.703-0.870) | 258 (pg/ml) | 0.136 | 0.625 | 0.868 | 0.820 | 0.541 | 0.903 |
| | Aβ42/Aβ40 | 0.763 (0.673-0.852) | 0.0556 | 0.145 | 0.781 | 0.690 | 0.708 | 0.385 | 0.927 |
| | NfL | 0.675 (0.579-0.771) | 19.3 (pg/ml) | 0.156 | 0.875 | 0.465 | 0.547 | 0.289 | 0.938 |

**Table S25. Classification accuracies of individual and combined plasma biomarker analytes for early tau PET status in the cognitively impaired sub-cohort.** The receiver operating characteristics area under the curve (AUC) point estimate (midpoint) and 95% confidence intervals are shown for classification of early tau PET status by individual or combined plasma biomarker analytes. Both the unadjusted AUC and the AUC adjusted for age, sex, and *APOE* genotype are provided. AUCs were compared using DeLong's test. The Benjamin-Hochberg procedure was used to adjust for multiple comparisons with the reference analyte, either within a company or across companies. The AUC for a model with covariates only was 0.736 (0.613-0.859).

| Company | Analytes | Unadjusted for covariates | | | Adjusted for covariates | | |
|---|---|---|---|---|---|---|---|
| | | AUC | Within-company comparisons p= | Across-company comparisons p= | AUC | Within-company comparisons p= | Across-company comparisons p= |
| C2N PrecivityAD2 | p-tau217 + Aβ42/Aβ40 | 0.932 (0.872-0.992) | REFERENCE | REFERENCE | 0.943 (0.887-0.999) | REFERENCE | REFERENCE |
| | %p-tau217+ Aβ42/Aβ40 | 0.924 (0.861-0.987) | 0.58 | | 0.933 (0.872-0.995) | 0.35 | |
| | p-tau217 | 0.923 (0.858-0.988) | 0.58 | | 0.937 (0.877-0.998) | 0.35 | |
| | %p-tau217 | 0.921 (0.857-0.985) | 0.58 | | 0.935 (0.873-0.997) | 0.35 | |
| | Aβ42/Aβ40 | 0.755 (0.639-0.870) | 0.0088 | | 0.808 (0.704-0.911) | 0.0232 | |
| Janssen LucentAD Quanterix | p-tau217 | 0.921 (0.855-0.986) | | 0.62 | 0.921 (0.857-0.986) | | 0.47 |
| Fujirebio Lumipulse | p-tau217 | 0.910 (0.845-0.975) | REFERENCE | 0.50 | 0.922 (0.858-0.986) | 0.54 | |
| | p-tau217 + Aβ42/Aβ40 | 0.906 (0.839-0.974) | 0.78 | | 0.926 (0.865-0.987) | REFERENCE | 0.47 |
| | Aβ42/Aβ40 | 0.744 (0.627-0.861) | 0.0089 | | 0.815 (0.713-0.917) | 0.0209 | |
| ALZpath Quanterix | p-tau217 | 0.901 (0.829-0.972) | | 0.50 | 0.930 (0.871-0.989) | | 0.56 |
| Roche NeuroToolKit | p-tau181 + Aβ42/Aβ40 + GFAP + NfL | 0.901 (0.829-0.974) | REFERENCE | 0.50 | 0.941 (0.886-0.996) | REFERENCE | 0.92 |
| | p-tau181 + Aβ42/Aβ40 + NfL | 0.897 (0.824-0.970) | 0.73 | | 0.941 (0.885-0.996) | 1.00 | |
| | p-tau181 + Aβ42/Aβ40 | 0.895 (0.820-0.970) | 0.73 | | 0.940 (0.885-0.995) | 1.00 | |
| | p-tau181 | 0.880 (0.802-0.957) | 0.50 | | 0.937 (0.880-0.994) | 0.83 | |
| | GFAP | 0.757 (0.645-0.869) | 0.0137 | | 0.830 (0.728-0.932) | 0.0298 | |
| | Aβ42/Aβ40 | 0.655 (0.528-0.783) | 0.0006 | | 0.763 (0.648-0.879) | 0.0058 | |
| | NfL | 0.617 (0.485-0.750) | 0.0003 | | 0.776 (0.665-0.888) | 0.0058 | |
| Quanterix Neurology 4-Plex | p-tau181 + Aβ42/Aβ40+ GFAP + NfL | 0.889 (0.809-0.968) | REFERENCE | 0.50 | 0.936 (0.876-0.996) | REFERENCE | 0.92 |
| | p-tau181 + Aβ42/Aβ40 + NfL | 0.875 (0.785-0.965) | 0.55 | | 0.932 (0.868-0.995) | 0.51 | |
| | p-tau181 + Aβ42/Aβ40 | 0.875 (0.786-0.964) | 0.55 | | 0.928 (0.863-0.994) | 0.42 | |
| | p-tau181 | 0.862 (0.771-0.953) | 0.37 | | 0.922 (0.856-0.988) | 0.28 | |
| | GFAP | 0.756 (0.637-0.874) | 0.032 | | 0.814 (0.704-0.924) | 0.035 | |
| | Aβ42/Aβ40 | 0.712 (0.579-0.844) | 0.03 | | 0.748 (0.623-0.873) | 0.0167 | |
| | NfL | 0.672 (0.535-0.809) | 0.0054 | | 0.793 (0.676-0.909) | 0.027 | |

**Table S26. Classification accuracies of individual plasma biomarker analytes for early tau PET status in the cognitively impaired sub-cohort.** The receiver operating characteristics area under the curve (AUC) point estimate (midpoint) and 95% confidence intervals are shown for classification of early tau PET status by plasma biomarker analytes. The single cut-off for the plasma biomarker that best distinguished early tau PET status based on the Youden index is shown, as well as the positive percent agreement (PPA), negative percent agreement (NPA), overall accuracy, positive predictive value (PPV), and negative predictive value (NPV) of the cut-off for early tau PET status in the cognitively impaired sub-cohort, which had a 48.7% rate of early tau PET positivity.

| Company | Analyte | AUC | Cut-off | Brier Score | PPA | NPA | Accuracy | PPV | NPV |
|---|---|---|---|---|---|---|---|---|---|
| **C2N PrecivityAD2** | p-tau217 | 0.923 (0.858-0.988) | 2.86 (pg/ml) | 0.119 | 0.966 | 0.814 | 0.875 | 0.778 | 0.972 |
| | %p-tau217 | 0.921 (0.857-0.985) | 7.21 (%) | 0.114 | 0.793 | 0.930 | 0.875 | 0.885 | 0.870 |
| | Aβ42/Aβ40 | 0.755 (0.639-0.870) | 0.0922 | 0.193 | 0.759 | 0.674 | 0.708 | 0.611 | 0.806 |
| **Janssen LucentAD Quanterix** | p-tau217 | 0.921 (0.855-0.986) | 0.0710 (pg/ml) | 0.126 | 0.966 | 0.791 | 0.861 | 0.757 | 0.971 |
| **Fujirebio Lumipulse** | p-tau217 | 0.910 (0.845-0.975) | 0.176 (pg/ml) | 0.137 | 1.000 | 0.698 | 0.819 | 0.690 | 1.000 |
| | Aβ42/Aβ40 | 0.744 (0.627-0.861) | 0.0818 | 0.201 | 0.621 | 0.767 | 0.708 | 0.643 | 0.750 |
| **ALZpath Quanterix** | p-tau217 | 0.901 (0.829-0.972) | 0.557 (pg/ml) | 0.140 | 0.931 | 0.791 | 0.847 | 0.750 | 0.944 |
| **Roche NeuroToolKit** | p-tau181 | 0.880 (0.802-0.957) | 1.14 (pg/ml) | 0.149 | 1.000 | 0.651 | 0.792 | 0.659 | 1.000 |
| | GFAP | 0.757 (0.645-0.869) | 0.103 (ng/ml) | 0.207 | 0.966 | 0.558 | 0.722 | 0.596 | 0.960 |
| | Aβ42/Aβ40 | 0.655 (0.528-0.783) | 0.117 | 0.223 | 0.586 | 0.674 | 0.639 | 0.548 | 0.707 |
| | NfL | 0.617 (0.485-0.750) | 4.47 (pg/ml) | 0.237 | 0.690 | 0.628 | 0.653 | 0.556 | 0.750 |
| **Quanterix Neurology 4-Plex** | p-tau181 | 0.862 (0.771-0.953) | 24.9 (pg/ml) | 0.159 | 0.792 | 0.795 | 0.794 | 0.704 | 0.861 |
| | GFAP | 0.756 (0.637-0.874) | 260 (pg/ml) | 0.200 | 0.667 | 0.795 | 0.746 | 0.667 | 0.795 |
| | Aβ42/Aβ40 | 0.712 (0.579-0.844) | 0.0567 | 0.222 | 0.833 | 0.615 | 0.698 | 0.571 | 0.857 |
| | NfL | 0.672 (0.535-0.809) | 23.0 (pg/ml) | 0.218 | 0.708 | 0.615 | 0.651 | 0.531 | 0.774 |

**Table S27. Classification accuracies of individual and combined plasma biomarker analytes for early tau PET status in the cognitively unimpaired sub-cohort.** The receiver operating characteristics area under the curve (AUC) point estimate (midpoint) and 95% confidence intervals are shown for classification of early tau PET status by individual or combined plasma biomarker analytes. Both the unadjusted AUC and the AUC adjusted for age, sex, and *APOE* genotype are provided. AUCs were compared using DeLong's test. The Benjamin-Hochberg procedure was used to adjust for multiple comparisons with the reference analyte, either within a company or across companies. The AUC for a model with covariates only was 0.883 (0.760-1.000).

| Company | Analytes | Unadjusted for covariates | | | Adjusted for covariates | | |
|---|---|---|---|---|---|---|---|
| | | AUC | Within-platform comparisons p= | Across-platform comparisons p= | AUC | Within-platform comparisons p= | Across-platform comparisons p= |
| Quanterix Neurology 4-Plex | p-tau181 + Aβ42/Aβ40 + GFAP + NfL | 0.882 (0.790-0.974) | REFERENCE | REFERENCE | 0.978 (0.938-1.000) | REFERENCE | REFERENCE |
| | p-tau181 + Aβ42/Aβ40 | 0.828 (0.718-0.938) | 0.11 | | 0.922 (0.854-0.991) | 0.0357 | |
| | p-tau181 + Aβ42/Aβ40 + NfL | 0.817 (0.704-0.929) | 0.11 | | 0.940 (0.870-1.000) | 0.0844 | |
| | Aβ42/Aβ40 | 0.812 (0.690-0.935) | 0.11 | | 0.921 (0.852-0.990) | 0.0357 | |
| | GFAP | 0.753 (0.565-0.940) | 0.16 | | 0.919 (0.815-1.000) | 0.092 | |
| | p-tau181 | 0.696 (0.483-0.909) | 0.16 | | 0.890 (0.769-1.000) | 0.0844 | |
| | NfL | 0.678 (0.506-0.849) | 0.11 | | 0.867 (0.710-1.000) | 0.0863 | |
| Roche NeuroToolKit | p-tau181 + Aβ42/Aβ40 + GFAP + NfL | 0.845 (0.759-0.931) | REFERENCE | 0.57 | 0.926 (0.825-1.000) | REFERENCE | 0.35 |
| | p-tau181 + Aβ42/Aβ40 + NfL | 0.744 (0.610-0.877) | 0.12 | | 0.904 (0.794-1.000) | 0.13 | |
| | Aβ42/Aβ40 | 0.739 (0.604-0.874) | 0.12 | | 0.882 (0.756-1.000) | 0.10 | |
| | GFAP | 0.732 (0.577-0.888) | 0.12 | | 0.922 (0.829-1.000) | 0.68 | |
| | p-tau181 + Aβ42/Aβ40 | 0.724 (0.606-0.842) | 0.0665 | | 0.888 (0.770-1.000) | 0.10 | |
| | p-tau181 | 0.668 (0.514-0.822) | 0.0967 | | 0.887 (0.769-1.000) | 0.10 | |
| | NfL | 0.554 (0.374-0.733) | 0.0293 | | 0.892 (0.762-1.000) | 0.13 | |
| C2N PrecivityAD2 | p-tau217 ratio | 0.805 (0.707-0.903) | REFERENCE | 0.33 | 0.932 (0.868-0.997) | 0.86 | |
| | p-tau217 ratio + Aβ42/Aβ40 | 0.799 (0.694-0.904) | 0.73 | | 0.934 (0.867-1.000) | 0.86 | |
| | p-tau217 + Aβ42/Aβ40 | 0.782 (0.677-0.887) | 0.26 | | 0.935 (0.880-0.990) | 0.84 | |
| | p-tau217 | 0.779 (0.673-0.885) | 0.26 | | 0.938 (0.884-0.991) | REFERENCE | 0.30 |
| | Aβ42/Aβ40 | 0.621 (0.455-0.787) | 0.0597 | | 0.897 (0.797-0.997) | 0.84 | |
| Fujirebio Lumipulse | p-tau217 | 0.741 (0.618-0.863) | REFERENCE | 0.13 | 0.887 (0.767-1.000) | 0.62 | |
| | p-tau217 + Aβ42/Aβ40 | 0.725 (0.586-0.863) | 0.88 | | 0.903 (0.812-0.994) | REFERENCE | 0.30 |
| | Aβ42/Aβ40 | 0.719 (0.572-0.867) | 0.88 | | 0.901 (0.807-0.996) | 0.62 | |
| ALZpath Quanterix | p-tau217 | 0.735 (0.601-0.870) | | 0.13 | 0.904 (0.803-1.000) | | 0.30 |
| Janssen LucentAD Quanterix | p-tau217 | 0.677 (0.522-0.833) | | 0.13 | 0.881 (0.755-1.000) | | 0.30 |

**Table S28. Classification accuracies of individual and combined plasma biomarker analytes for early tau PET status in the sub-cohort with amyloid PET >20 Centiloids.** The receiver operating characteristics area under the curve (AUC) point estimate (midpoint) and 95% confidence intervals are shown for classification of early tau PET status by individual or combined plasma biomarker analytes. Both the unadjusted AUC and the AUC adjusted for age, sex, and *APOE* genotype are provided. AUCs were compared using DeLong's test. The Benjamin-Hochberg procedure was used to adjust for multiple comparisons with the reference analyte, either within a company or across companies. The AUC for a model with covariates only was 0.688 (0.570-0.807).

| Company | Analytes | Unadjusted for covariates | | | Adjusted for covariates | | |
|---|---|---|---|---|---|---|---|
| | | AUC | Within-company comparisons p= | Across-company comparisons p= | AUC | Within-company comparisons p= | Across-company comparisons p= |
| Quanterix Neurology 4-Plex | p-tau181 + Aβ42/Aβ40 + NfL | 0.824 (0.724-0.924) | REFERENCE | REFERENCE | 0.847 (0.747-0.948) | REFERENCE | 0.62 |
| | p-tau181 + Aβ42/Aβ40 | 0.802 (0.697-0.907) | 0.34 | | 0.838 (0.737-0.940) | 0.32 | |
| | p-tau181 | 0.788 (0.684-0.893) | 0.29 | | 0.825 (0.721-0.930) | 0.24 | |
| | p-tau181 + Aβ42/Aβ40 + GFAP + NfL | 0.764 (0.652-0.875) | 0.23 | | 0.828 (0.734-0.921) | 0.54 | |
| | GFAP | 0.686 (0.559-0.813) | 0.12 | | 0.796 (0.694-0.898) | 0.37 | |
| | Aβ42/Aβ40 | 0.627 (0.495-0.759) | 0.0091 | | 0.705 (0.574-0.835) | 0.13 | |
| | NfL | 0.589 (0.456-0.723) | 0.0091 | | 0.697 (0.573-0.822) | 0.11 | |
| C2N PrecivityAD2 | %p-tau217+ Aβ42/Aβ40 | 0.805 (0.710-0.900) | REFERENCE | 0.79 | 0.879 (0.806-0.952) | REFERENCE | REFERENCE |
| | %p-tau217 | 0.792 (0.694-0.891) | 0.37 | | 0.872 (0.797-0.948) | 0.48 | |
| | p-tau217 + Aβ42/Aβ40 | 0.781 (0.681-0.880) | 0.32 | | 0.869 (0.792-0.945) | 0.48 | |
| | p-tau217 | 0.777 (0.677-0.878) | 0.32 | | 0.867 (0.790-0.943) | 0.48 | |
| | Aβ42/Aβ40 | 0.533 (0.409-0.657) | 0.0015 | | 0.694 (0.579-0.810) | 0.0049 | |
| Roche NeuroToolKit | p-tau181 + Aβ42/Aβ40 + GFAP + NfL | 0.799 (0.709-0.889) | REFERENCE | 0.79 | 0.850 (0.773-0.927) | REFERENCE | 0.55 |
| | p-tau181 + Aβ42/Aβ40 + NfL | 0.773 (0.674-0.872) | 0.48 | | 0.825 (0.740-0.911) | 0.30 | |
| | p-tau181 | 0.733 (0.627-0.838) | 0.27 | | 0.812 (0.724-0.901) | 0.24 | |
| | p-tau181 + Aβ42/Aβ40 | 0.724 (0.618-0.831) | 0.22 | | 0.814 (0.726-0.902) | 0.24 | |
| | GFAP | 0.675 (0.564-0.785) | 0.035 | | 0.801 (0.708-0.893) | 0.20 | |
| | Aβ42/Aβ40 | 0.590 (0.469-0.712) | 0.0058 | | 0.693 (0.575-0.812) | 0.017 | |
| | NfL | 0.505 (0.384-0.626) | 0.0009 | | 0.679 (0.560-0.798) | 0.017 | |
| Fujirebio Lumipulse | p-tau217 + Aβ42/Aβ40 | 0.747 (0.644-0.850) | REFERENCE | 0.49 | 0.823 (0.737-0.909) | REFERENCE | 0.062 |
| | p-tau217 | 0.745 (0.642-0.849) | 0.77 | | 0.823 (0.736-0.909) | 1.00 | |
| | Aβ42/Aβ40 | 0.576 (0.455-0.698) | 0.042 | | 0.721 (0.611-0.830) | 0.064 | |
| ALZpath Quanterix | p-tau217 | 0.745 (0.643-0.847) | | 0.49 | 0.828 (0.742-0.914) | | 0.12 |
| Janssen LucentAD Quanterix | p-tau217 | 0.744 (0.638-0.850) | | 0.49 | 0.814 (0.727-0.901) | | 0.062 |

**Table S29. Classification accuracies of individual plasma biomarker analytes for early tau PET status in the sub-cohort with amyloid PET >20 Centiloids.** The receiver operating characteristics area under the curve (AUC) point estimate (midpoint) and 95% confidence intervals are shown for classification of early tau PET status by plasma biomarker analytes. The single cut-off for the plasma biomarker that best distinguished early tau PET status based on the Youden index is shown, as well as the positive percent agreement (PPA), negative percent agreement (NPA), overall accuracy, positive percent agreement (PPA), negative percent agreement (NPA), overall accuracy, positive predictive value (PPV), and negative predictive value (NPV) of the cut-off for early tau PET status in the sub-cohort with amyloid PET >20 Centiloids, which had a 39.5% rate of early tau PET positivity.

| Company | Analyte | AUC | Cut-off | Brier Score | PPA | NPA | Accuracy | PPV | NPV |
|---|---|---|---|---|---|---|---|---|---|
| **C2N PrecivityAD2** | %p-tau217 | 0.792 (0.694-0.891) | 7.81 (%) | 0.168 | 0.694 | 0.818 | 0.769 | 0.714 | 0.804 |
| | p-tau217 | 0.777 (0.677-0.878) | 5.07 (pg/ml) | 0.182 | 0.639 | 0.855 | 0.769 | 0.742 | 0.783 |
| | Aβ42/Aβ40 | 0.533 (0.409-0.657) | 0.0787 | 0.238 | 0.222 | 0.891 | 0.626 | 0.571 | 0.636 |
| **Quanterix Neurology 4-Plex** | p-tau181 | 0.788 (0.684-0.893) | 23.6 (pg/ml) | 0.214 | 0.857 | 0.682 | 0.750 | 0.632 | 0.882 |
| | GFAP | 0.686 (0.559-0.813) | 258 (pg/ml) | 0.215 | 0.643 | 0.750 | 0.708 | 0.621 | 0.767 |
| | Aβ42/Aβ40 | 0.627 (0.495-0.759) | 0.0556 | 0.234 | 0.750 | 0.523 | 0.611 | 0.500 | 0.767 |
| | NfL | 0.589 (0.456-0.723) | 19.1 (pg/ml) | 0.237 | 0.857 | 0.386 | 0.569 | 0.471 | 0.810 |
| **Fujirebio Lumipulse** | p-tau217 | 0.745 (0.642-0.849) | 0.422 (pg/ml) | 0.207 | 0.556 | 0.855 | 0.736 | 0.714 | 0.746 |
| | Aβ42/Aβ40 | 0.576 (0.455-0.698) | 0.0882 | 0.237 | 0.806 | 0.400 | 0.560 | 0.468 | 0.759 |
| **ALZpath Quanterix** | p-tau217 | 0.745 (0.643-0.847) | 0.763 (pg/ml) | 0.207 | 0.694 | 0.709 | 0.703 | 0.610 | 0.780 |
| **Janssen LucentAD Quanterix** | p-tau217 | 0.744 (0.638-0.850) | 0.106 (pg/ml) | 0.207 | 0.694 | 0.800 | 0.758 | 0.694 | 0.800 |
| **Roche NeuroToolKit** | p-tau181 | 0.733 (0.627-0.838) | 1.47 (pg/ml) | 0.220 | 0.778 | 0.673 | 0.714 | 0.609 | 0.822 |
| | GFAP | 0.675 (0.564-0.785) | 0.157 (ng/ml) | 0.221 | 0.639 | 0.655 | 0.648 | 0.548 | 0.735 |
| | Aβ42/Aβ40 | 0.59 (0.469-0.712) | 0.111 | 0.232 | 0.500 | 0.636 | 0.582 | 0.474 | 0.660 |
| | NfL | 0.505 (0.384-0.626) | 3.70 (pg/ml) | 0.237 | 0.750 | 0.382 | 0.527 | 0.443 | 0.700 |

**Table S30. Classification accuracies of individual and combined plasma biomarker analytes for late tau PET status in the full cohort.** The receiver operating characteristics area under the curve (AUC) point estimate (midpoint) and 95% confidence intervals are shown for classification of late tau PET status by individual or combined plasma biomarker analytes. Both the unadjusted AUC and the AUC adjusted for age, sex, and *APOE* genotype are provided. AUCs were compared using DeLong's test. The Benjamin-Hochberg procedure was used to adjust for multiple comparisons with the reference analyte, either within a company or across companies. The AUC for a model with covariates only 0.667 (0.557-0.777).

| Company | Analytes | Unadjusted for covariates | | | Adjusted for covariates | | |
|---|---|---|---|---|---|---|---|
| | | AUC | Within-platform comparisons p= | Across-platform comparisons p= | AUC | Within-platform comparisons p= | Across-platform comparisons p= |
| C2N PrecivityAD2 | p-tau217 + Aβ42/Aβ40 | 0.901 (0.823-0.980) | REFERENCE | REFERENCE | 0.911 (0.835-0.986) | REFERENCE | REFERENCE |
| | p-tau217 | 0.896 (0.812-0.981) | 0.34 | | 0.911 (0.836-0.986) | 0.48 | |
| | %p-tau217 | 0.892 (0.803-0.981) | 0.34 | | 0.903 (0.828-0.979) | 0.46 | |
| | %p-tau217 + Aβ42/Aβ40 | 0.889 (0.797-0.982) | 0.34 | | 0.904 (0.828-0.980) | 0.46 | |
| | Aβ42/Aβ40 | 0.689 (0.584-0.795) | 0.0007 | | 0.754 (0.658-0.850) | 0.0056 | |
| Janssen LucentAD Quanterix | p-tau217 | 0.879 (0.796-0.961) | | 0.35 | 0.904 (0.831-0.977) | | 0.77 |
| Fujirebio Lumipulse | p-tau217 | 0.878 (0.797-0.958) | REFERENCE | 0.18 | 0.901 (0.829-0.974) | 0.62 | |
| | p-tau217 + Aβ42/Aβ40 | 0.858 (0.765-0.952) | 0.23 | | 0.903 (0.828-0.978) | REFERENCE | 0.77 |
| | Aβ42/Aβ40 | 0.678 (0.568-0.787) | 0.0001 | | 0.747 (0.654-0.841) | <0.0001 | |
| Roche NeuroToolKit | p-tau181 + Aβ42/Aβ40+ NfL | 0.869 (0.804-0.934) | REFERENCE | 0.35 | 0.898 (0.832-0.963) | REFERENCE | 0.77 |
| | p-tau181 + Aβ42/Aβ40 | 0.867 (0.798-0.935) | 0.82 | | 0.896 (0.830-0.962) | 0.78 | |
| | p-tau181 + Aβ42/Aβ40 + GFAP + NfL | 0.858 (0.785-0.930) | 0.51 | | 0.895 (0.826-0.964) | 0.81 | |
| | p-tau181 | 0.848 (0.762-0.934) | 0.53 | | 0.895 (0.828-0.961) | 0.78 | |
| | GFAP | 0.736 (0.625-0.848) | 0.019 | | 0.823 (0.735-0.910) | 0.029 | |
| | NfL | 0.667 (0.566-0.769) | 0.0001 | | 0.767 (0.680-0.853) | 0.0007 | |
| | Aβ42/Aβ40 | 0.636 (0.535-0.736) | <0.0001 | | 0.704 (0.607-0.801) | 0.0005 | |
| ALZpath Quanterix | p-tau217 | 0.843 (0.749-0.936) | | 0.046 | 0.891 (0.820-0.962) | | 0.77 |
| Quanterix Neurology 4-Plex | p-tau181 + Aβ42/Aβ40 + GFAP + NfL | 0.814 (0.717-0.910) | REFERENCE | 0.28 | 0.885 (0.811-0.960) | 0.88 | |
| | p-tau181 + Aβ42/Aβ40 | 0.797 (0.704-0.890) | 0.53 | | 0.888 (0.815-0.961) | REFERENCE | 0.77 |
| | p-tau181 + Aβ42/Aβ40 + NfL | 0.793 (0.699-0.888) | 0.53 | | 0.885 (0.812-0.958) | 0.78 | |
| | p-tau181 | 0.779 (0.683-0.875) | 0.49 | | 0.886 (0.813-0.959) | 0.88 | |
| | GFAP | 0.760 (0.647-0.872) | 0.089 | | 0.840 (0.748-0.931) | 0.41 | |
| | NfL | 0.670 (0.565-0.775) | 0.0084 | | 0.788 (0.699-0.877) | 0.020 | |
| | Aβ42/Aβ40 | 0.662 (0.553-0.770) | 0.024 | | 0.741 (0.639-0.843) | 0.020 | |

**Table S31. Classification accuracies of individual and combined plasma biomarker analytes for late tau PET status in the cognitively impaired sub-cohort.** The receiver operating characteristics area under the curve (AUC) point estimate (midpoint) and 95% confidence intervals are shown for classification of late tau PET status by individual or combined plasma biomarker analytes. Both the unadjusted AUC and the AUC adjusted for age, sex, and *APOE* genotype are provided. AUCs were compared using DeLong's test. The Benjamin-Hochberg procedure was used to adjust for multiple comparisons with the reference analyte, either within a company or across companies. The AUC for a model with covariates only 0.661 (0.520-0.803).

| Platform | Analytes | Unadjusted for covariates | | | Adjusted for covariates | | |
|---|---|---|---|---|---|---|---|
| | | AUC | Within-platform comparisons p= | Across-platform comparisons p= | AUC | Within-platform comparisons p= | Across-platform comparisons p= |
| C2N PrecivityAD2 | p-tau217 | 0.949 (0.892-1.000) | REFERENCE | REFERENCE | 0.948 (0.888-1.000) | REFERENCE | 1.00 |
| | p-tau217 + Aβ42/Aβ40 | 0.946 (0.884-1.000) | 0.60 | | 0.947 (0.886-1.000) | 0.69 | |
| | p-tau217 ratio + Aβ42/Aβ40 | 0.942 (0.881-1.000) | 0.59 | | 0.937 (0.868-1.000) | 0.60 | |
| | p-tau217 ratio | 0.941 (0.878-1.000) | 0.59 | | 0.938 (0.868-1.000) | 0.60 | |
| | Aβ42/Aβ40 | 0.704 (0.566-0.841) | 0.0008 | | 0.781 (0.660-0.902) | 0.0098 | |
| Fujirebio Lumipulse | p-tau217 + Aβ42/Aβ40 | 0.944 (0.889-0.999) | REFERENCE | 0.67 | 0.950 (0.887-1.000) | REFERENCE | REFERENCE |
| | p-tau217 | 0.941 (0.884-0.999) | 0.67 | | 0.944 (0.883-1.000) | 0.62 | |
| | Aβ42/Aβ40 | 0.689 (0.556-0.823) | <0.0001 | | 0.775 (0.661-0.888) | 0.0010 | |
| Janssen LucentAD Quanterix | p-tau217 | 0.931 (0.875-0.988) | | 0.65 | 0.938 (0.876-0.999) | | 1.00 |
| Roche NeuroToolKit | p-tau181 + Aβ42/Aβ40 + GFAP + NfL | 0.913 (0.848-0.978) | REFERENCE | 0.53 | 0.948 (0.901-0.994) | 0.73 | |
| | p-tau181 + Aβ42/Aβ40 + NfL | 0.913 (0.848-0.978) | 1.00 | | 0.949 (0.904-0.995) | 0.85 | |
| | p-tau181 + Aβ42/Aβ40 | 0.906 (0.836-0.976) | 0.56 | | 0.950 (0.905-0.996) | REFERENCE | 1.00 |
| | p-tau181 | 0.904 (0.833-0.975) | 0.56 | | 0.942 (0.892-0.992) | 0.30 | |
| | GFAP | 0.717 (0.592-0.843) | 0.0036 | | 0.823 (0.717-0.928) | 0.013 | |
| | NfL | 0.661 (0.529-0.792) | 0.0002 | | 0.785 (0.680-0.891) | 0.0045 | |
| | Aβ42/Aβ40 | 0.589 (0.451-0.728) | <0.0001 | | 0.692 (0.559-0.826) | 0.0009 | |
| ALZpath Quanterix | p-tau217 | 0.886 (0.806-0.967) | | 0.29 | 0.894 (0.818-0.969) | | 0.19 |
| Quanterix Neurology 4-Plex | p-tau181 + Aβ42/Aβ40 + GFAP + NfL | 0.881 (0.798-0.964) | REFERENCE | 0.48 | 0.948 (0.897-0.998) | REFERENCE | 1.00 |
| | p-tau181 + Aβ42/Aβ40 | 0.862 (0.772-0.951) | 0.35 | | 0.927 (0.867-0.988) | 0.10 | |
| | p-tau181 + Aβ42/Aβ40 + NfL | 0.859 (0.765-0.954) | 0.35 | | 0.946 (0.895-0.996) | 0.82 | |
| | p-tau181 | 0.855 (0.764-0.947) | 0.32 | | 0.927 (0.867-0.988) | 0.10 | |
| | GFAP | 0.782 (0.667-0.898) | 0.12 | | 0.855 (0.757-0.953) | 0.058 | |
| | NfL | 0.740 (0.611-0.870) | 0.044 | | 0.848 (0.755-0.941) | 0.058 | |
| | Aβ42/Aβ40 | 0.616 (0.471-0.760) | 0.0050 | | 0.717 (0.581-0.852) | 0.0075 | |

**Table S32. Classification accuracies of individual and combined plasma biomarker analytes for late tau PET status in the cognitively unimpaired sub-cohort.** The receiver operating characteristics area under the curve (AUC) point estimate (midpoint) and 95% confidence intervals are shown for classification of late tau PET status by individual or combined plasma biomarker analytes. Both the unadjusted AUC and the AUC adjusted for age, sex, and *APOE* genotype are provided. AUCs were compared using DeLong's test. The Benjamin-Hochberg procedure was used to adjust for multiple comparisons with the reference analyte, either within a company or across companies. The AUC for a model with covariates only 0.793 (0.616-0.971).

| Platform | Analytes | Unadjusted for covariates | | | Adjusted for covariates | | |
|---|---|---|---|---|---|---|---|
| | | AUC | Within-platform comparisons p= | Across-platform comparisons p= | AUC | Within-platform comparisons p= | Across-platform comparisons p= |
| C2N PrecivityAD2 | p-tau217 + Aβ42/Aβ40 | 0.761 (0.505-1.000) | REFERENCE | REFERENCE | 0.832 (0.590-1.000) | REFERENCE | 0.85 |
| | p-tau217 ratio + Aβ42/Aβ40 | 0.730 (0.448-1.000) | 0.27 | | 0.820 (0.577-1.000) | 0.70 | |
| | Aβ42/Aβ40 | 0.717 (0.545-0.888) | 0.69 | | 0.814 (0.650-0.978) | 0.77 | |
| | p-tau217 ratio | 0.703 (0.367-1.000) | 0.27 | | 0.817 (0.568-1.000) | 0.70 | |
| | p-tau217 | 0.702 (0.375-1.000) | 0.27 | | 0.826 (0.566-1.000) | 0.77 | |
| Roche NeuroToolKit | p-tau181 + Aβ42/Aβ40 | 0.708 (0.516-0.899) | REFERENCE | 0.76 | 0.808 (0.613-1.000) | 0.69 | |
| | p-tau181 + Aβ42/Aβ40 + NfL | 0.703 (0.497-0.909) | 0.77 | | 0.817 (0.636-0.997) | 0.69 | |
| | p-tau181 + Aβ42/Aβ40 + GFAP + NfL | 0.702 (0.490-0.913) | 0.77 | | 0.818 (0.640-0.996) | REFERENCE | 0.85 |
| | Aβ42/Aβ40 | 0.685 (0.465-0.905) | 0.77 | | 0.811 (0.634-0.987) | 0.69 | |
| | p-tau181 | 0.633 (0.318-0.949) | 0.77 | | 0.792 (0.574-1.000) | 0.69 | |
| | GFAP | 0.574 (0.228-0.920) | 0.77 | | 0.794 (0.616-0.972) | 0.69 | |
| | NfL | 0.555 (0.311-0.800) | 0.77 | | 0.800 (0.627-0.973) | 0.69 | |
| Quanterix Neurology 4-Plex | p-tau181 + Aβ42/Aβ40 + GFAP + NfL | 0.708 (0.491-0.924) | REFERENCE | 0.76 | 0.860 (0.687-1.000) | REFERENCE | REFERENCE |
| | p-tau181 + Aβ42/Aβ40 + NfL | 0.697 (0.482-0.912) | 0.41 | | 0.852 (0.668-1.000) | 0.33 | |
| | Aβ42/Aβ40 | 0.622 (0.372-0.871) | 0.19 | | 0.819 (0.590-1.000) | 0.33 | |
| | p-tau181 + Aβ42/Aβ40 | 0.619 (0.372-0.866) | 0.19 | | 0.811 (0.556-1.000) | 0.33 | |
| | NfL | 0.531 (0.309-0.753) | 0.37 | | 0.770 (0.528-1.000) | 0.30 | |
| | GFAP | 0.514 (0.131-0.897) | 0.37 | | 0.781 (0.537-1.000) | 0.30 | |
| | p-tau181 | 0.449 (0.130-0.768) | 0.37 | | 0.783 (0.468-1.000) | 0.33 | |
| Janssen LucentAD Quanterix | p-tau217 | 0.677 (0.351-1.000) | | 0.23 | 0.802 (0.550-1.000) | | 0.85 |
| Fujirebio Lumipulse | p-tau217 | 0.673 (0.377-0.970) | REFERENCE | 0.083 | 0.802 (0.624-0.979) | 0.73 | |
| | p-tau217 + Aβ42/Aβ40 | 0.641 (0.310-0.972) | 0.41 | | 0.809 (0.654-0.964) | REFERENCE | 0.85 |
| | Aβ42/Aβ40 | 0.536 (0.290-0.783) | 0.13 | | 0.794 (0.626-0.962) | 0.73 | |
| ALZpath Quanterix | p-tau217 | 0.613 (0.256-0.969) | | 0.083 | 0.808 (0.551-1.000) | | 0.85 |

**Table S33. Classification accuracies of individual and combined plasma biomarker analytes for late tau PET status in the sub-cohort with amyloid PET >20 Centiloids.** The receiver operating characteristics area under the curve (AUC) point estimate (midpoint) and 95% confidence intervals are shown for classification of late tau PET status by individual or combined plasma biomarker analytes. Both the unadjusted AUC and the AUC adjusted for age, sex, and *APOE* genotype are provided. AUCs were compared using DeLong's test. The Benjamin-Hochberg procedure was used to adjust for multiple comparisons with the reference analyte, either within a company or across companies. The AUC for a model with covariates only 0.660 (0.532-0.788).

| Platform | Analytes | Unadjusted for covariates | | | Adjusted for covariates | | |
|---|---|---|---|---|---|---|---|
| | | AUC | Within-platform comparisons p= | Across-platform comparisons p= | AUC | Within-platform comparisons p= | Across-platform comparisons p= |
| C2N PrecivityAD2 | p-tau217 ratio | 0.888 (0.788-0.989) | REFERENCE | REFERENCE | 0.888 (0.794-0.982) | 0.36 | |
| | p-tau217 + Aβ42/Aβ40 | 0.885 (0.788-0.982) | 0.83 | | 0.904 (0.817-0.991) | REFERENCE | REFERENCE |
| | p-tau217 ratio + Aβ42/Aβ40 | 0.884 (0.781-0.987) | 0.67 | | 0.891 (0.798-0.985) | 0.36 | |
| | p-tau217 | 0.881 (0.781-0.980) | 0.78 | | 0.903 (0.815-0.991) | 0.73 | |
| | Aβ42/Aβ40 | 0.589 (0.459-0.718) | 0.0003 | | 0.705 (0.577-0.833) | 0.016 | |
| Fujirebio Lumipulse | p-tau217 + Aβ42/Aβ40 | 0.855 (0.769-0.940) | REFERENCE | 0.36 | 0.881 (0.801-0.960) | REFERENCE | 0.48 |
| | p-tau217 | 0.835 (0.731-0.940) | 0.32 | | 0.867 (0.779-0.956) | 0.23 | |
| | Aβ42/Aβ40 | 0.546 (0.412-0.681) | <0.0001 | | 0.671 (0.544-0.797) | 0.0012 | |
| Janssen LucentAD Quanterix | p-tau217 | 0.818 (0.710-0.925) | | 0.0450 | 0.874 (0.783-0.966) | | 0.29 |
| Roche NeuroToolKit | p-tau181 + Aβ42/Aβ40 | 0.791 (0.687-0.895) | REFERENCE | 0.0817 | 0.850 (0.761-0.938) | REFERENCE | 0.29 |
| | p-tau181 + Aβ42/Aβ40 + GFAP + NfL | 0.784 (0.680-0.888) | 0.84 | | 0.845 (0.752-0.939) | 0.82 | |
| | p-tau181 + Aβ42/Aβ40 + NfL | 0.778 (0.673-0.882) | 0.84 | | 0.843 (0.752-0.933) | 0.68 | |
| | p-tau181 | 0.775 (0.660-0.889) | 0.79 | | 0.848 (0.759-0.937) | 0.76 | |
| | GFAP | 0.679 (0.553-0.804) | 0.096 | | 0.791 (0.683-0.899) | 0.30 | |
| | NfL | 0.592 (0.468-0.715) | 0.0006 | | 0.704 (0.587-0.821) | 0.0088 | |
| | Aβ42/Aβ40 | 0.528 (0.394-0.662) | 0.0029 | | 0.671 (0.542-0.799) | 0.0088 | |
| ALZpath Quanterix | p-tau217 | 0.773 (0.658-0.888) | | 0.033 | 0.832 (0.737-0.927) | | 0.28 |
| Quanterix Neurology 4-Plex | p-tau181 + Aβ42/Aβ40 + GFAP + NfL | 0.733 (0.606-0.859) | REFERENCE | 0.082 | 0.857 (0.760-0.954) | REFERENCE | 0.48 |
| | GFAP | 0.723 (0.590-0.855) | 0.72 | | 0.826 (0.721-0.930) | 0.19 | |
| | p-tau181 | 0.693 (0.565-0.821) | 0.59 | | 0.837 (0.734-0.940) | 0.56 | |
| | p-tau181 + Aβ42/Aβ40 | 0.683 (0.554-0.812) | 0.59 | | 0.831 (0.726-0.936) | 0.56 | |
| | p-tau181 + Aβ42/Aβ40+ NfL | 0.680 (0.550-0.809) | 0.59 | | 0.826 (0.721-0.930) | 0.51 | |
| | NfL | 0.612 (0.477-0.747) | 0.20 | | 0.761 (0.646-0.877) | 0.13 | |
| | Aβ42/Aβ40 | 0.525 (0.384-0.667) | 0.14 | | 0.691 (0.555-0.827) | 0.086 | |

**Table S34. Correlations between individual plasma biomarker analytes and early tau PET in the full cohort.** The unadjusted Spearman correlation with the early tau PET measure, and the partial Spearman correlation adjusting for age, sex, and *APOE* genotype, are shown with 95% confidence intervals. Correlations between the top performing analyte and other analytes were compared by bootstrapping.

| | | Unadjusted for covariates | | Adjusted for covariates | |
|---|---|---|---|---|---|
| **Company** | **Analyte** | **Spearman rho** | **p=** | **Partial Spearman rho** | **p=** |
| **C2N PrecivityAD2** | p-tau217 (pg/ml) | 0.579 (0.468 to 0.666) | REFERENCE | 0.562 (0.442 to 0.665) | REFERENCE |
| | %p-tau217 (%) | 0.559 (0.449 to 0.649) | 0.36 | 0.541 (0.426 to 0.640) | 0.35 |
| | Aβ42/Aβ40 | -0.264 (-0.401 to -0.121) | <0.0001 | -0.241 (-0.383 to -0.096) | <0.0001 |
| **Fujirebio Lumipulse** | p-tau217 (pg/ml) | 0.526 (0.416 to 0.620) | 0.11 | 0.506 (0.387 to 0.604) | 0.14 |
| | Aβ42/Aβ40 | -0.303 (-0.442 to -0.164) | <0.0001 | -0.282 (-0.419 to -0.147) | <0.0001 |
| **Janssen LucentAD Quanterix** | p-tau217 (pg/ml) | 0.499 (0.384 to 0.594) | 0.022 | 0.485 (0.353 to 0.593) | 0.053 |
| **ALZpath Quanterix** | p-tau217 (pg/ml) | 0.490 (0.371 to 0.586) | 0.011 | 0.489 (0.366 to 0.589) | 0.042 |
| **Roche NeuroToolKit** | p-tau181 (pg/ml) | 0.459 (0.341 to 0.568) | 0.0061 | 0.448 (0.314 to 0.561) | 0.018 |
| | GFAP (ng/ml) | 0.399 (0.267 to 0.512) | 0.0016 | 0.382 (0.225 to 0.506) | 0.0040 |
| | Aβ42/Aβ40 | -0.224 (-0.348 to -0.086) | <0.0001 | -0.205 (-0.330 to -0.060) | <0.0001 |
| | NfL (pg/mL) | 0.147 (0.012 to 0.289) | <0.0001 | 0.121 (-0.043 to 0.264) | <0.0001 |
| **Quanterix Neurology 4-Plex** | GFAP (pg/ml) | 0.418 (0.276 to 0.543) | 0.013 | 0.393 (0.227 to 0.548) | 0.018 |
| | p-tau181 (pg/ml) | 0.411 (0.266 to 0.535) | 0.013 | 0.391 (0.248 to 0.528) | 0.018 |
| | Aβ42/Aβ40 | -0.293 (-0.432 to -0.151) | 0.0004 | -0.252 (-0.410 to -0.094) | 0.0004 |
| | NfL (pg/mL) | 0.227 (0.081 to 0.371) | <0.0001 | 0.178 (0.017 to 0.346) | <0.0001 |

**Table S35. Correlations between individual plasma biomarker analytes and early tau PET in the cognitively impaired sub-cohort.** The unadjusted Spearman correlation with the early tau PET measure, and the partial Spearman correlation adjusting for age, sex, and *APOE* genotype, are shown with 95% confidence intervals. Correlations between the top performing analyte and other analytes were compared by bootstrapping.

| | | Unadjusted for covariates | | Adjusted for covariates | |
|---|---|---|---|---|---|
| **Company** | **Analyte** | **Spearman rho** | **p=** | **Partial Spearman rho** | **p=** |
| **C2N PrecivityAD2** | %p-tau217 (%) | 0.772 (0.661 to 0.836) | REFERENCE | 0.763 (0.607 to 0.843) | REFERENCE |
| | p-tau217 (pg/ml) | 0.755 (0.626 to 0.833) | 0.52 | 0.746 (0.581 to 0.840) | 0.56 |
| | Aβ42/Aβ40 | -0.442 (-0.616 to -0.233) | 0.0032 | -0.397 (-0.601 to -0.131) | 0.0036 |
| **Fujirebio Lumipulse** | p-tau217 (pg/ml) | 0.726 (0.591 to 0.818) | 0.28 | 0.716 (0.548 to 0.821) | 0.30 |
| | Aβ42/Aβ40 | -0.406 (-0.586 to -0.185) | 0.0025 | -0.373 (-0.583 to -0.145) | 0.0011 |
| **Janssen LucentAD Quanterix** | p-tau217 (pg/ml) | 0.710 (0.570 to 0.800) | 0.21 | 0.700 (0.544 to 0.798) | 0.28 |
| **ALZpath Quanterix** | p-tau217 (pg/ml) | 0.682 (0.543 to 0.772) | 0.065 | 0.690 (0.547 to 0.788) | 0.14 |
| **Roche NeuroToolKit** | p-tau181 (pg/ml) | 0.635 (0.470 to 0.748) | 0.049 | 0.647 (0.490 to 0.767) | 0.14 |
| | GFAP (ng/ml) | 0.451 (0.243 to 0.609) | 0.0032 | 0.447 (0.222 to 0.632) | 0.0039 |
| | Aβ42/Aβ40 | -0.298 (-0.516 to -0.054) | <0.0001 | -0.269 (-0.503 to -0.006) | <0.0001 |
| | NfL (pg/mL) | 0.185 (-0.043 to 0.396) | <0.0001 | 0.189 (-0.047 to 0.419) | <0.0001 |
| **Quanterix Neurology 4-Plex** | p-tau181 (pg/ml) | 0.583 (0.397 to 0.727) | 0.049 | 0.619 (0.407 to 0.755) | 0.062 |
| | GFAP (pg/ml) | 0.462 (0.243 to 0.641) | 0.0077 | 0.468 (0.222 to 0.650) | 0.0036 |
| | Aβ42/Aβ40 | -0.313 (-0.548 to -0.048) | <0.0001 | -0.260 (-0.502 to 0.034) | <0.0001 |
| | NfL (pg/mL) | 0.292 (0.040 to 0.519) | <0.0001 | 0.309 (0.048 to 0.535) | <0.0001 |

**Table S36. Correlations between individual plasma biomarker analytes and early tau PET in the cognitively unimpaired sub-cohort.** The unadjusted Spearman correlation with the early tau PET measure, and the partial Spearman correlation adjusting for age, sex, and *APOE* genotype, are shown with 95% confidence intervals. Correlations between the top performing analyte and other analytes were compared by bootstrapping.

| | | Unadjusted for covariates | | Adjusted for covariates | |
|---|---|---|---|---|---|
| **Company** | **Analyte** | **Spearman rho** | **p=** | **Partial Spearman rho** | **p=** |
| **C2N PrecivityAD2** | p-tau217 (pg/ml) | 0.401 (0.249 to 0.548) | REFERENCE | 0.380 (0.210 to 0.532) | REFERENCE |
| | p-tau217 ratio (%) | 0.352 (0.194 to 0.496) | 0.15 | 0.327 (0.152 to 0.483) | 0.18 |
| | Aβ42/Aβ40 | -0.114 (-0.296 to 0.079) | 0.011 | -0.110 (-0.289 to 0.088) | 0.017 |
| **Fujirebio Lumipulse** | p-tau217 (pg/ml) | 0.324 (0.154 to 0.473) | 0.18 | 0.296 (0.120 to 0.445) | 0.22 |
| | Aβ42/Aβ40 | -0.189 (-0.350 to -0.025) | 0.030 | -0.176 (-0.347 to 0.000) | 0.035 |
| **Quanterix Neurology 4-Plex** | GFAP (pg/ml) | 0.315 (0.125 to 0.504) | 0.36 | 0.273 (0.061 to 0.470) | 0.32 |
| | p-tau181 (pg/ml) | 0.250 (0.039 to 0.433) | 0.15 | 0.228 (0.018 to 0.441) | 0.19 |
| | Aβ42/Aβ40 | -0.165 (-0.359 to 0.044) | 0.030 | -0.128 (-0.334 to 0.086) | 0.035 |
| | NfL (pg/mL) | 0.151 (-0.069 to 0.359) | 0.030 | 0.090 (-0.123 to 0.298) | 0.035 |
| **ALZpath Quanterix** | p-tau217 (pg/ml) | 0.282 (0.112 to 0.443) | 0.061 | 0.274 (0.093 to 0.426) | 0.12 |
| **Janssen LucentAD Quanterix** | p-tau217 (pg/ml) | 0.281 (0.113 to 0.436) | 0.065 | 0.267 (0.086 to 0.417) | 0.12 |
| **Roche NeuroToolKit** | GFAP (ng/ml) | 0.265 (0.088 to 0.439) | 0.14 | 0.239 (0.035 to 0.412) | 0.18 |
| | p-tau181 (pg/ml) | 0.238 (0.062 to 0.403) | 0.030 | 0.212 (0.027 to 0.373) | 0.042 |
| | Aβ42/Aβ40 | -0.155 (-0.337 to 0.026) | 0.030 | -0.141 (-0.326 to 0.037) | 0.035 |
| | NfL (pg/mL) | 0.045 (-0.144 to 0.245) | 0.0067 | 0.011 (-0.190 to 0.203) | 0.017 |

**Table S37. Correlations between individual plasma biomarker analytes and early tau PET in the sub-cohort with amyloid PET >20 Centiloids.** The unadjusted Spearman correlation with the early tau PET measure, and the partial Spearman correlation adjusting for age, sex, and *APOE* genotype, are shown with 95% confidence intervals. Correlations between the top performing analyte and other analytes were compared by bootstrapping.

| | | Unadjusted for covariates | | Adjusted for covariates | |
|---|---|---|---|---|---|
| **Company** | **Analyte** | **Spearman rho** | **p=** | **Partial Spearman rho** | **p=** |
| **C2N PrecivityAD2** | p-tau217 (pg/ml) | 0.600 (0.450 to 0.719) | REFERENCE | 0.625 (0.439 to 0.751) | REFERENCE |
| | %p-tau217 (%) | 0.590 (0.421 to 0.716) | 0.80 | 0.604 (0.422 to 0.733) | 0.55 |
| | Aβ42/Aβ40 | -0.090 (-0.307 to 0.122) | <0.0001 | -0.081 (-0.314 to 0.154) | <0.0001 |
| **Janssen LucentAD Quanterix** | p-tau217 (pg/ml) | 0.545 (0.391 to 0.676) | 0.28 | 0.550 (0.369 to 0.686) | 0.15 |
| **Fujirebio Lumipulse** | p-tau217 (pg/ml) | 0.543 (0.395 to 0.663) | 0.28 | 0.536 (0.359 to 0.676) | 0.13 |
| | Aβ42/Aβ40 | -0.168 (-0.389 to 0.043) | <0.0001 | -0.183 (-0.397 to 0.044) | 0.0007 |
| **Quanterix Neurology 4-Plex** | p-tau181 (pg/ml) | 0.536 (0.391 to 0.651) | 0.53 | 0.542 (0.378 to 0.665) | 0.083 |
| | GFAP (pg/ml) | 0.416 (0.223 to 0.566) | 0.056 | 0.484 (0.259 to 0.639) | 0.017 |
| | Aβ42/Aβ40 | -0.255 (-0.449 to -0.019) | 0.015 | -0.238 (-0.462 to -0.012) | 0.0015 |
| | NfL (pg/mL) | 0.231 (0.009 to 0.420) | 0.0013 | 0.272 (0.044 to 0.474) | <0.0001 |
| **ALZpath Quanterix** | p-tau217 (pg/ml) | 0.521 (0.377 to 0.630) | 0.12 | 0.558 (0.379 to 0.679) | 0.15 |
| **Roche NeuroToolKit** | p-tau181 (pg/ml) | 0.458 (0.297 to 0.594) | 0.023 | 0.515 (0.334 to 0.642) | 0.079 |
| | GFAP (ng/ml) | 0.388 (0.225 to 0.525) | 0.019 | 0.451 (0.250 to 0.600) | 0.033 |
| | Aβ42/Aβ40 | -0.127 (-0.319 to 0.082) | 0.0006 | -0.051 (-0.268 to 0.184) | <0.0001 |
| | NfL (pg/mL) | 0.081 (-0.123 to 0.253) | <0.0001 | 0.152 (-0.058 to 0.341) | <0.0001 |

**Table S38. Correlations between individual plasma biomarker analytes and late tau PET in the full cohort.** The unadjusted Spearman correlation with the late tau PET measure, and the partial Spearman correlation adjusting for age, sex, and *APOE* genotype, are shown with 95% confidence intervals. Correlations between the top performing analyte and other analytes were compared by bootstrapping.

| | | Unadjusted for covariates | | Adjusted for covariates | |
|---|---|---|---|---|---|
| **Company** | **Analyte** | **Spearman rho** | **p=** | **Partial Spearman rho** | **p=** |
| **C2N PrecivityAD2** | p-tau217 (pg/ml) | 0.377 (0.243 to 0.507) | REFERENCE | 0.387 (0.241 to 0.511) | REFERENCE |
| | %p-tau217 (%) | 0.354 (0.215 to 0.481) | 0.32 | 0.362 (0.207 to 0.492) | 0.28 |
| | Aβ42/Aβ40 | -0.110 (-0.245 to 0.028) | <0.0001 | -0.106 (-0.253 to 0.034) | 0.0015 |
| **Fujirebio Lumipulse** | p-tau217 (pg/ml) | 0.327 (0.187 to 0.456) | 0.11 | 0.347 (0.198 to 0.481) | 0.23 |
| | Aβ42/Aβ40 | -0.130 (-0.271 to 0.012) | 0.0014 | -0.131 (-0.270 to 0.021) | <0.0001 |
| **Quanterix Neurology 4-Plex** | GFAP (pg/ml) | 0.295 (0.147 to 0.431) | 0.23 | 0.272 (0.097 to 0.430) | 0.13 |
| | p-tau181 (pg/ml) | 0.205 (0.050 to 0.360) | 0.018 | 0.228 (0.071 to 0.393) | 0.046 |
| | Aβ42/Aβ40 | -0.164 (-0.322 to -0.004) | 0.016 | -0.151 (-0.310 to -0.001) | 0.020 |
| | NfL (pg/mL) | 0.128 (-0.015 to 0.276) | 0.0019 | 0.109 (-0.050 to 0.275) | 0.0018 |
| **Janssen LucentAD Quanterix** | p-tau217 (pg/ml) | 0.276 (0.131 to 0.412) | 0.010 | 0.291 (0.133 to 0.435) | 0.021 |
| **Roche NeuroToolKit** | p-tau181 (pg/ml) | 0.269 (0.130 to 0.409) | 0.018 | 0.285 (0.121 to 0.430) | 0.036 |
| | GFAP (ng/ml) | 0.257 (0.109 to 0.394) | 0.062 | 0.258 (0.075 to 0.403) | 0.046 |
| | NfL (pg/mL) | 0.074 (-0.073 to 0.222) | 0.0016 | 0.073 (-0.101 to 0.217) | 0.0017 |
| | Aβ42/Aβ40 | -0.032 (-0.177 to 0.107) | 0.0011 | -0.026 (-0.168 to 0.125) | 0.0015 |
| **ALZpath Quanterix** | p-tau217 (pg/ml) | 0.239 (0.092 to 0.371) | <0.0001 | 0.269 (0.117 to 0.415) | 0.0022 |

**Table S39. Correlations between individual plasma biomarker analytes and late tau PET in the cognitively impaired sub-cohort.** The unadjusted Spearman correlation with the late tau PET measure, and the partial Spearman correlation adjusting for age, sex, and *APOE* genotype, are shown with 95% confidence intervals. Correlations between the top performing analyte and other analytes were compared by bootstrapping.

| | | Unadjusted for covariates | | Adjusted for covariates | |
|---|---|---|---|---|---|
| **Company** | **Analyte** | **Spearman rho** | **p=** | **Partial Spearman rho** | **p=** |
| **C2N PrecivityAD2** | p-tau217 (pg/ml) | 0.663 (0.466 to 0.793) | REFERENCE | 0.684 (0.510 to 0.805) | REFERENCE |
| | %p-tau217 (%) | 0.645 (0.445 to 0.775) | 0.53 | 0.665 (0.478 to 0.790) | 0.57 |
| | Aβ42/Aβ40 | -0.304 (-0.515 to -0.079) | 0.0033 | -0.328 (-0.534 to -0.084) | 0.0065 |
| **Fujirebio Lumipulse** | p-tau217 (pg/ml) | 0.645 (0.445 to 0.790) | 0.55 | 0.667 (0.476 to 0.797) | 0.57 |
| | Aβ42/Aβ40 | -0.298 (-0.497 to -0.055) | 0.0027 | -0.309 (-0.541 to -0.081) | 0.0028 |
| **Janssen LucentAD Quanterix** | p-tau217 (pg/ml) | 0.612 (0.421 to 0.746) | 0.26 | 0.637 (0.454 to 0.767) | 0.33 |
| **Roche NeuroToolKit** | p-tau181 (pg/ml) | 0.590 (0.406 to 0.728) | 0.23 | 0.613 (0.418 to 0.746) | 0.25 |
| | GFAP (ng/ml) | 0.377 (0.172 to 0.565) | 0.012 | 0.448 (0.242 to 0.618) | 0.019 |
| | NfL (pg/mL) | 0.169 (-0.063 to 0.372) | <0.0001 | 0.213 (-0.044 to 0.430) | <0.0001 |
| | Aβ42/Aβ40 | -0.093 (-0.355 to 0.161) | <0.0001 | -0.091 (-0.335 to 0.155) | <0.0001 |
| **ALZpath Quanterix** | p-tau217 (pg/ml) | 0.538 (0.330 to 0.692) | 0.030 | 0.567 (0.358 to 0.721) | 0.042 |
| **Quanterix Neurology 4-Plex** | p-tau181 (pg/ml) | 0.525 (0.309 to 0.682) | 0.18 | 0.582 (0.375 to 0.730) | 0.33 |
| | GFAP (pg/ml) | 0.463 (0.262 to 0.641) | 0.058 | 0.490 (0.261 to 0.650) | 0.064 |
| | NfL (pg/mL) | 0.283 (0.029 to 0.508) | 0.0011 | 0.312 (0.049 to 0.521) | 0.0028 |
| | Aβ42/Aβ40 | -0.200 (-0.423 to 0.057) | 0.0007 | -0.187 (-0.429 to 0.100) | 0.0028 |

**Table S40. Correlations between individual plasma biomarker analytes and late tau PET in the sub-cohort with amyloid PET >20 Centiloids.** The unadjusted Spearman correlation with the late tau PET measure, and the partial Spearman correlation adjusting for age, sex, and *APOE* genotype, are shown with 95% confidence intervals. Correlations between the top performing analyte and other analytes were compared by bootstrapping.

| | | Unadjusted for covariates | | Adjusted for covariates | |
|---|---|---|---|---|---|
| **Company** | **Analyte** | **Spearman rho** | **p=** | **Partial Spearman rho** | **p=** |
| **C2N PrecivityAD2** | p-tau217 (pg/ml) | 0.509 (0.312 to 0.675) | REFERENCE | 0.532 (0.312 to 0.700) | REFERENCE |
| | %p-tau217 (%) | 0.486 (0.285 to 0.652) | 0.55 | 0.502 (0.262 to 0.671) | 0.43 |
| | Aβ42/Aβ40 | 0.014 (-0.200 to 0.219) | 0.0008 | 0.008 (-0.218 to 0.218) | 0.0020 |
| **Fujirebio Lumipulse** | p-tau217 (pg/ml) | 0.476 (0.277 to 0.644) | 0.44 | 0.488 (0.259 to 0.666) | 0.33 |
| | Aβ42/Aβ40 | -0.016 (-0.243 to 0.198) | 0.0008 | -0.026 (-0.238 to 0.191) | 0.0020 |
| **Janssen LucentAD Quanterix** | p-tau217 (pg/ml) | 0.402 (0.191 to 0.575) | 0.028 | 0.428 (0.194 to 0.615) | 0.060 |
| **Roche NeuroToolKit** | p-tau181 (pg/ml) | 0.373 (0.184 to 0.558) | 0.028 | 0.418 (0.190 to 0.593) | 0.060 |
| | GFAP (ng/ml) | 0.290 (0.086 to 0.461) | 0.028 | 0.365 (0.148 to 0.546) | 0.060 |
| | NfL (pg/mL) | 0.070 (-0.139 to 0.255) | 0.0011 | 0.147 (-0.091 to 0.367) | 0.0020 |
| | Aβ42/Aβ40 | 0.025 (-0.178 to 0.236) | <0.0001 | 0.066 (-0.145 to 0.266) | 0.0020 |
| **Quanterix Neurology 4-Plex** | GFAP (pg/ml) | 0.353 (0.120 to 0.555) | 0.13 | 0.391 (0.143 to 0.585) | 0.11 |
| | p-tau181 (pg/ml) | 0.292 (0.066 to 0.476) | 0.028 | 0.320 (0.091 to 0.535) | 0.032 |
| | NfL (pg/mL) | 0.166 (-0.057 to 0.371) | 0.0033 | 0.219 (-0.015 to 0.429) | 0.0044 |
| | Aβ42/Aβ40 | -0.059 (-0.276 to 0.177) | 0.0008 | -0.066 (-0.311 to 0.157) | 0.0020 |
| **ALZpath Quanterix** | p-tau217 (pg/ml) | 0.350 (0.153 to 0.540) | 0.011 | 0.401 (0.179 to 0.582) | 0.032 |

**Table S41. Classification accuracies of individual and combined plasma biomarker analytes for cortical thickness status in the full cohort.** The receiver operating characteristics area under the curve (AUC) point estimate (midpoint) and 95% confidence intervals are shown for classification of cortical thickness status by individual or combined plasma biomarker analytes. Both the unadjusted AUC and the AUC adjusted for age, sex, and *APOE* genotype are provided. AUCs were compared using DeLong's test. The Benjamin-Hochberg procedure was used to adjust for multiple comparisons with the reference analyte, either within a company or across companies. The AUC for a model with covariates only was 0.758 (0.664-0.852).

| Company | Analytes | Unadjusted for covariates | | | Adjusted for covariates | | |
|---|---|---|---|---|---|---|---|
| | | AUC | Within-company comparisons p= | Across-company comparisons p= | AUC | Within-company comparisons p= | Across-company comparisons p= |
| Janssen LucentAD Quanterix | p-tau217 | 0.849 (0.765-0.933) | REFERENCE | REFERENCE | 0.896 (0.831-0.960) | REFERENCE | REFERENCE |
| Fujirebio Lumipulse | p-tau217 | 0.841 (0.765-0.916) | REFERENCE | 0.77 | 0.875 (0.802-0.948) | REFERENCE | 0.62 |
| | p-tau217 + Aβ42/Aβ40 | 0.814 (0.730-0.897) | 0.11 | | 0.873 (0.802-0.944) | 0.60 | |
| | Aβ42/Aβ40 | 0.633 (0.529-0.737) | 0.0003 | | 0.764 (0.673-0.855) | 0.010 | |
| C2N PrecivityAD2 | p-tau217 + Aβ42/Aβ40 | 0.840 (0.765-0.914) | REFERENCE | 0.77 | 0.888 (0.828-0.949) | REFERENCE | 0.65 |
| | p-tau217 | 0.839 (0.765-0.914) | 0.90 | | 0.885 (0.822-0.948) | 0.68 | |
| | %p-tau217+ Aβ42/Aβ40 | 0.819 (0.737-0.901) | 0.17 | | 0.883 (0.824-0.942) | 0.68 | |
| | %p-tau217 | 0.819 (0.737-0.901) | 0.17 | | 0.876 (0.812-0.939) | 0.68 | |
| | Aβ42/Aβ40 | 0.591 (0.488-0.694) | <0.0001 | | 0.756 (0.660-0.852) | 0.0025 | |
| Roche NeuroToolKit | p-tau181 + Aβ42/Aβ40 + GFAP + NfL | 0.838 (0.774-0.902) | REFERENCE | 0.77 | 0.874 (0.808-0.939) | 0.39 | |
| | p-tau181 + Aβ42/Aβ40 + NfL | 0.834 (0.768-0.901) | 0.52 | | 0.880 (0.821-0.939) | REFERENCE | 0.65 |
| | p-tau181 | 0.813 (0.744-0.883) | 0.38 | | 0.858 (0.788-0.929) | 0.44 | |
| | p-tau181 + Aβ42/Aβ40 | 0.810 (0.738-0.883) | 0.33 | | 0.865 (0.801-0.930) | 0.48 | |
| | NfL | 0.743 (0.663-0.823) | 0.014 | | 0.835 (0.760-0.909) | 0.0099 | |
| | GFAP | 0.677 (0.570-0.785) | 0.002 | | 0.806 (0.721-0.891) | 0.0099 | |
| | Aβ42/Aβ40 | 0.609 (0.499-0.720) | 0.0001 | | 0.778 (0.688-0.868) | 0.0099 | |
| ALZpath Quanterix | p-tau217 | 0.833 (0.755-0.912) | | 0.71 | 0.885 (0.821-0.950) | | 0.62 |
| Quanterix Neurology 4-Plex | p-tau181 + Aβ42/Aβ40 + NfL | 0.783 (0.707-0.860) | REFERENCE | 0.71 | 0.849 (0.767-0.930) | REFERENCE | 0.62 |
| | p-tau181 + Aβ42/Aβ40 + GFAP + NfL | 0.764 (0.674-0.853) | 0.32 | | 0.845 (0.760-0.929) | 0.73 | |
| | p-tau181 | 0.752 (0.650-0.855) | 0.32 | | 0.832 (0.749-0.915) | 0.37 | |
| | p-tau181 + Aβ42/Aβ40 | 0.749 (0.660-0.839) | 0.32 | | 0.831 (0.748-0.914) | 0.37 | |
| | NfL | 0.745 (0.662-0.829) | 0.32 | | 0.837 (0.756-0.918) | 0.68 | |
| | GFAP | 0.676 (0.559-0.793) | 0.093 | | 0.807 (0.713-0.901) | 0.34 | |
| | Aβ42/Aβ40 | 0.626 (0.506-0.745) | 0.093 | | 0.772 (0.680-0.865) | 0.21 | |

**Table S42. Classification accuracies of individual plasma biomarker analytes for cortical thickness status in the full cohort.** The receiver operating characteristics area under the curve (AUC) point estimate (midpoint) and 95% confidence intervals are shown for classification of cortical thickness status by plasma biomarker analytes. The single cut-off for the plasma biomarker that best distinguished cortical thickness status based on the Youden index is shown, as well as the positive percent agreement (PPA), negative percent agreement (NPA), overall accuracy, positive predictive value (PPV), and negative predictive value (NPV) of the cut-off for cortical thickness status in the full cohort, which had a 10.2% rate of cortical thickness positivity.

| Company | Analyte | AUC | Cut-off | Brier Score | PPA | NPA | Accuracy | PPV | NPV |
|---|---|---|---|---|---|---|---|---|---|
| **Janssen LucentAD Quanterix** | p-tau217 | 0.849 (0.765-0.933) | 0.0985 (pg/ml) | 0.072 | 0.815 | 0.841 | 0.838 | 0.367 | 0.976 |
| **Fujirebio Lumipulse** | p-tau217 | 0.841 (0.765-0.916) | 0.273 (pg/ml) | 0.081 | 0.778 | 0.770 | 0.771 | 0.276 | 0.968 |
| | Aβ42/Aβ40 | 0.633 (0.529-0.737) | 0.0843 | 0.091 | 0.630 | 0.686 | 0.680 | 0.185 | 0.943 |
| **C2N PrecivityAD2** | p-tau217 | 0.839 (0.765-0.914) | 4.08 (pg/ml) | 0.079 | 0.741 | 0.820 | 0.812 | 0.317 | 0.966 |
| | %p-tau217 | 0.819 (0.737-0.901) | 6.77 (%) | 0.081 | 0.778 | 0.791 | 0.789 | 0.296 | 0.969 |
| | Aβ42/Aβ40 | 0.591 (0.488-0.694) | 0.0971 | 0.090 | 0.889 | 0.356 | 0.410 | 0.135 | 0.966 |
| **ALZpath Quanterix** | p-tau217 | 0.833 (0.755-0.912) | 0.599 (pg/ml) | 0.078 | 0.815 | 0.741 | 0.748 | 0.262 | 0.973 |
| **Roche NeuroToolKit** | p-tau181 | 0.813 (0.744-0.883) | 1.32 (pg/ml) | 0.087 | 0.815 | 0.715 | 0.726 | 0.244 | 0.972 |
| | NfL | 0.743 (0.663-0.823) | 3.56 (pg/ml) | 0.089 | 0.963 | 0.431 | 0.485 | 0.160 | 0.990 |
| | GFAP | 0.677 (0.570-0.785) | 0.179 (ng/ml) | 0.090 | 0.481 | 0.854 | 0.816 | 0.271 | 0.936 |
| | Aβ42/Aβ40 | 0.609 (0.499-0.720) | 0.120 | 0.089 | 0.667 | 0.556 | 0.568 | 0.145 | 0.937 |
| **Quanterix Neurology 4-Plex** | p-tau181 | 0.752 (0.650-0.855) | 20.8 (pg/ml) | 0.086 | 0.773 | 0.601 | 0.617 | 0.170 | 0.962 |
| | NfL | 0.745 (0.662-0.829) | 20.8 (pg/ml) | 0.085 | 0.909 | 0.519 | 0.557 | 0.167 | 0.982 |
| | GFAP | 0.676 (0.559-0.793) | 169 (pg/ml) | 0.083 | 0.727 | 0.596 | 0.609 | 0.160 | 0.954 |
| | Aβ42/Aβ40 | 0.626 (0.506-0.745) | 0.0550 | 0.086 | 0.591 | 0.688 | 0.678 | 0.167 | 0.941 |

**Table S43. Classification accuracies of individual and combined plasma biomarker analytes for cortical thickness status in the cognitively impaired sub-cohort.** The receiver operating characteristics area under the curve (AUC) point estimate (midpoint) and 95% confidence intervals are shown for classification of cortical thickness status by individual or combined plasma biomarker analytes. Both the unadjusted AUC and the AUC adjusted for age, sex, and *APOE* genotype are provided. AUCs were compared using DeLong's test. The Benjamin-Hochberg procedure was used to adjust for multiple comparisons with the reference analyte, either within a company or across companies. The AUC for a model with covariates only was 0.719 (0.610-0.828).

| Company | Analytes | Unadjusted for covariates | | | Adjusted for covariates | | |
|---|---|---|---|---|---|---|---|
| | | AUC | Within-company comparisons p= | Across-company comparisons p= | AUC | Within-company comparisons p= | Across-company comparisons p= |
| Janssen LucentAD Quanterix | p-tau217 | 0.855 (0.772-0.938) | REFERENCE | REFERENCE | 0.913 (0.854-0.972) | REFERENCE | REFERENCE |
| Fujirebio Lumipulse | p-tau217 | 0.843 (0.758-0.928) | REFERENCE | 0.61 | 0.896 (0.824-0.968) | REFERENCE | 0.37 |
| | p-tau217 + Aβ42/Aβ40 | 0.842 (0.756-0.928) | 0.72 | | 0.895 (0.824-0.967) | 0.62 | |
| | Aβ42/Aβ40 | 0.611 (0.488-0.734) | 0.0001 | | 0.737 (0.631-0.842) | 0.0008 | |
| ALZpath Quanterix | p-tau217 | 0.836 (0.749-0.922) | | 0.41 | 0.902 (0.840-0.964) | | 0.49 |
| C2N PrecivityAD2 | p-tau217 | 0.824 (0.734-0.913) | REFERENCE | 0.41 | 0.872 (0.793-0.951) | 0.26 | |
| | p-tau217 + Aβ42/Aβ40 | 0.819 (0.729-0.909) | 0.24 | | 0.880 (0.806-0.954) | REFERENCE | 0.30 |
| | %p-tau217 | 0.802 (0.711-0.894) | 0.19 | | 0.856 (0.775-0.938) | 0.064 | |
| | %p-tau217+ Aβ42/Aβ40 | 0.797 (0.705-0.890) | 0.19 | | 0.857 (0.777-0.937) | 0.064 | |
| | Aβ42/Aβ40 | 0.603 (0.485-0.721) | 0.0016 | | 0.736 (0.622-0.851) | 0.003 | |
| Roche NeuroToolKit | p-tau181 + Aβ42/Aβ40 + GFAP + NfL | 0.803 (0.719-0.886) | REFERENCE | 0.41 | 0.881 (0.810-0.953) | REFERENCE | 0.30 |
| | p-tau181 + Aβ42/Aβ40 + NfL | 0.799 (0.710-0.887) | 0.86 | | 0.879 (0.804-0.953) | 0.50 | |
| | p-tau181 + Aβ42/Aβ40 | 0.786 (0.694-0.879) | 0.78 | | 0.858 (0.782-0.934) | 0.27 | |
| | p-tau181 | 0.785 (0.695-0.876) | 0.78 | | 0.859 (0.782-0.936) | 0.27 | |
| | NfL | 0.704 (0.602-0.805) | 0.0581 | | 0.811 (0.710-0.911) | 0.014 | |
| | GFAP | 0.631 (0.506-0.757) | 0.0294 | | 0.774 (0.660-0.887) | 0.0044 | |
| | Aβ42/Aβ40 | 0.617 (0.497-0.737) | 0.0259 | | 0.756 (0.649-0.863) | 0.0044 | |
| Quanterix Neurology 4-Plex | p-tau181 | 0.776 (0.672-0.880) | REFERENCE | 0.41 | 0.826 (0.727-0.925) | 0.22 | |
| | p-tau181 + Aβ42/Aβ40 + NfL | 0.770 (0.674-0.866) | 0.87 | | 0.852 (0.753-0.951) | REFERENCE | 0.37 |
| | p-tau181 + Aβ42/Aβ40 | 0.770 (0.666-0.874) | 0.78 | | 0.825 (0.725-0.925) | 0.22 | |
| | p-tau181 + Aβ42/Aβ40+ GFAP + NfL | 0.764 (0.662-0.865) | 0.87 | | 0.844 (0.739-0.950) | 0.36 | |
| | NfL | 0.727 (0.619-0.834) | 0.78 | | 0.815 (0.703-0.928) | 0.22 | |
| | GFAP | 0.652 (0.518-0.786) | 0.23 | | 0.771 (0.643-0.898) | 0.079 | |
| | Aβ42/Aβ40 | 0.584 (0.449-0.719) | 0.15 | | 0.723 (0.599-0.846) | 0.022 | |

**Table S44. Classification accuracies of individual plasma biomarker analytes for cortical thickness status in the cognitively impaired sub-cohort.** The receiver operating characteristics area under the curve (AUC) point estimate (midpoint) and 95% confidence intervals are shown for classification of cortical thickness status by plasma biomarker Analytes. The single cut-off for the plasma biomarker that best distinguished cortical thickness status based on the Youden index is shown, as well as the positive percent agreement (PPA), negative percent agreement (NPA), overall accuracy, positive predictive value (PPV), and negative predictive value (NPV) of the cut-off for cortical thickness status in the cognitively impaired sub-cohort, which had a 17.8% rate of cortical thickness positivity.

| Company | Analyte | AUC | Cut-off | Brier Score | PPA | NPA | Accuracy | PPV | NPV |
|---|---|---|---|---|---|---|---|---|---|
| **Janssen LucentAD Quanterix** | p-tau217 | 0.855 (0.772-0.938) | 0.0985 (pg/ml) | 0.107 | 0.870 | 0.783 | 0.798 | 0.465 | 0.965 |
| **Fujirebio Lumipulse** | p-tau217 | 0.843 (0.758-0.928) | 0.296 (pg/ml) | 0.111 | 0.826 | 0.745 | 0.760 | 0.413 | 0.952 |
| | Aβ42/Aβ40 | 0.611 (0.488-0.734) | 0.0822 | 0.146 | 0.565 | 0.708 | 0.682 | 0.295 | 0.882 |
| **ALZpath Quanterix** | p-tau217 | 0.836 (0.749-0.922) | 0.784 (pg/ml) | 0.116 | 0.739 | 0.849 | 0.829 | 0.515 | 0.938 |
| **C2N PrecivityAD2** | p-tau217 | 0.824 (0.734-0.913) | 4.08 (pg/ml) | 0.123 | 0.826 | 0.764 | 0.775 | 0.432 | 0.953 |
| | %p-tau217 | 0.802 (0.711-0.894) | 6.93 (%) | 0.130 | 0.826 | 0.736 | 0.752 | 0.404 | 0.951 |
| | Aβ42/Aβ40 | 0.603 (0.485-0.721) | 0.0972 | 0.144 | 0.913 | 0.377 | 0.473 | 0.241 | 0.952 |
| **Roche NeuroToolKit** | p-tau181 | 0.785 (0.695-0.876) | 1.32 (pg/ml) | 0.131 | 0.870 | 0.642 | 0.682 | 0.345 | 0.958 |
| | NfL | 0.704 (0.602-0.805) | 3.35 (pg/ml) | 0.140 | 1.000 | 0.358 | 0.473 | 0.253 | 1.000 |
| | GFAP | 0.631 (0.506-0.757) | 0.178 (ng/ml) | 0.144 | 0.478 | 0.821 | 0.760 | 0.367 | 0.879 |
| | Aβ42/Aβ40 | 0.617 (0.497-0.737) | 0.120 | 0.141 | 0.739 | 0.519 | 0.558 | 0.250 | 0.902 |
| **Quanterix Neurology 4-Plex** | p-tau181 | 0.776 (0.672-0.880) | 20.6 (pg/ml) | 0.138 | 0.947 | 0.576 | 0.640 | 0.316 | 0.981 |
| | NfL | 0.727 (0.619-0.834) | 20.8 (pg/ml) | 0.136 | 0.895 | 0.489 | 0.559 | 0.266 | 0.957 |
| | GFAP | 0.652 (0.518-0.786) | 170 (pg/ml) | 0.136 | 0.684 | 0.609 | 0.622 | 0.265 | 0.903 |
| | Aβ42/Aβ40 | 0.584 (0.449-0.719) | 0.0599 | 0.142 | 0.842 | 0.391 | 0.468 | 0.222 | 0.923 |

**Table S45. Classification accuracies of individual and combined plasma biomarker analytes for cortical thickness status in the sub-cohort with amyloid PET >20 Centiloids.** The receiver operating characteristics area under the curve (AUC) point estimate (midpoint) and 95% confidence intervals are shown for classification of cortical thickness status by individual or combined plasma biomarker analytes. Both the unadjusted AUC and the AUC adjusted for age, sex, and *APOE* genotype are provided. AUCs were compared using DeLong's test. The Benjamin-Hochberg procedure was used to adjust for multiple comparisons with the reference analyte, either within a company or across companies. The AUC for a model with covariates only was 0.752 (0.644-0.860).

| Company | Analytes | Unadjusted for covariates | | | Adjusted for covariates | | |
|---|---|---|---|---|---|---|---|
| | | AUC | Within-company comparisons p= | Across-company comparisons p= | AUC | Within-company comparisons p= | Across-company comparisons p= |
| Janssen LucentAD Quanterix | p-tau217 | 0.822 (0.728-0.915) | REFERENCE | REFERENCE | 0.893 (0.818-0.967) | | 0.85 |
| Fujirebio Lumipulse | p-tau217 + Aβ42/Aβ40 | 0.810 (0.716-0.903) | REFERENCE | 0.61 | 0.895 (0.826-0.964) | REFERENCE | REFERENCE |
| | p-tau217 | 0.810 (0.717-0.904) | 0.34 | | 0.895 (0.826-0.964) | 1.00 | |
| | Aβ42/Aβ40 | 0.527 (0.399-0.654) | 0.0002 | | 0.749 (0.640-0.858) | 0.0021 | |
| C2N PrecivityAD2 | p-tau217 + Aβ42/Aβ40 | 0.799 (0.700-0.897) | REFERENCE | 0.48 | 0.885 (0.817-0.952) | REFERENCE | 0.73 |
| | p-tau217 | 0.795 (0.700-0.890) | 0.78 | | 0.868 (0.791-0.946) | 0.24 | |
| | %p-tau217+ Aβ42/Aβ40 | 0.778 (0.672-0.883) | 0.59 | | 0.871 (0.800-0.943) | 0.49 | |
| | %p-tau217 | 0.769 (0.666-0.872) | 0.59 | | 0.855 (0.775-0.935) | 0.24 | |
| | Aβ42/Aβ40 | 0.547 (0.424-0.669) | 0.0058 | | 0.752 (0.639-0.866) | 0.0425 | |
| ALZpath Quanterix | p-tau217 | 0.790 (0.692-0.889) | | 0.48 | 0.879 (0.811-0.947) | | 0.57 |
| Roche NeuroToolKit | p-tau181 + Aβ42/Aβ40 + GFAP + NfL | 0.739 (0.649-0.829) | REFERENCE | 0.31 | 0.817 (0.715-0.919) | REFERENCE | 0.0975 |
| | p-tau181 + Aβ42/Aβ40 + NfL | 0.732 (0.640-0.824) | 0.79 | | 0.815 (0.716-0.914) | 0.81 | |
| | p-tau181 | 0.702 (0.599-0.805) | 0.45 | | 0.811 (0.714-0.908) | 0.81 | |
| | p-tau181 + Aβ42/Aβ40 | 0.695 (0.586-0.803) | 0.45 | | 0.803 (0.704-0.903) | 0.80 | |
| | NfL | 0.668 (0.556-0.779) | 0.26 | | 0.792 (0.686-0.898) | 0.45 | |
| | GFAP | 0.580 (0.445-0.715) | 0.14 | | 0.783 (0.673-0.893) | 0.38 | |
| | Aβ42/Aβ40 | 0.492 (0.349-0.636) | 0.0167 | | 0.753 (0.642-0.864) | 0.25 | |
| Quanterix Neurology 4-Plex | p-tau181 | 0.694 (0.580-0.809) | REFERENCE | 0.31 | 0.798 (0.683-0.913) | 0.81 | |
| | p-tau181 + Aβ42/Aβ40 | 0.681 (0.563-0.800) | 0.72 | | 0.798 (0.686-0.911) | 0.81 | |
| | p-tau181 + Aβ42/Aβ40 + NfL | 0.680 (0.557-0.803) | 0.72 | | 0.812 (0.699-0.925) | 1.00 | |
| | p-tau181 + Aβ42/Aβ40 + GFAP + NfL | 0.678 (0.547-0.810) | 0.72 | | 0.812 (0.689-0.934) | REFERENCE | 0.57 |
| | NfL | 0.655 (0.530-0.779) | 0.72 | | 0.775 (0.652-0.898) | 0.42 | |
| | GFAP | 0.586 (0.428-0.744) | 0.63 | | 0.772 (0.636-0.908) | 0.42 | |
| | Aβ42/Aβ40 | 0.522 (0.362-0.682) | 0.51 | | 0.745 (0.618-0.872) | 0.42 | |

**Table S46. Classification accuracies of individual plasma biomarker analytes for cortical thickness status in the sub-cohort with amyloid PET >20 Centiloids.** The receiver operating characteristics area under the curve (AUC) point estimate (midpoint) and 95% confidence intervals are shown for classification of cortical thickness status by plasma biomarker analytes. The single cut-off for the plasma biomarker that best distinguished cortical thickness status based on the Youden index is shown, as well as the positive percent agreement (PPA), negative percent agreement (NPA), overall accuracy, positive predictive value (PPV), and negative predictive value (NPV) of the cut-off for cortical thickness status in the sub-cohort with amyloid PET >20 Centiloids, which had a 18.0% rate of cortical thickness positivity.

| Company | Analyte | AUC | Cut-off | Brier Score | PPA | NPA | Accuracy | PPV | NPV |
|---|---|---|---|---|---|---|---|---|---|
| **Janssen LucentAD Quanterix** | p-tau217 | 0.822 (0.728-0.915) | 0.0985 (pg/ml) | 0.117 | 0.909 | 0.640 | 0.689 | 0.357 | 0.970 |
| **Fujirebio Lumipulse** | p-tau217 | 0.810 (0.717-0.904) | 0.321 (pg/ml) | 0.124 | 0.818 | 0.620 | 0.656 | 0.321 | 0.939 |
| | Aβ42/Aβ40 | 0.527 (0.399-0.654) | 0.0843 | 0.148 | 0.682 | 0.480 | 0.516 | 0.224 | 0.873 |
| **C2N PrecivityAD2** | p-tau217 | 0.795 ( 0.700-0.890) | 4.29 (pg/ml) | 0.131 | 0.864 | 0.640 | 0.680 | 0.345 | 0.955 |
| | %p-tau217 | 0.769 (0.666-0.872) | 7.84 (%) | 0.135 | 0.818 | 0.630 | 0.664 | 0.327 | 0.940 |
| | Aβ42/Aβ40 | 0.547 (0.424-0.669) | 0.0846 | 0.148 | 0.136 | 0.630 | 0.541 | 0.075 | 0.768 |
| **ALZpath Quanterix** | p-tau217 | 0.790 (0.692-0.889) | 0.836 (pg/ml) | 0.128 | 0.773 | 0.740 | 0.746 | 0.395 | 0.937 |
| **Roche NeuroToolKit** | p-tau181 | 0.702 (0.599-0.805) | 1.32 (pg/ml) | 0.144 | 0.909 | 0.480 | 0.557 | 0.278 | 0.960 |
| | NfL | 0.668 (0.556-0.779) | 4.94 (pg/ml) | 0.145 | 0.636 | 0.690 | 0.680 | 0.311 | 0.896 |
| | GFAP | 0.580 (0.445-0.715) | 0.179 (ng/ml) | 0.147 | 0.545 | 0.710 | 0.680 | 0.293 | 0.877 |
| | Aβ42/Aβ40 | 0.492 (0.349-0.636) | 0.111 | 0.147 | 0.318 | 0.600 | 0.549 | 0.149 | 0.800 |
| **Quanterix Neurology 4-Plex** | p-tau181 | 0.694 (0.580-0.809) | 20.8 (pg/ml) | 0.143 | 0.882 | 0.418 | 0.500 | 0.246 | 0.943 |
| | NfL | 0.655 (0.530-0.779) | 30.7 (pg/ml) | 0.145 | 0.529 | 0.772 | 0.729 | 0.333 | 0.884 |
| | GFAP | 0.586 (0.428-0.744) | 274 (pg/ml) | 0.143 | 0.353 | 0.785 | 0.708 | 0.261 | 0.849 |
| | Aβ42/Aβ40 | 0.522 (0.362-0.682) | 0.0550 | 0.145 | 0.647 | 0.519 | 0.542 | 0.224 | 0.872 |

**Table S47. Correlations between individual plasma biomarker analytes and cortical thickness in the full cohort.** The unadjusted Spearman correlation with the cortical thickness measure, and the partial Spearman correlation adjusting for age, sex, and *APOE* genotype, are shown with 95% confidence intervals. Correlations between the top performing analyte and other analytes were compared by bootstrapping.

| | | Unadjusted for covariates | | Adjusted for covariates | |
|---|---|---|---|---|---|
| **Company** | **Analyte** | **Spearman rho** | **p=** | **Partial Spearman rho** | **p=** |
| **Janssen LucentAD Quanterix** | p-tau217 (pg/ml) | -0.327 (-0.436 to -0.217) | REFERENCE | -0.320 (-0.434 to -0.198) | REFERENCE |
| **Roche NeuroToolKit** | p-tau181 (pg/ml) | -0.275 (-0.386 to -0.165) | 0.10 | -0.276 (-0.385 to -0.154) | 0.17 |
| | NfL (pg/mL) | -0.198 (-0.314 to -0.085) | 0.041 | -0.225 (-0.349 to -0.097) | 0.15 |
| | GFAP (ng/ml) | -0.171 (-0.287 to -0.049) | 0.016 | -0.197 (-0.312 to -0.074) | 0.070 |
| | Aβ42/Aβ40 | 0.050 (-0.076 to 0.170) | <0.0001 | 0.022 (-0.095 to 0.146) | <0.0001 |
| **C2N PrecivityAD2** | p-tau217 (pg/ml) | -0.274 (-0.388 to -0.159) | 0.079 | -0.268 (-0.388 to -0.134) | 0.12 |
| | %p-tau217 (%) | -0.266 (-0.378 to -0.149) | 0.10 | -0.254 (-0.373 to -0.125) | 0.12 |
| | Aβ42/Aβ40 | 0.072 (-0.054 to 0.187) | <0.0001 | 0.048 (-0.069 to 0.158) | 0.0007 |
| **ALZpath Quanterix** | p-tau217 (pg/ml) | -0.273 (-0.383 to -0.166) | 0.016 | -0.263 (-0.380 to -0.134) | 0.025 |
| **Fujirebio Lumipulse** | p-tau217 (pg/ml) | -0.256 (-0.369 to -0.138) | 0.018 | -0.242 (-0.357 to -0.117) | 0.027 |
| | Aβ42/Aβ40 | 0.060 (-0.067 to 0.183) | <0.0001 | 0.038 (-0.091 to 0.155) | <0.0001 |
| **Quanterix Neurology 4-Plex** | p-tau181 (pg/ml) | -0.194 (-0.317 to -0.066) | 0.010 | -0.182 (-0.318 to -0.046) | 0.070 |
| | GFAP (pg/ml) | -0.151 (-0.271 to -0.023) | 0.010 | -0.175 (-0.301 to -0.044) | 0.15 |
| | NfL (pg/mL) | -0.114 (-0.234 to 0.000) | 0.0008 | -0.124 (-0.260 to 0.003) | 0.044 |
| | Aβ42/Aβ40 | -0.030 (-0.161 to 0.096) | 0.0025 | -0.066 (-0.198 to 0.080) | 0.058 |

**Table S48. Correlations between individual plasma biomarker analytes and cortical thickness in the cognitively impaired sub-cohort.** The unadjusted Spearman correlation with the cortical thickness measure, and the partial Spearman correlation adjusting for age, sex, and *APOE* genotype, are shown with 95% confidence intervals. Correlations between the top performing analyte and other analytes were compared by bootstrapping.

| | | Unadjusted for covariates | | Adjusted for covariates | |
|---|---|---|---|---|---|
| **Company** | **Analyte** | **Spearman rho** | **p=** | **Partial Spearman rho** | **p=** |
| **Janssen LucentAD Quanterix** | p-tau217 (pg/ml) | -0.425 (-0.568 to -0.277) | REFERENCE | -0.424 (-0.564 to -0.274) | REFERENCE |
| **Fujirebio Lumipulse** | p-tau217 (pg/ml) | -0.384 (-0.520 to -0.222) | 0.23 | -0.386 (-0.521 to -0.233) | 0.26 |
| | Aβ42/Aβ40 | 0.078 (-0.097 to 0.248) | 0.0006 | 0.060 (-0.132 to 0.246) | 0.0017 |
| **C2N PrecivityAD2** | p-tau217 (pg/ml) | -0.356 (-0.497 to -0.196) | 0.079 | -0.365 (-0.504 to -0.185) | 0.13 |
| | %p-tau217 (%) | -0.356 (-0.497 to -0.197) | 0.15 | -0.364 (-0.517 to -0.187) | 0.19 |
| | Aβ42/Aβ40 | 0.105 (-0.067 to 0.269) | 0.0020 | 0.074 (-0.102 to 0.242) | 0.0017 |
| **ALZpath Quanterix** | p-tau217 (pg/ml) | -0.355 (-0.499 to -0.198) | 0.025 | -0.352 (-0.494 to -0.190) | 0.030 |
| **Roche NeuroToolKit** | p-tau181 (pg/ml) | -0.336 (-0.481 to -0.179) | 0.070 | -0.342 (-0.480 to -0.179) | 0.088 |
| | NfL (pg/mL) | -0.228 (-0.378 to -0.072) | 0.029 | -0.249 (-0.401 to -0.069) | 0.069 |
| | Aβ42/Aβ40 | 0.161 (-0.026 to 0.326) | 0.0046 | 0.157 (-0.015 to 0.330) | 0.010 |
| | GFAP (ng/ml) | -0.144 (-0.307 to 0.022) | 0.0028 | -0.184 (-0.330 to -0.011) | 0.012 |
| **Quanterix Neurology 4-Plex** | p-tau181 (pg/ml) | -0.235 (-0.413 to -0.039) | 0.0059 | -0.229 (-0.412 to -0.042) | 0.056 |
| | NfL (pg/mL) | -0.123 (-0.295 to 0.042) | 0.0006 | -0.116 (-0.295 to 0.070) | 0.017 |
| | GFAP (pg/ml) | -0.090 (-0.256 to 0.089) | <0.0001 | -0.115 (-0.292 to 0.077) | 0.017 |
| | Aβ42/Aβ40 | 0.021 (-0.160 to 0.212) | 0.0006 | -0.002 (-0.179 to 0.185) | 0.011 |

**Table S49. Correlations between individual plasma biomarker analytes and cortical thickness in the sub-cohort with amyloid PET >20 Centiloid.** The unadjusted Spearman correlation with the cortical thickness measure, and the partial Spearman correlation adjusting for age, sex, and *APOE* genotype, are shown with 95% confidence intervals. Correlations between the top performing analyte and other analytes were compared by bootstrapping.

| | | Unadjusted for covariates | | Adjusted for covariates | |
|---|---|---|---|---|---|
| **Company** | **Analyte** | **Spearman rho** | **p=** | **Partial Spearman rho** | **p=** |
| **Janssen LucentAD Quanterix** | p-tau217 (pg/ml) | -0.527 (-0.650 to -0.386) | REFERENCE | -0.524 (-0.648 to -0.395) | REFERENCE |
| **Fujirebio Lumipulse** | p-tau217 (pg/ml) | -0.477 (-0.608 to -0.334) | 0.27 | -0.471 (-0.608 to -0.322) | 0.26 |
| | Aβ42/Aβ40 | 0.041 (-0.137 to 0.222) | <0.0001 | 0.040 (-0.148 to 0.215) | <0.0001 |
| **C2N PrecivityAD2** | p-tau217 (pg/ml) | -0.458 (-0.585 to -0.317) | 0.087 | -0.459 (-0.587 to -0.305) | 0.12 |
| | %p-tau217 (%) | -0.415 (-0.551 to -0.270) | 0.046 | -0.408 (-0.542 to -0.260) | 0.042 |
| | Aβ42/Aβ40 | -0.016 (-0.193 to 0.163) | <0.0001 | -0.024 (-0.198 to 0.162) | <0.0001 |
| **ALZpath Quanterix** | p-tau217 (pg/ml) | -0.416 (-0.554 to -0.267) | 0.0079 | -0.413 (-0.553 to -0.258) | 0.013 |
| **Roche NeuroToolKit** | p-tau181 (pg/ml) | -0.389 (-0.523 to -0.237) | 0.015 | -0.401 (-0.536 to -0.258) | 0.028 |
| | NfL (pg/mL) | -0.277 (-0.431 to -0.111) | 0.0083 | -0.317 (-0.467 to -0.143) | 0.028 |
| | GFAP (ng/ml) | -0.153 (-0.319 to 0.025) | <0.0001 | -0.181 (-0.335 to -0.024) | <0.0001 |
| | Aβ42/Aβ40 | -0.038 (-0.231 to 0.134) | <0.0001 | -0.069 (-0.243 to 0.104) | 0.0007 |
| **Quanterix Neurology 4-Plex** | p-tau181 (pg/ml) | -0.327 (-0.496 to -0.139) | 0.0079 | -0.306 (-0.480 to -0.130) | 0.013 |
| | GFAP (pg/ml) | -0.190 (-0.361 to 0.001) | 0.0003 | -0.193 (-0.373 to 0.009) | 0.0054 |
| | NfL (pg/mL) | -0.171 (-0.347 to 0.012) | 0.0003 | -0.163 (-0.358 to 0.031) | 0.0021 |
| | Aβ42/Aβ40 | 0.007 (-0.209 to 0.199) | <0.0001 | -0.026 (-0.254 to 0.183) | <0.0001 |

**Table S50. Classification accuracies of individual and combined plasma biomarker analytes for cognitive status in the full cohort.** The receiver operating characteristics area under the curve (AUC) point estimate (midpoint) and 95% confidence intervals are shown for classification of cognitive status by individual or combined plasma biomarker analytes. Both the unadjusted AUC and the AUC adjusted for age, sex, and *APOE* genotype are provided. AUCs were compared using DeLong's test. The Benjamin-Hochberg procedure was used to adjust for multiple comparisons with the reference analyte, either within a company or across companies. The AUC for a model with covariates only was 0.559 (0.502-0.616).

| Company | Analytes | Unadjusted for covariates | | | Adjusted for covariates | | |
|---|---|---|---|---|---|---|---|
| | | AUC | Within-company comparisons p= | Across-company comparisons p= | AUC | Within-company comparisons p= | Across-company comparisons p= |
| C2N PrecivityAD2 | p-tau217 + Aβ42/Aβ40 | 0.680 (0.627-0.733) | REFERENCE | REFERENCE | 0.691 (0.639-0.743) | REFERENCE | REFERENCE |
| | %p-tau217 | 0.678 (0.625-0.731) | 0.85 | | 0.688 (0.636-0.740) | 0.80 | |
| | p-tau217 | 0.677 (0.625-0.730) | 0.84 | | 0.690 (0.638-0.742) | 0.80 | |
| | %p-tau217+ Aβ42/Aβ40 | 0.675 (0.622-0.728) | 0.84 | | 0.687 (0.635-0.739) | 0.80 | |
| | Aβ42/Aβ40 | 0.558 (0.501-0.615) | 0.0004 | | 0.572 (0.515-0.628) | <0.0001 | |
| Janssen LucentAD Quanterix | p-tau217 | 0.670 (0.615-0.724) | | 0.46 | 0.672 (0.618-0.725) | REFERENCE | 0.28 |
| Fujirebio Lumipulse | p-tau217 | 0.667 (0.613-0.721) | REFERENCE | 0.40 | 0.668 (0.614-0.721) | 0.99 | |
| | p-tau217 + Aβ42/Aβ40 | 0.660 (0.606-0.715) | 0.65 | | 0.668 (0.614-0.721) | REFERENCE | 0.28 |
| | Aβ42/Aβ40 | 0.591 (0.534-0.647) | 0.023 | | 0.601 (0.545-0.656) | 0.0008 | |
| ALZpath Quanterix | p-tau217 | 0.664 (0.610-0.718) | | 0.40 | 0.667 (0.614-0.721) | | 0.28 |
| Roche NeuroToolKit | p-tau181 + Aβ42/Aβ40 + NfL | 0.648 (0.593-0.703) | REFERENCE | 0.40 | 0.665 (0.612-0.719) | 0.38 | |
| | p-tau181 + Aβ42/Aβ40 + GFAP + NfL | 0.643 (0.587-0.698) | 0.48 | | 0.675 (0.622-0.728) | REFERENCE | 0.51 |
| | p-tau181 | 0.639 (0.584-0.694) | 0.48 | | 0.642 (0.587-0.696) | 0.11 | |
| | p-tau181 + Aβ42/Aβ40 | 0.638 (0.583-0.694) | 0.38 | | 0.642 (0.587-0.696) | 0.11 | |
| | NfL | 0.607 (0.551-0.663) | 0.14 | | 0.650 (0.596-0.705) | 0.17 | |
| | GFAP | 0.606 (0.550-0.662) | 0.14 | | 0.654 (0.599-0.708) | 0.17 | |
| | Aβ42/Aβ40 | 0.572 (0.516-0.629) | 0.049 | | 0.577 (0.521-0.634) | 0.0024 | |
| Quanterix Neurology 4-Plex | p-tau181 + Aβ42/Aβ40+ GFAP + NfL | 0.635 (0.576-0.695) | REFERENCE | 0.40 | 0.674 (0.618-0.731) | REFERENCE | 0.67 |
| | p-tau181 + Aβ42/Aβ40 + NfL | 0.629 (0.569-0.688) | 0.60 | | 0.662 (0.605-0.720) | 0.4 | |
| | p-tau181 + Aβ42/Aβ40 | 0.618 (0.559-0.678) | 0.30 | | 0.631 (0.572-0.690) | 0.065 | |
| | p-tau181 | 0.593 (0.533-0.653) | 0.12 | | 0.619 (0.560-0.679) | 0.051 | |
| | GFAP | 0.590 (0.530-0.650) | 0.10 | | 0.650 (0.592-0.709) | 0.15 | |
| | NfL | 0.583 (0.523-0.643) | 0.11 | | 0.639 (0.581-0.698) | 0.14 | |
| | Aβ42/Aβ40 | 0.582 (0.522-0.643) | 0.10 | | 0.601 (0.541-0.660) | 0.031 | |

**Table S51. Classification accuracies of individual plasma biomarker analytes for cognitive status in the full cohort.** The receiver operating characteristics area under the curve (AUC) point estimate (midpoint) and 95% confidence intervals are shown for classification of cognitive status by plasma biomarker analytes. The single cut-off for the plasma biomarker that best distinguished amyloid PET status based on the Youden index is shown, as well as the positive percent agreement (PPA), negative percent agreement (NPA), overall accuracy, positive predictive value (PPV), and negative predictive value (NPV) of the cut-off for cognitive impairment status in the full cohort, which had a 49.0% rate of cognitive impairment.

| Company | Analyte | AUC | Cut-off | Brier Score | PPA | NPA | Accuracy | PPV | NPV |
|---|---|---|---|---|---|---|---|---|---|
| C2N PrecivityAD2 | %p-tau217 | 0.678 (0.625-0.731) | 7.63 (%) | 0.222 | 0.385 | 0.895 | 0.645 | 0.779 | 0.603 |
| | p-tau217 | 0.677 (0.625-0.730) | 3.42 (pg/ml) | 0.223 | 0.479 | 0.800 | 0.643 | 0.697 | 0.615 |
| | Aβ42/Aβ40 | 0.558 (0.501-0.615) | 0.0929 | 0.247 | 0.594 | 0.520 | 0.556 | 0.543 | 0.571 |
| Janssen LucentAD Quanterix | p-tau217 | 0.670 (0.615-0.724) | 0.0695 (pg/ml) | 0.225 | 0.573 | 0.715 | 0.645 | 0.659 | 0.636 |
| Fujirebio Lumipulse | p-tau217 | 0.667 (0.613-0.721) | 0.273 (pg/ml) | 0.230 | 0.469 | 0.815 | 0.645 | 0.709 | 0.615 |
| | Aβ42/Aβ40 | 0.591 (0.534-0.647) | 0.0867 | 0.244 | 0.552 | 0.600 | 0.577 | 0.570 | 0.583 |
| ALZpath Quanterix | p-tau217 | 0.664 (0.610-0.718) | 0.423 (pg/ml) | 0.229 | 0.661 | 0.630 | 0.645 | 0.632 | 0.660 |
| Roche NeuroToolKit | p-tau181 | 0.639 (0.584-0.694) | 1.23 (pg/ml) | 0.238 | 0.594 | 0.690 | 0.643 | 0.648 | 0.639 |
| | NfL | 0.607 (0.551-0.663) | 4.66 (pg/ml) | 0.243 | 0.469 | 0.730 | 0.602 | 0.625 | 0.589 |
| | GFAP | 0.606 (0.550-0.662) | 0.148 (ng/ml) | 0.240 | 0.417 | 0.775 | 0.599 | 0.640 | 0.581 |
| | Aβ42/Aβ40 | 0.572 (0.516-0.629) | 0.127 | 0.247 | 0.734 | 0.455 | 0.592 | 0.564 | 0.641 |
| Quanterix Neurology 4-Plex | p-tau181 | 0.593 (0.533-0.653) | 25.5 (pg/ml) | 0.244 | 0.450 | 0.725 | 0.588 | 0.621 | 0.569 |
| | GFAP | 0.590 (0.530-0.650) | 207 (pg/ml) | 0.243 | 0.462 | 0.719 | 0.591 | 0.622 | 0.572 |
| | NfL | 0.583 (0.523-0.643) | 18.8 (pg/ml) | 0.245 | 0.737 | 0.421 | 0.579 | 0.560 | 0.615 |
| | Aβ42/Aβ40 | 0.582 (0.522-0.643) | 0.0609 | 0.245 | 0.719 | 0.450 | 0.585 | 0.567 | 0.616 |

**Table S52. Classification accuracies of individual and combined plasma biomarker analytes for cognitive status in the sub-cohort with amyloid PET >20 Centiloids.** The receiver operating characteristics area under the curve (AUC) point estimate (midpoint) and 95% confidence intervals are shown for classification of cognitive status by individual or combined plasma biomarker analytes. Both the unadjusted AUC and the AUC adjusted for age, sex, and *APOE* genotype are provided. AUCs were compared using DeLong's test. The Benjamin-Hochberg procedure was used to adjust for multiple comparisons with the reference analyte, either within a company or across companies. The AUC for a model with covariates only was 0.532 (0.448-0.615).

| Company | Analytes | Unadjusted for covariates | | | Adjusted for covariates | | |
|---|---|---|---|---|---|---|---|
| | | AUC | Within-company comparisons p= | Across-company comparisons p= | AUC | Within-company comparisons p= | Across-company comparisons p= |
| C2N PrecivityAD2 | %p-tau217+ Aβ42/Aβ40 | 0.720 (0.649-0.792) | REFERENCE | REFERENCE | 0.722 (0.650-0.793) | 0.96 | |
| | p-tau217 + Aβ42/Aβ40 | 0.718 (0.645-0.792) | 0.91 | | 0.723 (0.650-0.795) | REFERENCE | REFERENCE |
| | %p-tau217 | 0.717 (0.645-0.789) | 0.91 | | 0.718 (0.647-0.790) | 0.96 | |
| | p-tau217 | 0.716 (0.642-0.789) | 0.91 | | 0.721 (0.649-0.794) | 0.48 | |
| | Aβ42/Aβ40 | 0.500 (0.414-0.586) | 0.001 | | 0.519 (0.434-0.604) | 0.0003 | |
| Janssen LucentAD Quanterix | p-tau217 | 0.720 (0.646-0.795) | | 1.00 | 0.712 (0.639-0.786) | | 0.80 |
| Fujirebio Lumipulse | p-tau217 + Aβ42/Aβ40 | 0.710 (0.635-0.786) | REFERENCE | 0.95 | 0.709 (0.635-0.784) | REFERENCE | 0.80 |
| | p-tau217 | 0.708 (0.632-0.783) | 0.82 | | 0.708 (0.634-0.783) | 0.94 | |
| | Aβ42/Aβ40 | 0.555 (0.470-0.640) | 0.0032 | | 0.563 (0.478-0.649) | 0.0054 | |
| ALZpath Quanterix | p-tau217 | 0.692 (0.614-0.770) | | 0.66 | 0.688 (0.612-0.764) | | 0.51 |
| Roche NeuroToolKit | p-tau181 + Aβ42/Aβ40 + NfL | 0.681 (0.599-0.763) | REFERENCE | 0.66 | 0.683 (0.605-0.761) | 0.5 | |
| | p-tau181 + Aβ42/Aβ40 + GFAP + NfL | 0.664 (0.583-0.744) | 0.44 | | 0.698 (0.622-0.774) | REFERENCE | 0.80 |
| | p-tau181 + Aβ42/Aβ40 | 0.664 (0.583-0.746) | 0.38 | | 0.657 (0.578-0.736) | 0.29 | |
| | p-tau181 | 0.662 (0.580-0.743) | 0.38 | | 0.658 (0.579-0.737) | 0.29 | |
| | NfL | 0.643 (0.561-0.725) | 0.37 | | 0.666 (0.587-0.745) | 0.33 | |
| | GFAP | 0.636 (0.556-0.716) | 0.38 | | 0.676 (0.599-0.752) | 0.29 | |
| | Aβ42/Aβ40 | 0.528 (0.442-0.613) | 0.057 | | 0.530 (0.446-0.614) | 0.0051 | |
| Quanterix Neurology 4-Plex | p-tau181 + Aβ42/Aβ40 + GFAP + NfL | 0.651 (0.561-0.741) | REFERENCE | 0.66 | 0.716 (0.635-0.797) | REFERENCE | 0.90 |
| | p-tau181 + Aβ42/Aβ40 + NfL | 0.650 (0.558-0.742) | 0.96 | | 0.694 (0.609-0.778) | 0.39 | |
| | p-tau181 + Aβ42/Aβ40 | 0.631 (0.538-0.723) | 0.61 | | 0.661 (0.574-0.747) | 0.29 | |
| | GFAP | 0.626 (0.537-0.715) | 0.61 | | 0.699 (0.617-0.781) | 0.39 | |
| | p-tau181 | 0.625 (0.532-0.718) | 0.61 | | 0.662 (0.575-0.749) | 0.33 | |
| | NfL | 0.617 (0.525-0.710) | 0.61 | | 0.678 (0.593-0.764) | 0.39 | |
| | Aβ42/Aβ40 | 0.577 (0.485-0.670) | 0.61 | | 0.617 (0.527-0.706) | 0.12 | |

**Table S53. Classification accuracies of individual plasma biomarker analytes for cognitive status in the sub-cohort with amyloid PET >20 Centiloids.** The receiver operating characteristics area under the curve (AUC) point estimate (midpoint) and 95% confidence intervals are shown for classification of cognitive status by plasma biomarker analytes. The single cut-off for the plasma biomarker that best distinguished cognitive status based on the Youden index is shown, as well as the positive percent agreement (PPA), negative percent agreement (NPA), overall accuracy, positive predictive value (PPV), and negative predictive value (NPV) of the cut-off for cognitive impairment status in the sub-cohort with amyloid PET >20 Centiloids, which had a 60.2% rate of cognitive impairment.

| Company | Analyte | AUC | Cut-off | Brier Score | PPA | NPA | Accuracy | PPV | NPV |
|---|---|---|---|---|---|---|---|---|---|
| **Janssen LucentAD Quanterix** | p-tau217 | 0.720 (0.646-0.795) | 0.106 (pg/ml) | 0.210 | 0.591 | 0.776 | 0.665 | 0.800 | 0.557 |
| **C2N PrecivityAD2** | %p-tau217 | 0.717 (0.645-0.789) | 7.81 (%) | 0.206 | 0.635 | 0.750 | 0.681 | 0.793 | 0.576 |
| | p-tau217 | 0.716 (0.642-0.789) | 4.67 (pg/ml) | 0.208 | 0.565 | 0.763 | 0.644 | 0.783 | 0.537 |
| | Aβ42/Aβ40 | 0.500 (0.414-0.586) | 0.0839 | 0.239 | 0.296 | 0.592 | 0.414 | 0.523 | 0.357 |
| **Fujirebio Lumipulse** | p-tau217 | 0.708 (0.632-0.783) | 0.298 (pg/ml) | 0.214 | 0.696 | 0.632 | 0.670 | 0.741 | 0.578 |
| | Aβ42/Aβ40 | 0.555 (0.470-0.640) | 0.0820 | 0.238 | 0.557 | 0.579 | 0.565 | 0.667 | 0.463 |
| **ALZpath Quanterix** | p-tau217 | 0.692 (0.614-0.770) | 0.653 (pg/ml) | 0.217 | 0.730 | 0.618 | 0.686 | 0.743 | 0.603 |
| **Roche NeuroToolKit** | p-tau181 | 0.662 (0.580-0.743) | 1.17 (pg/ml) | 0.228 | 0.852 | 0.461 | 0.696 | 0.705 | 0.673 |
| | NfL | 0.643 (0.561-0.725) | 4.44 (pg/ml) | 0.232 | 0.643 | 0.632 | 0.639 | 0.725 | 0.539 |
| | GFAP | 0.636 (0.556-0.716) | 0.116 (ng/ml) | 0.227 | 0.783 | 0.421 | 0.639 | 0.672 | 0.561 |
| | Aβ42/Aβ40 | 0.528 (0.442-0.613) | 0.121 | 0.239 | 0.748 | 0.355 | 0.592 | 0.637 | 0.482 |
| **Quanterix Neurology 4-Plex** | GFAP | 0.626 (0.537-0.715) | 193 (pg/ml) | 0.226 | 0.714 | 0.525 | 0.642 | 0.707 | 0.533 |
| | p-tau181 | 0.625 (0.532-0.718) | 18.9 (pg/ml) | 0.231 | 0.857 | 0.393 | 0.679 | 0.694 | 0.632 |
| | NfL | 0.617 (0.525-0.710) | 20.4 (pg/ml) | 0.231 | 0.806 | 0.426 | 0.660 | 0.693 | 0.578 |
| | Aβ42/Aβ40 | 0.577 (0.485-0.670) | 0.0608 | 0.233 | 0.878 | 0.295 | 0.654 | 0.667 | 0.600 |

**Table S54. Correlations between individual plasma biomarker analytes and dementia symptoms.** The unadjusted Spearman correlation with the Clinical Dementia Rating Sum of Boxes, and the partial Spearman correlation adjusting for age, sex, and *APOE* genotype, are shown with 95% confidence intervals. Correlations between the top performing analyte and other analytes were compared by bootstrapping.

| | | Unadjusted for covariates | | Adjusted for covariates | |
|---|---|---|---|---|---|
| **Company** | **Analyte** | **Spearman rho** | **p=** | **Partial Spearman rho** | **p=** |
| **C2N PrecivityAD2** | p-tau217 (pg/ml) | 0.414 (0.325 to 0.505) | REFERENCE | 0.432 (0.331 to 0.517) | REFERENCE |
| | %p-tau217 (%) | 0.407 (0.313 to 0.494) | 0.65 | 0.415 (0.321 to 0.497) | 0.31 |
| | Aβ42/Aβ40 | -0.152 (-0.243 to -0.057) | <0.0001 | -0.145 (-0.238 to -0.040) | <0.0001 |
| **Fujirebio Lumipulse** | p-tau217 (pg/ml) | 0.403 (0.311 to 0.490) | 0.65 | 0.411 (0.312 to 0.492) | 0.36 |
| | Aβ42/Aβ40 | -0.213 (-0.307 to -0.121) | <0.0001 | -0.205 (-0.301 to -0.109) | <0.0001 |
| **Janssen LucentAD Quanterix** | p-tau217 (pg/ml) | 0.391 (0.295 to 0.482) | 0.34 | 0.402 (0.300 to 0.487) | 0.23 |
| **ALZpath Quanterix** | p-tau217 (pg/ml) | 0.374 (0.280 to 0.461) | 0.074 | 0.385 (0.281 to 0.470) | 0.042 |
| **Roche NeuroToolKit** | p-tau181 (pg/ml) | 0.331 (0.237 to 0.415) | 0.0038 | 0.352 (0.250 to 0.434) | 0.0081 |
| | NfL (pg/mL) | 0.261 (0.164 to 0.351) | 0.0022 | 0.315 (0.213 to 0.404) | 0.029 |
| | GFAP (ng/ml) | 0.257 (0.159 to 0.342) | <0.0001 | 0.306 (0.203 to 0.396) | 0.0048 |
| | Aβ42/Aβ40 | -0.155 (-0.251 to -0.064) | <0.0001 | -0.142 (-0.232 to -0.050) | <0.0001 |
| **Quanterix Neurology 4-Plex** | GFAP (pg/ml) | 0.240 (0.132 to 0.337) | <0.0001 | 0.291 (0.187 to 0.387) | 0.0033 |
| | p-tau181 (pg/ml) | 0.234 (0.131 to 0.332) | <0.0001 | 0.240 (0.140 to 0.339) | <0.0001 |
| | NfL (pg/mL) | 0.224 (0.118 to 0.321) | 0.0006 | 0.278 (0.175 to 0.373) | 0.0063 |
| | Aβ42/Aβ40 | -0.198 (-0.301 to -0.095) | <0.0001 | -0.188 (-0.284 to -0.082) | <0.0001 |

**Table S55. Correlations between individual plasma biomarker analytes and dementia symptoms in the sub-cohort with amyloid PET >20 Centiloids.** The unadjusted Spearman correlation with the Clinical Dementia Rating Sum of Boxes, and the partial Spearman correlation adjusting for age, sex, and *APOE* genotype, are shown with 95% confidence intervals. Correlations between the top performing analyte and other analytes were compared by bootstrapping.

| | | Unadjusted for covariates | | Adjusted for covariates | |
|---|---|---|---|---|---|
| **Company** | **Analyte** | **Spearman rho** | **p=** | **Partial Spearman rho** | **p=** |
| **Fujirebio Lumipulse** | p-tau217 (pg/ml) | 0.460 (0.337 to 0.570) | REFERENCE | 0.465 (0.339 to 0.571) | 0.89 |
| | Aβ42/Aβ40 | -0.099 (-0.242 to 0.049) | <0.0001 | -0.099 (-0.245 to 0.039) | <0.0001 |
| **C2N PrecivityAD2** | p-tau217 (pg/ml) | 0.457 (0.331 to 0.568) | 0.95 | 0.471 (0.356 to 0.575) | REFERENCE |
| | %p-tau217 (%) | 0.456 (0.339 to 0.569) | 0.95 | 0.460 (0.340 to 0.561) | 0.83 |
| | Aβ42/Aβ40 | 0.000 (-0.134 to 0.146) | <0.0001 | -0.001 (-0.151 to 0.142) | <0.0001 |
| **Janssen LucentAD Quanterix** | p-tau217 (pg/ml) | 0.448 (0.325 to 0.561) | 0.89 | 0.454 (0.317 to 0.562) | 0.75 |
| **ALZpath Quanterix** | p-tau217 (pg/ml) | 0.390 (0.252 to 0.508) | 0.10 | 0.400 (0.269 to 0.515) | 0.076 |
| **Roche NeuroToolKit** | p-tau181 (pg/ml) | 0.342 (0.220 to 0.458) | 0.019 | 0.362 (0.229 to 0.475) | 0.026 |
| | NfL (pg/mL) | 0.299 (0.171 to 0.429) | 0.031 | 0.355 (0.228 to 0.475) | 0.11 |
| | GFAP (ng/ml) | 0.269 (0.132 to 0.384) | 0.0068 | 0.325 (0.195 to 0.442) | 0.039 |
| | Aβ42/Aβ40 | 0.010 (-0.126 to 0.151) | <0.0001 | 0.023 (-0.125 to 0.166) | <0.0001 |
| **Quanterix Neurology 4-Plex** | GFAP (pg/ml) | 0.267 (0.121 to 0.403) | 0.0098 | 0.327 (0.176 to 0.453) | 0.040 |
| | p-tau181 (pg/ml) | 0.264 (0.110 to 0.407) | 0.0025 | 0.263 (0.109 to 0.408) | 0.0020 |
| | NfL (pg/mL) | 0.250 (0.095 to 0.392) | 0.0088 | 0.302 (0.163 to 0.438) | 0.034 |
| | Aβ42/Aβ40 | -0.157 (-0.321 to -0.012) | 0.0022 | -0.157 (-0.306 to -0.006) | 0.0011 |

### Appendix A. Derivation of cut-offs for tau PET and cortical thickness abnormality.

Utilizing the entire ADNI dataset, without limiting data to the plasma study subset, a mixture model was applied using the expectation-maximization algorithm within the R statistical environment. This model was independently fitted to three distinct Analytes: the mesial temporal meta-ROI flortaucipir SUVR (Figure A1), the temporal-parietal meta-ROI flortaucipir SUVR (Figure A2), and the meta-ROI cortical thickness measure (Figure A3). The following approach was applied to derive the cut-off thresholds: for both the mesial temporal meta-ROI flortaucipir SUVR and the temporal-parietal meta-ROI flortaucipir SUVR, thresholds were established at a value equal to the mean plus two standard deviations of the first component, which corresponds to the normal distribution within the mixture model. Conversely, the cut-off threshold for the meta-ROI cortical thickness was positioned at the mean minus two standard deviations of the second component, again aligning with the normal distribution curve represented within the model. The estimated cut-off thresholds derived from these calculations are reported in Table A1.

**Table A1. Cut-off thresholds for ADNI dataset, including mesial temporal and temporal-parietal meta-ROI flortaucipir SUVRs, and meta-ROI cortical thickness.** Thresholds are derived from mixture model analysis, indicating mean plus or minus two standard deviations for the corresponding normal distribution components.

| Outcome Analyte | Cut-off |
|---|---|
| mesial temporal meta-ROI flortaucipir SUVR | 1.328 SUVR |
| temporal-parietal meta-ROI flortaucipir SUVR | 1.224 SUVR |
| cortical thickness within the meta-ROI | 2.572 mm |

**Figure A1: Distribution of mesial temporal meta-ROI flortaucipir SUVR values across the ADNI dataset, with the mixture model fit displayed.**

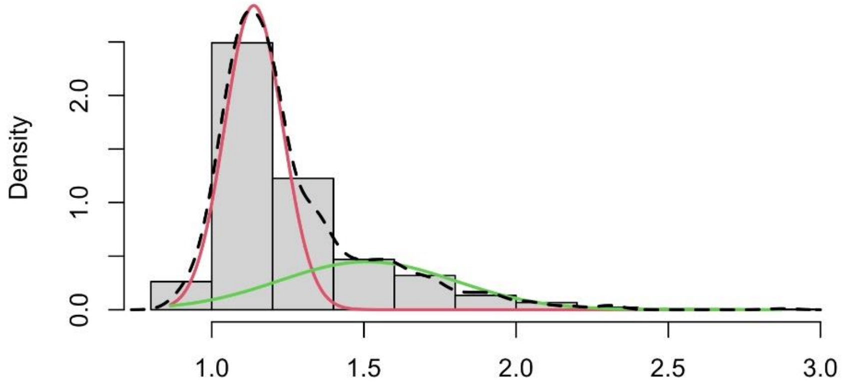

**Figure A2: Distribution of temporal-parietal meta-ROI flortaucipir SUVR values, with the mixture model overlay.**

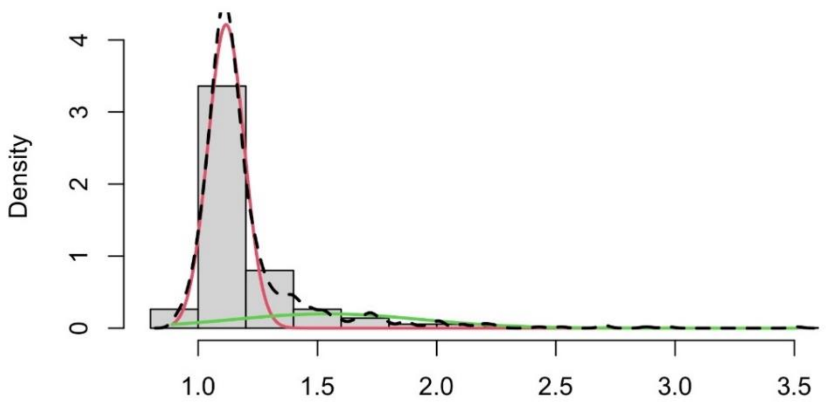

**Figure A3: Distribution of meta-ROI cortical thickness measurements with mixture model fit.**

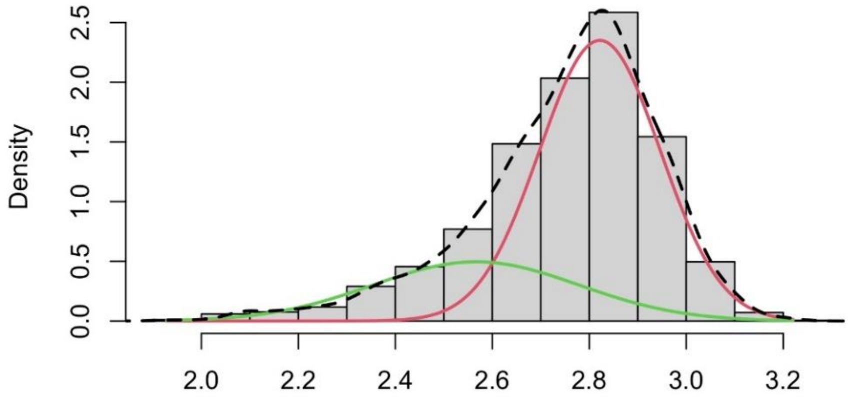

### Appendix B. Datasets and code used for analyses.

### Table B1. Demographic, cognitive, CSF, and imaging measures.

| | ADNI variable | LONI source dataset | Type of variable | Time from plasma collection |
|---|---|---|---|---|
| **Demographics** | | | | |
| Age at plasma collection (years) | AGE | PTDEMOG_25Mar2024 | continuous | 0 |
| Gender | PTGENDER | PTDEMOG_25Mar2024 | binary | NA |
| APOE ε4 carrier status | APOE4 | APOERES_02Apr2024 | binary | NA |
| APOE genotype | | APOERES_02Apr2024 | categorical | NA |
| Race | PTRACCAT | PTDEMOG_25Mar2024 | categorical | NA |
| Ethnicity | PTETHCAT | PTDEMOG_25Mar2024 | categorical | NA |
| Years of education | PTEDUCAT | PTDEMOG_25Mar2024 | continuous | NA |
| **Clinical dementia symptoms** | | | | |
| Diagnosis | DX | DXSUM_PDXCONV_02Apr2024 | categorical | <1 year |
| CDR_10 | | CDR_25Mar2024 | binary | <1 year |
| CDR | CDGLOBAL | CDR_25Mar2024 | categorical | <1 year |
| CDR-SB | CDRSB | CDR_25Mar2024 | continuous | <1 year |
| MMSE | MMSE | MMSE_02Apr2024 | continuous | <1 year |
| ADAS13 | ADAS13 | ADAS_ADNI1_02Apr2024 | continuous | <1 year |
| Date of clinical assessment | EXAMDATE | CDR_25Mar2024 | time | <1 year |
| Time from CDR to plasma collection | | CDR_25Mar2024 | continuous | NA |
| **CSF measures** | | | | |
| CSF_10 (based on pTau/Ab42) | | UPENNBIOMK_ROCHE_ELECSYS_02Apr2024 | binary | <1 year |
| Elecsys Aβ42 (pg/ml) | ABETA | UPENNBIOMK_ROCHE_ELECSYS_02Apr2024 | continuous | <1 year |
| Elecsys total tau (pg/ml) | TAU | UPENNBIOMK_ROCHE_ELECSYS_02Apr2024 | continuous | <1 year |
| Elecsys p-tau181 (pg/ml) | PTAU | UPENNBIOMK_ROCHE_ELECSYS_02Apr2024 | continuous | <1 year |
| Date of CSF collection | | UPENNBIOMK_ROCHE_ELECSYS_02Apr2024 | time | <1 year |
| Time from CSF to plasma collection | | UPENNBIOMK_ROCHE_ELECSYS_02Apr2024 | continuous | NA |
| **Amyloid PET** | | | | |
| CENT20_10 | | UCBERKELEY_AMY_6MM_25Mar2024 | binary | <0.5 year |
| CENT37_10 | | UCBERKELEY_AMY_6MM_25Mar2024 | binary | <0.5 year |
| Amyloid PET Centiloid | CENTILOIDS | UCBERKELEY_AMY_6MM_25Mar2024 | continuous | <0.5 year |
| Date of amyloid PET | | UCBERKELEY_AMY_6MM_25Mar2024 | time | <0.5 year |
| Time from amyloid PET to plasma collection | | UCBERKELEY_AMY_6MM_25Mar2024 | continuous | NA |
| **Tau PET** | | | | |
| Tau_early_10 | | | binary | <1 year |
| Tau_early | MesialTemporal | ADNI Tau ROIs | continuous | <1 year |
| Tau_late_10 | | | binary | <1 year |
| Tau_late | TemporoParietal | ADNI Tau ROIs | continuous | <1 year |
| Date of tau PET | | | time | <1 year |
| Time from tau PET to plasma collection | | | continuous | NA |
| **Cortical thickness** | | | | |
| Atrophy_10 | | ADNI metaROI thickness harmoized | binary | <1 year |
| Cortical signature volume | metaROI.AgeAdj | ADNI metaROI thickness harmoized | continuous | <1 year |
| Date of MRI | EXAMDATE | ADNI metaROI thickness harmoized | time | <1 year |
| Time from brain atrophy to plasma collection | | ADNI metaROI thickness harmoized | continuous | NA |

**Table B2. Plasma biomarkers.**

| |
|---|
| C2N_plasma_APS2 |
| C2N_plasma_Abeta42_Abeta40 |
| C2N_plasma_Abeta42 |
| C2N_plasma_Abeta40 |
| C2N_plasma_ptau217 |
| C2N_plasma_nptau217 |
| C2N_plasma_ptau217_ratio |
| Roche_plasma_Abeta42_Abeta40 |
| Roche_plasma_Ab42 |
| Roche_plasma_Ab40 |
| Roche_plasma_ptau181 |
| Roche_plasma_GFAP |
| Roche_plasma_NfL |
| Fuji_plasma_Ab42 |
| Fuji_plasma_Ab40 |
| Fuji_plasma_ptau217 |
| AlzPath_plasma_ptau217 |
| Janssen_plasma_ptau217 |
| QX_plasma_Ab42_Ab40 |
| QX_plasma_Ab42 |
| QX_plasma_Ab40 |
| QX_plasma_ptau181 |
| QX_plasma_GFAP |
| QX_plasma_NfL |

**Code B1. R script for combining plasma into a single dataset.**

```
# FNIH Project 2
# combine all plasma data
#### Coded: Kellen Petersen
#### Date as of commenting and documentation - 05/28/2024

# Merge unblinded data
library(tidyverse)
library(lubridate)
library(here)

#here::i_am("make_plasma_data_CLEAN.R")

# Load files
c2n <-
read.csv(here("adni_data","Study_2_unblinded_datasets","C2N_precivityresults_formatted_unblind_simp
le.csv"))
roche<- read.csv(here("adni_data","Study_2_unblinded_datasets","U_Gothenburg_Elecsys
results_formatted_unblind_simple.csv"))
fuji <- read.csv(here("adni_data","Study_2_unblinded_datasets","Indiana_U_Lumipulse
Results_formatted_unblind_data_simple.csv"))
alzpath<-
read.csv(here("adni_data","Study_2_unblinded_datasets","Quanterix_ALZpath_pTau217_results_formatted
_unblind_simple.csv"))
janssen<-
read.csv(here("adni_data","Study_2_unblinded_datasets","Quanterix_Janssen_pTau217_results_formatted
_unblind_simple.csv"))
quanterix<-
read.csv(here("adni_data","Study_2_unblinded_datasets","Quanterix_Simoa_pTau181_results__formatted_
unblind_simple.csv"))
quanterix2<-
read.csv(here("adni_data","Study_2_unblinded_datasets","Quanterix_Simoa_N4PE_results_formatted_unbl
ind_simple.csv"))

############################################################################
# Make datasets "wide"
# Calculate AB ratio variables as needed

c2n_wide <- c2n %>%
  select(RID,EXAMDATE,VISCODE2,TESTNAME,TESTVALUE) %>%
  pivot_wider(names_from = "TESTNAME", values_from = "TESTVALUE")
names(c2n_wide) <- c("RID","EXAMDATE","VISCODE2",
                     "C2N_plasma_Abeta40", "C2N_plasma_Abeta42",
                     "C2N_plasma_Abeta42_Abeta40",
                     "C2N_plasma_ptau217","C2N_plasma_nptau217",
                     "C2N_plasma_ptau217_ratio")

roche_wide <- roche %>%
  select(RID,EXAMDATE,VISCODE2,TESTNAME,TESTVALUE) %>%
  pivot_wider(names_from = "TESTNAME", values_from = "TESTVALUE")
names(roche_wide) <- c("RID","EXAMDATE","VISCODE2",
                       "Roche_plasma_Ab40","Roche_plasma_Ab42",
                       "Roche_plasma_GFAP","Roche_plasma_NfL",
                       "Roche_plasma_ptau181")
roche_wide$Roche_plasma_Ab40 <- as.numeric(roche_wide$Roche_plasma_Ab40)
roche_wide$Roche_plasma_Ab42 <- as.numeric(roche_wide$Roche_plasma_Ab42)
roche_wide$Roche_plasma_GFAP <- as.numeric(roche_wide$Roche_plasma_GFAP)
roche_wide$Roche_plasma_NfL <- as.numeric(roche_wide$Roche_plasma_NfL)
roche_wide$Roche_plasma_ptau181 <- as.numeric(roche_wide$Roche_plasma_ptau181)
roche_wide <- roche_wide %>%
  mutate(Roche_plasma_Ab42_Ab40 = Roche_plasma_Ab42/Roche_plasma_Ab40 * 0.001)

fuji_wide <- fuji %>%
  select(RID,EXAMDATE,VISCODE2,TESTNAME,TESTVALUE) %>%
  pivot_wider(names_from = "TESTNAME", values_from = "TESTVALUE")
```

```
names(fuji_wide) <- c("RID","EXAMDATE","VISCODE2",
                      "Fuji_plasma_ptau217",
                      "Fuji_plasma_Ab40","Fuji_plasma_Ab42")
fuji_wide <- fuji_wide %>%
  mutate(Fuji_plasma_Ab42_Ab40 = Fuji_plasma_Ab42/Fuji_plasma_Ab40)

alzpath_wide <- alzpath %>%
  select(RID,EXAMDATE,VISCODE2,TESTNAME,TESTVALUE) %>%
  pivot_wider(names_from = "TESTNAME", values_from = "TESTVALUE")
names(alzpath_wide) <- c("RID","EXAMDATE","VISCODE2",
                         "AlzPath_plasma_ptau217")
alzpath_wide$AlzPath_plasma_ptau217 <- as.numeric(alzpath_wide$AlzPath_plasma_ptau217)

janssen_wide <- janssen %>%
  select(RID,EXAMDATE,VISCODE2,TESTNAME,TESTVALUE) %>%
  pivot_wider(names_from = "TESTNAME", values_from = "TESTVALUE")
names(janssen_wide) <- c("RID","EXAMDATE","VISCODE2",
                         "Janssen_plasma_ptau217")

quanterix_wide <- quanterix %>%
  select(RID,EXAMDATE,VISCODE2,TESTNAME,TESTVALUE) %>%
  pivot_wider(names_from = "TESTNAME", values_from = "TESTVALUE")
names(quanterix_wide) <- c("RID","EXAMDATE","VISCODE2",
                           "QX_plasma_ptau181")
quanterix_wide$QX_plasma_ptau181 <- as.numeric(quanterix_wide$QX_plasma_ptau181)

quanterix2_wide <- quanterix2 %>%
  select(RID,EXAMDATE,VISCODE2,TESTNAME,TESTVALUE) %>%
  pivot_wider(names_from = "TESTNAME", values_from = "TESTVALUE")
names(quanterix2_wide) <- c("RID","EXAMDATE","VISCODE2",
                            "QX_plasma_Ab40","QX_plasma_Ab42",
                            "QX_plasma_GFAP","QX_plasma_NfL")
quanterix2_wide$QX_plasma_Ab40 <- as.numeric(quanterix2_wide$QX_plasma_Ab40)
quanterix2_wide$QX_plasma_Ab42 <- as.numeric(quanterix2_wide$QX_plasma_Ab42) # 1378 missing Ab42
quanterix2_wide$QX_plasma_GFAP <- as.numeric(quanterix2_wide$QX_plasma_GFAP)
quanterix2_wide$QX_plasma_NfL <- as.numeric(quanterix2_wide$QX_plasma_NfL)
quanterix2_wide$RID <- as.factor(quanterix2_wide$RID)
quanterix2_wide <- quanterix2_wide %>%
  mutate(QX_plasma_Ab42_Ab40 = QX_plasma_Ab42 / QX_plasma_Ab40)

# Combine Quanterix datasets
QX <- merge(quanterix_wide, quanterix2_wide, by=c("RID","EXAMDATE"), all=TRUE)
QX <- QX %>% arrange(RID,EXAMDATE)

# Remove VISCODEs
c2n_wide_b <- c2n_wide %>% select(-VISCODE2)
fuji_wide_b <- fuji_wide %>% select(-VISCODE2)
alzpath_wide_b <- alzpath_wide %>% select(-VISCODE2)
janssen_wide_b <- janssen_wide %>% select(-VISCODE2)
roche_wide_b <- roche_wide %>% select(-VISCODE2)
QX_b <- QX

# Merge
df  <- merge(c2n_wide_b, fuji_wide_b, by=c("RID","EXAMDATE"), all = TRUE)
df2 <- merge(df, alzpath_wide_b, by=c("RID","EXAMDATE"), all = TRUE)
df3 <- merge(df2, janssen_wide_b, by=c("RID","EXAMDATE"), all = TRUE)
df4 <- merge(df3, roche_wide_b, by=c("RID","EXAMDATE"), all = TRUE)
df5 <- merge(df4, QX_b, by=c("RID","EXAMDATE"), all = TRUE)

# Add RID_EXAMDATE and arrange
df5$EXAMDATE <- as.Date(df5$EXAMDATE, format = "%m/%d/%y")
df6 <- df5 %>% arrange(RID,EXAMDATE) %>%
  mutate(RID_EXAMDATE = paste(RID,EXAMDATE,sep = "_")) %>%
```

```
  relocate(RID_EXAMDATE, .after = 2)

# Write to csv
write.csv(df6, here("adni_data","Study_2_unblinded_datasets","plasma_master_CLEAN.csv"),
row.names=FALSE)
```

**Code B2. R script for merging plasma dataset with other variables.**

```
# FNIH Project 2
# make longitudinal dataset
#### Coded: Kellen Petersen
#### Date as of commenting and documentation - 05/28/2024

# load libraries
library(tidyverse)
library(here)

# here::i_am("make_master_dataset_CLEAN.R")

# Parameters
year <- 365.25
cut_years <- 1
cut_centiloids <- 20
cut_centiloids_sens <- 37
cut_tau_mesialtemporal <- 1.32826246868779 # mixEM
cut_tau_temporoparietal <- 1.26937256044181 # mixEM
cut_atrophy <- 2.57231252744212 # mixEM

################################################################################
# DEMOGRAPHICS
demo0 <- read.csv(here("adni_data", "PTDEMOG_25Mar2024.csv"))
demo <- demo0 %>%
  mutate(AGE = difftime(as.Date(VISDATE,"%m/%d/%Y"),as.Date(PTDOB,"%m/%d/%Y"), units = "days")) %>%
  mutate(VISDATE_DEMO = VISDATE) %>%
  select(RID,VISDATE_DEMO,PTDOB,AGE,PTGENDER,PTEDUCAT,PTRACCAT,PTETHCAT)
demo$AGE <- round(as.numeric(demo$AGE)/year,1)

demo <- demo %>%
  mutate(RACE = ifelse(PTRACCAT == 4, 0,
                       ifelse(PTRACCAT == 5, 1,
                              ifelse(PTRACCAT == -4, -4, 2 ))))
demo$VISDATE_DEMO <- format(as.Date(demo$VISDATE_DEMO, "%m/%d/%Y"), "%Y-%m-%d")
demo$PTDOB <- format(as.Date(demo$PTDOB, "%m/%d/%Y"), "%Y-%m-%d")

################################################################################
# APOE4: 3 different variables
apo0 <- read.csv(here("adni_data", "APOERES_02Apr2024.csv"))
apo0$APOE4_count <- rowSums(apo0[, c("APGEN1","APGEN2")] == 4)
apo0 <- apo0 %>% mutate(APOE4_positive = ifelse(APOE4_count > 0, 1, 0))
apo0$APOE_genotype <- paste(apo0$APGEN1, apo0$APGEN2, sep = "")
#apoe <- subset(apo, VISCODE == 'sc', c("RID", "APOE4_count", "APOE4_positive", "APOE_genotype"))
#apoe2 <- apoe[!duplicated(apoe$RID, fromLast = TRUE),]
apo <- apo0 %>%
  select(RID,APOE4_count, APOE4_positive, APOE_genotype)

################################################################################
# DIAGNOSIS
dx0 <- read.csv(here("adni_data", "DXSUM_PDXCONV_02Apr2024.csv"))
dx <- dx0 %>%
  mutate(EXAMDATE_DIAGNOSIS = EXAMDATE) %>%
  mutate(DX_123 = DIAGNOSIS) %>%
  select(RID, EXAMDATE_DIAGNOSIS, DX_123)
dx$EXAMDATE_DIAGNOSIS <- as.Date(dx$EXAMDATE_DIAGNOSIS, format = "%Y-%m-%d")

################################################################################
# CDR
cdr0 <- read.csv(here("adni_data", "CDR_25Mar2024.csv"))
cdr <- cdr0 %>%
  mutate(VISDATE_CDR = VISDATE) %>%
  mutate(CDR = CDGLOBAL) %>%
```

```
  mutate(CDR_10 = ifelse(CDR == -1, -1,
                       ifelse(CDR == 0, 0, 1))) %>%
  mutate(CDR_SOB = CDMEMORY + CDORIENT + CDJUDGE + CDCOMMUN + CDHOME + CDCARE) %>%
  select(RID, VISDATE_CDR, CDR, CDR_SOB, CDR_10)
cdr$VISDATE_CDR <- as.Date(cdr$VISDATE_CDR, format = "%Y-%m-%d")

################################################################################
# MMSE
mmse0 <- read.csv(here("adni_data", "MMSE_02Apr2024.csv"))

mmse0$VISCODE2 <- gsub("f", "bl", mmse0$VISCODE2)
mmse0$VISCODE2 <- gsub("sc", "bl", mmse0$VISCODE2)
mmse <- mmse0 %>%
  mutate(VISDATE_MMSE = VISDATE) %>%
  select(RID, VISCODE2, VISDATE_MMSE, MMSCORE) %>%
  arrange(RID, VISDATE_MMSE)
mmse$VISDATE_MMSE <- as.Date(mmse$VISDATE_MMSE, format = "%Y-%m-%d")

################################################################################
# CSF
# No PTID in the CSF file
csf0 <- read.csv(here("adni_data", "UPENNBIOMK_ROCHE_ELECSYS_02Apr2024.csv"))
csf <- csf0 %>%
  mutate(EXAMDATE_CSF = EXAMDATE) %>%
  mutate(RUNDATE_CSF = RUNDATE) %>%
  mutate(ABETA42_over_40  = ABETA42/ABETA40) %>%
  mutate(PTAU_over_ABETA42 = PTAU/ABETA42) %>%
  select(RID, EXAMDATE_CSF, ABETA42, PTAU, TAU, ABETA42_over_40, PTAU_over_ABETA42)
csf$EXAMDATE_CSF <- as.Date(csf$EXAMDATE_CSF, format = "%Y-%m-%d")

################################################################################
# AMYLOID PET
amy0 <- read.csv(here("adni_data", "UCBERKELEY_AMY_6MM_25Mar2024.csv"))
amy <- amy0 %>%
  mutate(SCANDATE_AMY = SCANDATE) %>%
  mutate(PROCESSDATE_AMY = PROCESSDATE) %>%
  select(RID, SCANDATE_AMY,      CENTILOIDS) %>%
  mutate(CENTILOIDS_10 = ifelse(CENTILOIDS > cut_centiloids, 1, 0)) %>%
  mutate(CENTILOIDS_10_sens = ifelse(CENTILOIDS > cut_centiloids_sens, 1, 0))
amy$SCANDATE_AMY <- as.Date(amy$SCANDATE_AMY, format = "%Y-%m-%d")

################################################################################
# TAU PET
tau0 <- read.csv(here("adni_data", "ADNI_Tau_ROIs.csv"))
tau <- tau0 %>%
  mutate(SCANDATE_TAU = SCANDATE) %>%
  select(RID,SCANDATE_TAU,MesialTemporal,TemporoParietal) %>%
  mutate(TAU_MesialTemporal_10 = ifelse(MesialTemporal > cut_tau_mesialtemporal, 1, 0)) %>%
  mutate(TAU_TemporoParietal_10 = ifelse(TemporoParietal > cut_tau_temporoparietal, 1, 0))
tau$SCANDATE_TAU <- as.Date(tau$SCANDATE_TAU, format = "%Y-%m-%d")

################################################################################
# ATROPHY
atrophy0 <- read.csv(here("adni_data", "ADNI_metaROI_thickness_harmonized_.csv"))
atrophy <- atrophy0 %>%
  mutate(SCANDATE_ATROPHY = EXAMDATE) %>%
  mutate(atrophy = metaROI.AgeAdj) %>%
  select(RID,SCANDATE_ATROPHY,atrophy) %>%
  mutate(atrophy_10 = ifelse(atrophy < cut_atrophy, 1, 0))
atrophy$SCANDATE_ATROPHY <- as.Date(atrophy$SCANDATE_ATROPHY, format = "%Y-%m-%d")
```

```
###############################################################################
# REMOVE 0 dataframes
rm(list = c("demo0", "apo0","dx0", "cdr0", "mmse0", "csf0", "amy0", "tau0", "atrophy0"))

###############################################################################
# SORT
demo <- demo %>% arrange(RID,VISDATE_DEMO)
apo <- apo %>% arrange(RID)
cdr <- cdr %>% arrange(RID, VISDATE_CDR)
dx <- dx %>% arrange(RID, EXAMDATE_DIAGNOSIS)
mmse <- mmse %>% arrange(RID, VISDATE_MMSE)
csf <- csf %>% arrange(RID, EXAMDATE_CSF)
amy <- amy %>% arrange(RID, SCANDATE_AMY)
tau <- tau %>% arrange(RID, SCANDATE_TAU)
atrophy <- atrophy %>% arrange(RID, SCANDATE_ATROPHY)

###############################################################################
# Load MASTER PLASMA file
plasma <- read.csv(here("adni_data", "Study_2_unblinded_datasets", "plasma_master_CLEAN.csv"))
plasma$EXAMDATE <- as.Date(plasma$EXAMDATE, format = "%Y-%m-%d")
IDs_plasma <- unique(plasma$RID)

###############################################################################
# MERGE
m_1 <- merge(plasma, cdr, by = c("RID"), all = TRUE) %>%
  mutate(difftime_years = as.numeric(abs(difftime(EXAMDATE, VISDATE_CDR, units = "days")/year)))
%>%
  group_by(RID_EXAMDATE) %>%
  slice(which.min(difftime_years)) %>%
  mutate(VISDATE_CDR = ifelse(all(difftime_years > cut_years), NA, VISDATE_CDR)) %>%
  mutate(CDR = ifelse(all(difftime_years > cut_years), NA, CDR)) %>%
  mutate(CDR_SOB = ifelse(all(difftime_years > cut_years), NA, CDR_SOB)) %>%
  mutate(CDR_10 = ifelse(all(difftime_years > cut_years), NA, CDR_10)) %>%
  mutate(difftime_years = ifelse(all(difftime_years > cut_years), NA, difftime_years)) %>%
  arrange(RID, EXAMDATE) %>%
  select(-difftime_years)
m_1$VISDATE_CDR <- as.Date(m_1$VISDATE_CDR, format = "%Y-%m-%d")

m_2 <- merge(m_1, dx, by = c("RID"), all = TRUE) %>%
  mutate(difftime_years = as.numeric(abs(difftime(EXAMDATE, EXAMDATE_DIAGNOSIS, units =
"days")/year))) %>%
  group_by(RID_EXAMDATE) %>%
  slice(which.min(difftime_years)) %>%
  mutate(EXAMDATE_DIAGNOSIS = ifelse(all(difftime_years > cut_years), NA, EXAMDATE_DIAGNOSIS)) %>%
  mutate(DX_123 = ifelse(all(difftime_years > cut_years), NA, DX_123))%>%
  mutate(difftime_years = ifelse(all(difftime_years > cut_years), NA, difftime_years)) %>%
  arrange(RID, EXAMDATE) %>%
  select(-difftime_years)
m_2$EXAMDATE_DIAGNOSIS <- as.Date(m_2$EXAMDATE_DIAGNOSIS, format = "%Y-%m-%d")

m_4 <- merge(m_2, mmse, by = c("RID"), all = TRUE) %>%
  mutate(difftime_years = as.numeric(abs(difftime(EXAMDATE, VISDATE_MMSE, units = "days")/year)))
%>%
  group_by(RID_EXAMDATE) %>%
  slice(which.min(difftime_years)) %>%
  mutate(VISDATE_MMSE = ifelse(all(difftime_years > cut_years), NA, VISDATE_MMSE)) %>%
  mutate(MMSCORE = ifelse(all(difftime_years > cut_years), NA, MMSCORE))%>%
  mutate(difftime_years = ifelse(all(difftime_years > cut_years), NA, difftime_years)) %>%
  arrange(RID, EXAMDATE) %>%
  select(-difftime_years)
m_4$VISDATE_MMSE <- as.Date(m_4$VISDATE_MMSE, format = "%Y-%m-%d")
```

```
m_5 <- merge(m_4, csf, by = c("RID"), all = TRUE) %>%
  mutate(difftime_years = as.numeric(abs(difftime(EXAMDATE, EXAMDATE_CSF, units = "days")/year)))
%>%
  mutate(difftime_years = ifelse(is.na(difftime_years),1000,difftime_years)) %>%
  group_by(RID_EXAMDATE) %>%
  slice(which.min(difftime_years)) %>%
  mutate(EXAMDATE_CSF = ifelse(all(difftime_years > cut_years), NA, EXAMDATE_CSF)) %>%
  mutate(PTAU_over_ABETA42 = ifelse(all(difftime_years > cut_years), NA, PTAU_over_ABETA42))%>%
  mutate(difftime_years = ifelse(all(difftime_years > cut_years), NA, difftime_years)) %>%
  arrange(RID, EXAMDATE) %>%
  select(-difftime_years) %>%
  filter(RID %in% IDs_plasma)
m_5$EXAMDATE_CSF <- as.Date(m_5$EXAMDATE_CSF, format = "%Y-%m-%d")

m_6 <- merge(m_5, amy, by = c("RID"), all = TRUE) %>%
  mutate(difftime_years = as.numeric(abs(difftime(EXAMDATE, SCANDATE_AMY, units = "days")/year)))
%>%
  mutate(difftime_years = ifelse(is.na(difftime_years),1000,difftime_years)) %>%
  group_by(RID_EXAMDATE) %>%
  slice(which.min(difftime_years)) %>%
  mutate(SCANDATE_AMY = ifelse(all(difftime_years > cut_years), NA, SCANDATE_AMY)) %>%
  mutate(CENTILOIDS = ifelse(all(difftime_years > cut_years), NA, CENTILOIDS))%>%
  mutate(CENTILOIDS_10 = ifelse(all(difftime_years > cut_years), NA, CENTILOIDS_10))%>%
  mutate(CENTILOIDS_10_sens = ifelse(all(difftime_years > cut_years), NA, CENTILOIDS_10_sens))%>%
  mutate(difftime_years = ifelse(all(difftime_years > cut_years), NA, difftime_years)) %>%
  arrange(RID, EXAMDATE) %>%
  select(-difftime_years) %>%
  filter(RID %in% IDs_plasma)
m_6$SCANDATE_AMY <- as.Date(m_6$SCANDATE_AMY, format = "%Y-%m-%d")

m_7 <- merge(m_6, tau, by = c("RID"), all = TRUE) %>%
  mutate(difftime_years = as.numeric(abs(difftime(EXAMDATE, SCANDATE_TAU, units = "days")/year)))
%>%
  mutate(difftime_years = ifelse(is.na(difftime_years),1000,difftime_years)) %>%
  group_by(RID_EXAMDATE) %>%
  slice(which.min(difftime_years)) %>%
  mutate(SCANDATE_TAU = ifelse(all(difftime_years > cut_years), NA, SCANDATE_TAU)) %>%
  mutate(MesialTemporal = ifelse(all(difftime_years > cut_years), NA, MesialTemporal))%>%
  mutate(TemporoParietal = ifelse(all(difftime_years > cut_years), NA, TemporoParietal))%>%
  mutate(TAU_MesialTemporal_10 = ifelse(all(difftime_years > cut_years), NA,
TAU_MesialTemporal_10))%>%
  mutate(TAU_TemporoParietal_10 = ifelse(all(difftime_years > cut_years), NA,
TAU_TemporoParietal_10))%>%
  mutate(difftime_years = ifelse(all(difftime_years > cut_years), NA, difftime_years)) %>%
  arrange(RID, EXAMDATE) %>%
  select(-difftime_years)%>%
  filter(RID %in% IDs_plasma)
m_7$SCANDATE_TAU <- as.Date(m_7$SCANDATE_TAU, format = "%Y-%m-%d")

m_8 <- merge(m_7, atrophy, by = c("RID"), all = TRUE) %>%
  mutate(difftime_years = as.numeric(abs(difftime(EXAMDATE, SCANDATE_ATROPHY, units =
"days")/year))) %>%
  mutate(difftime_years = ifelse(is.na(difftime_years),1000,difftime_years)) %>%
  group_by(RID_EXAMDATE) %>%
  slice(which.min(difftime_years)) %>%
  mutate(SCANDATE_ATROPHY = ifelse(all(difftime_years > cut_years), NA, SCANDATE_ATROPHY)) %>%
  mutate(atrophy = ifelse(all(difftime_years > cut_years), NA, atrophy))%>%
  mutate(atrophy_10 = ifelse(all(difftime_years > cut_years), NA, atrophy_10))%>%
  mutate(difftime_years = ifelse(all(difftime_years > cut_years), NA, difftime_years)) %>%
  arrange(RID, EXAMDATE) %>%
  select(-difftime_years)%>%
  filter(RID %in% IDs_plasma)
m_8$SCANDATE_ATROPHY <- as.Date(m_8$SCANDATE_ATROPHY, format = "%Y-%m-%d")
```

```
# ADD demo and apo at end
demo_noage <- demo %>% select(-AGE) %>%
  group_by(RID) %>%
  slice_head(n = 1)
demo_noage_apo <- merge(demo_noage, apo, by = c("RID"), all = TRUE) %>%
  arrange(RID, VISDATE_DEMO) %>%
  select(-VISDATE_DEMO)

m_9 <- merge(m_8, demo_noage_apo, by = c("RID"), all=FALSE) %>%
  arrange(RID, EXAMDATE)
m_9$PTDOB <- as.Date(m_9$PTDOB, format = "%Y-%m-%d")
m_9 <- m_9 %>%
  mutate(AGE = as.numeric(abs(difftime(EXAMDATE, PTDOB, units = "days")/year))) %>%
  mutate(AGE = round(AGE,1)) %>%
  arrange(RID, EXAMDATE)

# Write to csv
write.csv(m_9, here("adni_data","merged_data_CLEAN.csv"), row.names = FALSE)
```

**Code B3. R script for creating cross-sectional dataset used in analysis.**

```
# FNIH Project 2
# make cross-sectional dataset
#### Coded: Kellen Petersen
#### Date as of commenting and documentation - 05/28/2024

# Load libraries
library(tidyverse)
library(here)

#here::i_am("make_cross_sectional_dataset_CLEAN.R")

out_project <- here() #Folder where you want all output written to, a results and plot folder will
be written here by default
out_results <- paste0(out_project,"/results/")
out_plots <- paste0(out_project,"/plots/")

data_file <- here("adni_data","merged_data_CLEAN.csv") # file name + path here
data.in00 <- read.csv(data_file)

# Change -1 in cdr file to NA
data.in000 <- data.in00 %>%
  mutate(CDR = ifelse(CDR == -1, NA, CDR)) %>%
  mutate(CDR_SOB = ifelse(CDR == 0, 0, CDR_SOB))

# GROUP
data.in0 <- data.in000

################################################################################
c2n <-
read.csv(here("adni_data","Study_2_unblinded_datasets","C2N_precivityresults_formatted_unblind_simp
le.csv"))
roche<- read.csv(here("adni_data","Study_2_unblinded_datasets","U_Gothenburg_Elecsys
results_formatted_unblind_simple.csv"))
fuji <- read.csv(here("adni_data","Study_2_unblinded_datasets","Indiana_U_Lumipulse
Results_formatted_unblind_data_simple.csv"))
alzpath<-
read.csv(here("adni_data","Study_2_unblinded_datasets","Quanterix_ALZpath_pTau217_results_formatted
_unblind_simple.csv"))
janssen<-
read.csv(here("adni_data","Study_2_unblinded_datasets","Quanterix_Janssen_pTau217_results_formatted
_unblind_simple.csv"))
quanterix<-
read.csv(here("adni_data","Study_2_unblinded_datasets","Quanterix_Simoa_pTau181_results__formatted_
unblind_simple.csv"))
quanterix2<-
read.csv(here("adni_data","Study_2_unblinded_datasets","Quanterix_Simoa_N4PE_results_formatted_unbl
ind_simple.csv"))

IDs_other5 <- unique(c2n$RID)
IDs_quant <- unique(quanterix$RID)

ID_both <- intersect(IDs_other5, IDs_quant)
ID_only_other5 <- setdiff(IDs_other5, IDs_quant)
ID_only_quant <- setdiff(IDs_quant, IDs_other5)
ID_all <- union(ID_both, union(ID_only_other5, ID_only_quant))

ID_other5 <- union(ID_only_other5, ID_both)
ID_quant <- union(ID_only_quant, ID_both)

ID_2tp <- data.in0 %>% group_by(RID) %>% summarise(n_time = n()) %>% filter(n_time == 2) %>%
pull(RID)
ID_3tp <- data.in0 %>% group_by(RID) %>% summarise(n_time = n()) %>% filter(n_time == 3) %>%
pull(RID)
ID_4tp <- data.in0 %>% group_by(RID) %>% summarise(n_time = n()) %>% filter(n_time == 4) %>%
pull(RID)
```

```
ID_5tp <- data.in0 %>% group_by(RID) %>% summarise(n_time = n()) %>% filter(n_time == 5) %>%
pull(RID)

vars_ID <- c("RID","EXAMDATE","RID_EXAMDATE")
vars_c2n <- c("C2N_plasma_Abeta40", "C2N_plasma_Abeta42",
              "C2N_plasma_Abeta42_Abeta40", "C2N_plasma_ptau217",
              "C2N_plasma_nptau217", "C2N_plasma_ptau217_ratio")
vars_fuji <- c("Fuji_plasma_ptau217", "Fuji_plasma_Ab40", "Fuji_plasma_Ab42",
               "Fuji_plasma_Ab42_Ab40")
vars_alzpath <- c("AlzPath_plasma_ptau217")
vars_janssen <- c("Janssen_plasma_ptau217")
vars_roche <- c("Roche_plasma_Ab40", "Roche_plasma_Ab42", "Roche_plasma_GFAP",
                "Roche_plasma_NfL", "Roche_plasma_ptau181", "Roche_plasma_Ab42_Ab40")
vars_quanterix <- c("QX_plasma_ptau181", "QX_plasma_Ab40", "QX_plasma_Ab42",
                    "QX_plasma_GFAP", "QX_plasma_NfL", "QX_plasma_Ab42_Ab40")
vars_plasma5 <- c(vars_c2n, vars_fuji, vars_alzpath, vars_janssen, vars_roche)
vars_plasma <- c(vars_c2n, vars_fuji, vars_alzpath, vars_janssen, vars_roche, vars_quanterix)

vars_cdr <- c("VISDATE_CDR", "CDR", "CDR_SOB", "CDR_10")
vars_dx <- c("EXAMDATE_DIAGNOSIS", "DX_123")
vars_nptests <- c("VISDATE_MMSE", "MMSCORE")
vars_CSF <- c("EXAMDATE_CSF", "PTAU_over_ABETA42")
vars_AB <- c("SCANDATE_AMY", "CENTILOIDS", "CENTILOIDS_10", "CENTILOIDS_10_sens")
vars_TAU <- c("SCANDATE_TAU", "MesialTemporal", "TemporoParietal",
              "TAU_MesialTemporal_10", "TAU_TemporoParietal_10")
vars_MRI <- c("SCANDATE_ATROPHY", "atrophy", "atrophy_10")
vars_demo <- c("PTDOB", "PTGENDER", "PTEDUCAT", "PTRACCAT", "PTETHCAT", "RACE",
               "APOE4_count", "APOE4_positive", "APOE_genotype", "AGE")

# How many for each visit?
data.in0_ind <- data.in0 %>%
  group_by(RID) %>%
  mutate(index_RID_timepoint = row_number()) %>%
  ungroup()

################################################################################
#IDs <- unique(data.in0_ind$RID)
IDs <- ID_only_other5
vars_complete <- c(vars_ID, vars_plasma5, vars_cdr, vars_AB,vars_demo)
# IDs <- unique(data.in0$RID)
tmp1 <- data.in0_ind %>%
  filter(RID %in% IDs) %>%
  select(vars_complete) %>%
  filter(complete.cases(.))
cross1 <- data.in0_ind %>%
  filter(RID_EXAMDATE %in% tmp1$RID_EXAMDATE) %>%
  group_by(RID) %>%
  filter(EXAMDATE == max(EXAMDATE))

################################################################################
# ID_both <- intersect(IDs_other5, IDs_quant)
IDs <- ID_both
vars_complete <- c(vars_ID, vars_plasma5, vars_quanterix, vars_cdr, vars_AB,vars_demo)
#IDs <- unique(data.in0$RID)
tmp2 <- data.in0_ind %>%
  filter(RID %in% IDs) %>%
  select(vars_complete) %>%
  filter(complete.cases(.))
cross2 <- data.in0_ind %>%
  filter(RID_EXAMDATE %in% tmp2$RID_EXAMDATE) %>%
  group_by(RID) %>%
  filter(EXAMDATE == max(EXAMDATE))

################################################################################
# # ID_only_quant <- setdiff(IDs_quant, IDs_other5)
# IDs <- ID_only_quant
# vars_complete <- c(vars_ID, vars_quanterix, vars_cdr, vars_AB,vars_demo)
```

```
# #IDs <- unique(data.in0$RID)
# tmp3 <- data.in0_ind %>%
#   filter(RID %in% IDs) %>%
#   select(vars_complete) %>%
#   filter(complete.cases(.))
# cross3 <- data.in0_ind %>%
#   filter(RID_EXAMDATE %in% tmp3$RID_EXAMDATE) %>%
#   group_by(RID) %>%
#   filter(EXAMDATE == max(EXAMDATE))

cross <- rbind(cross1,cross2) %>%
  arrange(RID, EXAMDATE)

II <- which(cross$RID %in% union(ID_both,ID_only_other5))
cross$other5YN <- 0
cross$other5YN[II] <- 1

II <- which(cross$RID %in% union(ID_both,ID_only_quant))
cross$quantYN <- 0
cross$quantYN[II] <- 1

cross2 <- cross %>%
  mutate(RACE = ifelse(RACE==1,1,
                       ifelse(RACE==2,3,
                              ifelse(RACE==0,2,-99))))

# Write to file
write.csv(cross2, here("adni_data","cross_sectional_data_CLEAN.csv"), row.names = FALSE)
```

**Code B4. Function bank used in non-AUC/ROC analysis.**

```
#Function Bank and Variable Definitions for FNIH Project 2
#Crossectional Part of Study
#### Coded: Benjamin Saef
#### Date as of commenting and documentation - 05/28/2024
library(tidyr)
library(data.table)
library(RVAideMemoire)
library(pROC)
library(RColorBrewer)
library(MuMIn)

####### Variable creation - centralized location, just so that it can be made sure of across all
scripts

#analysis vars
#Covariates
sex_var <- "PTGENDER" #variable with sex
age_var <- "AGE" #variable with age
APOE_variable <- "APOE_genotype" #Variable with APOE genotypes

#C2N
pTau217_c2n <- "C2N_plasma_ptau217_ratio" #Variable representing p-tau217 ratio - c2n
pTau217conc_c2n <- "C2N_plasma_ptau217" #Variable representing p-tau217 concentration - c2n
abeta42_c2n <- "C2N_plasma_Abeta42" #Variable representing Ab42 - c2n
abeta40_c2n <- "C2N_plasma_Abeta42_Abeta40" #Variable representing Ab40 - c2n
abeta4240_c2n <- "C2N_plasma_Abeta42_Abeta40" #Variable representing Ab42/Ab40 - c2n

#Elecsys
abeta42_esys <- "Roche_plasma_Ab42" #Variable representing Ab42 - elecsys
abeta40_esys <- "Roche_plasma_Ab40" #Variable representing Ab40 - elecsys
abeta4240_esys <- "Roche_plasma_Ab42_Ab40" #Variable representing Ab42/Ab40 - elecsys
ptau_181_esys <- "Roche_plasma_ptau181" #Variable representing ptau181 - elecsys
GFAP_esys <- "Roche_plasma_GFAP" #Variable representing GFAP - elecsys
NfL_esys <- "Roche_plasma_NfL" #Variable representing NfL - elecsys

#Lumipulse
abeta42_lumi <- "Fuji_plasma_Ab42" #Variable representing Ab42 - lumipulse
abeta40_lumi <- "Fuji_plasma_Ab40" #Variable representing Ab40 - lumipulse
abeta4240_lumi <- "Fuji_plasma_Ab42_Ab40" #Variable representing Ab42/Ab40 - lumipulse
pTau217_lumi <- "Fuji_plasma_ptau217" #Variable representing p-tau217 ratio - lumipulse

#Simoa
alzpath_ptau217 <- "AlzPath_plasma_ptau217"  #Variable representing p-tau217 - AlzPath
janssen_ptau217 <- "Janssen_plasma_ptau217"  #Variable representing p-tau217 - Janssen

#Quanterix - Simoa
abeta42_quan <- "QX_plasma_Ab42" #Variable representing Ab42 - elecsys
abeta40_quan <- "QX_plasma_Ab40" #Variable representing Ab40 - elecsys
abeta4240_quan <- "QX_plasma_Ab42_Ab40" #Variable representing Ab42/Ab40 - elecsys
ptau_181_quan <- "QX_plasma_ptau181" #Variable representing ptau181 - elecsys
GFAP_quan <- "QX_plasma_GFAP" #Variable representing GFAP - elecsys
NfL_quan <- "QX_plasma_NfL" #Variable representing NfL - elecsys

#Variables for PET metrics
centiloid_Variable <- "CENTILOIDS" #Variable representing Centiloid
early_tau <- "MesialTemporal" #Variable representing Tau PET Centaur
late_tau <- "TemporoParietal"
atrophy <- "atrophy"
cdr_sum_of_boxes <- "CDR_SOB"
MMSE <- "MMSCORE"

#Making sure that the allele recordings fit neatly into the APOE levels we have selected
data.in[which(data.in[,APOE_variable]=="43"),APOE_variable] <- "34"
data.in[which(data.in[,APOE_variable]=="42"),APOE_variable] <- "24"
```

```
###Make sure APOE 33 is the reference level
data.in$APOE_Cat <- factor(data.in[,APOE_variable],
                           levels=c("33","34","24","22","23","44"))

#Function if needed to bring p-values in line with how they are outputted in SAS

pub_pval <- function(pval){

  if(pval < 0.0001){ pubpval <- "<0.0001"
  } else if(pval > 0.001){
    pubpval <- paste(signif(pval,2))
  }else{ pubpval <- paste(signif(pval,4))}
  return(pubpval)
}
#Function to Compare Rho
spdiff_rho <- function(data, indices) {
  d <- data[indices,] # allows boot to select sample
  cor_xz <- cor(d$x, d$z, method = "spearman",use = "complete.obs")
  cor_yz <- cor(d$y, d$z, method = "spearman",use = "complete.obs")
  diff <- abs(cor_xz) - abs(cor_yz)

  return(diff)
}
#Function to compare partial rhos
#Compare Rhos - needs fixing dependent on age, sex and APOE variables
#the covariate variables are currently hard coded because out how the bootstrap function works

spdiff_Partial_rho <- function(data, indices) {
  d <- data[indices,] # allows boot to select sample
  c_cov <- c("AGE","PTGENDER","APOE_Cat")   #change to reflect data
  cor_xz <- RVAideMemoire::pcor.test(d$x, d$z,d[,c_cov], method = "spearman",nrep = 1)
  cor_yz <- RVAideMemoire::pcor.test(d$y, d$z, d[,c_cov],method = "spearman",nrep = 1)
  diff <- abs(cor_xz$estimate) - abs(cor_yz$estimate)

  return(diff)
}
#Functions to create Characteristics Tables

###Creates entry focused around how many participants are positive for a specific variable (or just
which one has higher numerical code that would go
#second in a table)
positivity_dist_entry <- function(testvar,
                                  binvar,
                                  data_pos,
                                  data_neg,
                                  dataset,
                                  entryname){

  dataset <- dataset[which(complete.cases(dataset[,testvar])),]
  data_pos <- data_pos[which(complete.cases(data_pos[,testvar])),]
  data_neg <- data_neg[which(complete.cases(data_neg[,testvar])),]

  full_dist <- paste0(table(unlist(dataset[,testvar]))[2],
                      ", ",
round(100*table(unlist(dataset[,testvar]))[2]/length(unlist(dataset[,testvar])),2),
                      "%")
  pos_dist <- paste0(table(unlist(data_pos[,testvar]))[2],
                     ", ",
round(100*table(unlist(data_pos[,testvar]))[2]/length(unlist(data_pos[,testvar])),2),
                     "%")
  neg_dist <- paste0(table(unlist(data_neg[,testvar]))[2],
                     ", ",
```

```
round(100*table(unlist(data_neg[,testvar]))[2]/length(unlist(data_neg[,testvar])),2),
                     "%")

  chisq_test_Res <- chisq.test(unlist(dataset[,binvar]),
                               unlist(dataset[,testvar]),
                               correct=FALSE)
  pval_pre <- chisq_test_Res$p.value
  if(pval_pre < 0.0001){ pval <- "<0.0001"
  } else{ pval <- paste(signif(pval_pre,1))}

  posit_dist <-  c(entryname,
                   sum(!is.na(dataset[,testvar])),
                   full_dist,
                   sum(!is.na(data_neg[,testvar])),
                   neg_dist,
                   sum(!is.na(data_pos[,testvar])),
                   pos_dist,
                   pval)

  return(posit_dist)

}

####creates an entry focused around the median and first and third quantiles for continuous data
(usually non-normal)

create_analyte_dist_entry <-  function(Varid,binvar,g1_dataset,g2_dataset,full_dataset,labelid){
  g1_dataset <- g1_dataset
  g2_dataset <- g2_dataset
  full_dataset <- full_dataset
  pos_dat <- g1_dataset[which(!is.na(g1_dataset[,Varid])),]
  neg_dat <- g2_dataset[which(!is.na(g2_dataset[,Varid])),]
  all_dat <- full_dataset[which(!is.na(full_dataset[,Varid])),]
  all_sum <- signif(quantile(all_dat[,Varid],type=3), digits = 3)
  pos_sum <- signif(quantile(pos_dat[,Varid],type=3), digits = 3)
  neg_sum <- signif(quantile(neg_dat[,Varid],type=3), digits = 3)
  formula_of_model <- paste(Varid,binvar,sep="~")
  test_data <-  kruskal.test(as.formula(formula_of_model),data=all_dat)
  pval_pre <- test_data$p.value
  if(pval_pre < 0.0001){ pval <- "<0.0001"
  } else{ pval <- paste(signif(pval_pre,4))}
  entry_dist <- c(labelid,
                  sum(!is.na(all_dat[,Varid])),
                  paste0(all_sum["50%"]," (",
                         all_sum["25%"],
                         "-",all_sum["75%"],")"),
                  sum(!is.na(neg_dat[,Varid])),
                  paste0(neg_sum["50%"]," (",
                         neg_sum["25%"],
                         "-",neg_sum["75%"],")"),
                  sum(!is.na(pos_dat[,Varid])),
                  paste0(pos_sum["50%"]," (",
                         pos_sum["25%"],
                         "-",pos_sum["75%"],")"),
                  pval)

  return(entry_dist)
}

###Creates an entry focused around the means and standard deviation for the continuous data

create_mean_entry <-  function(Varid,binvar,g1_dataset,g2_dataset,full_dataset,labelid){
```

```
  pos_dat <- g1_dataset[which(!is.na(g1_dataset[,Varid])),]
  neg_dat <- g2_dataset[which(!is.na(g2_dataset[,Varid])),]
  all_dat <-full_dataset[which(!is.na(full_dataset[,Varid])),]
  all_sum <- signif(mean(all_dat[,Varid],na.rm=T), digits = 3)
  pos_sum <- signif(mean(pos_dat[,Varid],na.rm=T), digits = 3)
  neg_sum <- signif(mean(neg_dat[,Varid],na.rm=T), digits = 3)
  all_sd <- signif(sd(all_dat[,Varid],na.rm=T), digits = 3)
  pos_sd <- signif(sd(pos_dat[,Varid],na.rm=T), digits = 3)
  neg_sd <- signif(sd(neg_dat[,Varid],na.rm=T), digits = 3)

  formula_of_model <- paste(Varid,binvar,sep="~")
  test_data <-  kruskal.test(as.formula(formula_of_model),data=all_dat)
  pval_pre <- test_data$p.value
  if(pval_pre < 0.0001){ pval <- "<0.0001"
  } else{ pval <- paste(signif(pval_pre,4))}
  entry_dist <- c(labelid,
                  sum(!is.na(all_dat[,Varid])),
                  paste0(all_sum," (",
                         all_sd,")"),
                  sum(!is.na(neg_dat[,Varid])),
                  paste0(neg_sum," (",
                         neg_sd,")"),
                  sum(!is.na(pos_dat[,Varid])),
                  paste0(pos_sum," (",
                         pos_sd,")"),
                  pval)

  return(entry_dist)
}

###The following two are circumstantial and only to be used for their exact purpose. They may
require re-coding if things are differently
#coded in the target dataset

#Race (Black/White/Other) - need to be coded as 2/1/3 respectively
create_race_entry <- function(Varid,binvar,g1_dataset,g2_dataset,full_dataset,labelid){

  pos_dat <- g1_dataset[which(!is.na(g1_dataset[,Varid])),]
  neg_dat <- g2_dataset[which(!is.na(g2_dataset[,Varid])),]
  all_dat <-full_dataset[which(!is.na(full_dataset[,Varid])),]

full_dist <- paste0(table(all_dat[,Varid])["2"],"/",
                         table(all_dat[,Varid])["1"],"/",
                         table(all_dat[,Varid])["3"])
neg_dist <- paste0(table(neg_dat[,Varid])["2"],"/",
                          table(neg_dat[,Varid])["1"],"/",
                          table(neg_dat[,Varid])["3"])
pos_dist <- paste0(table(pos_dat[,Varid])["2"],"/",
                          table(pos_dat[,Varid])["1"],"/",
                          table(pos_dat[,Varid])["3"])

chisq_race <- chisq.test(all_dat[,binvar],
                         as.factor(all_dat[,Varid]),
                         correct=FALSE)

racial_characteristcs <- c("Race (Black/White/Other)",
                          sum(!is.na(all_dat[,Varid])),
                          full_dist,
                          sum(!is.na(neg_dat[,Varid])),
                          neg_dist,
                          sum(!is.na(pos_dat[,Varid])),
                          pos_dist,
                          signif(chisq_race$p.value,2))

return(racial_characteristcs)
```

```
}

###Creating this just in case we decide to do APOE in a different way this time
create_apoe_entry <- function(Varid,binvar,g1_dataset,g2_dataset,full_dataset,labelid){

  pos_dat <- g1_dataset[which(!is.na(g1_dataset[,Varid])),]
  neg_dat <- g2_dataset[which(!is.na(g2_dataset[,Varid])),]
  all_dat <-full_dataset[which(!is.na(full_dataset[,Varid])),]

  full_dist <- paste0(table(all_dat[,Varid])["22"],"/",
                      table(all_dat[,Varid])["23"],"/",
                      table(all_dat[,Varid])["24"],"/",
                      table(all_dat[,Varid])["33"],"/",
                      table(all_dat[,Varid])["34"],"/",
                      table(all_dat[,Varid])["44"])
  neg_dist <- paste0(table(neg_dat[,Varid])["22"],"/",
                     table(neg_dat[,Varid])["23"],"/",
                     table(neg_dat[,Varid])["24"],"/",
                     table(neg_dat[,Varid])["33"],"/",
                     table(neg_dat[,Varid])["34"],"/",
                     table(neg_dat[,Varid])["44"])
  pos_dist <- paste0(table(pos_dat[,Varid])["22"],"/",
                     table(pos_dat[,Varid])["23"],"/",
                     table(pos_dat[,Varid])["24"],"/",
                     table(pos_dat[,Varid])["33"],"/",
                     table(pos_dat[,Varid])["34"],"/",
                     table(pos_dat[,Varid])["44"])

  chisq_apoe <- chisq.test(all_dat[,binvar],
                           as.factor(all_dat[,Varid]),
                           correct=FALSE)

  apoe_characteristcs <- c("APOE Genotype (22/23/24/33/34/44)",
                             sum(!is.na(all_dat[,Varid])),
                             full_dist,
                             sum(!is.na(neg_dat[,Varid])),
                             neg_dist,
                             sum(!is.na(pos_dat[,Varid])),
                             pos_dist,
                             signif(chisq_apoe$p.value,2))

  return(apoe_characteristcs)

}

#### Pulling all functions together to construct tables

#dataset - input data - needs to contain all variables in binvar and varlist
#binvar and binvarlab - the binary variable which will determine the columns after the "all"
columns and the label will determine
#how the columns are named. both of these need to be strings
#varlist - list of variables, simple as it sounds, it needs to be a vector of strings
#typelist - here's where things get a little complicated. this needs to be the exact same length as
the varlist, as it's what
#determines how each entry is handled
#Here are the type listings
#A - this is the code for any analyte/non-normal variable. It will output an entry with the median
and interquartile range
#T - this code is very similar to the previous one, it's only use specifically for Tracers when
dealing with different measurement standards for
#SUVR, the namelist entry, where the labels are, will be used to determine which tracer this entry
will represent
```

```
#P - this is the code that is used to create entries based on binary variables. Specifically the
"positive" status that is measured
#is whatever R would place in the second slot of the variable when placed into the table function
#This is usually used for things like disease or biomarker status, but can be used for other things
as well
#namelist - list of labels that go along with varlist and typelist, needs to be in order
corresponding to those two
#filout - output destination for resulting excel table
char_table_full_bin <- function(dataset,binvar,
                                binvarlab,varlist,
                                typelist,namelist,
                                fileout){
  require(openxlsx)
  require(tidyverse)
  looplength <- length(varlist)
  dnames <- c("All",paste0(binvarlab," Negative"),
              paste0(binvarlab," Positive"))

  d1 = dataset[which(dataset[,binvar]==1),]
  d2 = dataset[which(dataset[,binvar]==0),]
  characteristics_table <- data.frame(matrix(nrow=looplength,ncol=(length(dnames)*2)+2))
  colnames(characteristics_table) <- c("Characteristic",
                                       paste(rep(dnames,each=2),
                                             c("n","Values"),sep=" "),"p=")

  for(i in 1:looplength){
    vartype <- typelist[i]
    varid <- varlist[i]
    label_name <- namelist[i]
    if(vartype=="A"){
      newentry  <- create_analyte_dist_entry(varid,
                                             binvar,
                                             d1,
                                             d2,
                                             dataset,
                                             label_name)
    }else if(vartype=="P"){
      newentry  <- positivity_dist_entry(varid,
                                         binvar,
                                         d1,
                                         d2,
                                         dataset,
                                         label_name)
    }

    else if(vartype=="T"){
      newentry <- create_mean_entry(varid,
                                    binvar,
                                    d1[which(d1$Tracer==label_name),],
                                    d2[which(d2$Tracer==label_name),],
                                    dataset[which(dataset$Tracer==label_name),],
                                    paste0(label_name," SUVR"))
    } else if(vartype=="M"){
      newentry  <- create_mean_entry(varid,
                                     binvar,
                                     d1,
                                     d2,
                                     dataset,
                                     label_name)
    } else if(vartype=="R"){
      newentry  <- create_race_entry(varid,
                        binvar,
                        d1,
                        d2,
                        dataset,
                        label_name)
```

```
    } else if(vartype=="4"){
      newentry  <-create_apoe_entry(varid,
                        binvar,
                        d1,
                        d2,
                        dataset,
                        label_name)
    }
    characteristics_table[i,] <- newentry
  }

  write.xlsx(characteristics_table,
             fileout,
             rowNames=FALSE,
             overwrite = TRUE)
  return(characteristics_table)

}

###Functions to run spearmans and partial spearmans

#Function to evaluate the comparison standard for the Spearman comparisons
#Just outputs the top analyte variable name
Eval_comp_standard <- function(data_input, #Dataset
                               var_a, #Phenotype - outcome variable
                               vars_list){ #list of analytes

  length_analyte <- length(vars_list)
  out_res <- as.data.frame(matrix(nrow=length_analyte,ncol=2))
  colnames(out_res) <- c("varname","rho")

  for(i_analyte in 1:length_analyte){

    mdl_rho=cor.test(data_input[,vars_list[i_analyte]],
                 data_input[,var_a], method = "spearman")
    out_res$varname[i_analyte]=vars_list[i_analyte]
    out_res$rho[i_analyte]=mdl_rho$estimate

  }

  out_res <- out_res[order(-abs(out_res$rho)),]
  TopVar <- out_res[1,"varname"]
  return(TopVar)
}

####
#Function to evaluate the comparison standard for the Partial Spearman comparisons
#Just outputs the top analyte variable name
Eval_comp_standard_partial <- function(data_input,#Dataset
                               var_a, #Phenotype - outcome variable
                               cov_var, #Covariates - Age, Gender, APOE genotype
                               vars_list){ #list of analytes
  df <- data_input
  length_analyte <- length(vars_list)
  out_res <- as.data.frame(matrix(nrow=length_analyte,ncol=2))
  colnames(out_res) <- c("varname","rho")

  for(i_analyte in 1:length_analyte){
    df2 <- df[which(complete.cases(df[,c(var_a,vars_list[i_analyte],cov_var)])),]
    mdl_rho=RVAideMemoire::pcor.test(df2[,var_a],
                                   df2[,vars_list[i_analyte]],
                                   df2[,cov_var],method = "spearman",
                                   conf.level = 0.95, nrep = 2)
```

```
    out_res$varname[i_analyte]=vars_list[i_analyte]
    out_res$rho[i_analyte]=mdl_rho$estimate

  }

  out_res <- out_res[order(-abs(out_res$rho)),]
  TopVar <- out_res[1,"varname"]
  return(TopVar)
}

###
spearman_plasma_runs <- function(data_input, # Data inputted
                                 var_a,  # variable A - continuous variable being correlated
                                 vars_to_cor,# vector of strings - continuous variables being
correlated with variable A
                                 var_comp, #variable in vars_to_cor vector that is used as standard
for comparisons (unadjusted)
                                 var_comp_p, #variable in vars_to_cor vector that is used as
standard for comparisons (adjusted)
                                 cov_var,    # covariates - variables used as covariates (age, sex,
APOE normally)
                                 analytes_labels, #labels for analytes in vars to cor
                                 group_lab){ #groups for analytes in vars to cor
  require(RVAideMemoire)
  require(boot)
 df  <- data_input
 print_headers <- c("Biomarker","group","Rho","Rho_L","Rho_H","p",

paste("pComp",var_comp,sep="_"),"Partial_Rho","PRho_L","PRho_H","p_partial",paste("Par_pComp",var_c
omp_p,sep="_"))
 results_print=data.frame(matrix(nrow = length(vars_to_cor),
                                 ncol = length(print_headers)))
 colnames(results_print) <- print_headers

 for(i_plasma in 1:length(vars_to_cor)){
   results_print$Biomarker[i_plasma]=analytes_labels[i_plasma]
   results_print$group[i_plasma]=group_lab[i_plasma]
   ###Spearman (unadjusted) Correlation Statistics below
   mdl_rho=cor.test(df[,vars_to_cor[i_plasma]],
                    df[,var_a], method = "spearman")
   b=RVAideMemoire::spearman.ci(df[,vars_to_cor[i_plasma]],
                                df[,var_a],
                                nrep = 1000,conf.level = 0.95)

   df2=data_input[,c(var_a,var_comp,vars_to_cor[i_plasma])]
   colnames(df2)=c("z","x","y")

   boot_results = boot(data = df2, statistic = spdiff_rho, R = 12500)
   results_under_H0 <- boot_results$t - mean(boot_results$t)
   boot_pvalue <- mean(abs(results_under_H0) >= abs(boot_results$t0))
   results_print[i_plasma,paste("pComp",var_comp,sep="_")]=pub_pval(boot_pvalue)

   results_print$Rho[i_plasma]=mdl_rho$estimate
   results_print$Rho_L[i_plasma]=b$conf.int[1]
   results_print$Rho_H[i_plasma]=b$conf.int[2]
   results_print$p[i_plasma]=paste(pub_pval(mdl_rho$p.value))

   ###Partial Correlation Statistics below
   df_for_cor=df[complete.cases(df[,c(var_a,vars_to_cor[i_plasma],cov_var)]),]
 mdl_rho=RVAideMemoire::pcor.test(df_for_cor[,var_a],
                                  df_for_cor[,vars_to_cor[i_plasma]],
                                  df_for_cor[,cov_var],method = "spearman",
                                  conf.level = 0.95, nrep = 1000)
```

```
 df2=df_for_cor[,c(var_a,var_comp_p,vars_to_cor[i_plasma],cov_var)]
 colnames(df2)=c("z","x","y",cov_var)

 set.seed(12345)
 boot_results = boot(data = df2, statistic = spdiff_Partial_rho, R = 12500)
 boot_ci_norm <- boot.ci(boot_results, type = "norm")
 results_under_H0 <- boot_results$t - mean(boot_results$t)
 boot_pvalue <- mean(abs(results_under_H0) >= abs(boot_results$t0))

 results_print$Partial_Rho[i_plasma]=mdl_rho$estimate
 results_print$PRho_L[i_plasma]=mdl_rho$conf.int[1]
 results_print$PRho_H[i_plasma]=mdl_rho$conf.int[2]
 results_print$p_partial[i_plasma]=paste(pub_pval(mdl_rho$p.value))
 results_print[i_plasma,paste("Par_pComp",var_comp_p,sep="_")]=pub_pval(boot_pvalue)
 }
 results_print <- results_print[order(results_print$group,-abs(results_print$Rho)),]
 return(results_print)

}

#The resulting dataset will include the following columns:
#Biomarker - Biomarker Name (from the analytes_labels vector)
#Rho - Spearman correlation statistic
#Rho_L - spearman rho - low confidence interval bound
#Rho_H - spearman rho - high confidence interval bound
#p - spearman correlation p-value
#pComp_'Analyte' - comparison p-value for the spearman correlations to the lead 'Analyte' chosen at
the top of the function
#Partial_Rho - Partial Spearman's Rho
#PRho_L - Partial Rho - Low confidence interval bound
#PRho_H - Partial Rho - High confidence interval bound
#p_partial - partial correlation p-value
#Par_pComp_'Analyte' - comparison p-value for the partial correlations to the lead 'Analyte' chosen
at the top of the function

###Functions to create forest plots

#creating function to create index. this just intakes data from spearman_plasma_runs function (DF1)
and outputs the plot index
#The plot index just has the biomarker, company, and order (plot_index) in it
index_biomarker_forest <- function(DF1){

  require(dplyr)
  df <- data.frame(DF1[,c("company","group","Rho","Biomarker")])
  rownames(df) <- NULL
  df$Rho <- as.numeric(df$Rho)
  df$company <- factor(df$company)
  df$company <-factor(df$company, levels = c(unique(df[order(-abs(df$Rho)),"company"])))
  DF <- df %>% group_by(group) %>%
    arrange(company,desc(abs(Rho))) %>%
    ungroup() %>%
    mutate(plot_index = desc(row_number()))

  return(DF[,c("Biomarker","company","plot_index")])
}

####Create Output forest plot from spearman function
##As of right now, is company agnostic, but not analyte agnostic due to hard coded analyte labels

spearman_gg_formanuscript <- function(DF1, #output from spearman_plasma_runs function
                                      breaks1, # a vector representing the axis ticks
                                      indicatorline, # position dashed line that goes on the
horizontal axis (coordinate flip - so technically y)
```

```
                                      df_plot_index, # output from index_biomarker_forest function,
used to order the forest plot
                                      title_plot="", # Put the title of the forest plot here
                                      rev_axis=FALSE, #if you'd like to reverse the horizontal Axis
                                      labels_all=FALSE, # whether or not you want the axis ticks
for analytes and the company labels
                                      amytag=FALSE){ #only flag this if wanting to hard set max of
the range

  require(dplyr)
  df <- data.frame(DF1[,c("company","Rho","Rho_L","Rho_H","Biomarker")])
  df <- merge(df,df_plot_index,by=c("Biomarker","company"))
  rownames(df) <- NULL
  colnames(df) <- c("analyte","company","AUC","AUC_Lower","AUC_Upper","plot_index")

  company_count <- table(df_plot_index$company)
  Index1 <- -1
  Company1 <- names(company_count)[1]
  Index2 <- Index1 - company_count[1]
  Company2 <- names(company_count)[2]
  Index3 <- Index2 - company_count[2]
  Company3 <- names(company_count)[3]
  Index4 <- Index3 - company_count[3]
  Company4<- names(company_count)[4]
  Index5 <- Index4 - company_count[4]
  Company5 <- names(company_count)[5]
  Index6 <- Index5 - company_count[5]
  Company6 <- names(company_count)[6]
  Index7 <- Index6 - company_count[6]

  df$AUC <- as.numeric(df$AUC)
  df$AUC_Lower <- as.numeric(df$AUC_Lower)
  df$AUC_Upper <- as.numeric(df$AUC_Upper)
  df$company <- factor(df$company)

  df$company <-factor(df$company, levels = c(unique(df[order(-abs(df$AUC)),"company"])))
  DF <- df

  DF$analyte_c <- NA
  DF[which(DF$analyte=="p-tau217 (pg/ml)"),"analyte_c"] <- "p-Tau 217"
  DF[grep(paste0("A","\U03B2","42/A","\U03B2","40"),DF$analyte),"analyte_c"] <- "Ab42/Ab40"
  DF[which(DF$analyte=="p-tau217 ratio (%)"),"analyte_c"] <- "p-Tau 217 ratio"
  DF[grep("GFAP",DF$analyte),"analyte_c"] <- "GFAP"
  DF[grep("NfL",DF$analyte),"analyte_c"] <- "NfL"
  DF[grep("p-tau181",DF$analyte),"analyte_c"] <- "p-Tau 181"

  DF$analyte_c <- factor(DF$analyte_c,levels=c("p-Tau 217 ratio",
                                             "p-Tau 217",
                                             "Ab42/Ab40",
                                             "p-Tau 181",
                                             "GFAP","NfL"))

  analyte_colors <- c("Ab42/Ab40" = "blue",
                      "p-Tau 217" = "#008000",
                      "p-Tau 181" = "#00FF00",
                      "NfL" = "maroon",
                      "GFAP" = "red",
                      "p-Tau 217 ratio" = "#145A32")

DF[which(DF$analyte_c=="Ab42/Ab40"),"AUC"] <- DF[which(DF$analyte_c=="Ab42/Ab40"),"AUC"]*-1
DF[which(DF$analyte_c=="Ab42/Ab40"),"AUC_Lower"] <-
DF[which(DF$analyte_c=="Ab42/Ab40"),"AUC_Lower"]*-1
```

```
DF[which(DF$analyte_c=="Ab42/Ab40"),"AUC_Upper"] <-
DF[which(DF$analyte_c=="Ab42/Ab40"),"AUC_Upper"]*-1

round_any = function(x, accuracy, f=round){f(x/ accuracy) * accuracy}

min_break_val <- min(breaks1)
min_y <-  round_any(min(c(DF$AUC_Lower,DF$AUC_Upper,min_break_val)) ,accuracy = 0.01 , f = floor)
min_y_text <- min_y-0.1
text_start <- min_y_text

max_y <-  round_any(max(DF$AUC_Upper) ,accuracy = 0.2 , f = ceiling)
max_y <- max(c(max(breaks1),max_y))
if(amytag==TRUE){
  max_y <- 0.9
}

if(rev_axis==TRUE){
  max_y <- max_y+0.1
  text_start <- max_y

}

  Spearman_test_plot <-  ggplot(data=DF,
                                aes(x = plot_index, #uses index as the x-axis (flipped to y later)
                                    y = AUC, # y-axis is spearman rho, flipped to x-axis later
                                    ymin = min_y, ymax = max_y ))+ #limited of y-axis (flipped to x
later)
    geom_rect(aes(xmin = Index1+0.5, xmax = Index2+0.5, ymin = min_y_text, ymax = max_y),
              fill = "#E5E4E2", alpha = 0.04) +
    geom_rect(aes(xmin = Index3+0.5, xmax = Index4+0.5, ymin = min_y_text, ymax = max_y),
              fill = "#E5E4E2", alpha = 0.04) +
    geom_rect(aes(xmin = Index5+0.5, xmax = Index6+0.5, ymin = min_y_text, ymax = max_y),
              fill = "#E5E4E2", alpha = 0.04) +
    geom_hline(aes(fill="black"),yintercept =indicatorline, linetype=2)+ #creates horizontal
(flipped to vertical) line at 0
    geom_point(aes(col=analyte_c))+ #sets the coloring based on whether its the summary measure or
not
    geom_errorbar(aes(ymin=(AUC_Lower), ymax=(AUC_Upper),col=analyte_c),width = 0, cex = 1,size=4)+
#formatting of forest lines
      labs(x = "", y = expression(paste("Spearman "   ,rho)))+theme_classic()+ #gives the title
based on the function call variable
    theme(plot.title = element_text(hjust = 0.5,size = 10), #these are all text formatting
          axis.ticks.y=element_line(size = 1, color = "black"),
          axis.text.x=element_text(size=6),
          axis.text.y=element_text(size=6),
          axis.title=element_text(size=7,face="bold"),
          strip.text.y = element_blank(),
          legend.position =  "none")+ #no legend
    scale_color_manual(values=analyte_colors)+
    scale_fill_manual(values = analyte_colors)+ #makes sure dots and lines are colored based on
color code
    coord_flip()+ggtitle(title_plot)

  if(rev_axis==TRUE){
    Spearman_test_plot <- Spearman_test_plot+scale_y_reverse(breaks=breaks1)

  }else {
    Spearman_test_plot <- Spearman_test_plot+scale_y_continuous(breaks=breaks1)

  }

  if(labels_all==TRUE){
Spearman_test_plot <- Spearman_test_plot+
```

```
      geom_text(aes(x = Index1, y = text_start, label = Company1, hjust =
0),size=3,family="Calibri") +
      geom_text(aes(x = Index2, y = text_start, label = Company2, hjust =
0),size=3,family="Calibri") +
      geom_text(aes(x = Index3, y = text_start, label = Company3, hjust =
0),size=3,family="Calibri") +
      geom_text(aes(x = Index4, y = text_start, label = Company4, hjust =
0),size=3,family="Calibri") +
      geom_text(aes(x = Index5, y = text_start, label = Company5, hjust =
0),size=3,family="Calibri") +
      geom_text(aes(x = Index6, y = text_start, label = Company6, hjust =
0),size=3,family="Calibri")+
     scale_x_continuous(breaks=DF$plot_index,labels=DF$analyte)
  }
  if(labels_all==FALSE){
    Spearman_test_plot <- Spearman_test_plot + theme(axis.text.y=element_blank(),
                                                      axis.ticks.y=element_blank())

  }

  return(Spearman_test_plot)

}

####This function's sole purpose is to clean up output from the spearman_plasma_runs functions
specifically tuned to our dataset and variables
## Will output a dataset with the following columnts and structure:
##company - self explanatory - company who made the assay
##measure    - which analyte is being represented
##Rho  - Spearman's Rho with the particular analyte and the outcome, will also include 95%
confidence interval - Rho (Rho Lower Bound - Rho Upper Bound)
##pcomp      - comparison between the top correlator Rho and the analyte - will read REFERENCE if
analyte is the reference
##pRho - Partial Spearman's Rho with the particular analyte and the outcome, will also include 95%
confidence interval - Rho (Rho Lower Bound - Rho Upper Bound)
##parcomp    - comparison between the top correlator partial Rho and the analyte - will read
REFERENCE if analyte is the reference

clean_spearman_table <- function(DF1){ # output from dataset

  pparnormal_comp <- grep("Par_pComp_",colnames(DF1))
  pnormal_comp  <- grep("pComp_",colnames(DF1))[1]
  Reference_bio  <- DF1[which(abs(DF1$Rho)==max(abs(DF1$Rho))),"Biomarker"]
  Reference_comp  <- as.character(DF1[which(abs(DF1$Rho)==max(abs(DF1$Rho))),"company"])
  Pref_bio   <- DF1[which(abs(DF1$Partial_Rho)==max(abs(DF1$Partial_Rho))),"Biomarker"]
  Pref_com   <- as.character(DF1[which(abs(DF1$Partial_Rho)==max(abs(DF1$Partial_Rho))),"company"])

 spearman_out  <- data.frame(matrix(nrow=nrow(DF1),ncol=6))

 colnames(spearman_out) <- c("company","measure","Rho","pcomp","pRho","parcomp")

 spearman_out$company <- as.factor(DF1$company)
 spearman_out$company <- factor(spearman_out$company,levels = c("C2N", "Fujirebio", "ALZpath",
"Janssen", "Roche", "Quanterix"))

 spearman_out$measure <- DF1$Biomarker
 spearman_out$Rho <- paste0(format(round(DF1$Rho,3),nsmall=3)," (",
                            format(round(DF1$Rho_L,3),nsmall=3)," to ",
                            format(round(DF1$Rho_H,3), nsmall=3),")")

 spearman_out[which(spearman_out$company==Reference_comp &
spearman_out$measure==Reference_bio),"pcomp"] <- "REFERENCE"
```

```
DF_comp <-  DF1[which(!(spearman_out$company==Reference_comp &
spearman_out$measure==Reference_bio)),]
DF_comp <- as.data.frame(DF_comp)
DF_comp$P_prep <- DF_comp[,pnormal_comp]
DF_comp[which(DF_comp[,pnormal_comp]=="<0.0001"),"P_prep"]  <- 0.0001
DF_comp$pcomp  <- format(round(p.adjust(as.numeric(DF_comp[,"P_prep"]),method = "BH"),4),nsmall=4)
DF_comp[which(DF_comp[,pnormal_comp]=="<0.0001"),"pcomp"] <- "<0.0001"
 spearman_out[which(spearman_out$company==Pref_com & spearman_out$measure==Pref_bio),"parcomp"] <-
"REFERENCE"
for(i_comparisons in 1:nrow(DF_comp)){

topind <-  which(spearman_out$company==DF_comp$company[i_comparisons] &
spearman_out$measure==DF_comp$Biomarker[i_comparisons])
spearman_out[topind,"pcomp"] <-  DF_comp$pcomp[i_comparisons]

}

spearman_out$pRho <- paste0(format(round(DF1$Partial_Rho,3),nsmall=3)," (",
                           format(round(DF1$PRho_L,3),nsmall=3)," to ",
                           format(round(DF1$PRho_H,3), nsmall=3),")")

DF_comp <-  DF1[which(!(spearman_out$company==Pref_com & spearman_out$measure==Pref_bio)),]
DF_comp <- as.data.frame(DF_comp)
DF_comp$Partial_prep <- DF_comp[,ppartial_comp]
DF_comp[which(DF_comp[,ppartial_comp]=="<0.0001"),"Partial_prep"]  <- 0.0001
DF_comp$ppart  <- format(round(p.adjust(as.numeric(DF_comp[,"Partial_prep"]),method =
"BH"),4),nsmall=4)
DF_comp[which(DF_comp[,ppartial_comp]=="<0.0001"),"ppart"] <- "<0.0001"

for(i_comparisons in 1:nrow(DF_comp)){

  topind <-  which(spearman_out$company==DF_comp$company[i_comparisons] &
spearman_out$measure==DF_comp$Biomarker[i_comparisons])
  spearman_out[topind,"parcomp"] <-  DF_comp$ppart[i_comparisons]

}
spearman_out[which(spearman_out$company==Pref_com & spearman_out$measure==Pref_bio),"parcomp"] <-
"REFERENCE"
spearman_out <- spearman_out[order(spearman_out$company),]

return(spearman_out)
}

##creates heatmap from data and a "ref_data" table
##the ref data must include the following:
# analyte code - code that represents which analyte the measure represents
# var_id - the variable name for the analyte
# fullname - the label for the variable that you want to show up on the heatmap
## data.in is just the dataset
## order_cor_analyte is what analyte's correlations do you want to determine the order of the
heatmap

create_cor_heatmap_for_project <-
function(data.in,ref_data,order_cor_analyte="C2N_plasma_ptau217_ratio"){
  require(tidyverse)
  require(Hmisc)
  require(reshape2)

  mydata.cor = cor(data.in[,ref_data$var_id], method = c("spearman"),use = "pairwise.complete.obs")

  ref_data$ptau_correlation <- abs(data.frame(mydata.cor)[,order_cor_analyte])
  ref_reorderd <-  ref_data %>% group_by(analyte_code) %>% mutate(mx = max(ptau_correlation)) %>%
    arrange(mx,ptau_correlation)
```

```
  ref_reorderd <- data.frame(ref_reorderd)

  mydata.cor <- mydata.cor[ref_reorderd$var_id,ref_reorderd$var_id]
  melted_cormat <- melt(mydata.cor)

  melted_cormat$Var1_Lab <- NA
  melted_cormat$Var2_Lab <- NA
  for(i_cycle in 1:nrow(ref_reorderd)){
    melted_cormat[which(melted_cormat$Var1==ref_reorderd$var_id[i_cycle]),"Var1_Lab"] <-
ref_reorderd$fullname[i_cycle]
    melted_cormat[which(melted_cormat$Var2==ref_reorderd$var_id[i_cycle]),"Var2_Lab"] <-
ref_reorderd$fullname[i_cycle]
  }

  melted_cormat$Var1_Lab <- factor(melted_cormat$Var1_Lab,levels=rev(ref_reorderd$fullname))
  melted_cormat$Var2_Lab <- factor(melted_cormat$Var2_Lab,levels=ref_reorderd$fullname)

  heatcolor_pallette <- c("#0087B3","white","red","black")
  color_scale <- c(0,0.5,0.999, 1)

  heatmap_of_cor <-  ggplot(melted_cormat, aes(Var1_Lab, Var2_Lab)) +
    geom_tile(aes(fill = abs(value))) +
    geom_text(aes(label = format(round(value, 2), nsmall = 2)),size=2) +
    scale_fill_gradientn(colours=heatcolor_pallette,values=color_scale)+
    theme(legend.position="none",
          axis.text.x = element_text(angle = 45, vjust = 1,
                                     size = 6, hjust = 1),axis.text.y=element_text(size=6),
          axis.title = element_blank())
  return(heatmap_of_cor)
}

##creates heatmap from data and a "ref_data" table
##the ref data must include the following:
# analyte code - code that represents which analyte the measure represents
# var_id - the variable name for the analyte
# fullname - the label for the variable that you want to show up on the heatmap
## data.in is just the dataset
## ref order is how you want to order the analyte groups
## order_cor_analyte is what analyte's correlations do you want to determine the order of the
heatmap within the analyte groups

create_cor_heatmap_for_project_custom <-
function(data.in,ref_data,reforder,order_cor_analyte="C2N_plasma_ptau217_ratio"){
  require(tidyverse)
  mydata.cor = cor(data.in[,ref_data$var_id], method = c("spearman"),use = "pairwise.complete.obs")

  ref_data$ptau_correlation <- abs(data.frame(mydata.cor)[,order_cor_analyte])
  ref_data$analyte_code <- factor(ref_data$analyte_code,levels=reforder)
  ref_reorderd <-  ref_data[order(ref_data$analyte_code,abs(ref_data$ptau_correlation)),]
  ref_reorderd <- data.frame(ref_reorderd)

  mydata.cor <- mydata.cor[ref_reorderd$var_id,ref_reorderd$var_id]
  melted_cormat <- melt(mydata.cor)

  melted_cormat$Var1_Lab <- NA
  melted_cormat$Var2_Lab <- NA
  for(i_cycle in 1:nrow(ref_reorderd)){
    melted_cormat[which(melted_cormat$Var1==ref_reorderd$var_id[i_cycle]),"Var1_Lab"] <-
ref_reorderd$fullname[i_cycle]
```

```
    melted_cormat[which(melted_cormat$Var2==ref_reorderd$var_id[i_cycle]),"Var2_Lab"] <-
ref_reorderd$fullname[i_cycle]
  }

  melted_cormat$Var1_Lab <- factor(melted_cormat$Var1_Lab,levels=rev(ref_reorderd$fullname))
  melted_cormat$Var2_Lab <- factor(melted_cormat$Var2_Lab,levels=ref_reorderd$fullname)

  heatcolor_pallette <- c("#0087B3","white","red","black")
  color_scale <- c(0,0.5,0.999, 1)

  heatmap_of_cor <-  ggplot(melted_cormat, aes(Var1_Lab, Var2_Lab)) +
    geom_tile(aes(fill = abs(value))) +
    geom_text(aes(label = format(round(value, 2), nsmall = 2)),size=2) +
    scale_fill_gradientn(colours=heatcolor_pallette,values=color_scale)+
    theme(legend.position="none",
          axis.text.x = element_text(angle = 45, vjust = 1,
                                     size = 6, hjust = 1),axis.text.y=element_text(size=6),
          axis.title = element_blank())
  return(heatmap_of_cor)
}

###

make_scatter_figure <- function(data.in, #input data
                                ref_data, #similar to ref_data for heatmaps, but also need group
variable with the codes for the company
                                metric,  # outcome variable ID
                                results_dataset, #results dataset from spearman_plasma_runs
                                phenotype_name, #outcome name
                                rev_axis=F){ #do you want to reverse the x-axis

  require(tidyr)
  require(cowplot)

  graph_data <- data.in[which(!is.na(data.in[,metric])),]
  pheno_name <- phenotype_name
  top_performers <- results_dataset %>% group_by(group) %>% filter(abs(Rho)==max(abs(Rho)))
  top_performers <- as.data.frame(top_performers)

  graph_data$participant_condition <- NA
  graph_data[which(graph_data$CENTILOIDS_10==0 & graph_data$CDR_10==0), "participant_condition"] <-
"A" #Blue: amyloid PET negative, CDR 0
  graph_data[which(graph_data$CENTILOIDS_10==0 & graph_data$CDR_10==1), "participant_condition"] <-
"B" #Yellowish green: amyloid PET negative, CDR >0
  graph_data[which(graph_data$CENTILOIDS_10==1 & graph_data$CDR_10==0), "participant_condition"] <-
"C" #Orange: amyloid PET positive, CDR 0
  graph_data[which(graph_data$CENTILOIDS_10==1 & graph_data$CDR_10==1), "participant_condition"] <-
"D" #Red: amyloid PET positive, CDR >0

  color_palette_scatter <- c("A" = "blue",
                             "B" = "#9acd32",
                             "C" = "orange",
                             "D" = "red")

  i_group <- "c2n"
  i_lab <- top_performers[which(top_performers$group==i_group),"Biomarker"]
  best_company <- ref_data[which(ref_data$group=="c2n" & ref_data$label==i_lab),"var_id"]

  scatter_c2n <- ggplot(data=graph_data %>%
                          arrange(participant_condition),
                        aes_string(x=metric,y=best_company))+
    geom_point(aes(colour=factor(participant_condition)),size=2,alpha=0.5)+
    scale_color_manual(values = color_palette_scatter)+
    theme_classic()+xlab(pheno_name)+ylab(paste("C2N \n",i_lab)) +
```

```
theme(legend.position="none",axis.text.x=element_text(size=12),axis.text.y=element_text(size=12),
          axis.title=element_text(size=12))

  i_group <- "fuji"
  i_lab <- top_performers[which(top_performers$group==i_group),"Biomarker"]
  best_company <- ref_data[which(ref_data$group=="fuji" & ref_data$label==i_lab),"var_id"]

  scatter_fuji <- ggplot(data=graph_data %>%
                           arrange(participant_condition),aes_string(x=metric,y=best_company))+
    geom_point(aes(colour=factor(participant_condition)),size=2,alpha=0.5)+
    scale_color_manual(values = color_palette_scatter)+
    theme_classic()+xlab(pheno_name)+ylab(paste("Fujirebio \n",i_lab)) +

theme(legend.position="none",axis.text.x=element_text(size=12),axis.text.y=element_text(size=12),
          axis.title=element_text(size=12))

  i_group <- "jan"
  i_lab <- top_performers[which(top_performers$group==i_group),"Biomarker"]
  best_company <- ref_data[which(ref_data$group=="jan" & ref_data$label==i_lab),"var_id"]

  scatter_janssen <- ggplot(data=graph_data %>%
                              arrange(participant_condition),aes_string(x=metric,y=best_company))+
    geom_point(aes(colour=factor(participant_condition)),size=2,alpha=0.5)+
    scale_color_manual(values = color_palette_scatter)+
    theme_classic()+xlab(pheno_name)+ylab(paste("Janssen \n",i_lab)) +

theme(legend.position="none",axis.text.x=element_text(size=12),axis.text.y=element_text(size=12),
          axis.title=element_text(size=12))

  i_group <- "roche"
  i_lab <- top_performers[which(top_performers$group==i_group),"Biomarker"]
  best_company <- ref_data[which(ref_data$group=="roche" & ref_data$label==i_lab),"var_id"]

  scatter_roche <- ggplot(data=graph_data %>%
                            arrange(participant_condition),aes_string(x=metric,y=best_company))+
    geom_point(aes(colour=factor(participant_condition)),size=2,alpha=0.5)+
    scale_color_manual(values = color_palette_scatter)+
    theme_classic()+xlab(pheno_name)+ylab(paste("Roche \n",i_lab)) +

theme(legend.position="none",axis.text.x=element_text(size=12),axis.text.y=element_text(size=12),
          axis.title=element_text(size=12))

  i_group <- "alz"
  i_lab <- top_performers[which(top_performers$group==i_group),"Biomarker"]
  best_company <- ref_data[which(ref_data$group=="alz" & ref_data$label==i_lab),"var_id"]

  scatter_alzpath  <- ggplot(data=graph_data %>%
                               arrange(participant_condition),aes_string(x=metric,y=best_company))+
    geom_point(aes(colour=factor(participant_condition)),size=2,alpha=0.5)+
    scale_color_manual(values = color_palette_scatter)+
    theme_classic()+xlab(pheno_name)+ylab(paste("ALZpath \n",i_lab)) +

theme(legend.position="none",axis.text.x=element_text(size=12),axis.text.y=element_text(size=12),
          axis.title=element_text(size=12))

  i_group <- "quan"
  i_lab <- top_performers[which(top_performers$group==i_group),"Biomarker"]
  best_company <- ref_data[which(ref_data$group=="quan" & ref_data$label==i_lab),"var_id"]

  scatter_quanterix  <- ggplot(data=graph_data %>%
                                 arrange(participant_condition),
                               aes_string(x=metric,y=best_company))+
    geom_point(aes(colour=factor(participant_condition)),size=2,alpha=0.5)+
    scale_color_manual(values = color_palette_scatter)+
    theme_classic()+xlab(pheno_name)+ylab(paste("Quanterix \n",i_lab)) +
```

```
theme(legend.position="none",axis.text.x=element_text(size=12),axis.text.y=element_text(size=12),
          axis.title=element_text(size=12))

  if(rev_axis==T){
    scatter_c2n <- scatter_c2n + scale_x_reverse()
    scatter_fuji <- scatter_fuji + scale_x_reverse()
    scatter_alzpath <- scatter_alzpath + scale_x_reverse()
    scatter_janssen <- scatter_janssen + scale_x_reverse()
    scatter_roche <- scatter_roche + scale_x_reverse()
    scatter_quanterix <- scatter_quanterix + scale_x_reverse()

  }

  order_of_plots <- top_performers[order(abs(top_performers$Rho),decreasing = T),"group"]
  plot_set_up <- list("alz"=scatter_alzpath,
                      "c2n"=scatter_c2n,
                      "fuji"=scatter_fuji,
                      "jan"=scatter_janssen,
                      "quan"=scatter_quanterix,
                      "roche"=scatter_roche)

  plot_ordered <- plot_set_up[order_of_plots]

  plot_out <- plot_grid(plot_ordered[[1]],plot_ordered[[2]],
                        plot_ordered[[3]],plot_ordered[[4]],
                        plot_ordered[[5]],plot_ordered[[6]],nrow = 3,ncol = 2,labels = "AUTO")

  return(plot_out)
}
```

**Code B5. R code for creating characteristics tables (uses function bank).**

```
#### Characteristics Table
#### Coded: Benjamin Saef
#### Date as of commenting and documentation - 05/28/2024

library(tidyr)
library(data.table)
library(RVAideMemoire)
library(pROC)
library(RColorBrewer)
library(MuMIn)
source("./Functions_for_Analysis.R")

##Set up project folders and
out_project <- "/FNIH_Project_2/Paper1" #Folder where you want all output written to, a results and
plot folder will be written here by default
out_results <- paste0(out_project,"/results/")
out_plots <- paste0(out_project,"/plots/")

dir.create(out_project,showWarnings = F)
dir.create(out_results,showWarnings = F)
dir.create(out_plots,showWarnings = F)
##Read files
data_file <- "/FNIH_Project_2/DATA_STUDY_2/cross_sectional_data_2024_04_23.csv"

last_crossectional <- read.csv(data_file)

#Fixed up race variable coding
last_crossectional <- as.data.frame(last_crossectional)
last_crossectional$RACE <- ifelse(last_crossectional$RACE==2,3,last_crossectional$RACE)
last_crossectional$RACE <- ifelse(last_crossectional$RACE==0,2,last_crossectional$RACE)

#Create Interval Variables

last_crossectional$PLASMA_CDR_INT <-
as.numeric(abs(difftime(last_crossectional$EXAMDATE,last_crossectional$VISDATE_CDR  ,units =
"days"))/365.25)
last_crossectional$PLASMA_TAU_INT <-
as.numeric(abs(difftime(last_crossectional$EXAMDATE,last_crossectional$SCANDATE_TAU,units =
"days"))/365.25)
last_crossectional$PLASMA_AMY_INT <-
as.numeric(abs(difftime(last_crossectional$EXAMDATE,last_crossectional$SCANDATE_AMY  ,units =
"days"))/365.25)
last_crossectional$PLASMA_MRI_INT <-
as.numeric(abs(difftime(last_crossectional$EXAMDATE,last_crossectional$SCANDATE_ATROPHY  ,units =
"days"))/365.25)
last_crossectional$PLASMA_CSF_INT <-
as.numeric(abs(difftime(last_crossectional$EXAMDATE,last_crossectional$EXAMDATE_CSF  ,units =
"days"))/365.25)
last_crossectional[which(is.na(last_crossectional$PTAU_over_ABETA42)),"PLASMA_CSF_INT"] <- NA

#list of variable IDs to create characteristic table entries for
varb_1 <-
c("AGE","PTGENDER","APOE_genotype","APOE4_positive","PTEDUCAT","CDR_SOB","PLASMA_CDR_INT","MMSCORE"
,"RACE",
            "ABETA42","TAU","PTAU","PTAU_over_ABETA42",#CSF
            "PLASMA_CSF_INT",

"C2N_plasma_Abeta42","C2N_plasma_Abeta40","C2N_plasma_Abeta42_Abeta40","C2N_plasma_ptau217","C2N_pl
asma_ptau217_ratio","C2N_plasma_nptau217",#C2N

"Roche_plasma_Ab42","Roche_plasma_Ab40","Roche_plasma_Ab42_Ab40","Roche_plasma_ptau181","Roche_plas
ma_GFAP","Roche_plasma_NfL",#Elecsys
```

```
"Fuji_plasma_Ab42","Fuji_plasma_Ab40","Fuji_plasma_Ab42_Ab40","Fuji_plasma_ptau217",#Lumipulse
            "AlzPath_plasma_ptau217" ,"Janssen_plasma_ptau217",#Simoa

"QX_plasma_Ab42","QX_plasma_Ab40","QX_plasma_Ab42_Ab40","QX_plasma_ptau181","QX_plasma_GFAP","QX_pl
asma_NfL", #Quanterix
            "PLASMA_AMY_INT",
            "CENTILOIDS","CENTILOIDS_10_sens",
            "PLASMA_TAU_INT",
            "MesialTemporal","TAU_MesialTemporal_10",
            "TemporoParietal","TAU_TemporoParietal_10",
            "atrophy","atrophy_10","PLASMA_MRI_INT") #Imaging Variables

#list of variable types to go in that table - please see Functions_for_Analysis.R to get more
detailed breakdown of types
type_1 <- c("A","P","4","P","A","A","A","A","R",
            "A","A","A","A",#CSF
            "A",
            "A","A","A","A","A","A",
            "A","A","A","A","A","A",
            "A","A","A","A",
            "A","A",
            "A","A","A","A","A","A",
            "A",
            "A","P",
            "A",
            "A","P",
            "A","P",
            "A","P","A")

#list of labels for the variables to go into the table
label_1 <- c("Age (Years)","Sex (% Female)","APOE4 Genotype (22/23/24/33/34/44)","APOE4 (%
Carrier)","Years of Education",
             "CDR Sum of Boxes","Plasma Collection to CDR Evaluation Time Interval","Mini-Mental
State Exam Score","Race (Black/White/Other)",
              "CSF Ab42","CSF Total Tau","CSF p-Tau 181","CSF p-Tau 181 / Ab42",#CSF
             "Time between Plasma Collection and CSF Collection",
             "Plasma Ab42","Plasma Ab40","Plasma Ab42/Ab40","Plasma p-tau217","Plasma p-tau217
Ratio","np-tau217 (pg/ml)", #C2N
             "Plasma Ab42","Plasma Ab40","Plasma Ab42/Ab40","Plasma p-Tau 181","Plasma
GFAP","Plasma Neurofilament Light",#Elecsys
             "Plasma Ab42","Plasma Ab40","Plasma Ab42/Ab40","Plasma p-tau217
Concentration",#Lumipulse
             "Plasma p-tau217 Concentration","Plasma p-tau217 Concentration",#Simoa
             "Plasma Ab42","Plasma Ab40","Plasma Ab42/Ab40","Plasma p-Tau 181","Plasma
GFAP","Plasma Neurofilament Light", #Quanterix
             "Time between Plasma Collection and Amyloid PET imaging",
             "Amyloid PET Centiloid","Amyloid PET Positivity (Centiloid > 37)",
             "Time between Plasma Collection and Tau PET imaging",
             "Tau PET Early","Tau PET Early Positivity",
             "Tau PET Late","Tau PET Late Positivity",
             "Cortical signature volume","Brain Atrophy Positivity","Time between Plasma Collection
and MRI")

####Function call - Function is in Functions_for_Analysis.R - Create full Data characteristics
tables
char_table_full_bin(last_crossectional,
                    "CENTILOIDS_10",
                    "Amyloid PET",
                    varb_1,
                    type_1,
                    label_1,

paste0(out_results,"Characteristics_Table_by_Centiloid20_positivity_05212024.xlsx"))
```

```
##Didn't make a function specifically for this, so decided to make this row separately

last_crossectional$CDR_CAT <- NA
last_crossectional[which(last_crossectional$CDR==0),"CDR_CAT"] <- "0"
last_crossectional[which(last_crossectional$CDR==0.5),"CDR_CAT"] <- "0.5"
last_crossectional[which(last_crossectional$CDR>0.5),"CDR_CAT"] <- "1+"

chisq.test(last_crossectional[,"CENTILOIDS_10"],
           as.factor(last_crossectional[,"CDR_CAT"]),
           correct=FALSE)

####
## CSF complete cases - characteristics tables
####
data_file_csf_only <- "/FNIH_Project_2/DATA_STUDY_2/cross_sectional_data_2024_04_30b_CSF_ONLY.csv"
CSF_only_data <- read.csv(data_file_csf_only)

#Creation of interval variables
CSF_only_data$PLASMA_CDR_INT <-
as.numeric(abs(difftime(CSF_only_data$EXAMDATE,CSF_only_data$VISDATE_CDR  ,units = "days"))/365.25)
CSF_only_data$PLASMA_TAU_INT <-
as.numeric(abs(difftime(CSF_only_data$EXAMDATE,CSF_only_data$SCANDATE_TAU,units = "days"))/365.25)
CSF_only_data$PLASMA_AMY_INT <-
as.numeric(abs(difftime(CSF_only_data$EXAMDATE,CSF_only_data$SCANDATE_AMY  ,units =
"days"))/365.25)
CSF_only_data$PLASMA_MRI_INT <-
as.numeric(abs(difftime(CSF_only_data$EXAMDATE,CSF_only_data$SCANDATE_ATROPHY  ,units =
"days"))/365.25)
CSF_only_data$PLASMA_CSF_INT <-
as.numeric(abs(difftime(CSF_only_data$EXAMDATE,CSF_only_data$EXAMDATE_CSF  ,units =
"days"))/365.25)

##didn't make separate function for CDR Category, so needed to make this custom to copy into table
table(CSF_only_data$CDR_CAT)
table(CSF_only_data[which(CSF_only_data$CENTILOIDS_10==0),]$CDR_CAT)
table(CSF_only_data[which(CSF_only_data$CENTILOIDS_10==1),]$CDR_CAT)
chisq.test(CSF_only_data[,"CENTILOIDS_10"],
           as.factor(CSF_only_data[,"CDR_CAT"]),
           correct=FALSE)

####Function call - Function is in Functions_for_Analysis.R
char_table_full_bin(CSF_only_data,
                    "CENTILOIDS_10",
                    "Amyloid PET",
                    varb_1,
                    type_1,
                    label_1,
                    paste0(out_results,"Characteristics_Table_by_CENT20_05162024_CSFONLY.xlsx"))

####
## Tau PET complete cases - characteristics tables
####

TAU_only_data <- subset(last_crossectional,!is.na(MesialTemporal))

table(TAU_only_data$CDR_CAT)
table(TAU_only_data[which(TAU_only_data$CENTILOIDS_10==0),]$CDR_CAT)
table(TAU_only_data[which(TAU_only_data$CENTILOIDS_10==1),]$CDR_CAT)

chisq.test(TAU_only_data[,"CENTILOIDS_10"],
           as.factor(TAU_only_data[,"CDR_CAT"]),
           correct=FALSE)

char_table_full_bin(TAU_only_data,
                    "CENTILOIDS_10",
```

```
                    "Amyloid PET",
                    varb_1,
                    type_1,
                    label_1,
                    paste0(out_results,"Characteristics_Table_by_CENT20_05212024_TAUONLY.xlsx"))

####
## Atrophy complete cases - characteristics tables
####
ATROPHY_only_data <- subset(last_crossectional,!is.na(atrophy))
ATROPHY_only_data$LP_ATROPHY_INT <-
as.numeric(abs(difftime(ATROPHY_only_data$EXAMDATE,ATROPHY_only_data$SCANDATE_ATROPHY,units =
"days"))/365.25)

table(ATROPHY_only_data$CDR_CAT)
table(ATROPHY_only_data[which(ATROPHY_only_data$CENTILOIDS_10==0),]$CDR_CAT)
table(ATROPHY_only_data[which(ATROPHY_only_data$CENTILOIDS_10==1),]$CDR_CAT)

chisq.test(ATROPHY_only_data[,"CENTILOIDS_10"],
           as.factor(ATROPHY_only_data[,"CDR_CAT"]),
           correct=FALSE)

char_table_full_bin(ATROPHY_only_data,
                    "CENTILOIDS_10",
                    "Amyloid PET",
                    varb_1,
                    type_1,
                    label_1,

paste0(out_results,"Characteristics_Table_by_CENT20_05212024_ATROPHYONLY.xlsx"))
```

**Code B6. R code for creating Spearman heatmaps (uses function bank).**

```
####Heatmap creation
#### Coded: Benjamin Saef
#### Date as of commenting and documentation - 05/28/2024

load("/FNIH_Project_2/Current_Analysis_data.rdata")
library(Hmisc)
library(reshape2)
out_project <- "/FNIH_Project_2/Paper1/Spearman_Analysis" #Folder where you want all output written
to, a results and plot folder will be written here by default
out_plots <- paste0(out_project,"/plots/heatmaps/")
dir.create(out_plots,showWarnings = F)

#C2N
pTau217_c2n <- "C2N_plasma_ptau217_ratio" #Variable representing p-tau217 ratio - c2n
pTau217conc_c2n <- "C2N_plasma_ptau217" #Variable representing p-tau217 concentration - c2n
abeta42_c2n <- "C2N_plasma_Abeta42" #Variable representing Ab42 - c2n
abeta40_c2n <- "C2N_plasma_Abeta42_Abeta40" #Variable representing Ab40 - c2n
abeta4240_c2n <- "C2N_plasma_Abeta42_Abeta40" #Variable representing Ab42/Ab40 - c2n

#Elecsys
abeta42_esys <- "Roche_plasma_Ab42" #Variable representing Ab42 - elecsys
abeta40_esys <- "Roche_plasma_Ab40" #Variable representing Ab40 - elecsys
abeta4240_esys <- "Roche_plasma_Ab42_Ab40" #Variable representing Ab42/Ab40 - elecsys
ptau_181_esys <- "Roche_plasma_ptau181" #Variable representing ptau181 - elecsys
GFAP_esys <- "Roche_plasma_GFAP" #Variable representing GFAP - elecsys
NfL_esys <- "Roche_plasma_NfL" #Variable representing NfL - elecsys

#Lumipulse

abeta42_lumi <- "Fuji_plasma_Ab42" #Variable representing Ab42 - lumipulse
abeta40_lumi <- "Fuji_plasma_Ab40" #Variable representing Ab40 - lumipulse
abeta4240_lumi <- "Fuji_plasma_Ab42_Ab40" #Variable representing Ab42/Ab40 - lumipulse
pTau217_lumi <- "Fuji_plasma_ptau217" #Variable representing p-tau217 ratio - lumipulse

ptau217_ab4240_lumi <- "" #Variable representing AB42/AB40 + p-tau217 ratio

#Simoa
alzpath_ptau217 <- "AlzPath_plasma_ptau217"  #Variable representing p-tau217 - AlzPath
janssen_ptau217 <- "Janssen_plasma_ptau217"  #Variable representing p-tau217 - Janssen

#Quanterix - Simoa
abeta42_quan <- "QX_plasma_Ab42" #Variable representing Ab42 - elecsys
abeta40_quan <- "QX_plasma_Ab40" #Variable representing Ab40 - elecsys
abeta4240_quan <- "QX_plasma_Ab42_Ab40" #Variable representing Ab42/Ab40 - elecsys
ptau_181_quan <- "QX_plasma_ptau181" #Variable representing ptau181 - elecsys
GFAP_quan <- "QX_plasma_GFAP" #Variable representing GFAP - elecsys
NfL_quan <- "QX_plasma_NfL" #Variable representing NfL - elecsys

a_var <- c( "ab4240", #Ab42/40
            "pt217", #ptau217
            "pt217", #pt217
            "ab4240", #Ab42/40
            "pt181", #ptau181
            "gfap", #GFAP
            "nfl", #NFL
            "ab4240",#Ab42/40
            "pt217",#ptau217
            "pt217",#ptau217
            "pt217",#ptau217
            "ab4240", #Ab42/40
            "pt181", #ptau181
            "gfap", #GFAP
```

```
            "nfl"#NFL
)

l_var <- c( "c2n", #Ab42/40
            "c2n", #ptau217
            "c2n",
            "roche", #Ab42/40
            "roche", #ptau181
            "roche", #GFAP
            "roche", #NFL
            "fuji",#Ab42/40
            "fuji",#ptau217
            "alz",#ptau217
            "jan",#ptau217
            "quan", #Ab42/40
            "quan", #ptau181
            "quan", #GFAP
            "quan"#NFL
)

##list out the different analytes - Labels
analytes <- c(paste0("A","\U03B2","42/A","\U03B2","40"), #Ab42/40
              "p-tau217 ratio (%)", #ptau217
              "p-tau217 (pg/ml)", #ptau217
              paste0("A","\U03B2","42/A","\U03B2","40"), #Ab42/40
              "p-tau181 (pg/ml)", #ptau181
              "GFAP (ng/ml)", #GFAP
              "NfL (pg/mL)", #NFL
              paste0("A","\U03B2","42/A","\U03B2","40"), #Ab42/40
              "p-tau217 (pg/ml)", #ptau217
              "p-tau217 (pg/ml)", #ptau217
              "p-tau217 (pg/ml)", #ptau217
              paste0("A","\U03B2","42/A","\U03B2","40"), #Ab42/40
              "p-tau181 (pg/ml)", #ptau181
              "GFAP (pg/ml)", #GFAP
              "NfL (pg/mL)" #NFL
)
#list out the variables representing those analytes.
i_var <- c(abeta4240_c2n, #Ab42/40 - c2n
           pTau217_c2n, #ptau217 - c2n
           pTau217conc_c2n, #ptau217 - c2n
           abeta4240_esys,#Ab42/40 - elecsys
           ptau_181_esys,#ptau181 - elecsys
           GFAP_esys,#GFAP - elecsys
           NfL_esys, #NFL - elecsys
           abeta4240_lumi,#Ab42/40 - lumipulse
           pTau217_lumi, #ptau217 - lumipulse
           alzpath_ptau217,#ptau217 - alzpath
           janssen_ptau217,#ptau217 - lumipulse
           abeta4240_quan,#Ab42/40 - quanterix
           ptau_181_quan,#ptau181 - quanterix
           GFAP_quan,#GFAP - quanterix
           NfL_quan #NFL - quanterix
)

ref_data <- data.frame(group=l_var,var_id=i_var,label=analytes,analyte_code=a_var)
ref_data$company <- recode(ref_data$group,"quan"="Quanterix","alz"="ALZpath","c2n"="C2N",
                           "fuji"="Fujirebio", "jan"="Janssen","roche" = "Roche")
ref_data$fullname <- paste(ref_data$company,ref_data$label)

#  a.  Correlation matrix A (matrix with colors and numbers of unadjusted Spearman correlations)
#i.    Entire cohort
#ii.   Color is based on absolute value (red higher, blue lower)
```

```
#iii.  All plasma measures versus one another

order_of_analytes <- rev(c("pt217","pt181","gfap","nfl","ab4240"))

####Creation of heatmap - analyte groups ordered by max absolute rho value
full_heatmap <- create_cor_heatmap_for_project(data.in,ref_data = ref_data)
####Creation of heatmap - analyte groups ordered by custom preference
full_heatmap_reorder <- create_cor_heatmap_for_project_custom(data.in,ref_data = ref_data,reforder
= order_of_analytes)

ggsave(paste0(out_plots,"heatmap_full_reorder.png"),
       plot=full_heatmap_reorder,
       device=png,dpi = 1200,width = 6,height=6)

#b.    Correlation matrix B (matrix with colors and numbers of unadjusted Spearman correlations)
#i.    Amyloid PET positive cohort
#ii.   Color is based on absolute value (red higher, blue lower)
#iii.  All plasma measures versus one another
####Creation of heatmap - analyte groups ordered by max absolute rho value
amyloid_positive_heatmap <-
create_cor_heatmap_for_project(subset(data.in,CENTILOIDS_10==1),ref_data = ref_data)
ggsave(paste0(out_plots,"heatmap_amyloid_pos.jpg"),
       plot=amyloid_positive_heatmap,
       device=jpeg,width = 6,height=6)

####Creation of heatmap - analyte groups ordered by custom preference
amyloid_positive_heatmap_reorder <-
create_cor_heatmap_for_project_custom(subset(data.in,CENTILOIDS_10==1),ref_data = ref_data,reforder
= order_of_analytes)

ggsave(paste0(out_plots,"heatmap_amypos_reorder.png"),
       plot=amyloid_positive_heatmap_reorder,
       device=png,dpi = 1200,width = 6,height=6)

#c.    Correlation matrix C (matrix with colors and numbers of unadjusted Spearman correlations)
#i.    Cognitively impaired cohort
#ii.   Color is based on absolute value (red higher, blue lower)
#iii.  All plasma measures versus one another
####Creation of heatmap - analyte groups ordered by max absolute rho value
cdr_positive_heatmap <- create_cor_heatmap_for_project(subset(data.in,CDR_10==1),ref_data =
ref_data)
ggsave(paste0(out_plots,"heatmap_cdr_pos.jpg"),
       plot=cdr_positive_heatmap,
       device=jpeg,width = 6,height=6)

####Creation of heatmap - analyte groups ordered by custom preference
cdr_positive_heatmap_reorder <-
create_cor_heatmap_for_project_custom(subset(data.in,CDR_10==1),ref_data = ref_data,reforder =
order_of_analytes)

ggsave(paste0(out_plots,"heatmap_cdr_pos_reorder.png"),
       plot=cdr_positive_heatmap_reorder,
       device=png,dpi = 1200,width = 6,height=6)

#d.    Correlation matrix D (matrix with colors and numbers of unadjusted Spearman correlations)
#i.    Cognitively unimpaired cohort
#ii.   Color is based on absolute value (red higher, blue lower)
#iii.  All plasma measures versus one another
####Creation of heatmap - analyte groups ordered by max absolute rho value
cdr_negative_heatmap <- create_cor_heatmap_for_project(subset(data.in,CDR_10==0),ref_data =
ref_data)
ggsave(paste0(out_plots,"heatmap_cdr_neg.jpg"),
       plot=cdr_negative_heatmap,
```

```
       device=jpeg,width = 6,height=6)

####Creation of heatmap - analyte groups ordered by custom preference
cdr_negative_heatmap_reorder <-
create_cor_heatmap_for_project_custom(subset(data.in,CDR_10==0),ref_data = ref_data,reforder =
order_of_analytes)

ggsave(paste0(out_plots,"heatmap_cdr_neg_reorder.png"),
       plot=cdr_negative_heatmap_reorder,
       device=png,dpi = 1200,width = 6,height=6)
```

**Code B7. R code for Spearman analysis (uses function bank).**

```
#####Spearman Analyses
#### Coded: Benjamin Saef
#### Date as of commenting and documentation - 05/28/2024

library(tidyr)
library(data.table)
library(RVAideMemoire)
library(pROC)
library(RColorBrewer)
library(MuMIn)
source("./Functions_for_Analysis.R")

#NO YEARS OF EDUCATION
#Make partial comparisons to top comparing partial
#Do algorithmic check on top performer before setting reference - new function?
#Remove all Quanterix only individuals

####Set seed - bootstrapping is involved so make sure
set.seed(12345)

#keep this script in the same folder as the "Functions_for_Analysis.R"
out_project <- "/FNIH_Project_2/Paper1/Spearman_Analysis" #Folder where you want all output written
to, a results and plot folder will be written here by default
out_results <- paste0(out_project,"/results/")
out_plots <- paste0(out_project,"/plots/")

dir.create(out_project,showWarnings = F)
dir.create(out_results,showWarnings = F)
dir.create(out_plots,showWarnings = F)

data_file <- "/FNIH_Project_2/DATA_STUDY_2/cross_sectional_data_2024_04_24b.csv"  # cross sectional
dataset

###read in dataset
data.in <- read.csv(data_file)

####This was done so I could verify the analysis by running analysis code in a different
programming language
####write.csv(data.in,"/FNIH_Project_2/DATA_STUDY_2/cross_sectional_data_2024_04_24b_FOR_SAS.csv",n
a = ".",row.names = F)
sex_var <- "PTGENDER" #variable with sex
age_var <- "AGE" #variable with age
APOE_variable <- "APOE_genotype" #Variable with APOE genotypes

#analysis vars
#C2N
pTau217_c2n <- "C2N_plasma_ptau217_ratio" #Variable representing p-tau217 ratio - c2n
pTau217conc_c2n <- "C2N_plasma_ptau217" #Variable representing p-tau217 concentration - c2n
abeta42_c2n <- "C2N_plasma_Abeta42" #Variable representing Ab42 - c2n
abeta40_c2n <- "C2N_plasma_Abeta42_Abeta40" #Variable representing Ab40 - c2n
abeta4240_c2n <- "C2N_plasma_Abeta42_Abeta40" #Variable representing Ab42/Ab40 - c2n

#Elecsys
abeta42_esys <- "Roche_plasma_Ab42" #Variable representing Ab42 - elecsys
abeta40_esys <- "Roche_plasma_Ab40" #Variable representing Ab40 - elecsys
abeta4240_esys <- "Roche_plasma_Ab42_Ab40" #Variable representing Ab42/Ab40 - elecsys
ptau_181_esys <- "Roche_plasma_ptau181" #Variable representing ptau181 - elecsys
GFAP_esys <- "Roche_plasma_GFAP" #Variable representing GFAP - elecsys
NfL_esys <- "Roche_plasma_NfL" #Variable representing NfL - elecsys

#Lumipulse
```

```
abeta42_lumi <- "Fuji_plasma_Ab42" #Variable representing Ab42 - lumipulse
abeta40_lumi <- "Fuji_plasma_Ab40" #Variable representing Ab40 - lumipulse
abeta4240_lumi <- "Fuji_plasma_Ab42_Ab40" #Variable representing Ab42/Ab40 - lumipulse
pTau217_lumi <- "Fuji_plasma_ptau217" #Variable representing p-tau217 ratio - lumipulse

#Simoa
alzpath_ptau217 <- "AlzPath_plasma_ptau217"  #Variable representing p-tau217 - AlzPath
janssen_ptau217 <- "Janssen_plasma_ptau217"  #Variable representing p-tau217 - Janssen

#Quanterix - Simoa
abeta42_quan <- "QX_plasma_Ab42" #Variable representing Ab42 - elecsys
abeta40_quan <- "QX_plasma_Ab40" #Variable representing Ab40 - elecsys
abeta4240_quan <- "QX_plasma_Ab42_Ab40" #Variable representing Ab42/Ab40 - elecsys
ptau_181_quan <- "QX_plasma_ptau181" #Variable representing ptau181 - elecsys
GFAP_quan <- "QX_plasma_GFAP" #Variable representing GFAP - elecsys
NfL_quan <- "QX_plasma_NfL" #Variable representing NfL - elecsys

#Variabels for PET metrics
centiloid_Variable <- "CENTILOIDS" #Variable representing Centiloid
early_tau <- "MesialTemporal" #Variable representing Tau PET Centaur
late_tau <- "TemporoParietal"
atrophy <- "atrophy"
cdr_sum_of_boxes <- "CDR_SOB"
MMSE <- "MMSCORE"

#Making sure that the allele recordings fit neatly into the APOE levels we have selected
data.in[which(data.in[,APOE_variable]=="43"),APOE_variable] <- "34"
data.in[which(data.in[,APOE_variable]=="42"),APOE_variable] <- "24"

###Make sure APOE 33 is the reference level
data.in$APOE_Cat <- factor(data.in[,APOE_variable],
                              levels=c("33","34","24","22","23","44"))

#Covariates
c_var <- c(age_var, #Age
           sex_var, #Sex
           "APOE_Cat") #APOE category variable

#####Analyses groups per platform
#Groupings if necessary - these are repeated for the purpose of comparisons within group keeping measures from same company together
l_var <- c( "c2n", #Ab42/40
            "c2n", #ptau217
            "c2n",
            "roche", #Ab42/40
            "roche", #ptau181
            "roche", #GFAP
            "roche", #NFL
            "fuji",#Ab42/40
            "fuji",#ptau217
            "alz",#ptau217
            "jan",#ptau217
            "quan", #Ab42/40
            "quan", #ptau181
            "quan", #GFAP
            "quan"#NFL
)

##list out the different analytes - Labels
analytes <- c(paste0("A","\U03B2","42/A","\U03B2","40"), #Ab42/40
              "p-tau217 ratio (%)", #ptau217
              "p-tau217 (pg/ml)", #ptau217
              paste0("A","\U03B2","42/A","\U03B2","40"), #Ab42/40
              "p-tau181 (pg/ml)", #ptau181
              "GFAP (ng/ml)", #GFAP
```

```
              "NfL (pg/mL)", #NFL
              paste0("A","\U03B2","42/A","\U03B2","40"), #Ab42/40
              "p-tau217 (pg/ml)", #ptau217
              "p-tau217 (pg/ml)", #ptau217
              "p-tau217 (pg/ml)", #ptau217
              paste0("A","\U03B2","42/A","\U03B2","40"), #Ab42/40
              "p-tau181 (pg/ml)", #ptau181
              "GFAP (pg/ml)", #GFAP
              "NfL (pg/mL)" #NFL
)
#list out the variables representing those analytes.
i_var <- c(abeta4240_c2n, #Ab42/40 - c2n
           pTau217_c2n, #ptau217 - c2n
           pTau217conc_c2n, #ptau217 - c2n
           abeta4240_esys,#Ab42/40 - elecsys
           ptau_181_esys,#ptau181 - elecsys
           GFAP_esys,#GFAP - elecsys
           NfL_esys, #NFL - elecsys
           abeta4240_lumi,#Ab42/40 - lumipulse
           pTau217_lumi, #ptau217 - lumipulse
           alzpath_ptau217,#ptau217 - alzpath
           janssen_ptau217,#ptau217 - lumipulse
           abeta4240_quan,#Ab42/40 - quanterix
           ptau_181_quan,#ptau181 - quanterix
           GFAP_quan,#GFAP - quanterix
           NfL_quan #NFL - quanterix
)

###output variable vector creation
o_vars <- c(centiloid_Variable, # Centiloid
            early_tau,#Early Tau
            late_tau, #Late Tau
            atrophy, #Atrophy
            cdr_sum_of_boxes, #CDR sum of boxes
            MMSE) #MMSE (not actually used)

spdiff_Partial_rho <- function(data, indices) { ###sort of a double check on the function creation
for the bootstratpping comparisons
  d <- data[indices,] # allows boot to select sample
  c_cov <- c(age_var,sex_var,"APOE_Cat")   #change to reflect data
  cor_xz <- RVAideMemoire::pcor.test(d$x, d$z,d[,c_cov], method = "spearman",nrep = 1)
  cor_yz <- RVAideMemoire::pcor.test(d$y, d$z, d[,c_cov],method = "spearman",nrep = 1)
  diff <- abs(cor_xz$estimate) - abs(cor_yz$estimate)

  return(diff)
}
###the above function also exists in Functions_for_Analysis.R

#####Centiloid

###Full Cohort

###Determine the comparison standard before running
comparison_standard <- Eval_comp_standard(data.in,o_vars[1],i_var)
comparison_standard_partial <- Eval_comp_standard_partial(data.in,o_vars[1],c_var,i_var)
ptm <- proc.time()
table_out_centiloid_full <-  spearman_plasma_runs(data.in, # Data inputted
                       o_vars[1],  # variable A - continuous variable being correlated
                       i_var,# vector of strings - continuous variables being correlated with
variable A
                       comparison_standard,
                       comparison_standard_partial, #variable in vars_to_cor vector that is used
as standard for comparisons
                       c_var,    # covariates - variables used as covariates (age, sex, APOE
normally)
```

```
                        analytes,
                        l_var)

#####Centiloid 20 - positive
comparison_standard <- Eval_comp_standard(subset(data.in,CENTILOIDS_10==1),o_vars[1],i_var)
comparison_standard_partial <-
Eval_comp_standard_partial(subset(data.in,CENTILOIDS_10==1),o_vars[1],c_var,i_var)
table_out_centiloid_amy20pos <-  spearman_plasma_runs(subset(data.in,CENTILOIDS_10==1), # Data
inputted
                                                      o_vars[1],  # variable A - continuous
variable being correlated
                                                      i_var,# vector of strings - continuous
variables being correlated with variable A
                                                      comparison_standard,
                                                      comparison_standard_partial, #variable in
vars_to_cor vector that is used as standard for comparisons
                                                      c_var,    # covariates - variables used as
covariates (age, sex, APOE normally)
                                                      analytes,
                                                      l_var)

####Centiloid 20 - negative
comparison_standard <- Eval_comp_standard(subset(data.in,CENTILOIDS_10==0),o_vars[1],i_var)
comparison_standard_partial <-
Eval_comp_standard_partial(subset(data.in,CENTILOIDS_10==0),o_vars[1],c_var,i_var)
ptm <- proc.time()
table_out_centiloid_amy20neg <-  spearman_plasma_runs(subset(data.in,CENTILOIDS_10==0), # Data
inputted
                                                      o_vars[1],  # variable A - continuous
variable being correlated
                                                      i_var,# vector of strings - continuous
variables being correlated with variable A
                                                      comparison_standard,
                                                      comparison_standard_partial, #variable in
vars_to_cor vector that is used as standard for comparisons
                                                      c_var,    # covariates - variables used as
covariates (age, sex, APOE normally)
                                                      analytes,
                                                      l_var)
proc.time() - ptm

####Centiloid 37 - positive
comparison_standard <- Eval_comp_standard(subset(data.in,CENTILOIDS_10_sens==1),o_vars[1],i_var)
comparison_standard_partial <-
Eval_comp_standard_partial(subset(data.in,CENTILOIDS_10_sens==1),o_vars[1],c_var,i_var)
table_out_centiloid_amy37pos <-  spearman_plasma_runs(subset(data.in,CENTILOIDS_10_sens==1), # Data
inputted
                                                      o_vars[1],  # variable A - continuous
variable being correlated
                                                      i_var,# vector of strings - continuous
variables being correlated with variable A
                                                      comparison_standard,
                                                      comparison_standard_partial, #variable in
vars_to_cor vector that is used as standard for comparisons
                                                      c_var,    # covariates - variables used as
covariates (age, sex, APOE normally)
                                                      analytes,
                                                      l_var)

####Centiloid 37 negative
comparison_standard <- Eval_comp_standard(subset(data.in,CENTILOIDS_10_sens==0),o_vars[1],i_var)
comparison_standard_partial <-
Eval_comp_standard_partial(subset(data.in,CENTILOIDS_10_sens==0),o_vars[1],c_var,i_var)
table_out_centiloid_amy37neg <-  spearman_plasma_runs(subset(data.in,CENTILOIDS_10_sens==0), # Data
inputted
```

```
                                                      o_vars[1],  # variable A - continuous
variable being correlated
                                                      i_var,# vector of strings - continuous
variables being correlated with variable A
                                                      comparison_standard,
                                                      comparison_standard_partial, #variable in
vars_to_cor vector that is used as standard for comparisons
                                                      c_var,    # covariates - variables used as
covariates (age, sex, APOE normally)
                                                      analytes,
                                                      l_var)

####CDR - positive
comparison_standard <- Eval_comp_standard(subset(data.in,CDR_10==1),o_vars[1],i_var)
comparison_standard_partial <-
Eval_comp_standard_partial(subset(data.in,CDR_10==1),o_vars[1],c_var,i_var)
table_out_centiloid_cdrpos <-  spearman_plasma_runs(subset(data.in,CDR_10==1), # Data inputted
                                                   o_vars[1],  # variable A - continuous variable
being correlated
                                                   i_var,# vector of strings - continuous variables
being correlated with variable A
                                                   comparison_standard,
                                                   comparison_standard_partial, #variable in
vars_to_cor vector that is used as standard for comparisons
                                                   c_var,    # covariates - variables used as
covariates (age, sex, APOE normally)
                                                   analytes,
                                                   l_var)

####CDR - negative
comparison_standard <- Eval_comp_standard(subset(data.in,CDR_10==0),o_vars[1],i_var)
comparison_standard_partial <-
Eval_comp_standard_partial(subset(data.in,CDR_10==0),o_vars[1],c_var,i_var)
table_out_centiloid_cdrneg <-  spearman_plasma_runs(subset(data.in,CDR_10==0), # Data inputted
                                                    o_vars[1],  # variable A - continuous variable
being correlated
                                                    i_var,# vector of strings - continuous
variables being correlated with variable A
                                                    comparison_standard,
                                                    comparison_standard_partial, #variable in
vars_to_cor vector that is used as standard for comparisons
                                                    c_var,    # covariates - variables used as
covariates (age, sex, APOE normally)
                                                    analytes,
                                                    l_var)

##### This function is only in this code, it loses use once the above 'analytes' vector is
corrected.
##### putting it here for documentation's purpose
relabel_spearman_res <- function(DF){

  DF$Biomarker <-recode(DF$Biomarker, "p-Tau 217 Ratio" = "p-tau217 ratio (%)",
                      "p-Tau 217 concentration" = "p-tau217 (pg/ml)",
                      "Neurofilament Light"="NfL (pg/mL)",
                      "GFAP"="GFAP (pg/ml)",
                      "p-Tau 181"="p-tau181 (pg/ml)")

  DF[which(DF$group=="roche" & DF$Biomarker=="GFAP (pg/ml)"),"Biomarker"] <- "GFAP (ng/ml)"
  DF[which(DF$Biomarker==paste0("Plasma A","\U03B2","42/A","\U03B2","40")),"Biomarker"] <-
paste0("A","\U03B2","42/A","\U03B2","40")

  DF$company <- factor(recode(DF$group,"quan"="Quanterix","alz"="ALZpath","c2n"="C2N",
                            "fuji"="Fujirebio", "jan"="Janssen","roche" = "Roche"))
  levels(DF$company) <- c("ALZpath","C2N", "Fujirebio",  "Janssen", "Quanterix","Roche")
```

```
  DF$company <-factor(DF$company, levels = c("C2N", "Fujirebio", "ALZpath", "Janssen", "Roche", 
"Quanterix"))

return(DF)

}

#####compile all the results data into a single list to save out and use in further output
list_of_cent_correlation_dataset <- list("Full Data"=table_out_centiloid_full,
                                         "Cent20 Positive Data"=table_out_centiloid_amy20pos,
                                         "Cent20 Negative Data"=table_out_centiloid_amy20neg,
                                         "Cent37 Positive Data"=table_out_centiloid_amy37pos,
                                         "Cent37 Negative Data"=table_out_centiloid_amy37neg,
                                         "CDR Positive Data"=table_out_centiloid_cdrpos,
                                         "CDR Negative Data"=table_out_centiloid_cdrneg)
####Due to length of table creation - save output data upon finishing
save(list_of_cent_correlation_dataset,
     file=paste0(out_results,"cent_correlations.rdata"))

########## deprecated - no longer necessary
#
#for(i in 1:length(list_of_cent_correlation_dataset)){
#
#
#  list_cent_new[[i]] <-  relabel_spearman_res(list_of_cent_correlation_dataset[[i]])
#
#}
#names(list_cent_new) <- names(list_of_cent_correlation_dataset)
##########
###Create Clean Plots
list_cent_new <- list()
list_of_cent_clean <- list()
for(i_clean in 1: length(list_of_cent_correlation_dataset)){
  DF <- list_of_cent_correlation_dataset[[i_clean]]

  DF$company <- factor(recode(DF$group,"quan"="Quanterix","alz"="ALZpath","c2n"="C2N",
                              "fuji"="Fujirebio", "jan"="Janssen","roche" = "Roche")) ### rename 
group to company, set as factor
  levels(DF$company) <- c("ALZpath","C2N", "Fujirebio",  "Janssen", "Quanterix","Roche") #set 
factor levels properly
  DF$company <-factor(DF$company, levels = c("C2N", "Fujirebio", "ALZpath", "Janssen", "Roche", 
"Quanterix")) # order factor properly

  list_cent_new[[i_clean]] <- DF ###add dataset with proper variables to go into plot 
creation/cleaning to new list
  list_of_cent_clean[[i_clean]]  <- clean_spearman_table(DF)
}

names(list_of_cent_clean) <- names(list_of_cent_correlation_dataset)

###Write out publication format tables
write.xlsx(list_of_cent_clean,
           file=paste0(out_results,"Centiloid_Correlations_05132024.xlsx"))

#####Tau Early

###Full cohort

###determine comparison standard for cohort, then run
comparison_standard <- Eval_comp_standard(data.in,o_vars[2],i_var)
comparison_standard_partial <- Eval_comp_standard_partial(data.in,o_vars[2],c_var,i_var)
ptm <- proc.time()
table_out_tauearly_full <-  spearman_plasma_runs(data.in, # Data inputted
```

```
                                                  o_vars[2],  # variable A - continuous variable
being correlated
                                                  i_var,# vector of strings - continuous variables
being correlated with variable A
                                                  comparison_standard,
                                                 comparison_standard_partial,#variable in
vars_to_cor vector that is used as standard for comparisons
                                                  c_var,    # covariates - variables used as
covariates (age, sex, APOE normally)
                                                  analytes,
                                                  l_var)
proc.time() - ptm

#### Centiloid 20 positive
ptm <- proc.time()
comparison_standard <- Eval_comp_standard(subset(data.in,CENTILOIDS_10==1),o_vars[2],i_var)
comparison_standard_partial <-
Eval_comp_standard_partial(subset(data.in,CENTILOIDS_10==1),o_vars[2],c_var,i_var)
table_out_tauearly_amy20pos <-  spearman_plasma_runs(subset(data.in,CENTILOIDS_10==1), # Data
inputted
                                                      o_vars[2],  # variable A - continuous
variable being correlated
                                                      i_var,# vector of strings - continuous
variables being correlated with variable A
                                                     comparison_standard,
                                                     comparison_standard_partial,#variable in
vars_to_cor vector that is used as standard for comparisons
                                                     c_var,    # covariates - variables used as
covariates (age, sex, APOE normally)
                                                      analytes,
                                                      l_var)

proc.time() - ptm

#### Centiloid 20 negative
ptm <- proc.time()
comparison_standard <- Eval_comp_standard(subset(data.in,CENTILOIDS_10==0),o_vars[2],i_var)
comparison_standard_partial <-
Eval_comp_standard_partial(subset(data.in,CENTILOIDS_10==0),o_vars[2],c_var,i_var)
table_out_tauearly_amy20neg <-  spearman_plasma_runs(subset(data.in,CENTILOIDS_10==0), # Data
inputted
                                                      o_vars[2],  # variable A - continuous
variable being correlated
                                                      i_var,# vector of strings - continuous
variables being correlated with variable A
                                                     comparison_standard,
                                                     comparison_standard_partial,#variable in
vars_to_cor vector that is used as standard for comparisons
                                                     c_var,    # covariates - variables used as
covariates (age, sex, APOE normally)
                                                      analytes,
                                                      l_var)

proc.time() - ptm

#### Centiloid 37 positive
ptm <- proc.time()
comparison_standard <- Eval_comp_standard(subset(data.in,CENTILOIDS_10_sens==1),o_vars[2],i_var)
comparison_standard_partial <-
Eval_comp_standard_partial(subset(data.in,CENTILOIDS_10_sens==1),o_vars[2],c_var,i_var)
table_out_tauearly_amy37pos <-  spearman_plasma_runs(subset(data.in,CENTILOIDS_10_sens==1), # Data
inputted
                                                      o_vars[2],  # variable A - continuous
variable being correlated
                                                      i_var,# vector of strings - continuous
variables being correlated with variable A
                                                     comparison_standard,
```

```
                                                     comparison_standard_partial,#variable in
vars_to_cor vector that is used as standard for comparisons
                                                     c_var,    # covariates - variables used as
covariates (age, sex, APOE normally)
                                                      analytes,
                                                      l_var)
proc.time() - ptm

#### Centiloid 37 negative

ptm <- proc.time()
comparison_standard <- Eval_comp_standard(subset(data.in,CENTILOIDS_10_sens==0),o_vars[2],i_var)
comparison_standard_partial <-
Eval_comp_standard_partial(subset(data.in,CENTILOIDS_10_sens==0),o_vars[2],c_var,i_var)
table_out_tauearly_amy37neg <-  spearman_plasma_runs(subset(data.in,CENTILOIDS_10_sens==0), # Data
inputted
                                                      o_vars[2],  # variable A - continuous
variable being correlated
                                                      i_var,# vector of strings - continuous
variables being correlated with variable A
                                                     comparison_standard,
                                                     comparison_standard_partial,#variable in
vars_to_cor vector that is used as standard for comparisons
                                                     c_var,    # covariates - variables used as
covariates (age, sex, APOE normally)
                                                      analytes,
                                                      l_var)

proc.time() - ptm

#### CDR positive
ptm <- proc.time()
comparison_standard <- Eval_comp_standard(subset(data.in,CDR_10==1),o_vars[2],i_var)
comparison_standard_partial <-
Eval_comp_standard_partial(subset(data.in,CDR_10==1),o_vars[2],c_var,i_var)
table_out_tauearly_cdrpos <-  spearman_plasma_runs(subset(data.in,CDR_10==1), # Data inputted
                                                    o_vars[2],  # variable A - continuous variable
being correlated
                                                    i_var,# vector of strings - continuous
variables being correlated with variable A
                                                   comparison_standard,
                                                   comparison_standard_partial,#variable in
vars_to_cor vector that is used as standard for comparisons
                                                   c_var,    # covariates - variables used as
covariates (age, sex, APOE normally)
                                                    analytes,
                                                    l_var)
proc.time() - ptm

#### CDR negative
ptm <- proc.time()
comparison_standard <- Eval_comp_standard(subset(data.in,CDR_10==0),o_vars[2],i_var)
comparison_standard_partial <-
Eval_comp_standard_partial(subset(data.in,CDR_10==0),o_vars[2],c_var,i_var)
table_out_tauearly_cdrneg <-  spearman_plasma_runs(subset(data.in,CDR_10==0), # Data inputted
                                                    o_vars[2],  # variable A - continuous variable
being correlated
                                                    i_var,# vector of strings - continuous
variables being correlated with variable A
                                                   comparison_standard,
                                                   comparison_standard_partial,#variable in
vars_to_cor vector that is used as standard for comparisons
                                                   c_var,    # covariates - variables used as
covariates (age, sex, APOE normally)
                                                    analytes,
                                                    l_var)
proc.time() - ptm
```

```
####Add all results tables to list
list_of_tauearly_correlation_dataset <- list("Full Data"=table_out_tauearly_full,
                                        "Cent20 Positive Data"=table_out_tauearly_amy20pos,
                                        "Cent20 Negative Data"=table_out_tauearly_amy20neg,
                                        "Cent37 Positive Data"=table_out_tauearly_amy37pos,
                                        "Cent37 Negative Data"=table_out_tauearly_amy37neg,
                                        "CDR Positive Data"=table_out_tauearly_cdrpos,
                                        "CDR Negative Data"=table_out_tauearly_cdrneg)
####Due to length of table creation - save output data upon finishing
save(list_of_tauearly_correlation_dataset,
     file=paste0(out_results,"tauearly_correlations.rdata"))

####Deprecated - no longer useful after fixing analytes vector
#list_tauearly_new <- list()
#for(i in 1:length(list_of_tauearly_correlation_dataset)){
#
#
#  list_tauearly_new[[i]] <-  relabel_spearman_res(list_of_tauearly_correlation_dataset[[i]])
#
#}
#
#names(list_tauearly_new) <- names(list_of_tauearly_correlation_dataset)

write.xlsx(list_of_tauearly_correlation_dataset,
           file=paste0(out_results,"TauEarly_Correlations.xlsx"))

###Create Clean Plots
list_tauearly_new <- list()
list_of_tauearly_clean <- list()
for(i_clean in 1: length(list_of_tauearly_correlation_dataset)){
  DF <- list_of_tauearly_correlation_dataset[[i_clean]]

  DF$company <- factor(recode(DF$group,"quan"="Quanterix","alz"="ALZpath","c2n"="C2N",
                              "fuji"="Fujirebio", "jan"="Janssen","roche" = "Roche")) ### rename
group to company, set as factor
  levels(DF$company) <- c("ALZpath","C2N", "Fujirebio",  "Janssen", "Quanterix","Roche") #set
factor levels properly
  DF$company <-factor(DF$company, levels = c("C2N", "Fujirebio", "ALZpath", "Janssen", "Roche",
"Quanterix")) # order factor properly

  list_tauearly_new[[i_clean]] <- DF ###add dataset with proper variables to go into plot
creation/cleaning to new list
  list_of_tauearly_clean[[i_clean]]  <- clean_spearman_table(DF)
}

names(list_of_tauearly_clean) <- names(list_of_tauearly_correlation_dataset)

#write out publication format tables
write.xlsx(list_of_tauearly_clean,
           file=paste0(out_results,"TauEarly_Correlations_05112024.xlsx"))

#####

####TAU LATE

####Full Data
#### Determine comparison standards for each cohort then run
comparison_standard <- Eval_comp_standard(data.in,o_vars[3],i_var)
comparison_standard_partial <- Eval_comp_standard_partial(data.in,o_vars[3],c_var,i_var)

ptm <- proc.time()
table_out_TauLate_full <-  spearman_plasma_runs(data.in, # Data inputted
                                               o_vars[3],  # variable A - continuous variable
being correlated
```

```
                                                 i_var,# vector of strings - continuous variables
being correlated with variable A
                                                 comparison_standard,
                                                comparison_standard_partial,#variable in
vars_to_cor vector that is used as standard for comparisons
                                                 c_var,    # covariates - variables used as
covariates (age, sex, APOE normally)
                                                 analytes,
                                                 l_var)
proc.time() - ptm

##### Amyloid 20 positive
ptm <- proc.time()
comparison_standard <- Eval_comp_standard(subset(data.in,CENTILOIDS_10==1),o_vars[3],i_var)
comparison_standard_partial <-
Eval_comp_standard_partial(subset(data.in,CENTILOIDS_10==1),o_vars[3],c_var,i_var)

table_out_TauLate_amy20pos <-  spearman_plasma_runs(subset(data.in,CENTILOIDS_10==1), # Data
inputted
                                                     o_vars[3],  # variable A - continuous variable
being correlated
                                                     i_var,# vector of strings - continuous
variables being correlated with variable A
                                                    comparison_standard,
                                                    comparison_standard_partial, #variable in
vars_to_cor vector that is used as standard for comparisons
                                                     c_var,    # covariates - variables used as
covariates (age, sex, APOE normally)
                                                     analytes,
                                                     l_var)

proc.time() - ptm

##### Amyloid 20 negative
ptm <- proc.time()
comparison_standard <- Eval_comp_standard(subset(data.in,CENTILOIDS_10==0),o_vars[3],i_var)
comparison_standard_partial <-
Eval_comp_standard_partial(subset(data.in,CENTILOIDS_10==0),o_vars[3],c_var,i_var)

table_out_TauLate_amy20neg <-  spearman_plasma_runs(subset(data.in,CENTILOIDS_10==0), # Data
inputted
                                                     o_vars[3],  # variable A - continuous variable
being correlated
                                                     i_var,# vector of strings - continuous
variables being correlated with variable A
                                                    comparison_standard,
                                                    var_comp_p = comparison_standard_partial,
                                                     c_var,    # covariates - variables used as
covariates (age, sex, APOE normally)
                                                     analytes,
                                                     l_var)
proc.time() - ptm

##### Amyloid 37 positive
ptm <- proc.time()
comparison_standard <- Eval_comp_standard(subset(data.in,CENTILOIDS_10_sens==1),o_vars[3],i_var)
comparison_standard_partial <-
Eval_comp_standard_partial(subset(data.in,CENTILOIDS_10_sens==1),o_vars[3],c_var,i_var)

table_out_TauLate_amy37pos <-  spearman_plasma_runs(subset(data.in,CENTILOIDS_10_sens==1), # Data
inputted
                                                     o_vars[3],  # variable A - continuous variable
being correlated
                                                     i_var,# vector of strings - continuous
variables being correlated with variable A
```

```
                                                    comparison_standard,
                                                    comparison_standard_partial, #variable in
vars_to_cor vector that is used as standard for comparisons
                                                     c_var,    # covariates - variables used as
covariates (age, sex, APOE normally)
                                                     analytes,
                                                     l_var)
proc.time() - ptm

##### Amyloid 37 negative
ptm <- proc.time()
comparison_standard <- Eval_comp_standard(subset(data.in,CENTILOIDS_10_sens==0),o_vars[3],i_var)
comparison_standard_partial <-
Eval_comp_standard_partial(subset(data.in,CENTILOIDS_10_sens==0),o_vars[3],c_var,i_var)

table_out_TauLate_amy37neg <-  spearman_plasma_runs(subset(data.in,CENTILOIDS_10_sens==0), # Data
inputted
                                                     o_vars[3],  # variable A - continuous variable
being correlated
                                                     i_var,# vector of strings - continuous
variables being correlated with variable A
                                                    comparison_standard,
                                                    comparison_standard_partial, #variable in
vars_to_cor vector that is used as standard for comparisons
                                                     c_var,    # covariates - variables used as
covariates (age, sex, APOE normally)
                                                     analytes,
                                                     l_var)
proc.time() - ptm

##### CDR positive
ptm <- proc.time()
comparison_standard <- Eval_comp_standard(subset(data.in,CDR_10==1),o_vars[3],i_var)
comparison_standard_partial <-
Eval_comp_standard_partial(subset(data.in,CDR_10==1),o_vars[3],c_var,i_var)

table_out_TauLate_cdrpos <-  spearman_plasma_runs(subset(data.in,CDR_10==1), # Data inputted
                                                   o_vars[3],  # variable A - continuous variable
being correlated
                                                   i_var,# vector of strings - continuous variables
being correlated with variable A
                                                  comparison_standard,
                                                  comparison_standard_partial, #variable in
vars_to_cor vector that is used as standard for comparisons
                                                   c_var,    # covariates - variables used as
covariates (age, sex, APOE normally)
                                                   analytes,
                                                   l_var)
proc.time() - ptm

##### CDR negative
ptm <- proc.time()
comparison_standard <- Eval_comp_standard(subset(data.in,CDR_10==0),o_vars[3],i_var)
comparison_standard_partial <-
Eval_comp_standard_partial(subset(data.in,CDR_10==0),o_vars[3],c_var,i_var)

table_out_TauLate_cdrneg <-  spearman_plasma_runs(subset(data.in,CDR_10==0), # Data inputted
                                                   o_vars[3],  # variable A - continuous variable
being correlated
                                                   i_var,# vector of strings - continuous variables
being correlated with variable A
                                                   comparison_standard,
                                                  comparison_standard_partial, #variable in
vars_to_cor vector that is used as standard for comparisons
                                                   c_var,    # covariates - variables used as
covariates (age, sex, APOE normally)
                                                   analytes,
```

```
                                                        l_var)

proc.time() - ptm

#####Deprecated as analytes vector fixed
#list_taulate_new <- list()
#for(i in 1:length(list_of_TauLate_correlation_dataset)){
#
#
#  list_taulate_new[[i]] <-  relabel_spearman_res(list_of_TauLate_correlation_dataset[[i]])
#
#}
#
#names(list_taulate_new) <- names(list_of_TauLate_correlation_dataset)

list_of_TauLate_correlation_dataset <- list("Full Data"=table_out_TauLate_full,
                                            "Cent20 Positive Data"=table_out_TauLate_amy20pos,
                                            "Cent20 Negative Data"=table_out_TauLate_amy20neg,
                                            "Cent37 Positive Data"=table_out_TauLate_amy37pos,
                                            "Cent37 Negative Data"=table_out_TauLate_amy37neg,
                                            "CDR Positive Data"=table_out_TauLate_cdrpos,
                                            "CDR Negative Data"=table_out_TauLate_cdrneg)
####Due to length of table creation - save output data upon finishing
save(list_of_TauLate_correlation_dataset,
     file=paste0(out_results,"taulate_correlations.rdata"))
write.xlsx(list_of_TauLate_correlation_dataset,
           file=paste0(out_results,"TauLate_Correlations.xlsx"))

###Create Clean Plots
list_taulate_new <- list()
list_of_taulate_clean <- list()
for(i_clean in 1: length(list_of_TauLate_correlation_dataset)){
  DF <- list_of_TauLate_correlation_dataset[[i_clean]]

  DF$company <- factor(recode(DF$group,"quan"="Quanterix","alz"="ALZpath","c2n"="C2N",
                              "fuji"="Fujirebio", "jan"="Janssen","roche" = "Roche"))
  levels(DF$company) <- c("ALZpath","C2N", "Fujirebio",  "Janssen", "Quanterix","Roche")
  DF$company <-factor(DF$company, levels = c("C2N", "Fujirebio", "ALZpath", "Janssen", "Roche",
"Quanterix"))

  list_taulate_new[[i_clean]] <- DF
  list_of_taulate_clean[[i_clean]]  <- clean_spearman_table(DF)
}

names(list_of_taulate_clean) <- names(list_of_TauLate_correlation_dataset)
write.xlsx(list_of_taulate_clean,
           file=paste0(out_results,"TauLate_Correlations_05112024.xlsx"))

#####Full data
####Select comparison standards for each cohort - then run
comparison_standard <- Eval_comp_standard(data.in,o_vars[4],i_var)
comparison_standard_partial <- Eval_comp_standard_partial(data.in,o_vars[4],c_var,i_var)
ptm <- proc.time()
table_out_Atrophy_full <-  spearman_plasma_runs(data.in, # Data inputted
                                                o_vars[4],  # variable A - continuous variable
being correlated
                                                i_var,# vector of strings - continuous variables
being correlated with variable A
                                                comparison_standard,
                                                comparison_standard_partial, #variable in
vars_to_cor vector that is used as standard for comparisons
                                                c_var,    # covariates - variables used as
covariates (age, sex, APOE normally)
                                                analytes,
```

```
                                                    l_var)
proc.time() - ptm

#####Centiloid 20 positive
ptm <- proc.time()
comparison_standard <- Eval_comp_standard(subset(data.in,CENTILOIDS_10==1),o_vars[4],i_var)
comparison_standard_partial <-
Eval_comp_standard_partial(subset(data.in,CENTILOIDS_10==1),o_vars[4],c_var,i_var)

table_out_Atrophy_amy20pos <-  spearman_plasma_runs(subset(data.in,CENTILOIDS_10==1), # Data
inputted
                                                    o_vars[4],  # variable A - continuous variable
being correlated
                                                    i_var,# vector of strings - continuous
variables being correlated with variable A
                                                    comparison_standard,
                                                    comparison_standard_partial, #variable in
vars_to_cor vector that is used as standard for comparisons
                                                    c_var,    # covariates - variables used as
covariates (age, sex, APOE normally)
                                                    analytes,
                                                    l_var)
proc.time() - ptm

#### Centiloid 20 negative
ptm <- proc.time()
comparison_standard <- Eval_comp_standard(subset(data.in,CENTILOIDS_10==0),o_vars[4],i_var)
comparison_standard_partial <-
Eval_comp_standard_partial(subset(data.in,CENTILOIDS_10==0),o_vars[4],c_var,i_var)
table_out_Atrophy_amy20neg <-  spearman_plasma_runs(subset(data.in,CENTILOIDS_10==0), # Data
inputted
                                                    o_vars[4],  # variable A - continuous variable
being correlated
                                                    i_var,# vector of strings - continuous
variables being correlated with variable A
                                                    comparison_standard,
                                                    comparison_standard_partial, #variable in
vars_to_cor vector that is used as standard for comparisons
                                                    c_var,    # covariates - variables used as
covariates (age, sex, APOE normally)
                                                    analytes,
                                                    l_var)

proc.time() - ptm

##### Centiloid 37 Positive
ptm <- proc.time()
comparison_standard <- Eval_comp_standard(subset(data.in,CENTILOIDS_10_sens==1),o_vars[4],i_var)
comparison_standard_partial <-
Eval_comp_standard_partial(subset(data.in,CENTILOIDS_10_sens==1),o_vars[4],c_var,i_var)

table_out_Atrophy_amy37pos <-  spearman_plasma_runs(subset(data.in,CENTILOIDS_10_sens==1), # Data
inputted
                                                    o_vars[4],  # variable A - continuous variable
being correlated
                                                    i_var,# vector of strings - continuous
variables being correlated with variable A
                                                    comparison_standard,
                                                    comparison_standard_partial, #variable in
vars_to_cor vector that is used as standard for comparisons
                                                    c_var,    # covariates - variables used as
covariates (age, sex, APOE normally)
                                                    analytes,
                                                    l_var)
proc.time() - ptm

##### Centiloid 37 negative
```

```
ptm <- proc.time()
comparison_standard <- Eval_comp_standard(subset(data.in,CENTILOIDS_10_sens==0),o_vars[4],i_var)
comparison_standard_partial <-
Eval_comp_standard_partial(subset(data.in,CENTILOIDS_10_sens==0),o_vars[4],c_var,i_var)

table_out_Atrophy_amy37neg <-  spearman_plasma_runs(subset(data.in,CENTILOIDS_10_sens==0), # Data
inputted
                                                    o_vars[4],  # variable A - continuous variable
being correlated
                                                    i_var,# vector of strings - continuous
variables being correlated with variable A
                                                    comparison_standard,
                                                    comparison_standard_partial, #variable in
vars_to_cor vector that is used as standard for comparisons
                                                    c_var,    # covariates - variables used as
covariates (age, sex, APOE normally)
                                                    analytes,
                                                    l_var)
proc.time() - ptm

##### CDR positive
ptm <- proc.time()
comparison_standard <- Eval_comp_standard(subset(data.in,CDR_10==1),o_vars[4],i_var)

comparison_standard_partial <-
Eval_comp_standard_partial(subset(data.in,CDR_10==1),o_vars[4],c_var,i_var)

table_out_Atrophy_cdrpos <-  spearman_plasma_runs(subset(data.in,CDR_10==1), # Data inputted
                                                  o_vars[4],  # variable A - continuous variable
being correlated
                                                  i_var,# vector of strings - continuous variables
being correlated with variable A
                                                  comparison_standard,
                                                  comparison_standard_partial, #variable in
vars_to_cor vector that is used as standard for comparisons
                                                  c_var,    # covariates - variables used as
covariates (age, sex, APOE normally)
                                                  analytes,
                                                  l_var)
proc.time() - ptm

##### CDR negative
ptm <- proc.time()
comparison_standard <- Eval_comp_standard(subset(data.in,CDR_10==0),o_vars[4],i_var)
comparison_standard_partial <-
Eval_comp_standard_partial(subset(data.in,CDR_10==0),o_vars[4],c_var,i_var)

table_out_Atrophy_cdrneg <-  spearman_plasma_runs(subset(data.in,CDR_10==0), # Data inputted
                                                  o_vars[4],  # variable A - continuous variable
being correlated
                                                  i_var,# vector of strings - continuous variables
being correlated with variable A
                                                  comparison_standard,
                                                  comparison_standard_partial, #variable in
vars_to_cor vector that is used as standard for comparisons
                                                  c_var,    # covariates - variables used as
covariates (age, sex, APOE normally)
                                                  analytes,
                                                  l_var)

#### Assemble outputs into list
list_of_Atrophy_correlation_dataset <- list("Full Data"=table_out_Atrophy_full,
                                            "Cent20 Positive Data"=table_out_Atrophy_amy20pos,
                                            "Cent20 Negative Data"=table_out_Atrophy_amy20neg,
                                            "Cent37 Positive Data"=table_out_Atrophy_amy37pos,
                                            "Cent37 Negative Data"=table_out_Atrophy_amy37neg,
                                            "CDR Positive Data"=table_out_Atrophy_cdrpos,
```

```
                                              "CDR Negative Data"=table_out_Atrophy_cdrneg)

####Due to length of table creation - save output data upon finishing
save(list_of_Atrophy_correlation_dataset,
     file=paste0(out_results,"atrophy_correlations.rdata"))

#### Clean up only necessary before analytes vector correction
#list_atrophy_new <- list()
#for(i in 1:length(list_of_Atrophy_correlation_dataset)){
#
#
#  list_atrophy_new[[i]] <-  relabel_spearman_res(list_of_Atrophy_correlation_dataset[[i]])
#
#}
#
#names(list_atrophy_new) <- names(list_of_Atrophy_correlation_dataset)
#####

write.xlsx(list_of_Atrophy_correlation_dataset,
           file=paste0(out_results,"Atrophy_Correlations.xlsx"))

###Create Clean Plots
list_atrophy_new <- list()
list_of_atrophy_clean <- list()
for(i_clean in 1: length(list_of_Atrophy_correlation_dataset)){
  DF <- list_of_Atrophy_correlation_dataset[[i_clean]]

  DF$company <- factor(recode(DF$group,"quan"="Quanterix","alz"="ALZpath","c2n"="C2N",
                              "fuji"="Fujirebio", "jan"="Janssen","roche" = "Roche"))
  levels(DF$company) <- c("ALZpath","C2N", "Fujirebio",  "Janssen", "Quanterix","Roche")
  DF$company <-factor(DF$company, levels = c("C2N", "Fujirebio", "ALZpath", "Janssen", "Roche",
"Quanterix"))
  list_atrophy_new[[i_clean]] <- DF

  list_of_atrophy_clean[[i_clean]]  <- clean_spearman_table(DF)
}

names(list_of_atrophy_clean) <- names(list_of_Atrophy_correlation_dataset)
write.xlsx(list_of_atrophy_clean,
           file=paste0(out_results,"Atrophy_Correlations_05132024.xlsx"))

#### Ended up not being used for final paper, here for documationation purposesd

#Atrophyplots <- list()
#for(i_plot in 1:length(list_atrophy_new)){
#  Atrophyplots[[i_plot]] <- spearman_gg_formanuscript(list_atrophy_new[[i_plot]],breaks1 =
plot_breaks[[4]],indicatorline = -0.25,rev_axis = TRUE)
#}

#for(i_plot in 1:length(list_atrophy_new)){
#  ggsave(paste0(out_plots,names(list_atrophy_new)[i_plot],"_Atrophy_Spearman_Forest.pdf"),
#         plot = Atrophyplots[[i_plot]],
#         device=cairo_pdf)#writes out spearman plot
#
#}

###CDR Sumbox

####Create comparison standards for unadjusted and adjusted spearmans
comparison_standard <- Eval_comp_standard(data.in,o_vars[5],i_var)
```

```
comparison_standard_partial <- Eval_comp_standard_partial(data.in,o_vars[5],c_var,i_var)

ptm <- proc.time()

####Run spearman table

### Full Data
table_out_sumbox_full <-  spearman_plasma_runs(data.in, # Data inputted
                                                o_vars[5],  # variable A - continuous variable
being correlated
                                                i_var,# vector of strings - continuous variables
being correlated with variable A
                                               comparison_standard,
                                               comparison_standard_partial, #variable in
vars_to_cor vector that is used as standard for comparisons
                                                c_var,    # covariates - variables used as
covariates (age, sex, APOE normally)
                                                analytes,
                                                l_var)

proc.time() - ptm

###### Centiloid 20 positive
ptm <- proc.time()
comparison_standard <- Eval_comp_standard(subset(data.in,CENTILOIDS_10==1),o_vars[5],i_var)
comparison_standard_partial <-
Eval_comp_standard_partial(subset(data.in,CENTILOIDS_10==1),o_vars[5],c_var,i_var)

table_out_sumbox_amy20pos <-  spearman_plasma_runs(subset(data.in,CENTILOIDS_10==1), # Data
inputted
                                                    o_vars[5],  # variable A - continuous variable
being correlated
                                                    i_var,# vector of strings - continuous
variables being correlated with variable A
                                                   comparison_standard,
                                                   comparison_standard_partial, #variable in
vars_to_cor vector that is used as standard for comparisons
                                                    c_var,    # covariates - variables used as
covariates (age, sex, APOE normally)
                                                    analytes,
                                                    l_var)
proc.time() - ptm

####Centiloid 20 Negative
ptm <- proc.time()
comparison_standard <- Eval_comp_standard(subset(data.in,CENTILOIDS_10==0),o_vars[5],i_var)
comparison_standard_partial <-
Eval_comp_standard_partial(subset(data.in,CENTILOIDS_10==0),o_vars[5],c_var,i_var)
table_out_sumbox_amy20neg <-  spearman_plasma_runs(subset(data.in,CENTILOIDS_10==0), # Data
inputted
                                                    o_vars[5],  # variable A - continuous variable
being correlated
                                                    i_var,# vector of strings - continuous
variables being correlated with variable A
                                                   comparison_standard,
                                                   comparison_standard_partial, #variable in
vars_to_cor vector that is used as standard for comparisons
                                                    c_var,    # covariates - variables used as
covariates (age, sex, APOE normally)
                                                    analytes,
                                                    l_var)
proc.time() - ptm

#### Centiloid 37 Positive
ptm <- proc.time()
comparison_standard <- Eval_comp_standard(subset(data.in,CENTILOIDS_10_sens==1),o_vars[5],i_var)
```

```
comparison_standard_partial <-
Eval_comp_standard_partial(subset(data.in,CENTILOIDS_10_sens==1),o_vars[5],c_var,i_var)
table_out_sumbox_amy37pos <-  spearman_plasma_runs(subset(data.in,CENTILOIDS_10_sens==1), # Data
inputted
                                                    o_vars[5],  # variable A - continuous variable
being correlated
                                                    i_var,# vector of strings - continuous
variables being correlated with variable A
                                                   comparison_standard,
                                                   comparison_standard_partial, #variable in
vars_to_cor vector that is used as standard for comparisons
                                                    c_var,    # covariates - variables used as
covariates (age, sex, APOE normally)
                                                    analytes,
                                                    l_var)
proc.time() - ptm

####Centiloid 37 Negative
ptm <- proc.time()
comparison_standard <- Eval_comp_standard(subset(data.in,CENTILOIDS_10_sens==0),o_vars[5],i_var)
comparison_standard_partial <-
Eval_comp_standard_partial(subset(data.in,CENTILOIDS_10_sens==0),o_vars[5],c_var,i_var)
table_out_sumbox_amy37neg <-  spearman_plasma_runs(subset(data.in,CENTILOIDS_10_sens==0), # Data
inputted
                                                    o_vars[5],  # variable A - continuous variable
being correlated
                                                    i_var,# vector of strings - continuous
variables being correlated with variable A
                                                   comparison_standard,
                                                   comparison_standard_partial, #variable in
vars_to_cor vector that is used as standard for comparisons
                                                    c_var,    # covariates - variables used as
covariates (age, sex, APOE normally)
                                                    analytes,
                                                    l_var)
proc.time() - ptm

#####CDR positive
ptm <- proc.time()
comparison_standard <- Eval_comp_standard(subset(data.in,CDR_10==1),o_vars[5],i_var)
comparison_standard_partial <-
Eval_comp_standard_partial(subset(data.in,CDR_10==1),o_vars[5],c_var,i_var)

table_out_sumbox_cdrpos <-  spearman_plasma_runs(subset(data.in,CDR_10==1), # Data inputted
                                                  o_vars[5],  # variable A - continuous variable
being correlated
                                                  i_var,# vector of strings - continuous variables
being correlated with variable A
                                                  comparison_standard,
                                                 comparison_standard_partial,#variable in
vars_to_cor vector that is used as standard for comparisons
                                                  c_var,    # covariates - variables used as
covariates (age, sex, APOE normally)
                                                  analytes,
                                                  l_var)
proc.time() - ptm
#####CDR negative - unnecessary for sum of boxes correlations
#table_out_sumbox_cdrneg <-  spearman_plasma_runs(subset(data.in,CDR_10==0), # Data inputted
#                                                  o_vars[5],  # variable A - continuous variable
being correlated
#                                                  i_var,# vector of strings - continuous variables
being correlated with variable A
#                                                  comparison_standard, #variable in vars_to_cor
vector that is used as standard for comparisons
#                                                 c_var,    # covariates - variables used as
covariates (age, sex, APOE normally)
#                                                  analytes,
```

```
#                                                   l_var)

#####Create list of results datasets
list_of_sumbox_correlation_dataset <- list("Full Data"=table_out_sumbox_full,
                                            "Cent20 Positive Data"=table_out_sumbox_amy20pos,
                                            "Cent20 Negative Data"=table_out_sumbox_amy20neg,
                                            "Cent37 Positive Data"=table_out_sumbox_amy37pos,
                                            "Cent37 Negative Data"=table_out_sumbox_amy37neg,
                                            "CDR Positive Data"=table_out_sumbox_cdrpos)

write.xlsx(list_of_sumbox_correlation_dataset,
           file=paste0(out_results,"sumbox_Correlations.xlsx"))

####Due to length of table creation - save output data upon finishing
save(list_of_sumbox_correlation_dataset,
     file=paste0(out_results,"cdrsumbox_correlations.rdata"))

###Create Clean tables - These will create publication quality tables
list_of_sumbox_clean <- list()
list_cdr_new <- list()
for(i_clean in 1: length(list_of_sumbox_correlation_dataset)){
  DF <- list_of_sumbox_correlation_dataset[[i_clean]] ###Asign Dataset to work on

  DF$company <- factor(recode(DF$group,"quan"="Quanterix","alz"="ALZpath","c2n"="C2N",
                              "fuji"="Fujirebio", "jan"="Janssen","roche" = "Roche")) ###Recode
group to company
  levels(DF$company) <- c("ALZpath","C2N", "Fujirebio",  "Janssen", "Quanterix","Roche")
  DF$company <-factor(DF$company, levels = c("C2N", "Fujirebio", "ALZpath", "Janssen", "Roche",
"Quanterix")) #Order company factor
  list_cdr_new[[i_clean]] <- DF # this changed after the analytes vector was corrected

  list_of_sumbox_clean[[i_clean]]  <- clean_spearman_table(DF) ###Call the cleaning function
}

names(list_of_sumbox_clean) <- names(list_of_sumbox_correlation_dataset) #make sure all the dataset
labels are correct

###Write out clean - publication formatted datasets
write.xlsx(list_of_sumbox_clean,
           file=paste0(out_results,"CDR_Sumbox_Correlations_05132024.xlsx"))

####

cdr_index <- index_biomarker_forest(table_out_cdr, #dataset to form basis for index
                       which(colnames(table_out_cdr)=="rho"), #column number for the metric being
used - basis
                       which(colnames(table_out_cdr)=="group"),#column number for group variable -
basis
                       "C2N", #way to distinguish groups within groups (basically, given this is
nested, this tells which to highlight)
                       which(colnames(table_out_cdr)=="Biomarker"), #Biomarker identification
variable where the function should search for a way to distinguish
                      l_var[-which(i_var==comparison_standard)])

cdr_plot <- Forest_plot_create(table_out_cdr, #creates forest plots for spearmans
                               cdr_index,
                               "CDR Sum of Boxes Correlations",
                               "Spearman Rho",
                               reverse_scales=FALSE)

##########These were more necessary when the
#list_cdr_new <- list()
#for(i in 1:length(list_of_sumbox_correlation_dataset)){
#
#
```

```
#  list_cdr_new[[i]] <-  relabel_spearman_res(list_of_sumbox_correlation_dataset[[i]])

#}

#names(list_cdr_new) <- names(list_of_sumbox_correlation_dataset)

save.image("/FNIH_Project_2/Current_Analysis_data.rdata")
###########################
#### Plots ##### All Functions can be found in Functions_for_Analysis.R
###########################

plot_breaks <- list() ##create list of breaks for each plot
plot_breaks[[1]] <- c(0.20, 0.40, 0.60, 0.80)
plot_breaks[[2]] <- c(0, 0.2, 0.4, 0.6, 0.8)
plot_breaks[[3]] <- c(0, 0.2, 0.4, 0.6, 0.8)
plot_breaks[[4]] <- c(0.2, 0,-0.2, -0.40)
plot_breaks[[5]] <- c(0.1, 0.3, 0.5)
names(plot_breaks) <- c("Centiloid","TauEarly","TauLate","Atrophy","CDR_SB")

refline_curr <- c(0.6,0.4,-0.2,0.3) ##create vector of positions for reference line (line is
arbitrary for visulization)
## we did try several solutions to make a line that would have some meaningful purpose within the
dataset but it ultimately didn't
## work out with our range of values

####Full cohort plots
cent_index <- index_biomarker_forest(list_cent_new[[1]]) ### input results dataset

##### Centiloid
centiloid_plot_all <- spearman_gg_formanuscript(list_cent_new[[1]], ### Results Date
                                                breaks1 = plot_breaks$Centiloid, #### Breaks
                                                df_plot_index = cent_index, #### Index from
Biomarker Forest index function
                                                indicatorline = refline_curr[1], ###Reference line
for plot
                                                labels_all = TRUE, ####Indicates whether or not the
labels will show up on this plot
                                                title_plot = "Amyloid PET", #### Title of plot
                                                amytag=TRUE) #### Specific paramenter to set axis
centiloid_plot_all <- centiloid_plot_all+ggtitle("Amyloid PET")

#### Tau Early
tauearly_plot_all <- spearman_gg_formanuscript(list_tauearly_new[[1]],breaks1 =
plot_breaks$TauEarly,
                                               df_plot_index = cent_index,indicatorline =
refline_curr[2],labels_all = FALSE,title_plot = "Early tau PET")
tauearly_plot_all <- tauearly_plot_all+ggtitle("Early tau PET")

#### Atrophy
atrophy_plot_all <- spearman_gg_formanuscript(list_atrophy_new[[1]],breaks1 = plot_breaks$Atrophy,
                                              df_plot_index = cent_index,indicatorline =
refline_curr[3],rev_axis = TRUE,labels_all = FALSE,
                                              title_plot = "Cortical thickness")
atrophy_plot_all <- atrophy_plot_all+ggtitle("Cortical thickness")

##### CDR Sum of Boxes
cdr_plot_all <- spearman_gg_formanuscript(list_cdr_new[[1]],breaks1 = plot_breaks$CDR_SB,
                                          df_plot_index = cent_index,indicatorline =
refline_curr[4],labels_all = FALSE,title_plot = "Dementia severity")

cdr_plot_all <- cdr_plot_all+ggtitle("Dementia severity")
plot_out <- centiloid_plot_all | tauearly_plot_all | atrophy_plot_all | cdr_plot_all

ggsave(paste0(out_plots,"Full_cohort_ATN.jpg"),
```

```
       plot=plot_out,
       device=jpeg,width = 8,height=4.02)
####Amyloid Positive cohort plots
cent_index <- index_biomarker_forest(list_cent_new[[2]])

##### Centiloid
centiloid_plot_amypos <- spearman_gg_formanuscript(list_cent_new[[2]],breaks1 =
c(plot_breaks$Centiloid,0,-0.2),
                                                   df_plot_index = cent_index,indicatorline =
0.6,labels_all = TRUE,title_plot = "Amyloid PET")
centiloid_plot_amypos <- centiloid_plot_amypos+ggtitle("Amyloid PET")

##### Tau Early
tauearly_plot_amypos <- spearman_gg_formanuscript(list_tauearly_new[[2]],breaks1 =
c(plot_breaks$TauEarly,-0.2),
                                                  df_plot_index = cent_index,indicatorline =
0.4,labels_all = FALSE,title_plot = "Early tau PET")
tauearly_plot_amypos <- tauearly_plot_amypos+ggtitle("Early tau PET")

##### Atrophy
atrophy_plot_amypos <- spearman_gg_formanuscript(list_atrophy_new[[2]],breaks1 =
c(plot_breaks$Atrophy,-0.6),
                                                 df_plot_index = cent_index,indicatorline = -
0.2,rev_axis = TRUE,labels_all = FALSE,
                                                 title_plot = "Cortical thickness")
atrophy_plot_amypos <- atrophy_plot_amypos+ggtitle("Cortical thickness")

##### CDR Sum of boxes
cdr_plot_amypos <- spearman_gg_formanuscript(list_cdr_new[[2]],breaks1 = c(plot_breaks$CDR_SB,-
0.1),
                                             df_plot_index = cent_index,indicatorline =
0.3,labels_all = FALSE,title_plot = "Dementia severity")

cdr_plot_amypos <- cdr_plot_amypos+ggtitle("Dementia severity")
plot_out <- centiloid_plot_amypos | tauearly_plot_amypos | atrophy_plot_amypos | cdr_plot_amypos

ggsave(paste0(out_plots,"Amyloid_PET_Positive_cohort_ATN.jpg"),
       plot=plot_out,
       device=jpeg,width = 8,height=4.02)

####Amyloid Negative cohort plots
cent_index <- index_biomarker_forest(list_cent_new[[3]])

###### Centiloid
centiloid_plot_amyneg <- spearman_gg_formanuscript(list_cent_new[[3]],breaks1 =
c(plot_breaks$Centiloid,0,-0.2),
                                                   df_plot_index = cent_index,indicatorline =
0.6,labels_all = TRUE,title_plot = "Amyloid PET")
centiloid_plot_amyneg <- centiloid_plot_amyneg+ggtitle("Amyloid PET")

##### Tau Early
tauearly_plot_amyneg <- spearman_gg_formanuscript(list_tauearly_new[[3]],breaks1 =
c(plot_breaks$TauEarly,-0.2),
                                                  df_plot_index = cent_index,indicatorline =
0.4,labels_all = FALSE,title_plot = "Early tau PET")
tauearly_plot_amyneg <- tauearly_plot_amyneg+ggtitle("Early tau PET")

##### Atrophy
atrophy_plot_amyneg <- spearman_gg_formanuscript(list_atrophy_new[[3]],breaks1 =
c(plot_breaks$Atrophy,-0.6),
                                                 df_plot_index = cent_index,indicatorline = -
0.2,rev_axis = TRUE,labels_all = FALSE,
                                                 title_plot = "Cortical thickness")
atrophy_plot_amyneg <- atrophy_plot_amyneg+ggtitle("Cortical thickness")
```

```
##### CDR Sum of Boxes
cdr_plot_amyneg <- spearman_gg_formanuscript(list_cdr_new[[3]],breaks1 = c(plot_breaks$CDR_SB,-0.1),
                                             df_plot_index = cent_index,indicatorline = 0.3,labels_all = FALSE,title_plot = "Dementia severity")

cdr_plot_amyneg <- cdr_plot_amyneg+ggtitle("Dementia severity")
plot_out <- centiloid_plot_amyneg | tauearly_plot_amyneg | atrophy_plot_amyneg | cdr_plot_amyneg

ggsave(paste0(out_plots,"Amyloid_PET_negative_cohort_ATN.jpg"),
       plot=plot_out,
       device=jpeg,width = 8,height=4.02)

######Cognitively Impaired Cohort Plots
cent_index <- index_biomarker_forest(list_cent_new[[6]])

#####Centiloid
centiloid_plot_cdrpos <- spearman_gg_formanuscript(list_cent_new[[6]],breaks1 = plot_breaks$Centiloid,
                                                   df_plot_index = cent_index,indicatorline = 0.6,labels_all = TRUE,title_plot = "Amyloid PET",
                                                   amytag=TRUE)
centiloid_plot_cdrpos <- centiloid_plot_cdrpos+ggtitle("Amyloid PET")

##### Tau Early
tauearly_plot_cdrpos <- spearman_gg_formanuscript(list_tauearly_new[[6]],breaks1 = plot_breaks$TauEarly,
                                                  df_plot_index = cent_index,indicatorline = 0.4,labels_all = FALSE,title_plot = "Early tau PET")
tauearly_plot_cdrpos <- tauearly_plot_cdrpos+ggtitle("Early tau PET")

###### Atrophy
atrophy_plot_cdrpos <- spearman_gg_formanuscript(list_atrophy_new[[6]],breaks1 = plot_breaks$Atrophy,
                                                 df_plot_index = cent_index,indicatorline = -0.2,rev_axis = TRUE,labels_all = FALSE,
                                                 title_plot = "Cortical thickness")
atrophy_plot_cdrpos <- atrophy_plot_cdrpos+ggtitle("Cortical thickness")

######CDR Sum of Boxes
cdr_plot_cdrpos <- spearman_gg_formanuscript(list_cdr_new[[6]],breaks1 = plot_breaks$CDR_SB,
                                             df_plot_index = cent_index,indicatorline = 0.3,labels_all = FALSE,title_plot = "Dementia severity")

cdr_plot_cdrpos <- cdr_plot_cdrpos+ggtitle("Dementia severity")
plot_out <- centiloid_plot_cdrpos | tauearly_plot_cdrpos | atrophy_plot_cdrpos | cdr_plot_cdrpos

ggsave(paste0(out_plots,"Cognitively_impaired_cohort_ATN.jpg"),
       plot=plot_out,
       device=jpeg,width = 8,height=4.02)

######Cognitively Unimpaired Cohort Plots
cent_index <- index_biomarker_forest(list_cent_new[[7]])

##### Centiloid
centiloid_plot_cdrneg <- spearman_gg_formanuscript(list_cent_new[[7]],breaks1 = c(0,plot_breaks$Centiloid),
                                                   df_plot_index = cent_index,indicatorline = 0.6,labels_all = TRUE,title_plot = "Amyloid PET",
                                                   amytag=TRUE)
centiloid_plot_cdrneg <- centiloid_plot_cdrneg+ggtitle("Amyloid PET")

#####Tau Early
```

```
tauearly_plot_cdrneg <- spearman_gg_formanuscript(list_tauearly_new[[7]],breaks1 =
plot_breaks$TauEarly,
                                                  df_plot_index = cent_index,indicatorline =
0.4,labels_all = FALSE,title_plot = "Early tau PET")
tauearly_plot_cdrneg <- tauearly_plot_cdrneg+ggtitle("Early tau PET")

#### Atrophy
atrophy_plot_cdrneg <- spearman_gg_formanuscript(list_atrophy_new[[7]],breaks1 = c(0.2,0.0,-0.2,-
0.4,-0.6),
                                                 df_plot_index = cent_index,indicatorline = -
0.2,rev_axis = TRUE,labels_all = FALSE,
                                                 title_plot = "Cortical thickness")
atrophy_plot_cdrneg <- atrophy_plot_cdrneg+ggtitle("Cortical thickness")

plot_out <- centiloid_plot_cdrneg | tauearly_plot_cdrneg | atrophy_plot_cdrneg

ggsave(paste0(out_plots,"Cognitively_unimpaired_cohort_ATN.jpg"),
       plot=plot_out,
       device=jpeg,width = 8,height=4.02)

####Centiloid All PLots
cent_index <- index_biomarker_forest(list_cent_new[[6]])

#### Amyloid Positive
centiloid_plot_amypos <- spearman_gg_formanuscript(list_cent_new[[2]],breaks1 =
c(plot_breaks$Centiloid,0,-0.2),
                                                   df_plot_index = cent_index,indicatorline =
0.6,labels_all = FALSE,title_plot = "Amyloid PET\n positive")
#### Amyloid Negative
centiloid_plot_amyneg <- spearman_gg_formanuscript(list_cent_new[[3]],breaks1 =
c(plot_breaks$Centiloid,0,-0.2),
                                                   df_plot_index = cent_index,indicatorline =
0.6,labels_all = FALSE,title_plot = "Amyloid PET\n negative")

#### CDR Positive
centiloid_plot_cdrpos <- spearman_gg_formanuscript(list_cent_new[[6]],breaks1 =
c(plot_breaks$Centiloid,0,-0.2),
                                                   df_plot_index = cent_index,indicatorline =
0.6,labels_all = TRUE,title_plot = "Cognitively\n impaired",
                                                   amytag=TRUE)
#### CDR Negative
centiloid_plot_cdrneg <- spearman_gg_formanuscript(list_cent_new[[7]],breaks1 =
c(plot_breaks$Centiloid,0,-0.2),
                                                   df_plot_index = cent_index,indicatorline =
0.6,labels_all = FALSE,title_plot = "Cognitively\n unimpaired",
                                                   amytag=TRUE)

#### Combine plots
plot_out <-  centiloid_plot_cdrpos | centiloid_plot_cdrneg | centiloid_plot_amypos |
centiloid_plot_amyneg

#### write out plots
ggsave(paste0(out_plots,"Centiloid_all_ATN.png"),
       plot=plot_out,
       device=png,dpi = 1200,width = 8,height=4.02)
```

**Code B8. R code for scatter plots (uses function bank).**

```
###Spearman Scatter Plots
#### Coded: Benjamin Saef
#### Date as of commenting and documentation - 05/28/2024

#Scatter plots for best performing analytes from each platform (Benjamin) versus amyloid PET (x-
axis):
#  Yellowish green: amyloid PET negative, CDR >0
#Blue: amyloid PET negative, CDR 0
#Orange: amyloid PET positive, CDR 0
#Red: amyloid PET positive, CDR >0
#a.    Amyloid PET Centiloid
#b.    Early Tau PET
#c.    Brain atrophy
#d.    Cognitive impairment
library(tidyr)
library(data.table)
library(RVAideMemoire)
library(pROC)
library(RColorBrewer)
library(MuMIn)

####Set project directory
out_project <- "/FNIH_Project_2/Paper1/Spearman_Analysis" #Folder where you want all output written
to, a results and plot folder will be written here by default
out_plots <- paste0(out_project,"/plots/scatter/")

dir.create(out_plots,showWarnings = F)

data_file <- "/FNIH_Project_2/DATA_STUDY_2/cross_sectional_data_2024_04_24b.csv"  # input cross-
sectional data

data.in <- read.csv(data_file)

#analysis vars
#C2N
pTau217_c2n <- "C2N_plasma_ptau217_ratio" #Variable representing p-tau217 ratio - c2n
pTau217conc_c2n <- "C2N_plasma_ptau217" #Variable representing p-tau217 concentration - c2n
abeta42_c2n <- "C2N_plasma_Abeta42" #Variable representing Ab42 - c2n
abeta40_c2n <- "C2N_plasma_Abeta42_Abeta40" #Variable representing Ab40 - c2n
abeta4240_c2n <- "C2N_plasma_Abeta42_Abeta40" #Variable representing Ab42/Ab40 - c2n

#Elecsys
abeta42_esys <- "Roche_plasma_Ab42" #Variable representing Ab42 - elecsys
abeta40_esys <- "Roche_plasma_Ab40" #Variable representing Ab40 - elecsys
abeta4240_esys <- "Roche_plasma_Ab42_Ab40" #Variable representing Ab42/Ab40 - elecsys
ptau_181_esys <- "Roche_plasma_ptau181" #Variable representing ptau181 - elecsys
GFAP_esys <- "Roche_plasma_GFAP" #Variable representing GFAP - elecsys
NfL_esys <- "Roche_plasma_NfL" #Variable representing NfL - elecsys

#Lumipulse

abeta42_lumi <- "Fuji_plasma_Ab42" #Variable representing Ab42 - lumipulse
abeta40_lumi <- "Fuji_plasma_Ab40" #Variable representing Ab40 - lumipulse
abeta4240_lumi <- "Fuji_plasma_Ab42_Ab40" #Variable representing Ab42/Ab40 - lumipulse
pTau217_lumi <- "Fuji_plasma_ptau217" #Variable representing p-tau217 ratio - lumipulse

#Simoa
alzpath_ptau217 <- "AlzPath_plasma_ptau217"  #Variable representing p-tau217 - AlzPath
janssen_ptau217 <- "Janssen_plasma_ptau217"  #Variable representing p-tau217 - Janssen

#Quanterix - Simoa
```

```
abeta42_quan <- "QX_plasma_Ab42" #Variable representing Ab42 - elecsys
abeta40_quan <- "QX_plasma_Ab40" #Variable representing Ab40 - elecsys
abeta4240_quan <- "QX_plasma_Ab42_Ab40" #Variable representing Ab42/Ab40 - elecsys
ptau_181_quan <- "QX_plasma_ptau181" #Variable representing ptau181 - elecsys
GFAP_quan <- "QX_plasma_GFAP" #Variable representing GFAP - elecsys
NfL_quan <- "QX_plasma_NfL" #Variable representing NfL - elecsys

#Variabels for PET metrics
centiloid_Variable <- "CENTILOIDS" #Variable representing Centiloid
early_tau <- "MesialTemporal" #Variable representing Tau PET Centaur
late_tau <- "TemporoParietal"
atrophy <- "atrophy"

l_var <- c( "ab4240", #Ab42/40
            "pt217", #ptau217
            "APS", #APS
            "ab4240", #Ab42/40
            "pt181", #ptau181
            "gfap", #GFAP
            "nfl", #NFL
            "ab4240",#Ab42/40
            "pt217",#ptau217
            "pt217",#ptau217
            "pt217",#ptau217
            "ab4240", #Ab42/40
            "pt181", #ptau181
            "gfap", #GFAP
            "nfl"#NFL
)

##list out the different analytes - Labels
analytes <- c(paste0("A","\U03B2","42/A","\U03B2","40"), #Ab42/40
              "p-tau217 ratio (%)", #ptau217
              "p-tau217 (pg/ml)", #ptau217
              paste0("A","\U03B2","42/A","\U03B2","40"), #Ab42/40
              "p-tau181 (pg/ml)", #ptau181
              "GFAP (ng/ml)", #GFAP
              "NfL (pg/mL)", #NFL
              paste0("A","\U03B2","42/A","\U03B2","40"), #Ab42/40
              "p-tau217 (pg/ml)", #ptau217
              "p-tau217 (pg/ml)", #ptau217
              "p-tau217 (pg/ml)", #ptau217
              paste0("A","\U03B2","42/A","\U03B2","40"), #Ab42/40
              "p-tau181 (pg/ml)", #ptau181
              "GFAP (pg/ml)", #GFAP
              "NfL (pg/mL)" #NFL
)
#list out the variables representing those analytes.
i_var <- c(abeta4240_c2n, #Ab42/40 - c2n
           pTau217_c2n, #ptau217 - c2n
           APS_c2n,#aps - c2n
           abeta4240_esys,#Ab42/40 - elecsys
           ptau_181_esys,#ptau181 - elecsys
           GFAP_esys,#GFAP - elecsys
           NfL_esys, #NFL - elecsys
           abeta4240_lumi,#Ab42/40 - lumipulse
           pTau217_lumi, #ptau217 - lumipulse
           alzpath_ptau217,#ptau217 - alzpath
           janssen_ptau217,#ptau217 - lumipulse
           abeta4240_quan,#Ab42/40 - quanterix
           ptau_181_quan,#ptau181 - quanterix
           GFAP_quan,#GFAP - quanterix
           NfL_quan #NFL - quanterix
)
```

```
ref_data <- data.frame(group=l_var,var_id=i_var,label=analytes)

###Create Scatter plot figure for Centiloid results
load("/FNIH_Project_2/Paper1/Spearman_Analysis/results/cent_correlations.rdata") #load in results
data from Spearman_Analysis.R code
plot_out <- make_scatter_figure(data.in,
                    ref_data,
                    o_vars[1],
                    list_of_cent_correlation_dataset[[1]],
                    "Amyloid PET Centiloid")
ggsave(paste0(out_plots,"Centiloid_Scatter_ATN.png"),
       plot=plot_out,
       device=png,dpi = 1200,width = 8,height=12)

###Create Scatter plot figure for Early Tau results
load("/FNIH_Project_2/Paper1/Spearman_Analysis/results/tauearly_correlations.rdata") #load in
results data from Spearman_Analysis.R code
plot_out <- make_scatter_figure(data.in,
                                ref_data,
                                o_vars[2],
                                list_of_tauearly_correlation_dataset[[1]],
                                "Early Tau PET")
ggsave(paste0(out_plots,"EarlyTau_Scatter_ATN.png"),
       plot=plot_out,
       device=png,dpi = 1200,width = 8,height=12)

load("/FNIH_Project_2/Paper1/Spearman_Analysis/results/taulate_correlations.rdata") #load in
results data from Spearman_Analysis.R code
###Create Scatter plot figure for Late Tau results
plot_out <- make_scatter_figure(data.in,
                                ref_data,
                                o_vars[3],
                                list_of_TauLate_correlation_dataset[[1]],
                                "Late Tau PET")
ggsave(paste0(out_plots,"LateTau_Scatter_ATN.png"),
       plot=plot_out,
       device=png,dpi = 1200,width = 8,height=12)

load("/FNIH_Project_2/Paper1/Spearman_Analysis/results/atrophy_correlations.rdata") #load in
results data from Spearman_Analysis.R code
###Create Scatter plot figure for Atrophy/Cortical Thickness
plot_out <- make_scatter_figure(data.in,
                                ref_data,
                                o_vars[4],
                                list_of_Atrophy_correlation_dataset[[1]],
                                "Cortical thickness",rev_axis = T)
ggsave(paste0(out_plots,"Atrophy_Scatter_ATN.png"),
       plot=plot_out,
       device=png,dpi = 1200,width = 8,height=12)

load("/FNIH_Project_2/Paper1/Spearman_Analysis/results/cdrsumbox_correlations.rdata") #load in
results data from Spearman_Analysis.R code
###Create Scatter plot figure for CDR Sum of boxes
plot_out <- make_scatter_figure(data.in,
                                ref_data,
                                o_vars[5],
                                list_of_sumbox_correlation_dataset[[1]],
                                "Dementia severity [CDR Sum of Boxes]")
ggsave(paste0(out_plots,"CDRSumbox_Scatter_ATN.png"),
       plot=plot_out,
       device=png,dpi = 1200,width = 8,height=12)
```

### Code B9. R code for creating ROC plots (requires B10).

```
# FNIH Project 2
# make ROC plots
#### Coded: Kellen Petersen
#### Date as of commenting and documentation - 05/28/2024

# Load libraries
library(tidyverse)
library(data.table)
library(RVAideMemoire)
library(pROC)
library(RColorBrewer)
library(MuMIn)
library(here)
library(cowplot)
library(ggeasy)
library(showtext)
library(ggtext)  # Load the ggtext package
library(ggpubr)  # Load the ggpubr package

#here::i_am("make_roc_plots_CLEAN.R")

# Load make_model file
source(here("make_model_roc_plot_CLEAN.R"))

# Load data
data_file <- here("adni_data","cross_sectional_data_2024_04_30.csv")
data.in00 <- read.csv(data_file)

# Choose group (here: entire cohort)
df <- data.in00

# Parameters: with covariates (True) or not (False)
covars_TF <- FALSE

# Outcome variables
vars_outcome <- c("CENTILOIDS_10")
#vars_outcome <- c("TAU_MesialTemporal_10")
#vars_outcome <- c("atrophy_10")
#vars_outcome <- c("impaired_10")

# Run Make Models
result1 <- make_model(df,covars_TF,vars_outcome)
T1 <- result1$T
M1 <- result1$MODELS_df
REF <- result1$REF

# Re-arrange data frame and choose best performing models
Tbest <- T1 %>%
  group_by(company) %>%
  slice(1) %>%
  ungroup() %>%
  arrange(desc(AUC))

# Make labels
Tbest$analyte2 <- Tbest$analyte
Tbest$analyte2 <- gsub("b", "β", Tbest$analyte2)
Tbest$company <- gsub("AlzPath", "ALZpath", Tbest$company)
Tbest$labels <- paste0(Tbest$company, ": ", Tbest$analyte2)
order <- Tbest$original_order
order <- c(order,26)
labels <- Tbest$labels
labels[which(labels == "covariates: covariates")] <- "Covariates (age, sex, APOE genotype)"

# Make ROC plot list
roc_list <- setNames(
```

```
  object = list(
    M1[[order[1]]][[2]],
    M1[[order[2]]][[2]],
    M1[[order[3]]][[2]],
    M1[[order[4]]][[2]],
    M1[[order[5]]][[2]],
    M1[[order[6]]][[2]],
    M1[[25]][[2]]
  ),
  nm = labels
)

# Colors for the legend
company_colors <- c("blue", "darkgreen", "red", "purple", "darkorange", "brown", "black")
company_names <- c("C2N", "Fujirebio", "ALZpath", "Janssen", "Roche", "Quanterix", "covariates")

# Create a new column 'colors' in your data frame
Tbest$colors <- company_colors[match(Tbest$company, company_names)]
COLS <- Tbest$colors

# Plotting with ggroc and adding a legend
title0 <- "Amyloid PET"
roc_plot <- ggroc(roc_list, linetype = 1, size = 1) +
  geom_abline(intercept = 1, slope = 1, linetype = "dotted", color = "black") +
  scale_color_manual(values = COLS, name = "Best performing models") +
  theme_cowplot(12) +
  coord_fixed() +
  labs(x = "NPA", y = "PPA", title = title0) +
  theme(plot.title = element_text(hjust = 0.5))
roc_plot <- ggpar(roc_plot,
                  legend.text = ggtext::element_markdown(),
                  legend.title = ggtext::element_markdown())

# Save plot
ggsave(here("results_clean","roc_Amyloid_CLEAN.jpg"), roc_plot, device = "jpg", dpi = 500, width =
11, height = 6, units = "in")
```

**Code B10. R code for creating models used in plotting ROC curves.**

```
# FNIH Project 2
# make models for ROC plots
#### Coded: Kellen Petersen
#### Date as of commenting and documentation - 05/28/2024

make_model <- function(data.in0,covars_TF,vars_outcome){
  data.in <- data.in0

  #Covariates
  age_var <- "AGE" #variable with age
  sex_var <- "PTGENDER" #variable with sex
  educ_var <- "PTEDUCAT" #variable with education

  APOE_variable <- "APOE_genotype" #Variable with APOE genotypes
  data.in$APOE_Cat <- factor(data.in[[APOE_variable]], levels = c("33", "34", "24", "22", "23",
"44"))
  list_covariates <- list(c(age_var, sex_var, "APOE_Cat"))

  if (covars_TF) {
    vars_covariates <- list_covariates
  } else {
    vars_covariates <- NULL
  }

  # Assays Lists for each platform
  assays_c2n       <- c("C2N_plasma_Abeta42_Abeta40", "C2N_plasma_ptau217",
"C2N_plasma_ptau217_ratio")
  assays_fuji      <- c("Fuji_plasma_Ab42_Ab40","Fuji_plasma_ptau217")
  assays_alzpath   <- c("AlzPath_plasma_ptau217")
  assays_janssen   <- c("Janssen_plasma_ptau217")
  assays_roche     <-
c("Roche_plasma_Ab42_Ab40","Roche_plasma_ptau181","Roche_plasma_NfL","Roche_plasma_GFAP")
  assays_quanterix <- c("QX_plasma_Ab42_Ab40","QX_plasma_ptau181","QX_plasma_NfL","QX_plasma_GFAP")
  assays_csfratio  <- c("PTAU_over_ABETA42")

  # Create list of model indices
  combos_c2n       <- list(assays_c2n[c(3,1)],assays_c2n[c(2,1)],
                           assays_c2n[c(3)],assays_c2n[c(2)],assays_c2n[c(1)])            # 5
models
  combos_fuji      <- list(assays_fuji[c(2,1)],assays_fuji[c(2)],assays_fuji[c(1)])        # 3
models
  combos_alzath    <- list(assays_alzpath)                                                # 1
model
  combos_janssen   <- list(assays_janssen)                                                # 1
model
  combos_roche     <- list(assays_roche[c(2,1,4,3)],assays_roche[c(2,1,3)],
                           assays_roche[c(2,1)],
                           assays_roche[c(2)],assays_roche[c(1)],
                           assays_roche[c(4)],assays_roche[c(3)])                         # 7
models
  combos_quanterix <- list(assays_quanterix[c(2,1,4,3)],assays_quanterix[c(2,1,3)],
                           assays_quanterix[c(2,1)],
                           assays_quanterix[c(2)],assays_quanterix[c(1)],
                           assays_quanterix[c(4)],assays_quanterix[c(3)])                 # 7
models
  combos_csfratio   <- list(assays_csfratio)                                              # 1
model

  combos_all <- list()
  combos_all[[1]] <- combos_c2n[1]
  combos_all[[2]] <- combos_c2n[2]
  combos_all[[3]] <- combos_c2n[3]
  combos_all[[4]] <- combos_c2n[4]
  combos_all[[5]] <- combos_c2n[5]
```

```
combos_all[[6]] <- combos_fuji[1]
combos_all[[7]] <- combos_fuji[2]
combos_all[[8]] <- combos_fuji[3]

combos_all[[9]] <- combos_alzath[1]

combos_all[[10]] <- combos_janssen[1]

combos_all[[11]] <- combos_roche[1]
combos_all[[12]] <- combos_roche[2]
combos_all[[13]] <- combos_roche[3]
combos_all[[14]] <- combos_roche[4]
combos_all[[15]] <- combos_roche[5]
combos_all[[16]] <- combos_roche[6]
combos_all[[17]] <- combos_roche[7]

combos_all[[18]] <- combos_quanterix[1]
combos_all[[19]] <- combos_quanterix[2]
combos_all[[20]] <- combos_quanterix[3]
combos_all[[21]] <- combos_quanterix[4]
combos_all[[22]] <- combos_quanterix[5]
combos_all[[23]] <- combos_quanterix[6]
combos_all[[24]] <- combos_quanterix[7]

Combos <- combos_all

# Company label
C_c2n <- rep("C2N", 5)
C_fuji <- rep("Fujirebio", 3)
C_alzpath <- rep("AlzPath", 1)
C_janssen <- rep("Janssen", 1)
C_roche <- rep("Roche", 7)
C_5csfratio <- rep("CSF", 1)
C_quanterix <- rep("Quanterix", 7)
C <- c(C_c2n, C_fuji, C_alzpath, C_janssen, C_roche, C_quanterix)
CC <- C

# Analyte label
A_c2n <- c("p-tau217 ratio + Ab42/Ab40","p-tau217 + Ab42/Ab40",
           "p-tau217 ratio","p-tau217","Ab42/Ab40")
A_fuji <- c("p-tau217 + Ab42/Ab40", "p-tau217", "Ab42/Ab40")
A_alzpath <- c("p-tau217")
A_janssen <- c("p-tau217")
A_roche <- c("p-tau181 + Ab42/Ab40 + GFAP + NfL",
             "p-tau181 + Ab42/Ab40 + NfL",
             "p-tau181 + Ab42/Ab40",
             "p-tau181","Ab42/Ab40",
             "GFAP","NfL")
A_5csfratio <- c("CSF PTAU/AB42")
A_quanterix <- c("p-tau181 + Ab42/Ab40 + GFAP + NfL",
                 "p-tau181 + Ab42/Ab40 + NfL",
                 "p-tau181 + Ab42/Ab40",
                 "p-tau181","Ab42/Ab40",
                 "GFAP","NfL")
A_qcsfratio <- c("CSF PTAU/AB42")

A <- c(A_c2n, A_fuji, A_alzpath, A_janssen, A_roche, A_quanterix)
AA <- A

# List of single analytes
single_analytes <- c("p-tau217 ratio",
                     "Ab42/Ab40",
                     "p-tau217",
                     "p-tau181",
                     "GFAP",
                     "NfL",
                     "CSF PTAU/AB42")
```

```
  # Run the models
  OUT_df <- list()
  MODELS_df <- list()
  k <- 0
  data.in <- data.in
  # Loop through the combinations of predictors
  for (i_outcome in 1:length(vars_outcome)) {
    for (i_assay_combos in 1:length(Combos)) {

      outcome <- vars_outcome[i_outcome]
      pred1 <- Combos[[i_assay_combos]]
      pred2 <- vars_covariates
      preds <- c(unlist(pred1), unlist(pred2))

      formula_str <- paste(outcome, " ~ ", paste(preds, collapse = " + "))
      model_formula <- as.formula(formula_str)

      temp_data <- data.in[complete.cases(data.in[,preds]), c(outcome,preds)] %>%
        na.omit()

      mdl_logistic <- glm(model_formula, data = temp_data, family = "binomial")
      mdl_pred <- predict(mdl_logistic, data = temp_data, type = "response")
      brierScore <- mean((mdl_pred-temp_data[[outcome]])^2)/dim(temp_data)[1]

      roc_tmp <- pROC::roc(temp_data[[outcome]],mdl_pred,plot=FALSE,print.auc=FALSE) #this will get
the ROC output

      # YOUDEN INDEX
      youden_index <- pROC::coords(roc_tmp, "best", ret="all")
      optimal_cutoff <- youden_index$threshold

      T <- data.frame(roc_tmp$thresholds, roc_tmp$sensitivities, roc_tmp$specificities)
      auc_tmp <- pROC::auc(roc_tmp)
      ci_AUC  <- ci.auc(roc_tmp) #get confidence interview

      sensitivity <- youden_index$sensitivity
      specificity <- youden_index$specificity

      # Sensitivity = 90%
      II_sensX <- which(roc_tmp$sensitivities >= .90)
      I_sensX <- max(II_sensX)
      sensX_threshold <- roc_tmp$thresholds[I_sensX]
      sensX_specificity <- roc_tmp$specificities[I_sensX]

      # Specificity = 90%
      II_specX <- which(roc_tmp$specificities >= .90)
      I_specX <- min(II_specX)
      specX_threshold <- roc_tmp$thresholds[I_specX]
      specX_sensitivities <- roc_tmp$sensitivities[I_specX]

      num_intermediate <- sum( (mdl_pred > sensX_threshold) & (mdl_pred < specX_threshold) )
      num_intermediate

      out_v <- c()
      out_v[1] <- CC[i_assay_combos] # company label
      out_v[2] <- round(roc_tmp$auc,3) # AUC rounded to 2 decimals
      out_v[3] <- round(ci_AUC[1],3) # AUC lower bound
      out_v[4] <- round(ci_AUC[3],3) # AUC upper bound
      out_v[5] <- paste0(formatC(sensitivity, digits=3, format="fg"))
      out_v[6] <- paste0(formatC(specificity, digits=3, format="fg"))
      out_v[7] <- paste0(formatC(youden_index$ppv, digits=3, format="fg"))
      out_v[8] <- paste0(formatC(youden_index$npv, digits=3, format="fg"))
      out_v[9] <- paste0(formatC(youden_index$accuracy, digits=3, format="fg"))
```

```
      tmp <- formatC((log(optimal_cutoff / (1 - optimal_cutoff)) - mdl_logistic$coefficients[1]) /
mdl_logistic$coefficients[2], digits=4, format="fg")
      if (AA[i_assay_combos] %in% single_analytes) {out_v[10] <- tmp}
      else {out_v[10] <- -99}

      out_v[11] <- AA[i_assay_combos] # analyte label

      out_v[12] <- formatC(sensX_threshold, digits=3, format="fg")
      out_v[13] <- formatC(sensX_specificity, digits=3, format="fg")

      out_v[14] <- formatC(specX_threshold, digits=3, format="fg")
      out_v[15] <- formatC(specX_sensitivities, digits=3, format="fg")

      out_v[16] <- num_intermediate

      out_v[17] <- formula_str
      out_v[18] <- outcome
      out_v[19] <- paste(pred1, collapse = " + ")
      out_v[20] <- covars_TF
      dd <- dim(temp_data)
      out_v[21] <- dd[1]
      out_v[22] <- brierScore

      model <- list()
      model[[1]] <- mdl_logistic
      model[[2]] <- roc_tmp

      k <- i_assay_combos + length(combos_all)*(i_outcome-1)

      OUT_df[[k]] <- out_v
      MODELS_df[[k]] <- model
    }

    # Run a single time for the covariates only
    outcome <- vars_outcome[i_outcome]
    pred2 <- vars_covariates
    preds <- c(unlist(list_covariates))

    formula_str <- paste(outcome, " ~ ", paste(preds, collapse = " + "))
    model_formula <- as.formula(formula_str)

    temp_data <- data.in[complete.cases(data.in[,preds]), c(outcome,preds)] %>%
      na.omit()

    mdl_logistic <- glm(model_formula, data = temp_data, family = "binomial")
    mdl_pred <- predict(mdl_logistic, data = temp_data, type = "response")
    brierScore <- mean((mdl_pred-temp_data[[outcome]])^2)

    roc_tmp <- pROC::roc(temp_data[[outcome]],mdl_pred,plot=FALSE,print.auc=FALSE) #this will get
the ROC output

    youden_index <- pROC::coords(roc_tmp, "best", ret="all")
    optimal_cutoff <- youden_index$threshold

    T <- data.frame(roc_tmp$thresholds, roc_tmp$sensitivities, roc_tmp$specificities)
    auc_tmp <- pROC::auc(roc_tmp)
    ci_AUC  <- ci.auc(roc_tmp) #get confidence intervals

    sensitivity <- youden_index$sensitivity
    specificity <- youden_index$specificity

    # Sensitivity = 95%
    II_sensX <- which(roc_tmp$sensitivities >= .90)
    I_sensX <- max(II_sensX)
    sensX_threshold <- roc_tmp$thresholds[I_sensX]
    sensX_specificity <- roc_tmp$specificities[I_sensX]
```

```
  # Specificity = 95%
  II_specX <- which(roc_tmp$specificities >= .90)
  I_specX <- min(II_specX)
  specX_threshold <- roc_tmp$thresholds[I_specX]
  specX_sensitivities <- roc_tmp$sensitivities[I_specX]

  num_intermediate <- sum( (mdl_pred > sensX_threshold) & (mdl_pred < specX_threshold) )
  num_intermediate

  out_v <- c()
  out_v[1] <- "covariates" # company label
  out_v[2] <- round(roc_tmp$auc,3) # AUC rounded to 2 decimals
  out_v[3] <- round(ci_AUC[1],3) # AUC lower bound
  out_v[4] <- round(ci_AUC[3],3) # AUC upper bound
  out_v[5] <- paste0(formatC(sensitivity, digits=3, format="fg"))
  out_v[6] <- paste0(formatC(specificity, digits=3, format="fg"))
  out_v[7] <- paste0(formatC(youden_index$ppv, digits=3, format="fg"))
  out_v[8] <- paste0(formatC(youden_index$npv, digits=3, format="fg"))
  out_v[9] <- paste0(formatC(youden_index$accuracy, digits=3, format="fg"))

  out_v[10] <- -99

  out_v[11] <- "covariates" # analyte label

  out_v[12] <- formatC(sensX_threshold, digits=3, format="fg")
  out_v[13] <- formatC(sensX_specificity, digits=3, format="fg")

  out_v[14] <- formatC(specX_threshold, digits=3, format="fg")
  out_v[15] <- formatC(specX_sensitivities, digits=3, format="fg")

  out_v[16] <- num_intermediate

  out_v[17] <- formula_str
  out_v[18] <- outcome
  out_v[19] <- " "
  out_v[20] <- covars_TF
  out_v[21] <- dd[1]
  out_v[22] <- brierScore

  model <- list()
  model[[1]] <- mdl_logistic
  model[[2]] <- roc_tmp

  k <- 1 + length(combos_all) #+ length(combos_all)*(i_outcome-1)

  OUT_df[[k]] <- out_v
  MODELS_df[[k]] <- model
}

# Make output into dataframe
df_out <- t(data.frame(OUT_df))
df_out <- data.frame(df_out)
rownames(df_out) <- NULL
colnames(df_out) <- c("company","AUC","AUC_Lower","AUC_Upper",
                    "sensitivity","specificity","ppv","npv","accuracy",
                    "cutoff_value","analyte",
                    "90% Sens Threshold","90% Sens - Specificity",
                    "90% Spec Threshold","90% Spec - Sensitivity",
                    "Num_Intermediate","formula","outcome","predictors",
                    "covars_TF","n_cases","Brier_Score")

df_out$AUC <- as.numeric(df_out$AUC)
df_out$AUC_Lower <- as.numeric(df_out$AUC_Lower)
df_out$AUC_Upper <- as.numeric(df_out$AUC_Upper)
df_out$Num_Intermediate <- as.numeric(df_out$Num_Intermediate)
df_out$n_cases <- as.numeric(df_out$n_cases)
df_out <- df_out %>%
```

```
    mutate(intermediate_results = paste(Num_Intermediate, " (",
round(Num_Intermediate/n_cases,2)*100, "%)", sep = ""))

  DF <- df_out
  DFB <- DF %>%
    mutate(index_0 = row_number()) %>%
    filter(outcome == vars_outcome[1] ) %>%
    arrange(company,AUC) %>%
    group_by(company) %>%
    mutate(index = row_number())
  DFB$company <- factor(DFB$company, levels = c("C2N", "Fujirebio", "AlzPath","Janssen", "Roche",
"Quanterix","CSF","covariates"))
  DFB <- DFB %>%
    mutate(companyNums = as.numeric(company)) %>%
    arrange(companyNums,desc(AUC)) %>%
    ungroup() %>%
    mutate(plot_index = desc(row_number()))
  DFB$analyte <- factor(DFB$analyte,
                        levels = c("p-tau217 ratio + Ab42/Ab40",
                                   "p-tau217 ratio",
                                   "Ab42/Ab40",
                                   "p-tau217 + Ab42/Ab40",
                                   "p-tau217",
                                   "p-tau181 + Ab42/Ab40 + GFAP + NfL",
                                   "p-tau181 + Ab42/Ab40 + NfL",
                                   "p-tau181 + Ab42/Ab40",
                                   "p-tau181",
                                   "GFAP", "NfL",
                                   "CSF PTAU/AB42",
                                   "covariates"))
  DFB2 <- DFB %>%
    mutate(AUC_CI = paste0(format(AUC,nsmall=3)," (",format(AUC_Lower,nsmall=3), "-
",format(AUC_Upper,nsmall=3),")")) %>%
    select("company","analyte",
           "AUC_CI",
           "sensitivity","specificity",
           "90% Sens Threshold","90% Sens - Specificity",
           "90% Spec Threshold","90% Spec - Sensitivity",
           "intermediate_results")

  # Final output dataframe T
  T <- DFB %>%
    select(company,AUC,AUC_Lower,AUC_Upper,analyte,Brier_Score) %>%
    mutate(original_order = row_number()) %>%
    group_by(company) %>%
    mutate(max_AUC = max(AUC)) %>%
    ungroup() %>%
    arrange(desc(max_AUC), company, desc(AUC)) %>%
    select(-max_AUC) %>%
    filter(company != "CSF")

  REF <- data.frame(company = C, analyte = A) %>%
    mutate(index = row_number()) %>%
    filter(company != "CSF")

  return(list(T = T, MODELS_df = MODELS_df, REF = REF))
}
```

**Code B11. R code for obtaining AUC values for covariate-only models.**

```
# FNIH Project 2
# get AUCs for covariates-only models
#### Coded: Kellen Petersen
#### Date as of commenting and documentation - 05/28/2024

# Load libraries
library(tidyverse)
library(pROC)
library(here)

#here::i_am("get_AUC_covariatesOnly.R")

data_file <- here("adni_data","cross_sectional_data_CLEAN.csv")
data.in00 <- read.csv(data_file)

# Define binary variable for cognitive impairment
data.in00$impaired_10 <- ifelse(data.in00$CDR > 0, 1, 0)

# Define cohort
data.in0 <- data.in00
# data.in0 <- data.in00 %>% filter(CDR == 0)
# data.in0 <- data.in00 %>% filter(CDR > 0)
# data.in0 <- data.in00 %>% filter(CENTILOIDS_10 == 0)
# data.in0 <- data.in00 %>% filter(CENTILOIDS_10 == 1)
# data.in0 <- data.in00 %>% filter(CENTILOIDS_10_sens == 0)
# data.in0 <- data.in00 %>% filter(CENTILOIDS_10_sens == 1)

# Parameters for covariates (included or not?)
covars_TF <- TRUE

# Outcome variables
vars_outcome <- c("CENTILOIDS_10")
#vars_outcome <- c("CENTILOIDS_10_sens")
#vars_outcome <- c("TAU_MesialTemporal_10")
#vars_outcome <- c("TAU_TemporoParietal_10")
#vars_outcome <- c("atrophy_10")
#vars_outcome <- c("impaired_10")

#Covariates
age_var <- "AGE" #variable with age
sex_var <- "PTGENDER" #variable with sex
educ_var <- "PTEDUCAT" #variable with education

APOE_variable <- "APOE_genotype" #Variable with APOE genotypes
data.in0$APOE_Cat <- factor(data.in0[[APOE_variable]], levels = c("33", "34", "24", "22", "23",
"44"))
list_covariates <- list(c(age_var, sex_var, "APOE_Cat"))
if (covars_TF) {
  vars_covariates <- list_covariates
} else {
  vars_covariates <- NULL
}

# Dataset that goes into the model
data.in <- data.in0

# Make model formula
outcome <- vars_outcome
pred1 <- NULL
pred2 <- vars_covariates
preds <- c(unlist(pred1), unlist(pred2))
formula_str <- paste(outcome, " ~ ", paste(preds, collapse = " + "))
model_formula <- as.formula(formula_str)

# Make relevant dataset
```

```
temp_data <- data.in[complete.cases(data.in[,preds]), c(outcome,preds)] %>%
  na.omit()

# Fit logistic regression model
mdl_logistic <- glm(model_formula, data = temp_data, family = "binomial")
mdl_pred <- predict(mdl_logistic, data = temp_data, type = "response")

# ROC and AUC
roc_tmp <- pROC::roc(temp_data[[outcome]],mdl_pred,plot=FALSE,print.auc=FALSE) #this will get the
ROC output
auc_tmp <- pROC::auc(roc_tmp)
ci_AUC  <- ci.auc(roc_tmp)

# Output
out_v <- c()
out_v[1] <- round(roc_tmp$auc,3) # AUC rounded to 3 decimals
out_v[2] <- round(ci_AUC[1],3) # AUC lower bound
out_v[3] <- round(ci_AUC[3],3) # AUC upper bound

outp <- paste0("AUC (CI): ", format(out_v[1],nsmall=3)," (",format(out_v[2],nsmall=3), "-
",format(out_v[3],nsmall=3),")")
print(outp)
```

**Code B12. R code for creating AUC tables.**

```
# FNIH Project 2
# make AUC tables
#### Coded: Kellen Petersen
#### Date as of commenting and documentation - 05/28/2024

# Load libraries
library(tidyverse)
library(data.table)
library(RVAideMemoire)
library(pROC)
library(RColorBrewer)
library(MuMIn)
library(here)
library(cowplot)
library(ggeasy)

#here::i_am("make_AUC_tables_2_CLEAN.R")

# Load data
data_file <- here("adni_data","cross_sectional_data_CLEAN.csv")
data.in00 <- read.csv(data_file)

# Define output folder
out_folder <- "results_clean"

# Choose outcome
info1 <- "outcomeCENT20"
#info1 <- "outcomeCENT37"
#info1 <- "outcomeEarlyTau"
#info1 <- "outcomeLateTau"
#info1 <- "outcomeAtrophy"
#info1 <- "outcomeImpairment"

# Choose cohort
info2 <- "cohortAll"
#info2 <- "cohortCU"
#info2 <- "cohortCI"
#info2 <- "cohortAB20n"
#info2 <- "cohortAB20p"
#info2 <- "cohortAB37n"
#info2 <- "cohortAB37p"

# Make output file names
outfile_singlecutoff <- paste0(info1,"_",info2,"_singleCutoff",".csv")
outfile_AUCs <- paste0(info1,"_",info2,"_AUCs",".csv")
outfile_twoCuts <- paste0(info1,"_",info2,"_twoCuts",".csv")

# Make cohort dataframe
data.in0 <- data.in00
#data.in0 <- data.in00 %>% filter(CDR == 0)
#data.in0 <- data.in00 %>% filter(CDR > 0)
#data.in0 <- data.in00 %>% filter(CENTILOIDS_10 == 0)
#data.in0 <- data.in00 %>% filter(CENTILOIDS_10 == 1)
#data.in0 <- data.in00 %>% filter(CENTILOIDS_10_sens == 0)
#data.in0 <- data.in00 %>% filter(CENTILOIDS_10_sens == 1)

# Define binary variable for cognitive impairment
data.in0$impaired_10 <- ifelse(data.in0$CDR > 0, 1, 0)

# Rename dataframe
data.in.cohort <- data.in0

# Parameters (covariates included?)
covars_TF <- FALSE
```

```
# Outcome variables
vars_outcome <- c("CENTILOIDS_10")
#vars_outcome <- c("CENTILOIDS_10_sens")
#vars_outcome <- c("TAU_MesialTemporal_10")
#vars_outcome <- c("TAU_TemporoParietal_10")
#vars_outcome <- c("atrophy_10")
#vars_outcome <- c("impaired_10")

#Covariates
age_var <- "AGE" #variable with age
sex_var <- "PTGENDER" #variable with sex
educ_var <- "PTEDUCAT" #variable with education
APOE_variable <- "APOE_genotype" #Variable with APOE genotypes
data.in.cohort$APOE_Cat <- factor(data.in.cohort[[APOE_variable]],
                                  levels = c("33", "34", "24", "22", "23", "44"))
list_covariates <- list(c(age_var, sex_var, "APOE_Cat"))
if (covars_TF) {
  vars_covariates <- list_covariates
} else {
  vars_covariates <- NULL
}

# Assays Lists
assays_c2n       <- c("C2N_plasma_Abeta42_Abeta40", "C2N_plasma_ptau217",
"C2N_plasma_ptau217_ratio")
assays_fuji      <- c("Fuji_plasma_Ab42_Ab40","Fuji_plasma_ptau217")
assays_alzpath   <- c("AlzPath_plasma_ptau217")
assays_janssen   <- c("Janssen_plasma_ptau217")
assays_roche     <-
c("Roche_plasma_Ab42_Ab40","Roche_plasma_ptau181","Roche_plasma_NfL","Roche_plasma_GFAP")
assays_quanterix <- c("QX_plasma_Ab42_Ab40","QX_plasma_ptau181","QX_plasma_NfL","QX_plasma_GFAP")
assays_csfratio  <- c("PTAU_over_ABETA42")

# Create list of model indices
combos_c2n       <- list(assays_c2n[c(3,1)],assays_c2n[c(2,1)],
                         assays_c2n[c(3)],assays_c2n[c(2)],assays_c2n[c(1)])              # 5
models
combos_fuji      <- list(assays_fuji[c(2,1)],assays_fuji[c(2)],assays_fuji[c(1)])         # 3
models
combos_alzath    <- list(assays_alzpath) # 1 model
combos_janssen   <- list(assays_janssen) # 1 model
combos_roche     <- list(assays_roche[c(2,1,4,3)],assays_roche[c(2,1,3)],
                         assays_roche[c(2,1)],
                         assays_roche[c(2)],assays_roche[c(1)],
                         assays_roche[c(4)],assays_roche[c(3)]) # 7 models
combos_quanterix <- list(assays_quanterix[c(2,1,4,3)],assays_quanterix[c(2,1,3)],
                         assays_quanterix[c(2,1)],
                         assays_quanterix[c(2)],assays_quanterix[c(1)],
                         assays_quanterix[c(4)],assays_quanterix[c(3)]) # 7 models
combos_csfratio   <- list(assays_csfratio) # 1 model

combos_all <- list()
combos_all[[1]] <- combos_c2n[1]
combos_all[[2]] <- combos_c2n[2]
combos_all[[3]] <- combos_c2n[3]
combos_all[[4]] <- combos_c2n[4]
combos_all[[5]] <- combos_c2n[5]

combos_all[[6]] <- combos_fuji[1]
combos_all[[7]] <- combos_fuji[2]
combos_all[[8]] <- combos_fuji[3]

combos_all[[9]] <- combos_alzath[1]

combos_all[[10]] <- combos_janssen[1]
```

```
combos_all[[11]] <- combos_roche[1]
combos_all[[12]] <- combos_roche[2]
combos_all[[13]] <- combos_roche[3]
combos_all[[14]] <- combos_roche[4]
combos_all[[15]] <- combos_roche[5]
combos_all[[16]] <- combos_roche[6]
combos_all[[17]] <- combos_roche[7]

combos_all[[18]] <- combos_quanterix[1]
combos_all[[19]] <- combos_quanterix[2]
combos_all[[20]] <- combos_quanterix[3]
combos_all[[21]] <- combos_quanterix[4]
combos_all[[22]] <- combos_quanterix[5]
combos_all[[23]] <- combos_quanterix[6]
combos_all[[24]] <- combos_quanterix[7]

Combos <- combos_all

# Company label
C_c2n <- rep("C2N", 5)
C_fuji <- rep("Fujirebio", 3)
C_alzpath <- rep("AlzPath", 1)
C_janssen <- rep("Janssen", 1)
C_roche <- rep("Roche", 7)
C_5csfratio <- rep("CSF", 1)
C_quanterix <- rep("Quanterix", 7)
C <- c(C_c2n, C_fuji, C_alzpath, C_janssen, C_roche, C_quanterix)
CC <- C

# Analyte label
A_c2n <- c("p-tau217 ratio + Ab42/Ab40","p-tau217 + Ab42/Ab40",
           "p-tau217 ratio","p-tau217","Ab42/Ab40")
A_fuji <- c("p-tau217 + Ab42/Ab40", "p-tau217", "Ab42/Ab40")
A_alzpath <- c("p-tau217")
A_janssen <- c("p-tau217")
A_roche <- c("p-tau181 + Ab42/Ab40 + GFAP + NfL",
             "p-tau181 + Ab42/Ab40 + NfL",
             "p-tau181 + Ab42/Ab40",
             "p-tau181","Ab42/Ab40",
             "GFAP","NfL")
A_quanterix <- c("p-tau181 + Ab42/Ab40 + GFAP + NfL",
                 "p-tau181 + Ab42/Ab40 + NfL",
                 "p-tau181 + Ab42/Ab40",
                 "p-tau181","Ab42/Ab40",
                 "GFAP","NfL")
A <- c(A_c2n, A_fuji, A_alzpath, A_janssen, A_roche, A_quanterix)
AA <- A

# List of single analytes
single_analytes <- c("p-tau217 ratio",
                     "Ab42/Ab40",
                     "p-tau217",
                     "p-tau181",
                     "GFAP",
                     "NfL",
                     "CSF PTAU/AB42")

OUT_df <- list()
MODELS_df <- list()
k <- 0
data.in <- data.in.cohort

# Loop through the combinations of predictors
for (i_outcome in 1:length(vars_outcome)) {
  for (i_assay_combos in 1:length(Combos)) {

    outcome <- vars_outcome[i_outcome]
```

```
    pred1 <- Combos[[i_assay_combos]]
    pred2 <- vars_covariates
    preds <- c(unlist(pred1), unlist(pred2))

    formula_str <- paste(outcome, " ~ ", paste(preds, collapse = " + "))
    model_formula <- as.formula(formula_str)

    temp_data <- data.in[complete.cases(data.in[,preds]), c(outcome,preds)] %>%
      na.omit()

    mdl_logistic <- glm(model_formula, data = temp_data, family = "binomial")
    mdl_pred <- predict(mdl_logistic, data = temp_data, type = "response")

    roc_tmp <- pROC::roc(temp_data[[outcome]],mdl_pred,plot=FALSE,print.auc=FALSE
    youden_index <- coords(roc_tmp, "best", ret="all")
    optimal_cutoff <- youden_index$threshold

    T <- data.frame(roc_tmp$thresholds, roc_tmp$sensitivities, roc_tmp$specificities)
    auc_tmp <- pROC::auc(roc_tmp)
    ci_AUC  <- ci.auc(roc_tmp) #get confidence interval

    sensitivity <- youden_index$sensitivity
    specificity <- youden_index$specificity

    # Sensitivity = 90%
    II_sensX <- which(roc_tmp$sensitivities >= .90)
    I_sensX <- max(II_sensX)
    sensX_threshold <- roc_tmp$thresholds[I_sensX]
    sensX_specificity <- roc_tmp$specificities[I_sensX]

    # Specificity = 90%
    II_specX <- which(roc_tmp$specificities >= .90)
    I_specX <- min(II_specX)
    specX_threshold <- roc_tmp$thresholds[I_specX]
    specX_sensitivities <- roc_tmp$sensitivities[I_specX]

    num_intermediate <- sum( (mdl_pred > sensX_threshold) & (mdl_pred < specX_threshold) )
    num_intermediate

    out_v <- c()
    out_v[1] <- CC[i_assay_combos] # company label
    out_v[2] <- round(roc_tmp$auc,3) # AUC rounded to 3 decimals
    out_v[3] <- round(ci_AUC[1],3) # AUC lower bound
    out_v[4] <- round(ci_AUC[3],3) # AUC upper bound
    out_v[5] <- paste0(formatC(sensitivity, digits=3, format="fg"))
    out_v[6] <- paste0(formatC(specificity, digits=3, format="fg"))
    out_v[7] <- paste0(formatC(youden_index$ppv, digits=3, format="fg"))
    out_v[8] <- paste0(formatC(youden_index$npv, digits=3, format="fg"))
    out_v[9] <- paste0(formatC(youden_index$accuracy, digits=3, format="fg"))

    tmp <- formatC((log(optimal_cutoff / (1 - optimal_cutoff)) - mdl_logistic$coefficients[1]) /
mdl_logistic$coefficients[2], digits=4, format="fg")
    if (AA[i_assay_combos] %in% single_analytes) {out_v[10] <- tmp}
    else {out_v[10] <- -99}

    out_v[11] <- AA[i_assay_combos] # analyte label

    out_v[12] <- formatC(sensX_threshold, digits=3, format="fg")
    out_v[13] <- formatC(sensX_specificity, digits=3, format="fg")

    out_v[14] <- formatC(specX_threshold, digits=3, format="fg")
    out_v[15] <- formatC(specX_sensitivities, digits=3, format="fg")

    out_v[16] <- num_intermediate

    out_v[17] <- formula_str
    out_v[18] <- outcome
```

```
    out_v[19] <- paste(pred1, collapse = " + ")
    out_v[20] <- covars_TF

    dd <- dim(temp_data)
    out_v[21] <- dd[1]

    brierScore <- mean((mdl_pred-temp_data[[outcome]])^2)
    out_v[22] <- brierScore

    model <- list()
    model[[1]] <- mdl_logistic
    model[[2]] <- roc_tmp

    k <- i_assay_combos + length(combos_all)*(i_outcome-1)
    OUT_df[[k]] <- out_v
    MODELS_df[[k]] <- model
  }
}

################################################################################

df_out <- t(data.frame(OUT_df))
df_out <- data.frame(df_out)
rownames(df_out) <- NULL
colnames(df_out) <- c("company","AUC","AUC_Lower","AUC_Upper",
                      "sensitivity","specificity","ppv","npv","accuracy",
                      "cutoff_value","analyte",
                      "90% Sens Threshold","90% Sens - Specificity",
                      "90% Spec Threshold","90% Spec - Sensitivity",
                      "Num_Intermediate","formula","outcome","predictors",
                      "covars_TF","n_cases","Brier_Score")

df_out$AUC <- as.numeric(df_out$AUC)
df_out$AUC_Lower <- as.numeric(df_out$AUC_Lower)
df_out$AUC_Upper <- as.numeric(df_out$AUC_Upper)
df_out$Num_Intermediate <- as.numeric(df_out$Num_Intermediate)
df_out$n_cases <- as.numeric(df_out$n_cases)
df_out <- df_out %>%
  mutate(intermediate_results = paste(Num_Intermediate, " (",
round(Num_Intermediate/n_cases,2)*100, "%)", sep = ""))
DF <- df_out

DFB <- DF %>%
  mutate(index_0 = row_number()) %>%
  filter(outcome == vars_outcome[1] ) %>%
  arrange(company,AUC) %>%
  group_by(company) %>%
  mutate(index = row_number())
DFB$company <- factor(DFB$company, levels = c("C2N", "Fujirebio", "AlzPath","Janssen", "Roche",
"Quanterix","CSF"))

DFB <- DFB %>%
  mutate(companyNums = as.numeric(company)) %>%
  arrange(companyNums,desc(AUC)) %>%
  ungroup() %>%
  mutate(plot_index = desc(row_number()))

DFB$analyte <- factor(DFB$analyte,
                      levels = c("p-tau217 ratio + Ab42/Ab40",
                                 "p-tau217 ratio",
                                 "Ab42/Ab40",
                                 "p-tau217 + Ab42/Ab40",
                                 "p-tau217",
                                 "p-tau181 + Ab42/Ab40 + GFAP + NfL",
                                 "p-tau181 + Ab42/Ab40 + NfL",
                                 "p-tau181 + Ab42/Ab40",
                                 "p-tau181",
```

```
                                 "GFAP", "NfL",
                                 "CSF PTAU/AB42"))

DFB2 <- DFB %>%
  mutate(AUC_CI = paste0(format(AUC,nsmall=3)," (",format(AUC_Lower,nsmall=3), "-
",format(AUC_Upper,nsmall=3),")")) %>%
  select("company","analyte",
         "AUC_CI",
         "sensitivity","specificity",
         "90% Sens Threshold","90% Sens - Specificity",
         "90% Spec Threshold","90% Spec - Sensitivity",
         "intermediate_results")

################################################################################
#  Single analyte table
################################################################################
T1 <- DFB %>%
  mutate(AUC_CI = paste0(formatC(AUC, digits=3, format="fg")," (",
                       formatC(AUC_Lower, digits=3, format="fg"), "-",
                       format(AUC_Upper, digits=3, format="fg"),")")) %>%
  select("company","analyte","AUC",
         "AUC_CI","cutoff_value",
         "sensitivity","specificity","accuracy","Brier_Score","ppv","npv") %>%
  filter(analyte %in% single_analytes) %>%
  rename("Platform" = "company",
         "Analyte" = "analyte",
         "AUCraw" = "AUC",
         "AUC" = "AUC_CI",
         "Cutoff Value" = "cutoff_value",
         "Sensitivity" = "sensitivity",
         "Specificity" = "specificity",
         "Accuracy" = "accuracy",
         "Brier Score" = "Brier_Score",
         "PPV" = "ppv",
         "NPV" = "npv",) %>%
  filter(Platform != "CSF")

T2 <- T1 %>%
  group_by(Platform) %>%
  mutate(max_AUCraw = max(AUCraw)) %>%
  ungroup() %>%
  arrange(desc(max_AUCraw), Platform, desc(AUCraw)) %>%
  select(-max_AUCraw,-AUCraw)

write.csv(T2, file = here(out_folder,outfile_singlecutoff), row.names = FALSE)

################################################################################
################## Make AUC Table: with & w/o covariates  ######################
################################################################################

AUCunadj <- DFB %>%
  mutate(AUC_CI = paste0(format(AUC,nsmall=3)," (",format(AUC_Lower,nsmall=3), "-
",format(AUC_Upper,nsmall=3),")")) %>%
  select("company","analyte","AUC_CI","AUC") %>%
  group_by(company) %>%
  mutate(max_AUC = max(AUC)) %>%
  ungroup() %>%
  arrange(desc(max_AUC),company,desc(AUC))%>%
  select(-max_AUC,-AUC) %>%
  rename("AUC (unadjusted)" = "AUC_CI")

covars_TF <- TRUE
if (covars_TF) {
  vars_covariates <- list_covariates
} else {
  vars_covariates <- NULL
```

```
}

OUT_5adj <- list()
MODELS_5adj <- list()
k <- 0
data.in <- data.in.cohort
# Loop through the combinations of predictors
for (i_outcome in 1:length(vars_outcome)) {
  for (i_assay_combos in 1:length(Combos)) {

    outcome <- vars_outcome[i_outcome]
    pred1 <- Combos[[i_assay_combos]]
    pred2 <- vars_covariates
    preds <- c(unlist(pred1), unlist(pred2))

    formula_str <- paste(outcome, " ~ ", paste(preds, collapse = " + "))
    model_formula <- as.formula(formula_str)

    temp_data <- data.in[complete.cases(data.in[,preds]), c(outcome,preds)] %>%
      na.omit()

    mdl_logistic <- glm(model_formula, data = temp_data, family = "binomial")
    mdl_pred <- predict(mdl_logistic, data = temp_data, type = "response")

    roc_tmp <- pROC::roc(temp_data[[outcome]],mdl_pred,plot=FALSE,print.auc=FALSE)
    youden_index <- coords(roc_tmp, "best", ret="all")
    optimal_cutoff <- youden_index$threshold

    T <- data.frame(roc_tmp$thresholds, roc_tmp$sensitivities, roc_tmp$specificities)
    auc_tmp <- pROC::auc(roc_tmp)
    ci_AUC  <- ci.auc(roc_tmp) #get confidence interval

    sensitivity <- youden_index$sensitivity
    specificity <- youden_index$specificity

    # Sensitivity = 90%
    II_sensX <- which(roc_tmp$sensitivities >= .90)
    I_sensX <- max(II_sensX)
    sensX_threshold <- roc_tmp$thresholds[I_sensX]
    sensX_specificity <- roc_tmp$specificities[I_sensX]

    # Specificity = 90%
    II_specX <- which(roc_tmp$specificities >= .90)
    I_specX <- min(II_specX)
    specX_threshold <- roc_tmp$thresholds[I_specX]
    specX_sensitivities <- roc_tmp$sensitivities[I_specX]

    num_intermediate <- sum( (mdl_pred > sensX_threshold) & (mdl_pred < specX_threshold) )

    out_v <- c()
    out_v[1] <- CC[i_assay_combos] # company label
    out_v[2] <- formatC(roc_tmp$auc, digits=3, format="fg") # AUC rounded to 2 decimals
    out_v[3] <- formatC(ci_AUC[1], digits=3, format="fg") # AUC lower bound
    out_v[4] <- formatC(ci_AUC[3], digits=3, format="fg") # AUC upper bound
    out_v[5] <- formatC(sensitivity, digits=3, format="fg")
    out_v[6] <- formatC(specificity, digits=3, format="fg")
    out_v[7] <- formatC(youden_index$ppv, digits=3, format="fg")
    out_v[8] <- formatC(youden_index$npv, digits=3, format="fg")
    out_v[9] <- formatC(youden_index$accuracy, digits=3, format="fg")

    tmp <- formatC((log(optimal_cutoff / (1 - optimal_cutoff)) - mdl_logistic$coefficients[1]) /
mdl_logistic$coefficients[2],digits=4,format="fg")
    if (AA[i_assay_combos] %in% single_analytes) {out_v[10] <- tmp}
    else {out_v[10] <- -99}
    out_v[11] <- AA[i_assay_combos] # analyte label

    out_v[12] <- formatC(sensX_threshold, digits=3, format="fg")
```

```
    out_v[13] <- formatC(sensX_specificity, digits=3, format="fg")

    out_v[14] <- formatC(specX_threshold, digits=3, format="fg")
    out_v[15] <- formatC(specX_sensitivities, digits=3, format="fg")

    out_v[16] <- num_intermediate

    out_v[17] <- formula_str
    out_v[18] <- outcome
    out_v[19] <- paste(pred1, collapse = " + ")
    out_v[20] <- covars_TF
    dd <- dim(temp_data)
    out_v[21] <- dd[1]

    brierScore <- mean((mdl_pred-temp_data[[outcome]])^2)
    out_v[22] <- brierScore

    model <- list()
    model[[1]] <- mdl_logistic
    model[[2]] <- roc_tmp

    k <- i_assay_combos + length(combos_all)*(i_outcome-1)
    OUT_5adj[[k]] <- out_v
    MODELS_5adj[[k]] <- model
  }
}

# datafrace for models adjusted (a) for covariates
df_outa <- t(data.frame(OUT_5adj))
df_outa <- data.frame(df_outa)
rownames(df_outa) <- NULL
colnames(df_outa) <- c("company","AUC","AUC_Lower","AUC_Upper",
                       "sensitivity","specificity","ppv","npv","accuracy",
                       "cutoff_value","analyte",
                       "90% Sens Threshold","90% Sens - Specificity",
                       "90% Spec Threshold","90% Spec - Sensitivity",
                       "Num_Intermediate","formula","outcome","predictors",
                       "covars_TF","n_cases","Brier_Score")

df_outa$AUC <- as.numeric(df_outa$AUC)
df_outa$AUC_Lower <- as.numeric(df_outa$AUC_Lower)
df_outa$AUC_Upper <- as.numeric(df_outa$AUC_Upper)
df_outa$Num_Intermediate <- as.numeric(df_outa$Num_Intermediate)
df_outa$n_cases <- as.numeric(df_outa$n_cases)
df_outa <- df_outa %>%
  mutate(intermediate_results = paste(Num_Intermediate, " (",
round(Num_Intermediate/n_cases,2)*100, "%)", sep = ""))

DFa <- df_outa

DFBa <- DFa %>%
  mutate(index_0 = row_number()) %>%
  filter(outcome == vars_outcome[1] ) %>%
  arrange(company,AUC) %>%
  group_by(company) %>%
  mutate(index = row_number())
DFBa$company <- factor(DFBa$company,
                       levels = c("C2N", "Fujirebio", "AlzPath","Janssen", "Roche",
"Quanterix","CSF"))

DFBa <- DFBa %>%
  mutate(companyNums = as.numeric(company)) %>%
  arrange(companyNums,desc(AUC)) %>%
  ungroup() %>%
  mutate(plot_index = desc(row_number()))
DFBa$analyte <- factor(DFBa$analyte,
```

```
                      levels = c("p-tau217 ratio + Ab42/Ab40",
                                 "p-tau217 ratio",
                                 "Ab42/Ab40",
                                 "p-tau217 + Ab42/Ab40",
                                 "p-tau217",
                                 "p-tau181 + Ab42/Ab40 + GFAP + NfL",
                                 "p-tau181 + Ab42/Ab40 + NfL",
                                 "p-tau181 + Ab42/Ab40",
                                 "p-tau181",
                                 "GFAP", "NfL",
                                 "CSF PTAU/AB42"))

DFBa2 <- DFBa %>%
  mutate(AUC_CI = paste0(AUC," (",AUC_Lower, "-",AUC_Upper,")")) %>%
  select("company","analyte",
         "AUC_CI",
         "sensitivity","specificity",
         "90% Sens Threshold","90% Sens - Specificity",
         "90% Spec Threshold","90% Spec - Sensitivity",
         "intermediate_results")
AUCadj <- DFBa %>%
  mutate(AUC_CI = paste0(format(AUC,nsmall=3)," (",format(AUC_Lower,nsmall=3), "-
",format(AUC_Upper,nsmall=3),")")) %>%
  select("company","analyte","AUC_CI","AUC") %>%
  group_by(company) %>%
  mutate(max_AUC = max(AUC)) %>%
  ungroup() %>%
  arrange(desc(max_AUC),company,desc(AUC))%>%
  select(-max_AUC,-AUC) %>%
  rename("AUC (adjusted)" = "AUC_CI")

AUCboth <- full_join(AUCunadj,AUCadj, by = c("company","analyte"))
AUCboth <- AUCboth %>%
  filter(company != "CSF") %>%
  rename(Platform = company,
         Analytes = analyte)

AUCboth$p_unadj <- NA
AUCboth$p_adj <- NA

REF <- data.frame(Platform = C, Analytes = A) %>%
  mutate(index = row_number()) %>%
  filter(Platform != "CSF")
Platforms <- unique(AUCboth$Platform)
Platforms <- Platforms[Platforms != c("AlzPath","Janssen")]

# Loop through each company and perform Delong's test
for (platform in Platforms) {
  # Filter the data for the current company
  platform_data <- AUCboth[AUCboth$Platform == platform, ]

  # Extract the AUC values
  ref_platform <- platform_data$Platform[1]
  ref_analyte <- platform_data$Analytes[1]

  i_ref <- which(REF$Platform == ref_platform & REF$Analytes == ref_analyte)
  roc0 <- MODELS_df[[i_ref]][[2]]

  a_list <- platform_data$Analytes
  a_list <- a_list[-1]
  ps0 <- rep(NA, length(a_list))
  ps00 <- rep(NA, length(a_list))
  ps <- rep(NA, length(a_list))
  psAdj <- rep(NA, length(a_list))

  j <- 1
  for (a in a_list) {
```

```
    i_a <- which(REF$Platform == ref_platform & REF$Analytes == a)

    roc1 <- MODELS_df[[i_a]][[2]]
    t <- roc.test(roc0, roc1, method = "delong")
    ps0[j] <- t$p.value
    j <- j + 1
  }
  p_adjusted <- p.adjust(ps0, method="BH")
  pRaw <- c(NA, ps0)
  pAdj <- c(NA, p_adjusted)

  a_list <- platform_data$Analytes
  k0 <- which(AUCboth$Platform == platform & AUCboth$Analytes == a_list[1])
  kN <- which(AUCboth$Platform == platform & AUCboth$Analytes == a_list[length(a_list)])
  AUCboth$p_unadj[k0:kN] <- pRaw
  AUCboth$p_adj[k0:kN] <- pAdj
}

AUCbest <- AUCboth %>%
  mutate(original_order = row_number()) %>%
  group_by(Platform) %>%
  slice(1) %>%
  ungroup() %>%
  arrange(original_order) %>%
  select(Platform,Analytes,-original_order)
AUCbest$p_best <- NA

for (i in 2:nrow(AUCbest)) {
  ref_platform <- AUCbest$Platform[1]
  ref_analyte <- AUCbest$Analytes[1]
  i_ref <- which(REF$Platform == ref_platform & REF$Analytes == ref_analyte)

  roc0 <- MODELS_df[[i_ref]][[2]]

  comp_platform <- AUCbest$Platform[i]
  comp_analyte <- AUCbest$Analytes[i]
  i_comp <- which(REF$Platform == comp_platform & REF$Analytes == comp_analyte)

  roc1 <- MODELS_df[[i_comp]][[2]]

  t <- roc.test(roc0, roc1, method = "delong")
  p <- t$p.value
  AUCbest$p_best[i] <- p
}

pbest <- AUCbest$p_best
pbest <- pbest[-1]

pbest_adjusted <- p.adjust(pbest, method="BH")
AUCbest$p_best_adj <- c(NA, pbest_adjusted)

AUCboth2 <- AUCboth %>%
  left_join(AUCbest, by = c("Platform","Analytes"))

T_auc_noCov <- AUCboth2 %>%
  mutate(p_unadj = ifelse(p_unadj<0.0001,"<0.0001",
                          ifelse(p_unadj>0.1,round(p_unadj,2),
                                 ifelse(is.na(p_unadj),NA,format(round(p_unadj, 4), scientific =
FALSE)))),
         p_adj = ifelse(p_adj<0.0001,"<0.0001",
                        ifelse(p_adj>0.1,round(p_adj,2),
                               ifelse(is.na(p_adj),NA,format(round(p_adj, 4), scientific =
FALSE)))),
         p_best = ifelse(p_best<0.0001,"<0.0001",
                         ifelse(p_best>0.1,round(p_best,2),
                                ifelse(is.na(p_best),NA,format(round(p_best, 4), scientific =
FALSE)))),
```

```
        p_best_adj = ifelse(p_best_adj<0.0001,"<0.0001",
                            ifelse(p_best_adj>0.1,round(p_best_adj,2),
                                   ifelse(is.na(p_best_adj),NA,format(round(p_best_adj, 4),
scientific = FALSE))))) %>%
  rename("No Cov - unadjusted p=" = p_unadj,
         "No Cov - adjusted p=" = p_adj,
         "No Cov - Best unadjusted  p=" = p_best,
         "No Cov - Best adjusted p=" = p_best_adj)

################################################################################
################################################################################

AUCadj <- AUCadj %>%
  rename(Platform = company, Analytes = analyte) %>%
  filter(Platform != "CSF")

AUCboth <- AUCadj %>% #### Replace AUCboth with AUCadj
  select(Platform,Analytes)
AUCboth$p_unadjCov <- NA
AUCboth$p_adjCov <- NA

REF <- data.frame(Platform = C, Analytes = A) %>%
  mutate(index = row_number()) %>%
  filter(Platform != "CSF")
Platforms <- unique(AUCadj$Platform)
Platforms <- Platforms[Platforms != c("AlzPath","Janssen")]

# Loop through each company and perform Delong's test
for (platform in Platforms) {
  # Filter the data for the current company
  platform_data <- AUCboth[AUCboth$Platform == platform, ]

  # Extract the AUC values
  ref_platform <- platform_data$Platform[1]
  ref_analyte <- platform_data$Analytes[1]

  i_ref <- which(REF$Platform == ref_platform & REF$Analytes == ref_analyte)
  roc0 <- MODELS_5adj[[i_ref]][[2]]

  a_list <- platform_data$Analytes
  a_list <- a_list[-1]
  ps0 <- rep(NA, length(a_list))
  ps00 <- rep(NA, length(a_list))
  ps <- rep(NA, length(a_list))
  psAdj <- rep(NA, length(a_list))

  j <- 1
  for (a in a_list) {
    i_a <- which(REF$Platform == ref_platform & REF$Analytes == a)

    roc1 <- MODELS_5adj[[i_a]][[2]]
    t <- roc.test(roc0, roc1, method = "delong")
    ps0[j] <- t$p.value
    j <- j + 1
  }
  p_adjusted <- p.adjust(ps0, method="BH")
  pRaw <- c(NA, ps0)
  pAdj <- c(NA, p_adjusted)

  a_list <- platform_data$Analytes
  k0 <- which(AUCboth$Platform == platform & AUCboth$Analytes == a_list[1])
  kN <- which(AUCboth$Platform == platform & AUCboth$Analytes == a_list[length(a_list)])
  AUCboth$p_unadjCov[k0:kN] <- pRaw
  AUCboth$p_adjCov[k0:kN] <- pAdj
}

AUCbest <- AUCboth %>%
```

```
  mutate(original_order = row_number()) %>%
  group_by(Platform) %>%
  slice(1) %>%
  ungroup() %>%
  arrange(original_order) %>%
  select(Platform,Analytes,-original_order)
AUCbest$p_bestCov <- NA

for (i in 2:nrow(AUCbest)) {
  ref_platform <- AUCbest$Platform[1]
  ref_analyte <- AUCbest$Analytes[1]
  i_ref <- which(REF$Platform == ref_platform & REF$Analytes == ref_analyte)
  roc0 <- MODELS_5adj[[i_ref]][[2]]

  comp_platform <- AUCbest$Platform[i]
  comp_analyte <- AUCbest$Analytes[i]
  i_comp <- which(REF$Platform == comp_platform & REF$Analytes == comp_analyte)

  roc1 <- MODELS_5adj[[i_comp]][[2]]

  t <- roc.test(roc0, roc1, method = "delong")
  p <- t$p.value
  AUCbest$p_bestCov[i] <- p
}

pbest <- AUCbest$p_bestCov
pbest <- pbest[-1]

pbest_adjusted <- p.adjust(pbest, method="BH")
AUCbest$p_best_adjCov <- c(NA, pbest_adjusted)

AUCboth2 <- AUCboth %>%
  left_join(AUCbest, by = c("Platform","Analytes"))

T_auc_Cov <- AUCboth2 %>%
  mutate(p_unadjCov = ifelse(p_unadjCov<0.0001,"<0.0001",
                             ifelse(p_unadjCov>0.1,round(p_unadjCov,2),
                                    ifelse(is.na(p_unadjCov),NA,format(round(p_unadjCov, 4),
scientific = FALSE)))),
         p_adjCov = ifelse(p_adjCov<0.0001,"<0.0001",
                           ifelse(p_adjCov>0.1,round(p_adjCov,2),
                                  ifelse(is.na(p_adjCov),NA,format(round(p_adjCov, 4), scientific =
FALSE)))),
         p_bestCov = ifelse(p_bestCov<0.0001,"<0.0001",
                            ifelse(p_bestCov>0.1,round(p_bestCov,2),
                                   ifelse(is.na(p_bestCov),NA,format(round(p_bestCov, 4),
scientific = FALSE)))),
         p_best_adjCov = ifelse(p_best_adjCov<0.0001,"<0.0001",
                                ifelse(p_best_adjCov>0.1,round(p_best_adjCov,2),
                                       ifelse(is.na(p_best_adjCov),NA,format(round(p_best_adjCov,
4), scientific = FALSE))))) %>%
  rename("Cov - unadjusted p=" = p_unadjCov,
         "Cov - adjusted p=" = p_adjCov,
         "Cov - Best unadjusted  p=" = p_bestCov,
         "Cov - Best adjusted p=" = p_best_adjCov)

T_auc <- full_join(T_auc_noCov, T_auc_Cov, by = c("Platform","Analytes"))

T_auc2 <- T_auc %>%
  select(Platform, Analytes,
         "AUC (unadjusted)", "No Cov - adjusted p=", "No Cov - Best adjusted p=",
         "AUC (adjusted)","Cov - adjusted p=","Cov - Best adjusted p=")

write.csv(T_auc2, file = here(out_folder,outfile_AUCs), row.names = FALSE)
```

**Code B13. R code for creating AUC forest plots (requires B14).**

```
# FNIH Project 2
# make AUC Forest plots
#### Coded: Kellen Petersen
#### Date as of commenting and documentation - 05/31/2024

# Load libraries
library(tidyverse)
library(data.table)
library(RVAideMemoire)
library(pROC)
library(RColorBrewer)
library(MuMIn)
library(here)
library(cowplot)
library(ggeasy)
library(scales)
library(patchwork)

# Load functions
source("functions_forest.R")

# Load data
#here::i_am("make_forest_using_function.R")

# Load data
data_file <- here("cross_sectional_data_CLEAN.csv")
data.in00 <- read.csv(data_file)

# Choose cohort and coavariates
data.in0 <- data.in00 # Entire cohort
covars_TF <- FALSE # without covariates

# Make models for outcome 1
vars_outcome <- c("CENTILOIDS_10")
T1 <- make_model(data.in0,covars_TF,vars_outcome)
breaks1 = c(0, 0.50, 0.733, 1)
AUC_indicatorline = 0.733
Y <- c(.4, 1)
p1 <- getAUCs_withLabels(T1,breaks1,AUC_indicatorline,rev_axis=FALSE,Y[1],Y[2])
T1$key <- paste(T1$company, T1$analyte)

# Make models for outcome 2
vars_outcome <- c("TAU_MesialTemporal_10")
T2 <- make_model(data.in0,covars_TF,vars_outcome)
T2$key <- paste(T2$company, T2$analyte)
order_index2 <- match(T1$key, T2$key)
T2_ordered <- T2[order_index2, ]
T2_ordered$key <- NULL
breaks1 =c(0, 0.50, 0.767, 1)
AUC_indicatorline = 0.767
Y <- c(.35, 1)
p2 <- getAUCs_withoutLabels(T2_ordered,breaks1,AUC_indicatorline,rev_axis=FALSE,Y[1],Y[2])

# Make models for outcome 3
vars_outcome <- c("atrophy_10")
T3 <- make_model(data.in0,covars_TF,vars_outcome)
T3$key <- paste(T3$company, T3$analyte)
order_index3 <- match(T1$key, T3$key)
T3_ordered <- T3[order_index3, ]
T3_ordered$key <- NULL
breaks1 = c(0.50, 0.758, 1)
AUC_indicatorline = 0.758
Y <- c(0.35, 1)
p3 <- getAUCs_withoutLabels(T3_ordered,breaks1,AUC_indicatorline,rev_axis=FALSE,Y[1],Y[2])
```

```
# Make models for outcome 4
data.in0$impaired_10 <- ifelse(data.in0$CDR > 0, 1, 0)
data.in0$impaired_10 <- as.factor(data.in0$impaired_10)
vars_outcome <- c("impaired_10")
T4 <- make_model(data.in0,covars_TF,vars_outcome)
T4$key <- paste(T4$company, T4$analyte)
order_index4 <- match(T1$key, T4$key)
T4_ordered <- T4[order_index4, ]
T4_ordered$key <- NULL
breaks1 = c(0, 0.45, 0.559, .75)
AUC_indicatorline = 0.559
Y <- c(.35, .85)
p4 <- getAUCs_withoutLabels(T4_ordered,breaks1,AUC_indicatorline,rev_axis=FALSE,Y[1],Y[2])

# Add titles
p1b <- p1 +
  ggtitle("Amyloid PET") +
  theme(plot.title = element_text(hjust = 0.5,size = 10),
        plot.subtitle = element_text(hjust = 0.5))
p2b <- p2 +
  ggtitle("Early tau PET") +
  theme(plot.title = element_text(hjust = 0.5,size = 10),
        plot.subtitle = element_text(hjust = 0.5))
p3b <- p3 +
  ggtitle("Cortical thickness") +
  theme(plot.title = element_text(hjust = 0.5,size = 10),
        plot.subtitle = element_text(hjust = 0.5))
p4b <- p4 +
  ggtitle("Cognitive impairment") +
  theme(plot.title = element_text(hjust = 0.5,size = 10),
        plot.subtitle = element_text(hjust = 0.5))

# Combine plots
pp <- p1b | p2b | p3b | p4b
pp

# Save plot
# ggsave("Forest_AUC_CLEAN.png", pp, device = "png", dpi = 500, width = 11, height = 6, units =
"in")
```

**Code B14. R code for functions used in making AUC forest plots.**

```
# FNIH Project 2
# Function to make AUC Forest plots
#### Coded: Kellen Petersen
#### Date as of commenting and documentation - 05/31/2024

getAUCs_withLabels <- function(T,breaks1,indicatorline,rev_axis=FALSE,y1,y2){
  df <- T %>%
    select(company,AUC,AUC_Lower,AUC_Upper,analyte)

  df$AUC <- as.numeric(df$AUC)
  df$AUC_Lower <- as.numeric(df$AUC_Lower)
  df$AUC_Upper <- as.numeric(df$AUC_Upper)
  DF <- df %>%
    mutate(index_0 = row_number()) %>%
    group_by(company) %>%
    mutate(index = row_number()) %>%
    ungroup() %>%
    mutate(plot_index = desc(row_number()))
  DF$analyte_c <- DF$analyte
  DF$analyte_c <- factor(DF$analyte_c,levels=c("p-tau217 ratio + Ab42/Ab40",
                                              "p-tau217 ratio",
                                              "p-tau217 + Ab42/Ab40",
                                              "p-tau217",
                                              "Ab42/Ab40",
                                              "p-tau181 + Ab42/Ab40 + GFAP + NfL",
                                              "p-tau181 + Ab42/Ab40 + NfL",
                                              "p-tau181 + Ab42/Ab40",
                                              "p-tau181",
                                              "GFAP",
                                              "NfL"))
  AB <- paste0("A","\U03B2","42/A","\U03B2","40")
  DF$analyte_l <- DF$analyte
  DF$analyte_l <- as.character(DF$analyte_l)
  DF[which(DF$analyte=="p-tau217 + Ab42/Ab40"),"analyte_l"] <- paste0("p-tau217 + ",AB)
  DF[which(DF$analyte=="p-tau217 ratio + Ab42/Ab40"),"analyte_l"] <- paste0("%p-tau217 + ",AB)
  DF[which(DF$analyte=="p-tau217 ratio"),"analyte_l"] <- paste0("%p-tau217")
  DF[which(DF$analyte=="Ab42/Ab40"),"analyte_l"] <- AB
  DF[which(DF$analyte=="p-tau181 + Ab42/Ab40"),"analyte_l"] <- paste0("p-tau181 + ",AB)
  DF[which(DF$analyte=="p-tau181 + Ab42/Ab40 + NfL"),"analyte_l"] <- paste0("p-tau181 + ",AB," +
NfL")
  DF[which(DF$analyte=="p-tau181 + Ab42/Ab40 + GFAP + NfL"),"analyte_l"] <- paste0("p-tau181 +
",AB," + GFAP + NfL")

  analyte_colors <- c("p-tau217 ratio + Ab42/Ab40" = "black",
                      "p-tau217 + Ab42/Ab40" = "darkgray",

                      "p-tau217 ratio" = "#145A32",
                      "p-tau217" = "#008000",
                      "p-tau181" = "#00FF00",

                      "Ab42/Ab40" = "blue",
                      "NfL" = "maroon",
                      "GFAP" = "red",

                      "p-tau181 + Ab42/Ab40 + GFAP + NfL" = "purple",
                      "p-tau181 + Ab42/Ab40 + NfL" = "orange",
                      "p-tau181 + Ab42/Ab40" = "cyan"
  )

  min_y <- round(min(DF$AUC_Lower),digits = 2)
  min_y <- y1
  min_y_text <- min_y-0.1
  text_start <- min_y_text
  round_any = function(x, accuracy, f=round){f(x/ accuracy) * accuracy}
  max_y <-  round_any(max(DF$AUC_Upper) ,accuracy = 0.2 , f = ceiling)
```

```
  max_y <- max(c(max(breaks1),max_y))
  if(rev_axis==TRUE){
    max_y <- max_y+0.1
    text_start <- max_y
  }

  company_count <- table(DF$company)
  complab <- DF %>%
    group_by(company) %>%
    slice(1) %>%
    ungroup() %>%
    arrange(desc(plot_index))

  C2N_Index_Point <- as.numeric(complab[complab$company=="C2N","plot_index"])
  Fujirebio_Index_Point <- as.numeric(complab[complab$company=="Fujirebio","plot_index"])
  ALZPath_Index_Point <- as.numeric(complab[complab$company=="AlzPath","plot_index"])
  Janssen_Index_Point <- as.numeric(complab[complab$company=="Janssen","plot_index"])
  Roche_Index_Point <- as.numeric(complab[complab$company=="Roche","plot_index"])
  Quanterix_Index_Point <- as.numeric(complab[complab$company=="Quanterix","plot_index"])

  Indices <- complab$plot_index
  Endpoint <- as.numeric(Quanterix_Index_Point - company_count["Quanterix"])
  title_plot <- ""

  test_plot <-  ggplot(data=DF,
                              aes(x = plot_index, #uses index as the x-axis (flipped to y later)
                                  y = AUC, # y-axis is spearman rho, flipped to x-axis later
                                  ymin = min_y, ymax = max_y ))+ #limited of y-axis (flipped to x
later)
    geom_rect(aes(xmin = Indices[1]+0.5, xmax = Indices[2]+0.5, ymin = min_y_text, ymax = max_y),
              fill = "#E5E4E2", alpha = 0.04) +
    geom_rect(aes(xmin = Indices[3]+0.5, xmax = Indices[4]+0.5, ymin = min_y_text, ymax = max_y),
              fill = "#E5E4E2", alpha = 0.04) +
    geom_rect(aes(xmin = Indices[5]+0.5, xmax = Indices[6]+0.5, ymin = min_y_text, ymax = max_y),
              fill = "#E5E4E2", alpha = 0.04) +
    geom_hline(aes(fill="black"),yintercept =indicatorline, linetype=2)+ #creates horizontal
(flipped to vertical) line at 0
    geom_point(aes(col=analyte_c))+ #sets the coloring based on whether its the summary measure or
not
    geom_errorbar(aes(ymin=(AUC_Lower), ymax=(AUC_Upper),col=analyte_c),width = 0, cex = 1,size=4)+
#formatting of forest lines
    geom_text(aes(x = C2N_Index_Point, y = text_start, label = "C2N", hjust =
0),size=3,family="Calibri") +
    geom_text(aes(x = Fujirebio_Index_Point, y = text_start, label = "Fujirebio", hjust =
0),size=3,family="Calibri") +
    geom_text(aes(x = ALZPath_Index_Point, y = text_start, label = "ALZpath", hjust =
0),size=3,family="Calibri") +
    geom_text(aes(x = Janssen_Index_Point, y = text_start, label = "Janssen", hjust =
0),size=3,family="Calibri") +
    geom_text(aes(x = Roche_Index_Point, y = text_start, label = "Roche", hjust =
0),size=3,family="Calibri") +
    geom_text(aes(x = Quanterix_Index_Point, y = text_start, label = "Quanterix", hjust =
0),size=3,family="Calibri")+
    labs(x = " ",
         y = "AUC", #y axis will have no label, x axis will be spearman rho
         title = title_plot)+
    theme_classic()+ #gives the title based on the function call variable
    theme(plot.title=element_text(size=16,face="bold"), #these are all text formatting
          axis.ticks.y=element_line(size = 1, color = "black"),
          axis.text.x=element_text(size=7,face="bold"),
          axis.text.y=element_text(size=8),
          axis.title=element_text(size=12,face="bold"),
          strip.text.y = element_blank(),
          legend.position =  "none")+ #no legend
    scale_x_continuous(breaks=DF$plot_index,labels=DF$analyte_l)+
    scale_color_manual(values=analyte_colors)+
```

```
    scale_fill_manual(values = analyte_colors)+ #makes sure dots and lines are colored based on
color code
    coord_flip()

  test_plot <- test_plot +
    scale_y_continuous(limits = c(y1-.1,y2),
                       breaks = breaks1,
                       labels = scales::number_format(accuracy = 0.001))
  return(test_plot)
}
```

```
getAUCs_withoutLabels <- function(T,breaks1,indicatorline,rev_axis=FALSE,y1,y2){
  df <- T %>%
    select(company,AUC,AUC_Lower,AUC_Upper,analyte)

  df$AUC <- as.numeric(df$AUC)
  df$AUC_Lower <- as.numeric(df$AUC_Lower)
  df$AUC_Upper <- as.numeric(df$AUC_Upper)
  DF <- df %>%
    mutate(index_0 = row_number()) %>%
    group_by(company) %>%
    mutate(index = row_number()) %>%
    ungroup() %>%
    mutate(plot_index = desc(row_number()))
  DF$analyte_c <- DF$analyte
  DF$analyte_c <- factor(DF$analyte_c,levels=c("p-tau217 ratio + Ab42/Ab40",
                                               "p-tau217 ratio",
                                               "p-tau217 + Ab42/Ab40",
                                               "p-tau217",
                                               "Ab42/Ab40",
                                               "p-tau181 + Ab42/Ab40 + GFAP + NfL",
                                               "p-tau181 + Ab42/Ab40 + NfL",
                                               "p-tau181 + Ab42/Ab40",
                                               "p-tau181",
                                               "GFAP",
                                               "NfL"))
  AB <- paste0("A","\U03B2","42/A","\U03B2","40")
  DF$analyte_l <- DF$analyte
  DF$analyte_l <- as.character(DF$analyte_l)
  DF[which(DF$analyte=="p-tau217 + Ab42/Ab40"),"analyte_l"] <- paste0("p-tau217 + ",AB)
  DF[which(DF$analyte=="p-tau217 ratio + Ab42/Ab40"),"analyte_l"] <- paste0("p-tau217 ratio + ",AB)
  DF[which(DF$analyte=="Ab42/Ab40"),"analyte_l"] <- AB
  DF[which(DF$analyte=="p-tau181 + Ab42/Ab40"),"analyte_l"] <- paste0("p-tau181 + ",AB)
  DF[which(DF$analyte=="p-tau181 + Ab42/Ab40 + NfL"),"analyte_l"] <- paste0("p-tau181 + ",AB," +
NfL")
  DF[which(DF$analyte=="p-tau181 + Ab42/Ab40 + GFAP + NfL"),"analyte_l"] <- paste0("p-tau181 +
",AB," + GFAP + NfL")

  analyte_colors <- c("p-tau217 ratio + Ab42/Ab40" = "black",
                      "p-tau217 + Ab42/Ab40" = "darkgray",

                      "p-tau217 ratio" = "#145A32",
                      "p-tau217" = "#008000",
                      "p-tau181" = "#00FF00",

                      "Ab42/Ab40" = "blue",
                      "NfL" = "maroon",
                      "GFAP" = "red",

                      "p-tau181 + Ab42/Ab40 + GFAP + NfL" = "purple",
```

```
                     "p-tau181 + Ab42/Ab40 + NfL" = "orange",
                     "p-tau181 + Ab42/Ab40" = "cyan"
  )

  min_y <- round(min(DF$AUC_Lower),digits = 2)
  min_y <- y1
  min_y_text <- min_y-0
  text_start <- min_y_text
  round_any = function(x, accuracy, f=round){f(x/ accuracy) * accuracy}
  max_y <-  round_any(max(DF$AUC_Upper) ,accuracy = 0.05 , f = ceiling)
  print(max_y)
  max_y <- 0
  max_y <- max(c(max(breaks1),max_y))
  print(max_y)
  if(rev_axis==TRUE){
    max_y <- max_y+0.1
    text_start <- max_y
  }

  company_count <- table(DF$company)
  complab <- DF %>%
    group_by(company) %>%
    slice(1) %>%
    ungroup() %>%
    arrange(desc(plot_index))

  C2N_Index_Point <- as.numeric(complab[complab$company=="C2N","plot_index"])
  Fujirebio_Index_Point <- as.numeric(complab[complab$company=="Fujirebio","plot_index"])
  ALZPath_Index_Point <- as.numeric(complab[complab$company=="AlzPath","plot_index"])
  Janssen_Index_Point <- as.numeric(complab[complab$company=="Janssen","plot_index"])
  Roche_Index_Point <- as.numeric(complab[complab$company=="Roche","plot_index"])
  Quanterix_Index_Point <- as.numeric(complab[complab$company=="Quanterix","plot_index"])

  Indices <- complab$plot_index
  Endpoint <- as.numeric(Quanterix_Index_Point - company_count["Quanterix"])
  title_plot <- ""

  test_plot <-  ggplot(data=DF,
                              aes(x = plot_index, #uses index as the x-axis (flipped to y later)
                                  y = AUC, # y-axis is spearman rho, flipped to x-axis later
                                  ymin = min_y, ymax = max_y ))+ #limited of y-axis (flipped to x
later)
    geom_rect(aes(xmin = Indices[1]+0.5, xmax = Indices[2]+0.5, ymin = min_y_text, ymax = max_y),
              fill = "#E5E4E2", alpha = 0.04) +
    geom_rect(aes(xmin = Indices[3]+0.5, xmax = Indices[4]+0.5, ymin = min_y_text, ymax = max_y),
              fill = "#E5E4E2", alpha = 0.04) +
    geom_rect(aes(xmin = Indices[5]+0.5, xmax = Indices[6]+0.5, ymin = min_y_text, ymax = max_y),
              fill = "#E5E4E2", alpha = 0.04) +
    geom_hline(aes(fill="black"),yintercept =indicatorline, linetype=2)+ #creates horizontal
(flipped to vertical) line at 0
    geom_point(aes(col=analyte_c))+ #sets the coloring based on whether its the summary measure or
not
    geom_errorbar(aes(ymin=(AUC_Lower), ymax=(AUC_Upper),col=analyte_c),width = 0, cex = 1,size=4)+
#formatting of forest lines
    labs(x = " ",
         y = "AUC", #y axis will have no label, x axis will be spearman rho
         title = title_plot)+
    theme_classic()+ #gives the title based on the function call variable
    theme(plot.title=element_text(size=16,face="bold"), #these are all text formatting
          axis.ticks.y=element_line(size = 1, color = "black"),
          axis.text.x=element_text(size=7,face="bold"),
          axis.text.y=element_text(size=8),
          axis.title=element_text(size=12,face="bold"),
          strip.text.y = element_blank(),
          legend.position =  "none")+ #no legend
    scale_x_continuous(breaks=DF$plot_index,labels=DF$analyte_l)+
```

```
    scale_color_manual(values=analyte_colors)+
    scale_fill_manual(values = analyte_colors)+ #makes sure dots and lines are colored based on
color code
    theme(axis.text.y=element_blank(), # Remove x axis labels
          axis.ticks.y=element_blank()) + # Remove x axis ticks
    coord_flip() #+#+

  test_plot <- test_plot +
    scale_y_continuous(limits = c(y1,y2),
                       breaks = breaks1,
                       labels = scales::number_format(accuracy = 0.001))

  return(test_plot)
}

make_model <- function(data.in0,covars_TF,vars_outcome){
  data.in.5 <- data.in0
  #Covariates
  age_var <- "AGE" #variable with age
  sex_var <- "PTGENDER" #variable with sex
  educ_var <- "PTEDUCAT" #variable with education

  APOE_variable <- "APOE_genotype" #Variable with APOE genotypes
  data.in.5$APOE_Cat <- factor(data.in.5[[APOE_variable]], levels = c("33", "34", "24", "22", "23",
"44"))
  list_covariates <- list(c(age_var, sex_var, "APOE_Cat"))
  if (covars_TF) {
    vars_covariates <- list_covariates
  } else {
    vars_covariates <- NULL
  }

  # Assays Lists
  assays_c2n       <- c("C2N_plasma_Abeta42_Abeta40", "C2N_plasma_ptau217",
"C2N_plasma_ptau217_ratio")
  assays_fuji      <- c("Fuji_plasma_Ab42_Ab40","Fuji_plasma_ptau217")
  assays_alzpath   <- c("AlzPath_plasma_ptau217")
  assays_janssen   <- c("Janssen_plasma_ptau217")
  assays_roche     <-
c("Roche_plasma_Ab42_Ab40","Roche_plasma_ptau181","Roche_plasma_NfL","Roche_plasma_GFAP")
  assays_quanterix <- c("QX_plasma_Ab42_Ab40","QX_plasma_ptau181","QX_plasma_NfL","QX_plasma_GFAP")
  assays_csfratio  <- c("PTAU_over_ABETA42")

  # Create list of model indices
  combos_c2n       <- list(assays_c2n[c(3,1)],assays_c2n[c(2,1)],
                           assays_c2n[c(3)],assays_c2n[c(2)],assays_c2n[c(1)])              # 5
models
  combos_fuji      <- list(assays_fuji[c(2,1)],assays_fuji[c(2)],assays_fuji[c(1)])         # 3
models
  combos_alzath    <- list(assays_alzpath)                                                  # 1
model
  combos_janssen   <- list(assays_janssen)                                                  # 1
model
  combos_roche     <- list(assays_roche[c(2,1,4,3)],assays_roche[c(2,1,3)],
                           assays_roche[c(2,1)],
                           assays_roche[c(2)],assays_roche[c(1)],
                           assays_roche[c(4)],assays_roche[c(3)])                           # 7
models
  combos_quanterix <- list(assays_quanterix[c(2,1,4,3)],assays_quanterix[c(2,1,3)],
                           assays_quanterix[c(2,1)],
                           assays_quanterix[c(2)],assays_quanterix[c(1)],
```

```
                         assays_quanterix[c(4)],assays_quanterix[c(3)])                  # 7
models
  combos_csfratio   <- list(assays_csfratio)                                             # 1
model

  combos_all <- list()
  combos_all[[1]] <- combos_c2n[1]
  combos_all[[2]] <- combos_c2n[2]
  combos_all[[3]] <- combos_c2n[3]
  combos_all[[4]] <- combos_c2n[4]
  combos_all[[5]] <- combos_c2n[5]

  combos_all[[6]] <- combos_fuji[1]
  combos_all[[7]] <- combos_fuji[2]
  combos_all[[8]] <- combos_fuji[3]

  combos_all[[9]] <- combos_alzath[1]

  combos_all[[10]] <- combos_janssen[1]

  combos_all[[11]] <- combos_roche[1]
  combos_all[[12]] <- combos_roche[2]
  combos_all[[13]] <- combos_roche[3]
  combos_all[[14]] <- combos_roche[4]
  combos_all[[15]] <- combos_roche[5]
  combos_all[[16]] <- combos_roche[6]
  combos_all[[17]] <- combos_roche[7]

  combos_all[[18]] <- combos_quanterix[1]
  combos_all[[19]] <- combos_quanterix[2]
  combos_all[[20]] <- combos_quanterix[3]
  combos_all[[21]] <- combos_quanterix[4]
  combos_all[[22]] <- combos_quanterix[5]
  combos_all[[23]] <- combos_quanterix[6]
  combos_all[[24]] <- combos_quanterix[7]

  Combos <- combos_all

  # Company label
  C_c2n <- rep("C2N", 5)
  C_fuji <- rep("Fujirebio", 3)
  C_alzpath <- rep("AlzPath", 1)
  C_janssen <- rep("Janssen", 1)
  C_roche <- rep("Roche", 7)
  C_quanterix <- rep("Quanterix", 7)
  C <- c(C_c2n, C_fuji, C_alzpath, C_janssen, C_roche, C_quanterix)
  CC <- C

  # Analyte label
  A_c2n <- c("p-tau217 ratio + Ab42/Ab40","p-tau217 + Ab42/Ab40",
             "p-tau217 ratio","p-tau217","Ab42/Ab40")
  A_fuji <- c("p-tau217 + Ab42/Ab40", "p-tau217", "Ab42/Ab40")
  A_alzpath <- c("p-tau217")
  A_janssen <- c("p-tau217")
  A_roche <- c("p-tau181 + Ab42/Ab40 + GFAP + NfL",
               "p-tau181 + Ab42/Ab40 + NfL",
               "p-tau181 + Ab42/Ab40",
               "p-tau181","Ab42/Ab40",
               "GFAP","NfL")
  A_quanterix <- c("p-tau181 + Ab42/Ab40 + GFAP + NfL",
                   "p-tau181 + Ab42/Ab40 + NfL",
                   "p-tau181 + Ab42/Ab40",
                   "p-tau181","Ab42/Ab40",
                   "GFAP","NfL")
  A_qcsfratio <- c("CSF PTAU/AB42")
  A <- c(A_c2n, A_fuji, A_alzpath, A_janssen, A_roche, A_quanterix)
  AA <- A
```

```
single_analytes <- c("p-tau217 ratio",
                     "Ab42/Ab40",
                     "p-tau217",
                     "p-tau181",
                     "GFAP",
                     "NfL",
                     "CSF PTAU/AB42")

OUT_5 <- list()
MODELS_5 <- list()
k <- 0
data.in <- data.in.5
# Loop through the combinations of predictors
for (i_outcome in 1:length(vars_outcome)) {
  for (i_assay_combos in 1:length(Combos)) {

    outcome <- vars_outcome[i_outcome]
    pred1 <- Combos[[i_assay_combos]]
    pred2 <- vars_covariates
    preds <- c(unlist(pred1), unlist(pred2))

    print(i_assay_combos)
    print(preds)

    formula_str <- paste(outcome, " ~ ", paste(preds, collapse = " + "))
    model_formula <- as.formula(formula_str)

    temp_data <- data.in[complete.cases(data.in[,preds]), c(outcome,preds)] %>%
      na.omit()

    mdl_logistic <- glm(model_formula, data = temp_data, family = "binomial")
    mdl_pred <- predict(mdl_logistic, data = temp_data, type = "response")

    roc_tmp <- pROC::roc(temp_data[[outcome]],mdl_pred,plot=FALSE,print.auc=FALSE)
    youden_index <- coords(roc_tmp, "best", ret="all")
    optimal_cutoff <- youden_index$threshold

    T <- data.frame(roc_tmp$thresholds, roc_tmp$sensitivities, roc_tmp$specificities)
    auc_tmp <- pROC::auc(roc_tmp)
    ci_AUC  <- ci.auc(roc_tmp) #get confidence interview

    sensitivity <- youden_index$sensitivity
    specificity <- youden_index$specificity

    # Sensitivity = 90%
    II_sensX <- which(roc_tmp$sensitivities >= .90)
    I_sensX <- max(II_sensX)
    sensX_threshold <- roc_tmp$thresholds[I_sensX]
    sensX_specificity <- roc_tmp$specificities[I_sensX]

    # Specificity = 90%
    II_specX <- which(roc_tmp$specificities >= .90)
    I_specX <- min(II_specX)
    specX_threshold <- roc_tmp$thresholds[I_specX]
    specX_sensitivities <- roc_tmp$sensitivities[I_specX]

    num_intermediate <- sum( (mdl_pred > sensX_threshold) & (mdl_pred < specX_threshold) )
    num_intermediate

    out_v <- c()
    out_v[1] <- CC[i_assay_combos] # company label
    out_v[2] <- round(roc_tmp$auc,3) # AUC rounded to 2 decimals
    out_v[3] <- round(ci_AUC[1],3) # AUC lower bound
    out_v[4] <- round(ci_AUC[3],3) # AUC upper bound
    out_v[5] <- AA[i_assay_combos] # analyte label
```

```
      model <- list()
      model[[1]] <- mdl_logistic
      model[[2]] <- roc_tmp

      k <- i_assay_combos + length(combos_all)*(i_outcome-1)
      print(i_assay_combos)
      print(i_outcome)
      print(k)
      print(optimal_cutoff)
      OUT_5[[k]] <- out_v
      MODELS_5[[k]] <- model
    }
  }

  df_out <- t(data.frame(OUT_5))
  df_out <- data.frame(df_out)
  rownames(df_out) <- NULL
  colnames(df_out) <- c("company","AUC","AUC_Lower","AUC_Upper","analyte")

  df_out$AUC <- as.numeric(df_out$AUC)
  df_out$AUC_Lower <- as.numeric(df_out$AUC_Lower)
  df_out$AUC_Upper <- as.numeric(df_out$AUC_Upper)
  DF <- df_out
  DFB <- DF %>%
    mutate(index_0 = row_number()) %>%
    filter(outcome == vars_outcome[1] ) %>%
    arrange(company,AUC) %>%
    group_by(company) %>%
    mutate(index = row_number())
  DFB$company <- factor(DFB$company, levels = c("C2N", "Fujirebio", "AlzPath","Janssen", "Roche",
"Quanterix","CSF"))
  DFB <- DFB %>%
    mutate(companyNums = as.numeric(company)) %>%
    arrange(companyNums,desc(AUC)) %>%
    ungroup() %>%
    mutate(plot_index = desc(row_number()))
  DFB$analyte <- factor(DFB$analyte,
                        levels = c("p-tau217 ratio + Ab42/Ab40",
                                   "p-tau217 ratio",
                                   "Ab42/Ab40",
                                   "p-tau217 + Ab42/Ab40",
                                   "p-tau217",
                                   "p-tau181 + Ab42/Ab40 + GFAP + NfL",
                                   "p-tau181 + Ab42/Ab40 + NfL",
                                   "p-tau181 + Ab42/Ab40",
                                   "p-tau181",
                                   "GFAP", "NfL",
                                   "CSF PTAU/AB42"))

  T <- DFB %>%
    select(company,AUC,AUC_Lower,AUC_Upper,analyte) %>%
    group_by(company) %>%
    mutate(max_AUC = max(AUC)) %>%
    ungroup() %>%
    arrange(desc(max_AUC), company, desc(AUC)) %>%
    select(-max_AUC) %>%
    filter(company != "CSF")

  return(T)
}
```

**Appendix C. External quality control analyses.**

Coefficient of variance (CV), calculated as the standard deviation divided by the mean times 100, in measured values for aliquots of two pooled plasma controls with known higher or lower p-tau217 values.

**ALZpath Quanterix**

| Assay | QC control | CV |
|---|---|---|
| p-tau217 | high | 8.02% |
| p-tau217 | low | 8.93% |

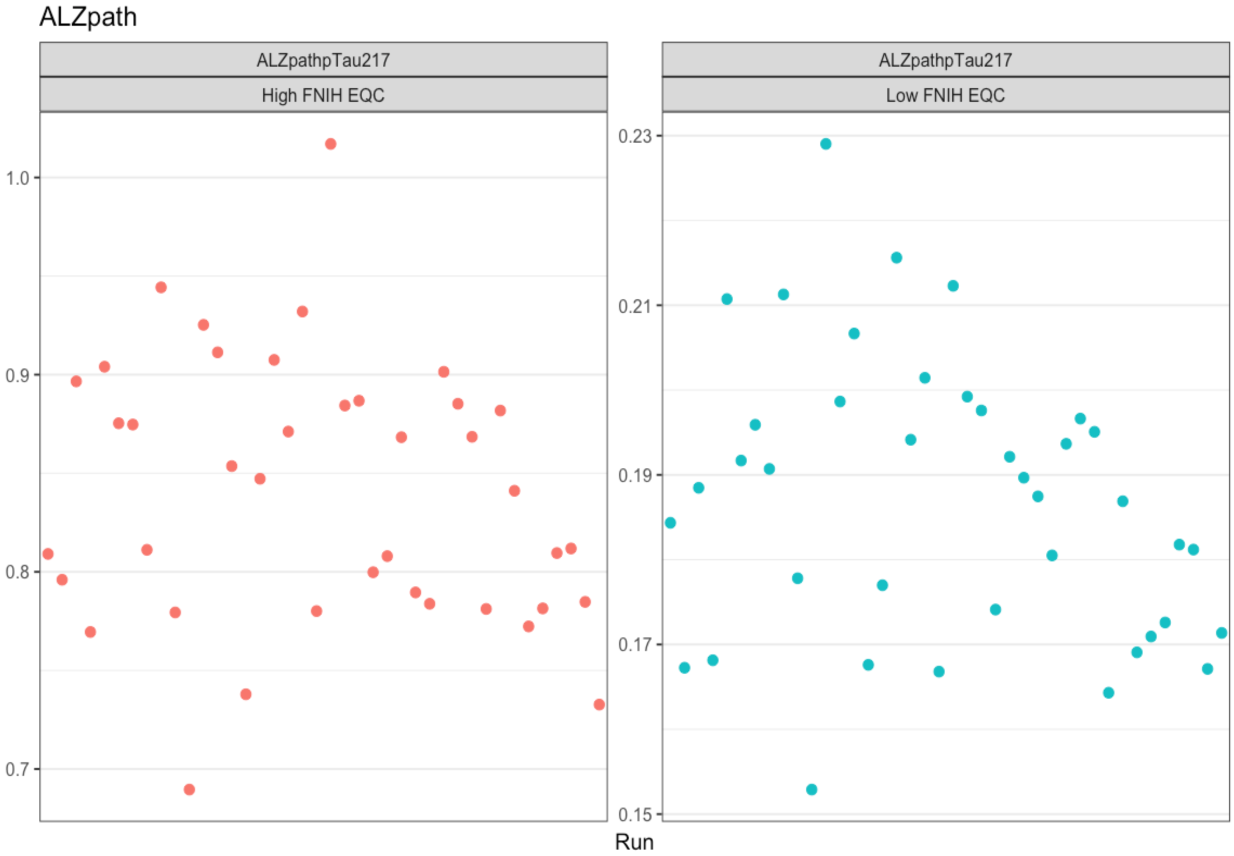

**C2N PrecivityAD2**

| Assay | QC control | CV |
|---|---|---|
| Aβ40 | high | 5.41% |
| Aβ40 | low | 3.23% |
| Aβ42 | high | 9.43% |
| Aβ42 | low | 8.82% |
| Aβ42/Aβ40 | high | 7.33% |
| Aβ42/Aβ40 | low | 7.86% |
| np-tau217 | high | 6.26% |
| np-tau217 | low | 10.36% |
| p-tau217 | high | 7.50% |
| %p-tau217 | high | 5.24% |

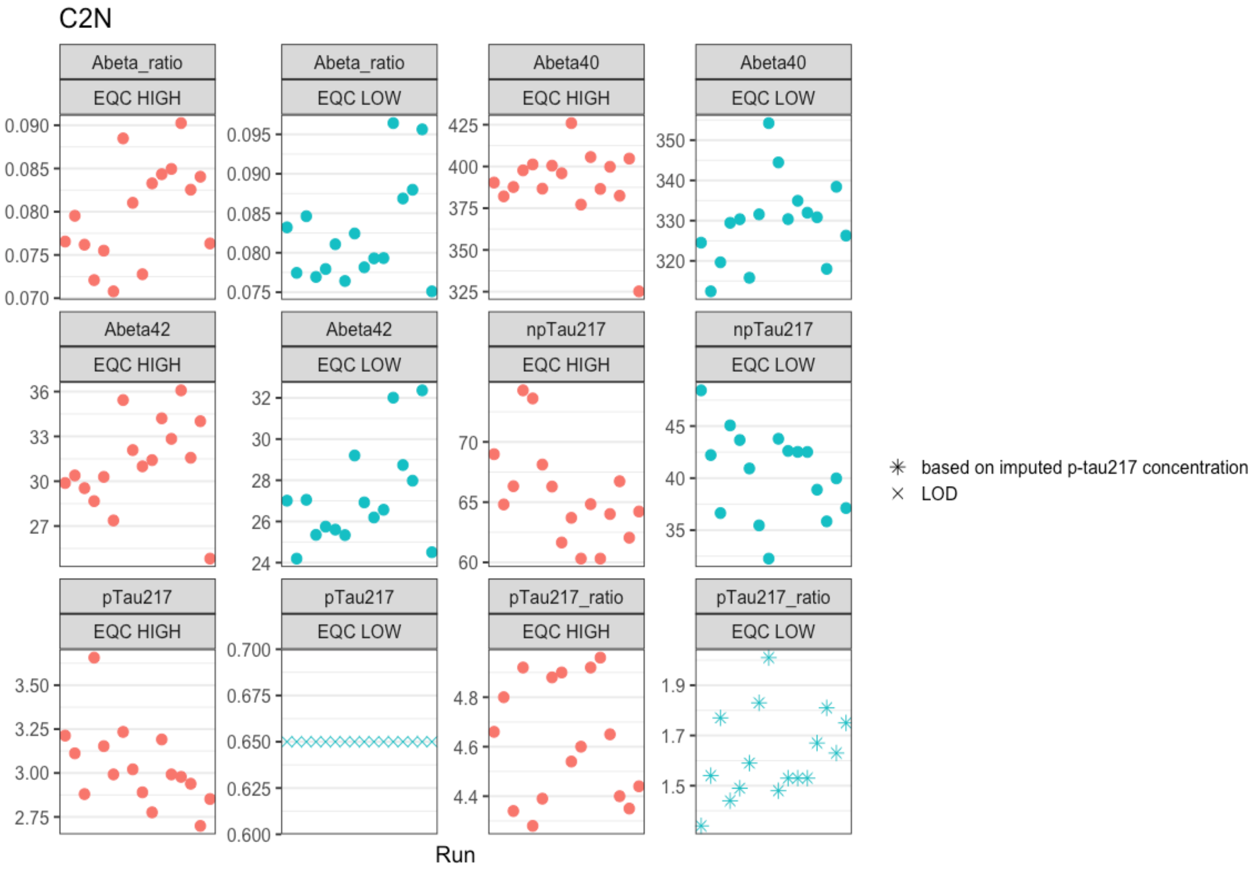

**Fujirebio Lumipulse**

| Assay | QC control | CV |
|---|---|---|
| p-tau217 | high | 11.47% |
| p-tau217 | low | 33.62% |

Fujirebio

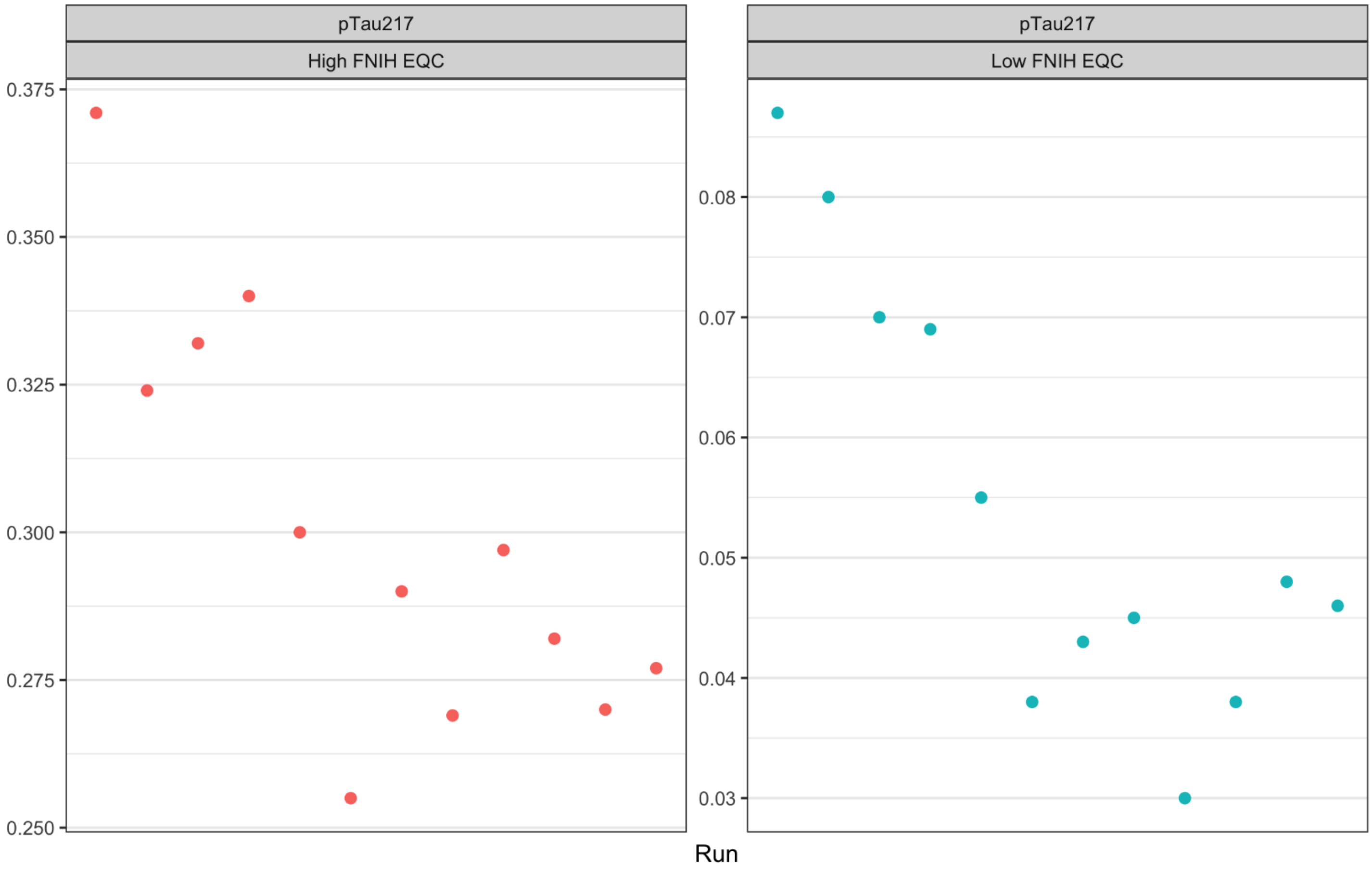

**Janssen LucentAD Quanterix**

| Assay | QC | CV |
|---|---|---|
| p-tau217 | high | 5.63% |
| p-tau217 | low | 7.65% |

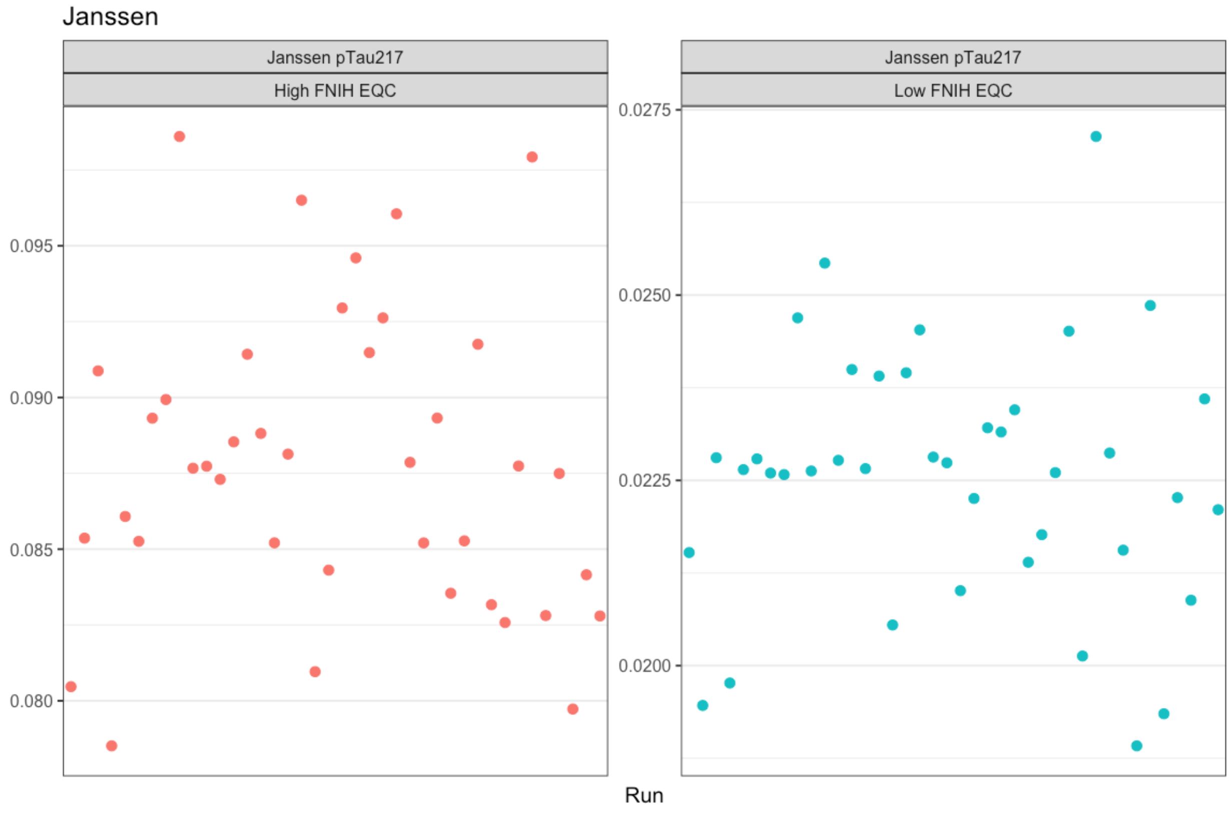

**Roche NeuroToolKit**

| Assay | QC control | CV |
|---|---|---|
| Aβ40 | high | 5.5% |
| Aβ40 | low | 7.57% |
| Aβ42 | high | 4.77% |
| Aβ42 | low | 3.24% |
| GFAP | high | 1.95% |
| GFAP | low | 3.38% |
| NFL | high | 2.09% |
| NFL | low | 3.12% |
| p-tau181 | high | 4.5% |
| p-tau181 | low | 6.84% |

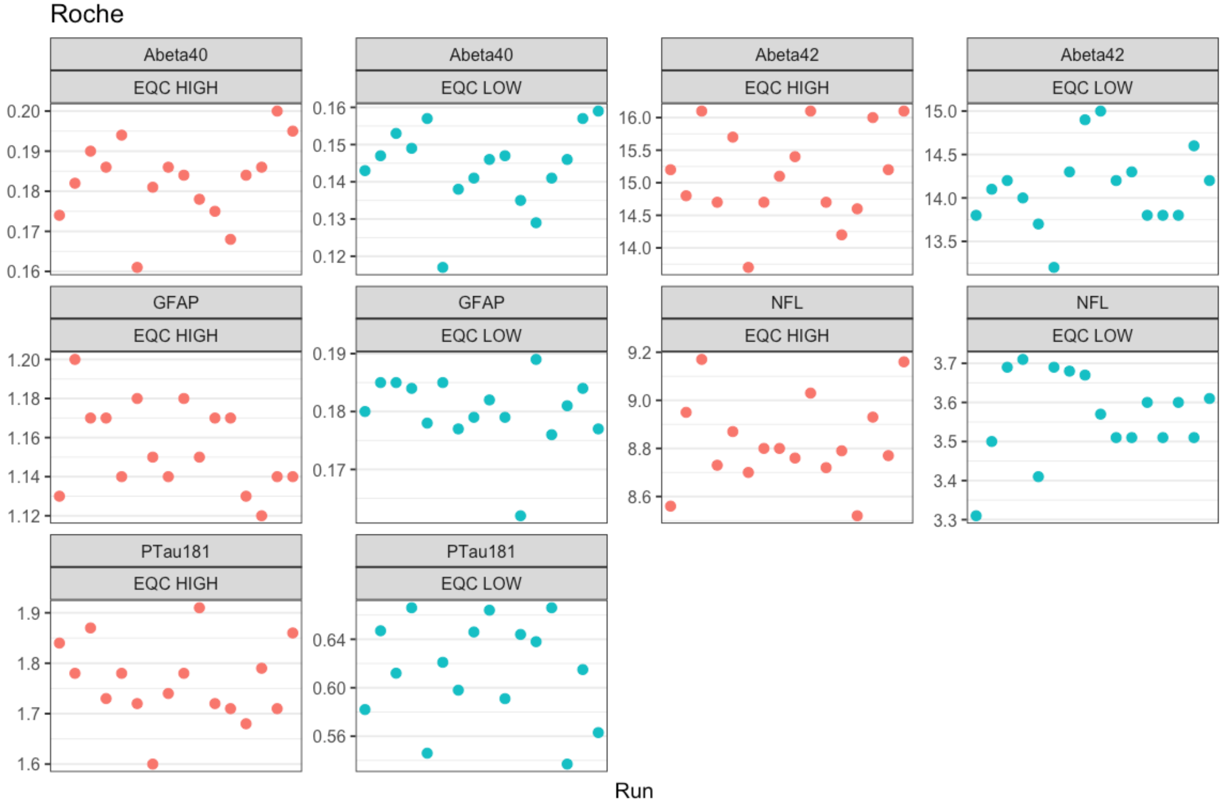

**Quanterix Neurology 4-Plex**

| Assay | QC control | CV |
|---|---|---|
| Aβ40 | high | 4.20% |
| Aβ40 | low | 4.07% |
| Aβ42 | high | 4.08% |
| Aβ42 | low | 5.05% |
| GFAP | high | 6.51% |
| GFAP | low | 4.89% |
| NfL | high | 5.79% |
| NfL | low | 4.89% |
| p-tau181 | high | 3.16% |
| p-tau181 | low | 5.97% |

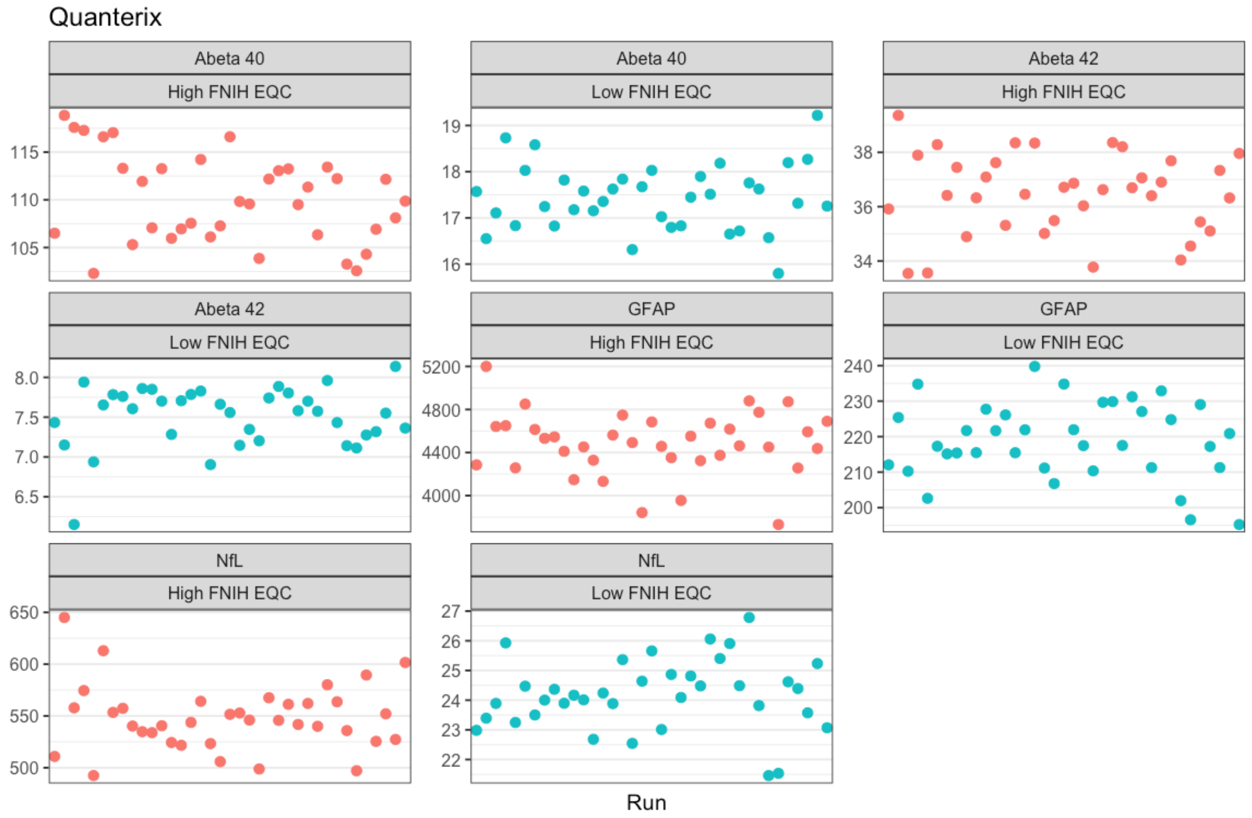

Quanterix pTau181

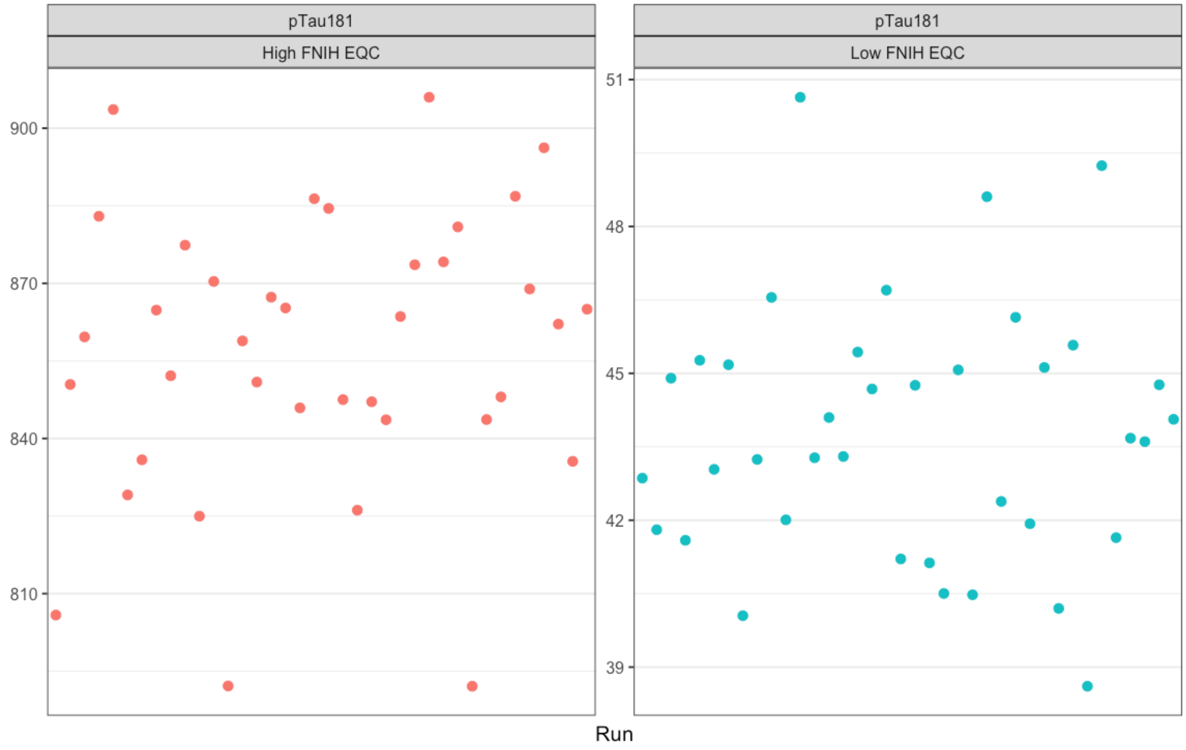
